# ZNF683-positive tissue-resident memory T cells organize treatment-refractory niches with targetable circuits in inclusion body myositis

**DOI:** 10.64898/2026.11.16.26367991

**Authors:** Francia Victoria A. De Los Reyes, Shinichiro Hayashi, Mie Kato, Jantima Tanboon, Satoru Noguchi, Ichizo Nishino

## Abstract

Inclusion body myositis (IBM) is a progressive autoimmune myositis refractory to conventional immunosuppression, yet mechanisms sustaining chronic inflammation remain unresolved. Integrating comparative single-nucleus RNA sequencing, spatial transcriptomics, and orthogonal validation across autoimmune myositis, we identified ZNF683+IL7R+ cytotoxic tissue-resident memory T (TRM) cells with natural killer cell-like features enriched in IBM. Trajectory analysis positioned an early-disease ZBTB16+ cytotoxic T cell state upstream of ZNF683+IL7R+TRM. *ZNF683* overexpression induced memory and natural killer cell-like transcriptional programs while attenuating glucocorticoid-response signatures, linking this program to treatment refractoriness. Ligand–receptor modeling nominated candidate targetable circuits involving IL-15–IL15RA, XCL1–XCR1, and HMGB1/CXCL12–CXCR4. Spatial transcriptomics localized ZNF683+TRM to stress-associated IBM niches. These findings define a disease-specific TRM-centered circuit linked to treatment refractoriness and candidate therapeutic vulnerabilities in IBM.

**One-sentence summary:** ZNF683+IL7R+ TRM cells organize targetable treatment-refractory IBM niches involving IL-15-IL-15RA, XCL1-XCR1, and HMGB1/CXCL12-CXCR4 axes.

## INTRODUCTION

Inclusion body myositis (IBM) is a sporadic, progressive autoimmune myositis (AM) that most commonly occurs in adults over 50 years (1–4). IBM remains refractory to conventional immunosuppression and lacks disease-modifying therapy. A defining pathological feature is cytotoxic CD8+ T cell invasion of non-necrotic myofibers, but the cellular programs and tissue circuits that sustain this chronic inflammation despite treatment remain incompletely understood (5–11). Identifying IBM-specific immune mechanisms, rather than inflammatory programs shared broadly across AM subtypes, is therefore essential for developing mechanism-based biomarkers and targetable therapeutic strategies.

Previous studies characterized IBM-associated T cells as KLRG1-expressing, highly differentiated cytotoxic T cells (12, 13). However, a KLRG1-centered terminal differentiation model alone does not fully explain how cytotoxic T cells persist in muscle, maintain IFN-γ-associated inflammatory programs, and interact with diseased myofibers and interstitial populations (14–17). The absence of an identified exogenous pathogen raises the possibility that endogenous antigen-presentation pathways, including lipid antigen presentation and natural killer T cell-like programs, contribute to immune persistence in IBM. Studies implicating natural killer T cell-like programs in autoimmune and neuroinflammatory diseases support this possibility (18–23).

In this context, it is important to consider that invariant natural killer T cells (iNKT cells) recognize CD1d-bound lipid antigens presented by dendritic cells, macrophages, and B cells, and can adopt cytotoxic or helper-like effector programs (24, 25). As such, transcriptional features shared with iNKT and natural killer cells may therefore provide a framework for examining innate-like cytotoxic T cell states in IBM (26, 27).

Here, by integrating comparative single-nucleus RNA sequencing, spatial transcriptomics, orthogonal validation across independent cohorts, and analysis of a *ZNF683*-overexpression dataset, we identify a ZNF683+IL7R+ cytotoxic tissue-resident memory T cell state with natural killer cell-like transcriptional programs and attenuated glucocorticoid-response signatures, suggesting a transcriptional basis for treatment refractoriness. We further map candidate targetable intercellular circuits involving IL-15–IL15RA, XCL1–XCR1, and HMGB1/CXCL12–CXCR4 across dendritic cells, myofibers, and stromal populations. These findings define an IBM-specific, treatment-refractory immune–myofiber–interstitial niche and nominate cellular and molecular vulnerabilities for therapeutic exploration.

## RESULTS

### Single-nucleus RNA sequencing reveals an IBM-enriched cellular landscape

To identify IBM-enriched inflammatory features and candidate intercellular circuits, we assembled a muscle pathology-defined discovery cohort for snRNA-seq and orthogonal validation (Fig. 1A). The discovery cohort comprised 25 donors: 4 with IBM, 5 with anti-synthetase syndrome (ASyS), 9 with dermatomyositis (DM), 5 with immune-mediated necrotizing myopathy (IMNM), and 2 adult nondisease controls. ASyS subtypes were anti-Jo-1 (n=2), anti-OJ (n=2), and anti-PL-7 (n=1). DM subtypes were anti-MDA5 (n=2), anti-Mi-2 (n=2), anti-NXP-2 (n=2), anti-TIF1-γ (n=2), and anti-SAE (n=1). IMNM subtypes were anti-HMGCR (n=2) and anti-SRP (n=3) (Fig. 1B; table S1). Analysis of 127,888 nuclei resolved 13 transcriptionally distinct nuclei classes based on canonical gene-signature markers, establishing the cellular framework for identifying the inflammatory features that distinguish IBM from other AM subtypes (Fig. 1, B to D; fig. S1, A and B; tables S1 to S3) (29–33). IBM muscle showed relative enrichment of cytotoxic T cells (TC; *CD8A*, *CD8B*, *THEMIS*, *CD3D*, *CD247*), helper T cells (TH; *CD4*, *THEMIS*, *CD3D*, *CD247*), and antigen-presenting cells (APC; *MSR1*, *MRC1*, *CD86*, *ITGAX*). Proportions of fast (*MYH1*, *MYH2*; MyoF) and slow (*MYH7*; MyoS) myofiber nuclei were relatively reduced, and regenerating myofiber nuclei (*MYH3*, *MYH8*; MyoR) were expanded. Muscle stem cells (MuSC; *PAX7*, *NCAM1*) were stable across conditions. Fibro-adipogenic precursors/mesenchymal stromal cells (FAP/MSC; *PDGFRA*, *BMPER*, *APOD*), pericytes (PC; *PDGFRB*), endothelial cells (EC; *PECAM1*, *VWF*), lymphatic endothelial cells (LEC; *PECAM1*, *VWF*, *MSR1*), B cells (BC; *CD19*, *MS4A1*), and plasma cells (PlC; *XBP1*, *SDC1*) varied among conditions without IBM-specific expansion.

**Fig. 1.**
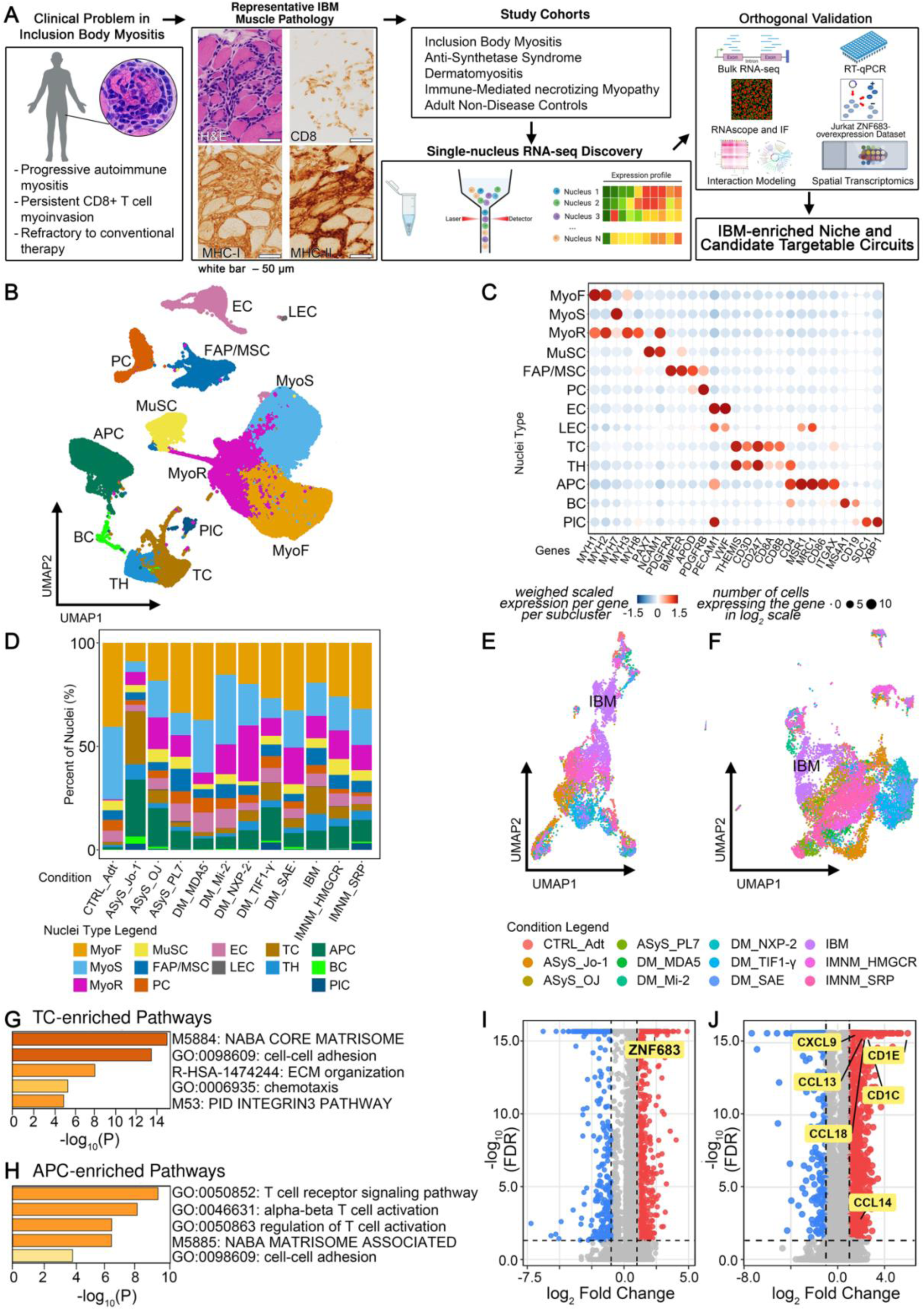
Translational study design and single-nucleus landscape of inclusion body myositis. (A) Workflow linking representative IBM muscle pathology, study cohorts, and single-nucleus RNA-seq discovery. Representative diagnostic IBM muscle-biopsy findings: hematoxylin and eosin (H&E), CD8, major histocompatibility complex class I (MHC-I), and class II (MHC-II) staining. Scale bars, 50 μm. snRNA-seq findings were evaluated using bulk RNA-seq, reverse transcription-quantitative polymerase chain reaction (RT-qPCR), RNAscope and immunofluorescence (IF), a deposited Jurkat *ZNF683*-overexpression dataset, intercellular interaction modeling, and spatial transcriptomics to define an IBM-enriched cellular niche and nominate candidate targetable circuits. (B) Uniform manifold approximation and projection (UMAP) of 13 nuclei classes from integrated snRNA-seq data. (C) Dot plot of marker genes defining each nuclei class. Color: weighted scaled expression within each class. Dot size: log₂-scaled number of expressing nuclei. (**D)** Nuclei-class composition across conditions. (**E and F)** UMAPs of T cells (TC) and antigen-presenting cells (APC) nuclei colored by condition. (G and H) Pathway enrichment of genes upregulated in IBM TC and APC. (**I and J)** Volcano plots showing differential gene expression in IBM versus other AM subtypes in TC nuclei (*ZNF683* highlighted) and APC nuclei (*CD1C*, *CD1E*, *CCL13*, *CCL14*, *CCL18*, and *CXCL9* highlighted). Axes: log₂ fold change and −log₁₀ FDR.

High-resolution reclustering of TC (Fig. 1E) and APC (Fig. 1F) showed that IBM nuclei occupied transcriptionally discrete regions within both compartments. Pathway enrichment analysis of IBM-upregulated genes (Fig. 1G–H; tables S4 and S5) identified cell adhesion, chemotaxis, and extracellular matrix-associated programs in TC nuclei, and T cell signaling, T cell activation, and matrisome remodeling pathways in APC nuclei. These programs suggested coordinated immune-cell positioning and tissue remodeling within IBM muscle. Differential expression analysis identified *ZNF683* (34, 35) as one of the most upregulated genes in IBM TC nuclei relative to other AM subtypes (Fig. 1I). In APC nuclei, IBM was associated with increased expression of *CD1C* and *CD1E*, which encode CD1 family molecules involved in lipid antigen presentation, together with chemokines *CCL13*, *CCL14*, *CCL18*, and *CXCL9* (Fig. 1J) (36–39). These transcriptional changes are consistent with APC programs that may support antigen presentation, T cell recruitment, positioning, and macrophage-mediated activation in IBM muscle.

### IBM enriches ZNF683+IL7R+ cytotoxic TRM cells

TC subclustering resolved 12 gene signature-defined subpopulations across conditions (Fig. 2A; fig. S2A) (22, 23, 25, 26, 40–42). The cluster label iNKT-like_TC denotes a CD8+ T cell population with transcriptional and TCR repertoire features reminiscent of iNKT-like biology, including *THEMIS*, *ZBTB16*, and *CRTAM* expression, without implying bona fide CD1d-restricted iNKT cell identity or function. T cells with NK-like cytotoxic gene signatures but without iNKT-like features were annotated as NK-like cytotoxic T cells (NK-like_TC; *THEMIS*, *CCL4*, *CCL5*, *DNAJB1*, *KLRK1*). Subcluster proportions varied across conditions (Fig. 2B; fig. S2B; tables S6 and S7), with ZNF683+IL7R+TRM preferentially enriched in IBM. Within IBM (Fig. 2, C and D), iNKT-like_TC expressed *ZBTB16*, *CRTAM*, and *NR4A1* (43–45). NK-like_TC expressed cytotoxic and natural killer (NK) cell-associated genes, including *KLRK1*, *CCL4*, and *CCL5*, together with the stress-associated gene *DNAJB1*, and showed lower *ZBTB16*, *CRTAM*, and *NR4A1* expression (46, 47). A small NK cell subset (NKc) was identified within the broader T/NK cell compartment, characterized by *NCAM1* and *ZBTB16* expression, absence of *THEMIS*, and detectable *CD8A*/*CD8B* transcripts.

**Fig. 2.**
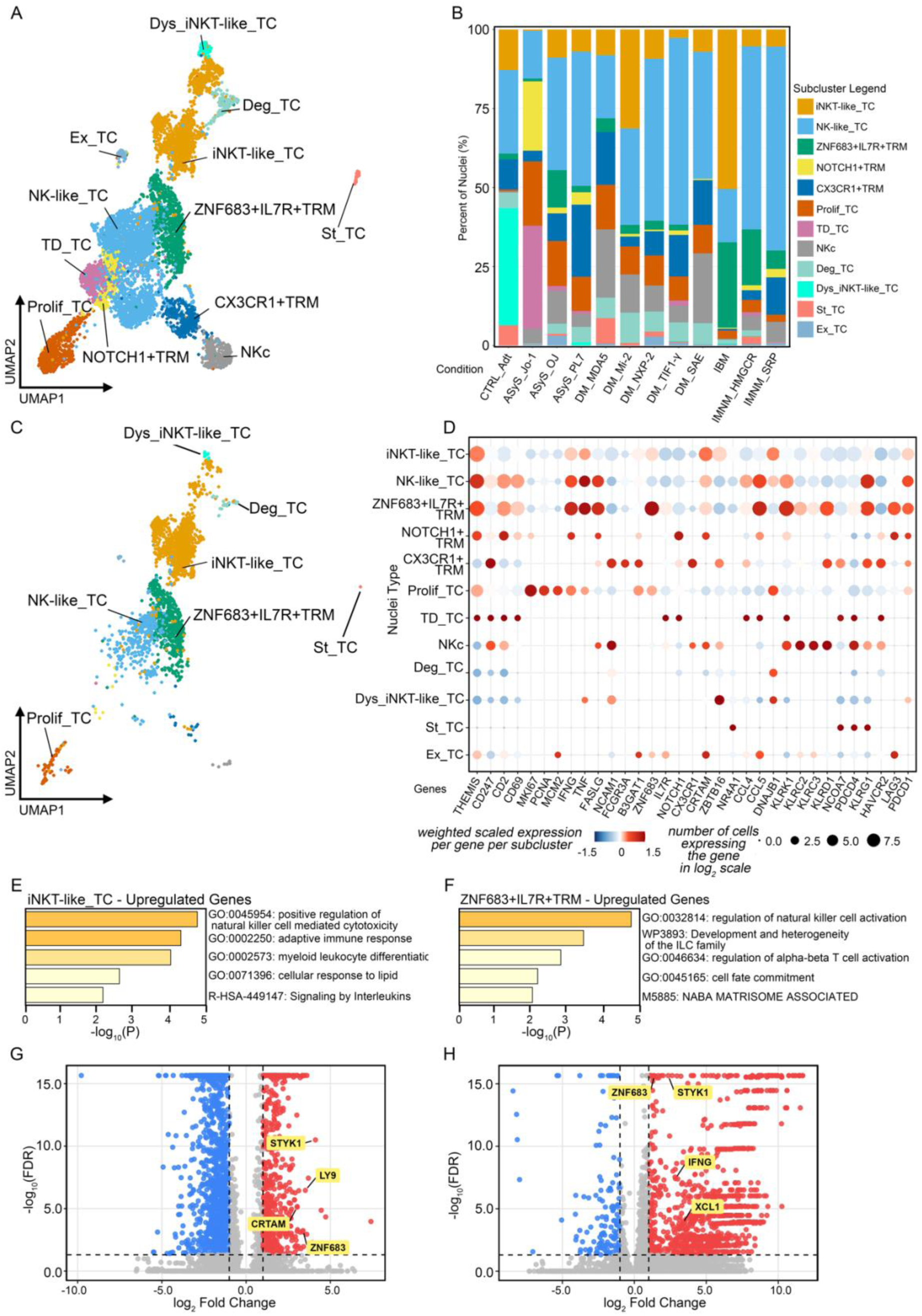
T cell subcluster architecture identifies IBM-enriched ZNF683+IL7R+ tissue-resident memory T cells. **(A)** UMAP of 12 T cell and NK cell subclusters from integrated snRNA-seq data. (**B)** Subcluster composition across conditions. CX3CR1+TRM predominates in adult control muscle, whereas ZNF683+IL7R+TRM were preferentially enriched in IBM. (**C)** UMAP of TC subclusters restricted to IBM nuclei. (**D)** Gene signature dot plot across IBM TC subclusters showing differential expression of activation, cytotoxicity, tissue-residency, and NK/NKT-like markers. (**E and F)** Pathway enrichment of genes upregulated in iNKT-like_TC (**E)** and ZNF683+IL7R+TRM (**F).** iNKT-like_TC was enriched for NK-like cytotoxicity, adaptive immune response, and lipid-sense programs, whereas ZNF683+IL7R+TRM was enriched for NK cell activation and innate lymphoid cell-related and T cell activation programs. (G and H) Volcano plots showing differentially expressed genes in iNKT-like_TC (G) and ZNF683+IL7R+TRM (H) compared with other TC subclusters. Selected shared genes, including *ZNF683* and *STYK1*, and subcluster-associated genes, including *CRTAM* and *LY9* in iNKT-like_TC and *IFNG* and *XCL1* in ZNF683+IL7R+TRM, are indicated. Axes show log₂ fold change and −log₁₀ false discovery rate (FDR).

ZNF683+IL7R+TRM formed a transcriptionally coherent cluster distinguished from other tissue-resident memory T cell (TRM) populations, including CX3CR1+TRM and NOTCH1+TRM, by strong *ZNF683* expression. *ZNF683* encodes HOBIT, a transcriptional regulator of tissue residency across TRM and NKT populations (34, 35, 59).

ZNF683+IL7R+TRM also expressed *CD69* and showed low *CX3CR1* expression, consistent with tissue retention and reduced recirculation (48, 49). This population additionally co-expressed *KLRG1*, *PDCD1*, *HAVCR2*, and *LAG3*, markers associated with terminal differentiation and functional restraint in chronically stimulated T cells (50–54). It retained inflammatory effector programs marked by *IFNG*, which can promote myofiber stress and immune amplification (99), and *XCL1*, which can recruit XCR1+ conventional type 1 dendritic cells (cDC1) and support antigen cross-presentation (94, 95). *IL7R* co-expression distinguishes this population from more terminally differentiated *ZNF683*-high states in which *IL7R* is typically downregulated, suggesting a memory-associated survival program that may support long-term tissue persistence (35, 51).

Pathway enrichment analysis (Fig. 2, E and F; tables S8 and S9) identified NK cell-mediated cytotoxicity and cellular response to lipid programs in iNKT-like_TC, together with NK cell activation and innate lymphoid cell family transcriptional signatures in ZNF683+IL7R+TRM. Both subclusters expressed *ZNF683* and *STYK1* (a kinase enriched in the murine NK-cell transcriptional signature and implicated in cell trafficking) (60, 61), although *ZNF683* expression was markedly higher in ZNF683+IL7R+TRM. *CRTAM* and *LY9* were preferentially expressed in iNKT-like_TC (Fig. 2, G and H). CRTAM has been reported on activated IFN-γ-producing human iNKT cells (62, 63), whereas *LY9*/SLAMF3 regulates iNKT cell development and activation in mice (64). These findings are interpreted as supportive transcriptional features rather than lineage-defining markers. *IFNG* and *XCL1* were enriched in ZNF683+IL7R+TRM (55–59, 65, 66). TCR sequences associated with iNKT and atypical NKT lineages were detected in iNKT-like_TC, NK-like_TC, and ZNF683+IL7R+TRM in IBM (fig. S3, A and B) (67, 68). Direct CD1d restriction was not tested, and comparable TCR-sequence testing was not performed in other AM subtypes, limiting both lineage assignment and assessment of disease specificity at the repertoire level.

### Temporal T cell states in IBM

TC subcluster proportions varied with disease duration across IBM samples (Fig. 3A; table S10). Samples from patients with shorter disease duration (≤24 months) tended to show higher proportions of iNKT-like_TC, whereas those with longer disease duration (>24 months) tended to contain increased proportions of ZNF683+IL7R+TRM and NK-like_TC. Inter-individual variability was observed within this pattern, including one patient with 56 months of disease duration (IBM_4) who showed a higher iNKT-like_TC proportion than the patient with 48 months (IBM_3).

**Fig. 3.**
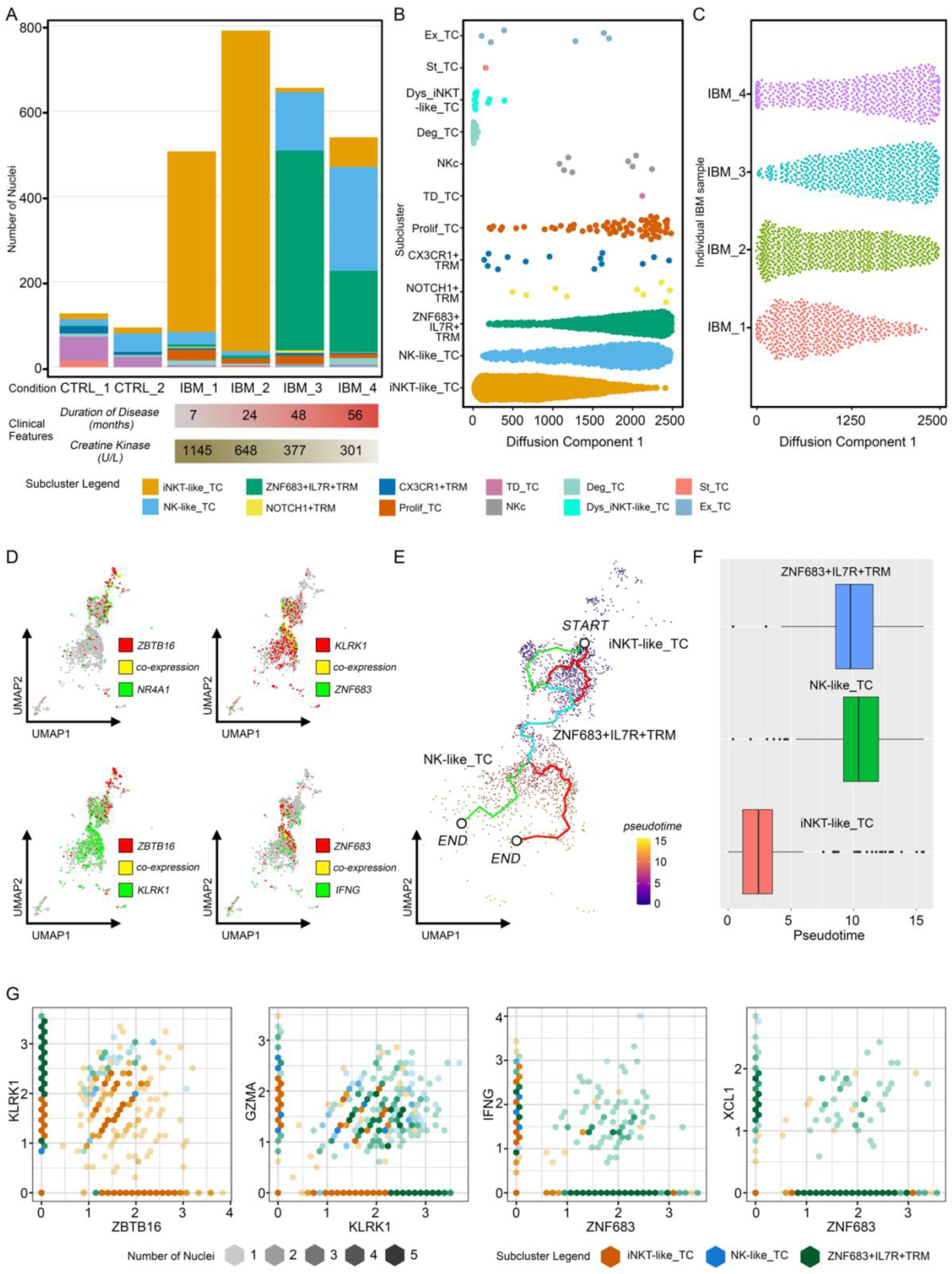
Pseudotime and co-expression analyses suggest a disease duration-associated transcriptional continuum from iNKT-like cytotoxic TC to ZNF683+IL7R+TRM. (A) TC subcluster composition across IBM samples and controls showed iNKT-like_TC relatively enriched in early disease (≤24 months). ZNF683+IL7R+TRM were increased in samples with longer disease duration. Disease duration (months) and serum creatine kinase levels are indicated below each sample. (B–C) Diffusion component analysis of major TC subclusters (B) and individual IBM samples (C). iNKT-like_TC occupied the low end of diffusion component 1, with ZNF683+IL7R+TRM at progressively higher values, stratifying by duration and suggesting a transcriptional continuum. (D) Co-expression dot plot of *ZBTB16*, *NR4A1*, *KLRK1*, *ZNF683*, and *IFNG* across major TC subclusters. *ZBTB16* and *NR4A1* were enriched in iNKT-like_TC. *ZNF683* and *IFNG* were concentrated in ZNF683+IL7R+TRM. (**E–F)** Pseudotime analysis inferring transcriptional ordering from iNKT-like_TC, set at the START node, toward ZNF683+IL7R+TRM and NK-like_TC end states. The UMAP with START and END nodes is in **E**. Pseudotime distributions by subcluster are in **F**. (G) Pairwise co-expression analyses proposing shared expression of cytotoxic genes, including *KLRK1* and *GZMA*, across iNKT-like_TC, NK-like_TC, and ZNF683+IL7R+TRM, with co-enrichment of *IFNG* and *XCL1* with *ZNF683* in ZNF683+IL7R+TRM. Point size reflects nuclei count per observation.

Diffusion component analysis identified a transcriptional continuum along diffusion component 1, with iNKT-like_TC occupying the low end and ZNF683+IL7R+TRM distributed toward progressively higher values (Fig. 3B). The distribution was consistent with a maturation continuum supported by shared TCR repertoire features (67, 68). Early disease duration samples localized toward the low end of this axis, whereas samples with longer disease duration localized closer to ZNF683+IL7R+TRM and NK-like_TC states (Fig. 3C).

Co-expression analysis showed enrichment of *ZBTB16* and *NR4A1* in iNKT-like_TC, prominent *KLRK1* expression in NK-like_TC, and concentration of *ZNF683*/*IFNG* in ZNF683+IL7R+TRM (Fig. 3D). Pseudotime analysis, with iNKT-like_TC set as the start state, placed iNKT-like_TC at earlier positions and ordered ZNF683+IL7R+TRM and NK-like_TC toward separate terminal states (Fig. 3, E and F). This pattern supported a probabilistic continuum rather than a linear differentiation trajectory.

Pairwise co-expression analysis showed that *KLRK1* and *GZMA* were broadly expressed across iNKT-like_TC, NK-like_TC, and ZNF683+IL7R+TRM, consistent with shared cytotoxic gene programs across these states (Fig. 3G). In contrast, *IFNG* and *XCL1* were co-enriched with *ZNF683* in ZNF683+IL7R+TRM, suggesting selective amplification of inflammatory and chemotactic effector programs in this state. Per-condition feature plots confirmed IBM specificity of *ZNF683*, *IL7R*, *XCL1*, and *KLRG1* co-expression within this cluster (fig. S4, A to D). Together, these data support a model in which an iNKT-like_TC-predominant early state is followed by ZNF683+IL7R+TRM accumulation, establishing a persistent cytotoxic effector program.

### Transcript-level validation of *ZNF683* expression across IBM disease duration

RNA in situ hybridization with immunofluorescence demonstrated *ZNF683* mRNA signal in regions enriched for CD8+ cells (Fig. 4, A to D), with concordant mRNA detection in areas surrounding myofibers lacking p62-positive aggregates on co-staining (Fig. 4, A to D), consistent with ZNF683+ TRM enrichment at non-degenerating myofiber regions (table S11). To validate snRNA-seq findings in independent samples, bulk RNA-seq compared IBM (n=13) with other AM subtypes (ASyS_Jo-1, n=13; ASyS_OJ, n=13; DM_MDA5, n=12; DM_Mi-2, n=13; DM_NXP-2, n=12; DM_TIF1-γ, n=16; IMNM_HMGCR, n=11; IMNM_SRP, n=11) and adult controls (n=4) (Fig. 4E; tables S12 and S13). *ZNF683* was the top upregulated gene in IBM TC, co-upregulated with genes encoding cytotoxic and innate T cell surface markers (*CD8A*, *CD247*, *CD28*, *KLRC1*, *KLRC3*, *KLRC4*, *KLRK1*, *KLRB1*, *KLRD1*), the *KLRC4–KLRK1* readthrough transcript, cytokines (*IFNG*, *XCL1*, *CCL4*, *CCL5*), proteases (*GZMA*, *GZMK*, *GZMH*, *GZMB*, *GZMM*), and the pore-forming protein *PRF1* (69–71). This indicates that *ZNF683* upregulation in IBM occurs as part of a coordinated cytotoxic T cell program. IBM showed significantly higher TC-related signature expression than all other AM subtypes examined (χ²=51.89, df=8, *p*=1.77×10⁻⁸) (Fig. 4F; table S14). Within IBM, *ZNF683* expression (CPM) was significantly higher in cases with >24 months disease duration (n=8) than ≤24 months (n=5) (Fig. 4G; table S15).

**Fig. 4.**
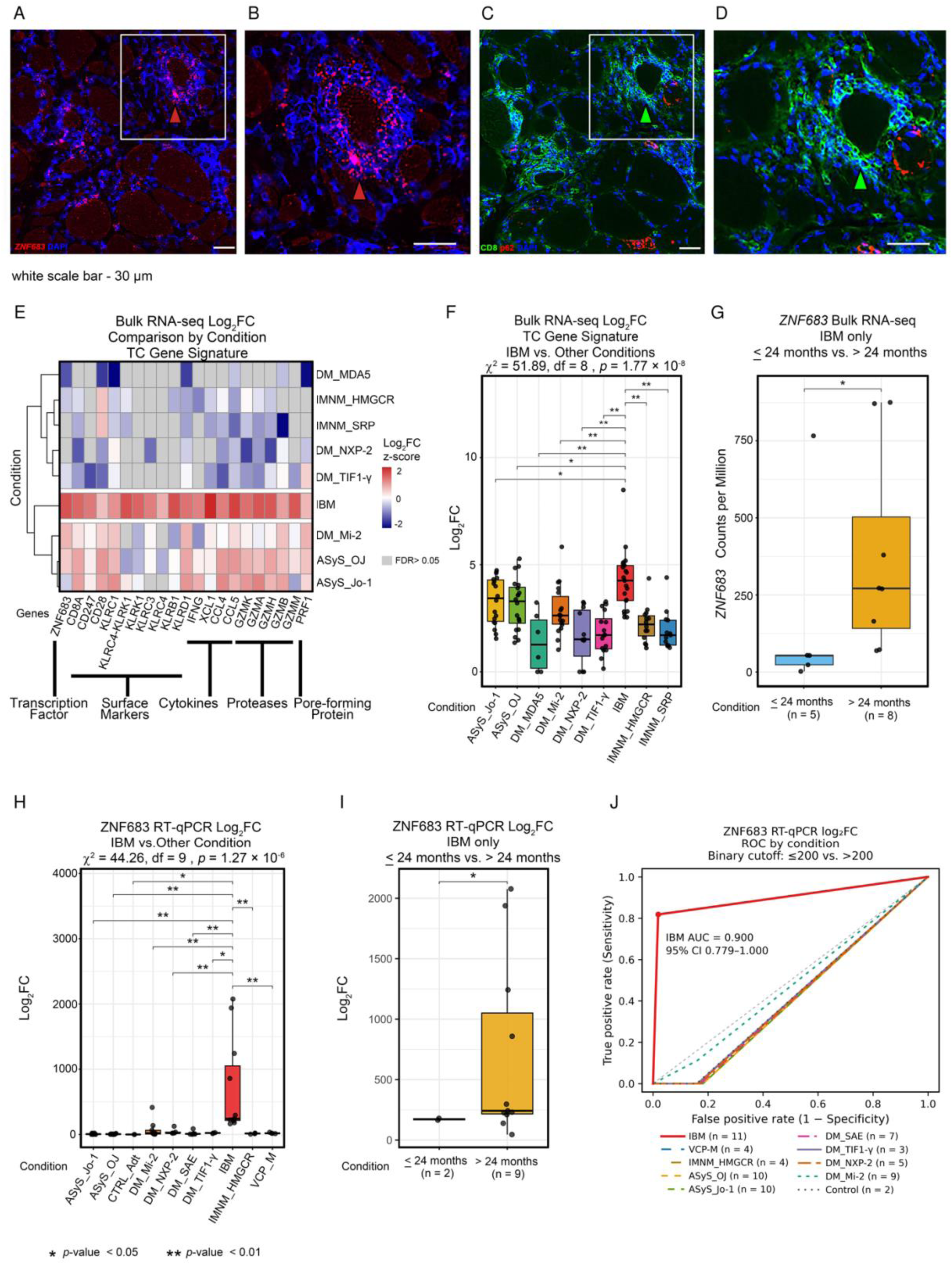
*ZNF683* expression is selectively upregulated in IBM muscle and higher with increased disease duration. (A and B) RNAscope detection of *ZNF683* transcripts (red) with DAPI (blue). (C and D) Corresponding CD8 (green), p62 (red), and DAPI (blue) co-staining. B and D enlarge boxed regions in A and C. White scale bars, 30 μm. (E) Bulk RNA-seq log₂ fold change (log₂FC) heatmap of a coordinated cytotoxic T cell gene signature across conditions. Gene categories: transcription factor (*ZNF683*), surface markers, cytokines (*IFNG*, *XCL1*, *CCL4*, *CCL5*), proteases, and pore-forming protein (*PRF1*). (F) Bulk RNA-seq log₂FC distribution of the TC gene signature across conditions; IBM expression is significantly higher than all other AM subtypes (Kruskal–Wallis χ² = 51.89, df = 8, *p* = 1.77 × 10⁻⁸). (G) Bulk RNA-seq *ZNF683* expression (CPM) stratified by IBM disease duration: >24 months (n = 8) vs. ≤24 months (n = 5). (H) RT-qPCR validation of *ZNF683* expression across conditions; IBM shows significantly higher expression than all other groups (Kruskal–Wallis χ² = 44.258, df = 9, *p* = 1.27 × 10⁻⁶). (I) RT-qPCR *ZNF683* expression stratified by IBM disease duration: >24 months (n = 9) vs. ≤24 months (n = 2); two-sided Welch t-test, t = −2.508, df = 8.016, *p* = 0.036. (J) Binary ROC curves of *ZNF683* RT-qPCR log₂FC (≤200=0, >200=1) for each condition versus all remaining samples. IBM (n=11) versus non-IBM (n=54): AUC 0.900 (95% DeLong CI 0.779–1.000, upper bound capped at 1), sensitivity 81.818% (9/11), and specificity 98.148% (53/54). Red point: >200 cutoff for IBM. Control denotes nondisease adult controls. Some condition curves overlap.

RT-qPCR confirmed elevated *ZNF683* expression in IBM (n=11) relative to other AM subtypes and non-myositis controls (χ²=44.258, df=9, *p*=1.27×10⁻⁶; post hoc pairwise *p*=0.001–0.038) (Fig. 4H; tables S16 and S17). Within IBM, RT-qPCR showed higher *ZNF683* expression in cases with >24 months disease duration (n=9) than ≤24 months (n=2) (two-sided Welch t=−2.508, df=8.016, *p*=0.036, 95% CI=−1229.882 to −51.734 for the shorter-minus-longer difference on the log₂FC scale) (Fig. 4I; table S18). RT-qPCR log₂FC expression thresholds were mild (>50), moderate (>100), and marked (>200). Using a binary RT-qPCR log₂FC score (0 for ≤200 and 1 for >200), IBM (n=11) was compared with all non-IBM samples (n=54), yielding an AUC of 0.900 (95% DeLong CI, 0.779–1.000; upper bound capped at 1), sensitivity of 81.818% (9/11), and specificity of 98.148% (53/54) (Fig. 4J; table S19). Figure 4J and table S19 also show the binary analysis for each remaining condition versus all other samples. Convergent evidence across snRNA-seq, bulk RNA-seq, and RT-qPCR in independent cohorts supports *ZNF683* as an IBM-specific marker whose accumulation with disease duration appears consistent with the transcriptional continuum identified by trajectory analysis.

### CD8+ T cell myoinvasion is sustained across the full disease course in IBM

Muscle biopsies spanning 11 to 108 months of symptom duration demonstrated endomysial CD8+ cytotoxic T cell infiltration with invasion of non-necrotic myofibers at all timepoints (fig. S5, A to D). CD8+ density did not scale with disease duration, and CD68+ macrophage infiltration was observed across all strata. Myoinvasion was noted at morphologically intact myofibers lacking rimmed vacuoles (fig. S5C) and in regions of otherwise normal-appearing muscle (fig. S5D), consistent with the established IBM pathological hallmark of cytotoxic T cell targeting of non-degenerating fibers. Together, these findings indicate that CD8+ T cell myoinvasion in IBM is directed at antigenically competent, non-degenerating fibers across the full disease course, suggesting that sustained antigen exposure contributes to the chronic cytotoxic T cell response. This provides the histopathological basis for the transcriptionally distinct CD8+ T cell states identified by snRNA-seq.

### Protein validation of temporal CD8+ T cell states in IBM

Immunofluorescence independently supported the transcriptional continuum identified by snRNA-seq, showing disease duration-associated shifts in CD8+ T cell subcluster marker expression (Fig. 5, A to F). PLZF (*ZBTB16*) expression was significantly higher in patients with ≤24 months’ disease duration than in those with >24 months (OR=0.63, 95% CI 0.50– 0.80, *p*=1.42×10⁻⁴; Fig. 5, A and B). Conversely, KLRK1 and ZNF683 expression were higher with longer disease duration (KLRK1: OR=3.26, 95% CI 2.57–4.16, *p*=1.38×10⁻²⁴; ZNF683: OR=4.52, 95% CI 3.38–6.06, *p*=3.59×10⁻²⁷; Fig. 5, C to F). ZNF683+ CD8+ T cells adjacent to CAV3+ myofiber boundaries were enriched in longer disease duration, consistent with spatial accumulation of ZNF683+ TRM at the myofiber interface (Fig. 5, E and F). Patient clinical data are in table S20.

**Fig. 5.**
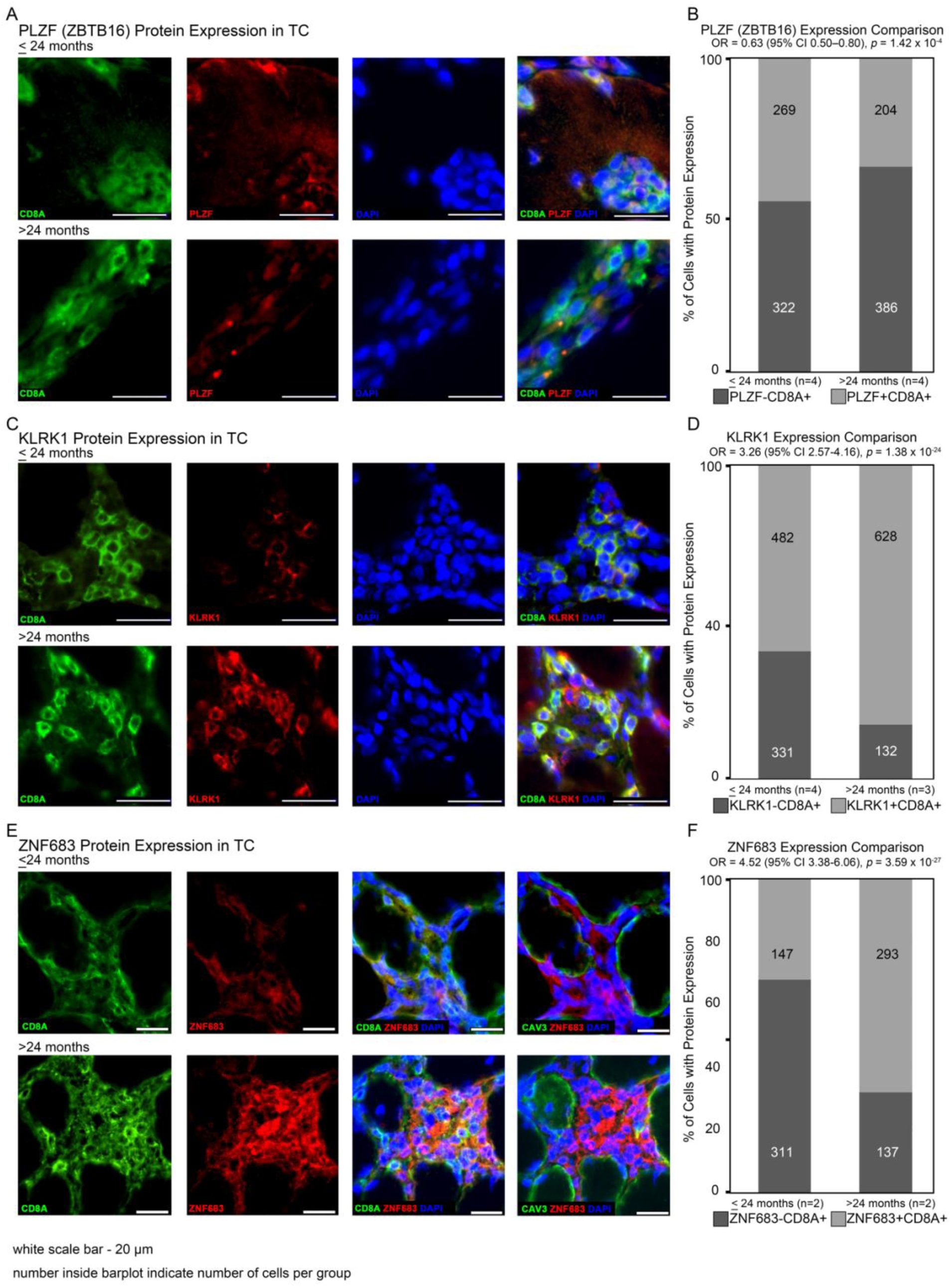
Protein-level validation of major CD8+ T cell subcluster markers in IBM muscle. **(A)** Representative immunofluorescence images of PLZF (encoded by *ZBTB16*) protein expression in CD8+ T cells from IBM muscle biopsies stratified by disease duration (≤24 months, n=4; >24 months, n=4). CD8+ T cells were identified by anti-CD8A immunostaining. Nuclei were counterstained with DAPI. (**B)** Bar plots show proportions of PLZF+ and PLZF− cells within the CD8+ compartment. Numbers within bars indicate total CD8+ T cells analyzed per group. (**C)** Representative immunofluorescence images of KLRK1 protein expression in CD8+ T cells from IBM biopsies stratified by disease duration (≤24 months, n=4; >24 months, n=3). Staining and quantification were performed as in **A**. (**D)** As in B, for KLRK1. (**E)** Representative immunofluorescence images of ZNF683 protein expression in CD8+ T cells (≤24 months, n=2; >24 months, n=2). Co-staining with CAV3 demarcates myofiber boundaries and establishes the spatial proximity of ZNF683+ CD8+ T cells to the myofiber interface. (**F)** As in B, for ZNF683. For all panels, odds ratios (OR) with 95% confidence intervals and two-sided *p* values are shown. Scale bars, 20 μm.

### Transcriptional consequences of *ZNF683* overexpression in Jurkat cells

To examine the transcriptional consequences of *ZNF683* expression in a tractable cell model, publicly available RNA-seq data from Jurkat cells with experimentally induced *ZNF683* overexpression (35) were analyzed (Fig. 6, A and B; tables S21 to S23). Upregulated pathways included leukocyte activation, cytokine signaling, and interferon signaling. Downregulated pathways included glucocorticoid response, unfolded protein response, stress-induced premature senescence, and nutrient-level response (72–77). Representative upregulated genes included the cytokine signaling components *JAK3* and *IL12RB2*, chemokine receptors *CCR2* and *CCR5*, interferon regulatory factors *IRF5* and *IRF6*, and memory-associated *CD44*. *HMOX1* was among the downregulated genes (Fig. 6C; tables S24 and S25). Curated gene-set heatmaps showed higher memory T cell- and NK cell-associated expression and lower stress-response expression in *ZNF683*-overexpressing cells (HOBIT_POS) than in controls (HOBIT_NEG) (Fig. 6, D to F; tables S26 to S28). The stress-response set in table S28 included *DDIT4* (REDD1), a glucocorticoid-responsive stress regulator (105), with a reported log₂FC of −1.66. This provides a specific glucocorticoid-related gene that may suggest broad cellular stress responses associated with glucocorticoid treatment. These results associate *ZNF683* overexpression with altered memory, NK-like, and stress-response programs.

**Fig. 6.**
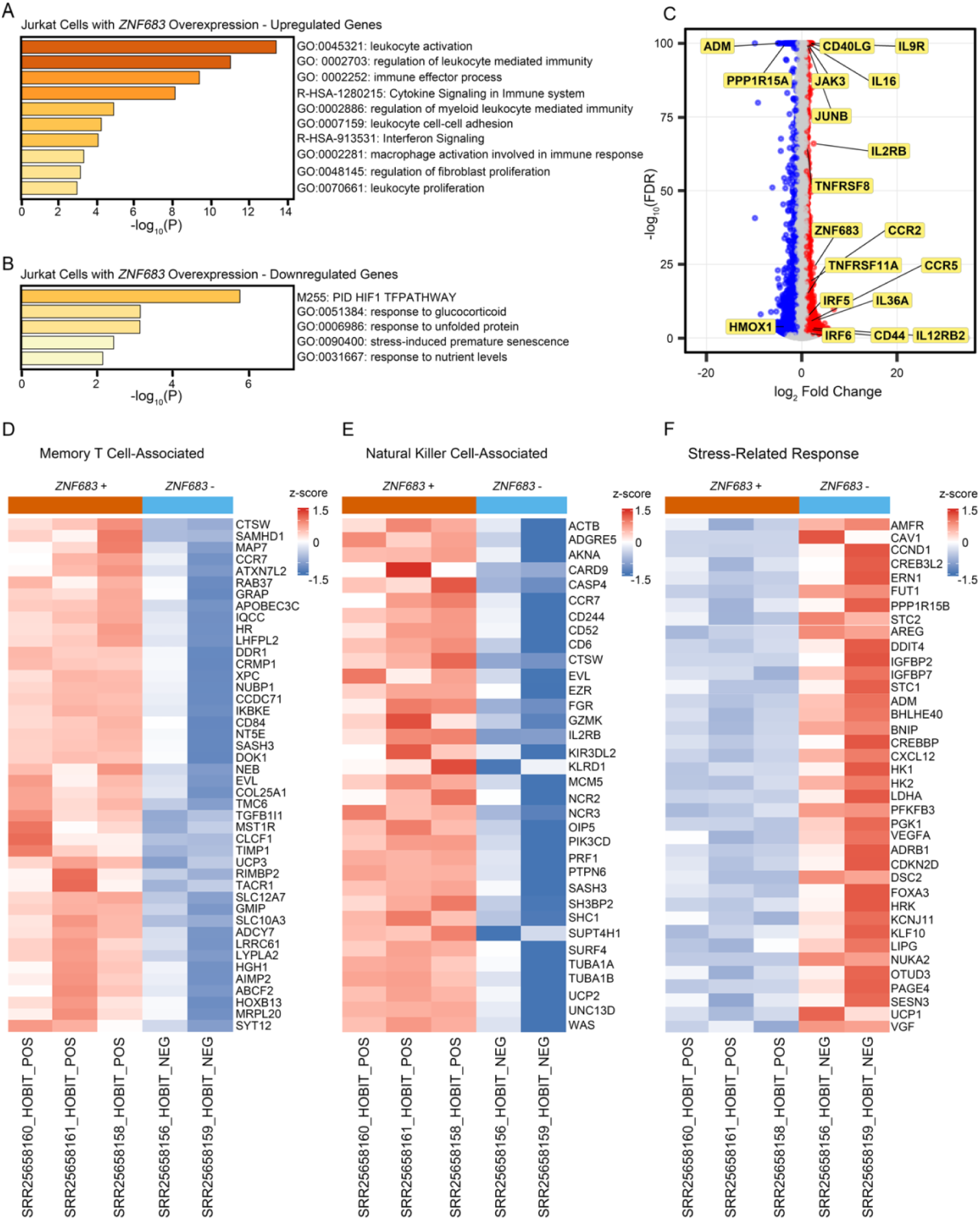
*ZNF683* overexpression in Jurkat cells activates cytokine and memory T cell transcription signatures while suppressing stress-response pathways. **(A–B)** Pathway enrichment of genes upregulated (**A**) or downregulated (**B**) following *ZNF683* overexpression. Upregulated pathways include leukocyte activation, cytokine signaling, and interferon signaling. Downregulated pathways include glucocorticoid response, unfolded protein response, and stress-induced premature senescence. (**C)** Volcano plot of differential gene expression following *ZNF683* overexpression. Upregulated genes include cytokine signaling components (*JAK3*, *IL12RB2*), chemokine receptors (*CCR2*, *CCR5*), interferon regulatory factors (*IRF5*, *IRF6*), and memory T cell marker *CD44*. *HMOX1* is among the most significantly downregulated genes, consistent with suppression of oxidative stress response. Axes: log₂ fold change (x) and −log₁₀ FDR (y). (**D–E)** Heatmaps of z-score-scaled expression of memory T cell-associated (**D**) and NK cell-associated (**E**) gene signatures in *ZNF683*-overexpressing (HOBIT_POS) and control (HOBIT_NEG) Jurkat cells. Both transcription signatures are enriched in HOBIT_POS relative to HOBIT_NEG. (**F)** Heatmap of z-score-scaled stress-response gene signatures. Relative downregulation in HOBIT_POS is consistent with suppression of glucocorticoid and stress-response transcription signatures. Z-scores were used for visualization only and do not define differential expression.

### Macrophage and dendritic cell composition in IBM

To contextualize TC within the broader inflammatory environment, APC subclustering using established gene signatures (78–82) resolved eight subpopulations with condition-specific compositional differences (Fig. 7, A and B; fig. S6, A and B; tables S29 and S30). The dominant population comprised *MRC1*-high macrophages with low *CD80*/*CD86* expression (M2), followed by macrophages with mixed *CD80*/*CD86*/*MRC1*/*CD68* expression (M1/2) and a smaller *CD80*/*CD86*-high subset (M1) (83). Conventional dendritic cell subclusters cDC1 and cDC2 were identified, with cDC1 defined by high *FLT3*, *CLEC9A*, and *XCR1* expression, and cDC2 by *FLT3* and *CD1C* co-expression, alongside a *CD2*-expressing dendritic cell population (84–87). *XCR1* expression was restricted to cDC1 (Fig. 7D; fig. S6A), identifying cDC1 as the candidate interaction partner for XCL1-mediated TRM– dendritic cell crosstalk. IBM APC nuclei formed a discrete cluster (Fig. 7C). M2 macrophages comprised the largest subpopulation across all four IBM samples regardless of disease duration (Fig. 7E; table S31; n=2 per group).

**Fig. 7.**
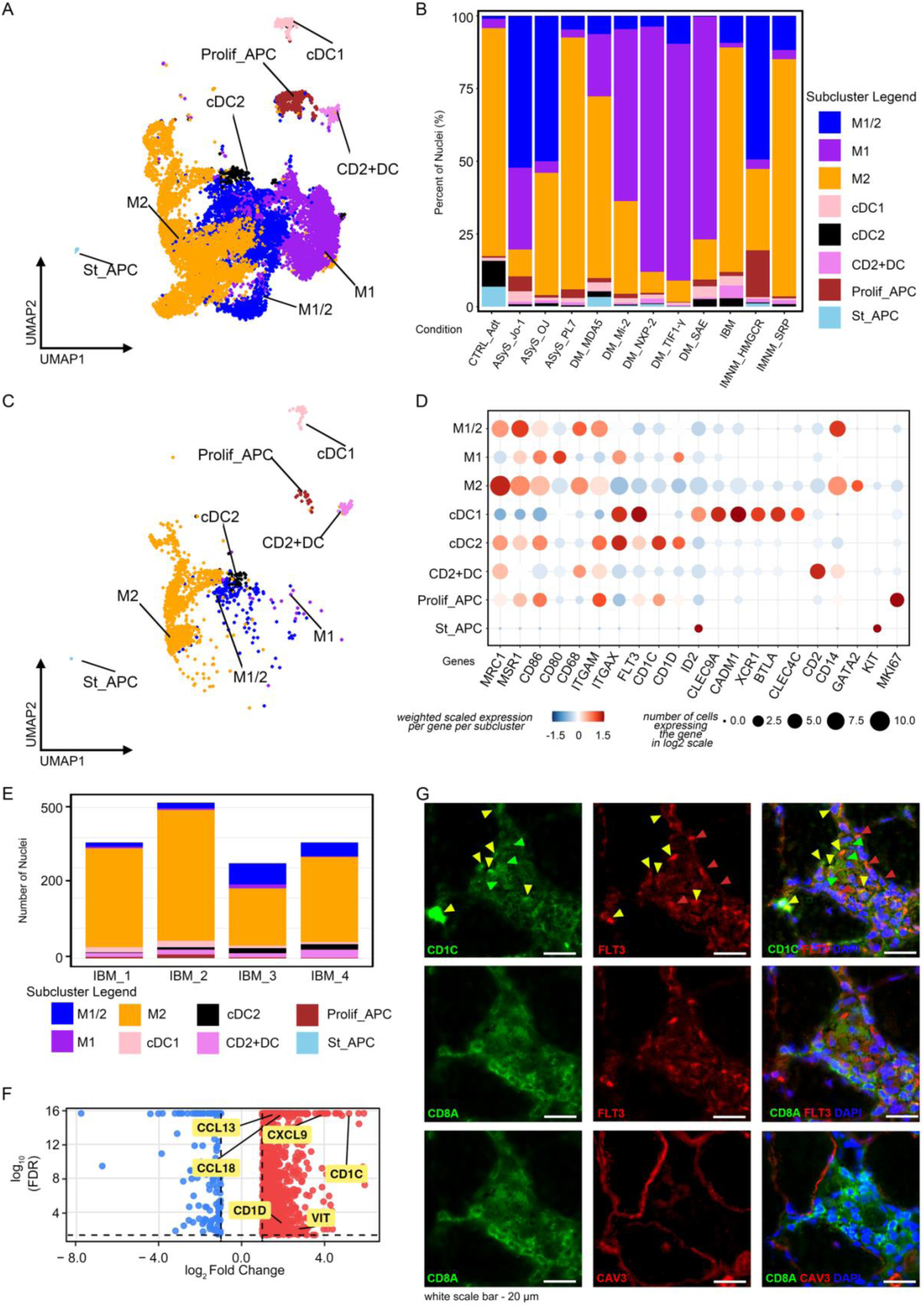
APC subcluster characterization in IBM muscle. **(A)** UMAP of APC nuclei across all conditions, resolving macrophage (M1, M2, M1/2), dendritic cell (cDC1, cDC2, CD2+DC), proliferative (Prolif_APC), and stem cell-like (St_APC) subclusters. (**B)** APC subcluster proportions across conditions. (**C)** IBM-only APC UMAP. (**D)** Weighted scaled expression and proportion of nuclei expressing canonical macrophage and dendritic cell markers across IBM APC subclusters. (**E)** APC subcluster composition in individual IBM samples (n=2 per disease duration group), showing M2 macrophage dominance across disease duration strata. (**F)** Volcano plot of IBM M2 macrophages versus those from all other conditions. Upregulated genes *CD1C*, *CD1D*, *CCL13*, *CCL18*, *CXCL9*, and *VIT* are highlighted. Axes show log₂ fold change and −log₁₀(FDR). (**G)** Immunofluorescence images from IBM muscle showing spatial organization of cDC1 (FLT3+), cDC2 (FLT3+CD1C+), M2 macrophages (CD1C+), and cytotoxic T cells (CD8A+) at the myofiber interface (CAV3+). Arrowheads indicate representative cells. White scale bars, 20 μm.

Differential expression analysis comparing IBM M2 macrophages with those from other AM (Fig. 7F; table S5) revealed upregulation of *CD1C* and *CD1D*, lipid antigen-presentation markers co-expressed in M2 macrophages and cDC2 (Fig. 7D), alongside *CCL13*, *CCL18*, and *CXCL9*, supporting TRM retention and activation through CCR5- and CXCR3-mediated chemotaxis (37, 88–90), and *VIT*, implicated in ECM remodeling (91). Immunofluorescence confirmed the spatial co-occurrence of APC populations with CD8A+ T cells at the myofiber interface (Fig. 7G; table S32), demonstrating proximity between CD1C+FLT3+ cDC2, CD1C+ M2 macrophages, and CD8A+ T cells near CAV3+ myofibers, and between FLT3+ cDC1 and CD8A+ T cells. This topological organization, with TRM near CD1-expressing APC and CAV3+ myofibers, proposes the in situ cellular basis for the interaction network in the following section.

The IBM APC transcriptional profile may indicate signals sustaining tissue-resident T cells with upregulation of CD1-family lipid antigen-presentation molecules. This is consistent with a microenvironment that maintains the resident cytotoxic T cell compartment and is not observed at equivalent magnitude in other AM.

### Inflammatory myofiber–interstitial cellular landscape in IBM

Probabilistic ligand–receptor analysis of TC–APC interactions (Fig. 8A; table S33) identified signaling pairs linking ZNF683+IL7R+ TRM with APC subpopulations. XCL1–XCR1 was detected exclusively in the TRM–cDC1 pairing, consistent with selective cDC1 recruitment for antigen cross-presentation (92–95). IFNG–IFNGR1/IFNGR2 interactions were inferred between TRM and all dendritic cell and macrophage populations, and TNF superfamily interactions (including FASLG and TNFSF10 with cognate receptors) were detected across multiple APC subtypes. Reciprocal APC-to-TRM signals (Fig. 8B; table S34) included IL-15–IL15RA/IL2RB/IL2RG and IL-7–IL2RG/IL7R from dendritic cells and macrophages, alongside CCL13–CCR5 and CXCL11–CXCR3 (94–96). Three additional chemokine axes with established experimental support in cytotoxic T cell recruitment, CXCL9–CXCR3, CXCL10–CXCR3, and CCL18–CCR8, fell below LIANA detection thresholds and were retained through custom computation. These are reported as complementary analyses supporting the biological interpretation rather than as directly inferred interactions (tables S35 to S37). Bulk RNA-seq showed IBM-specific upregulation of both APC- and TC-associated interaction genes across conditions (fig. S7, A and D). These inferred APC–TRM interactions are consistent with a microenvironment providing signaling continuity of the kind associated with *ZNF683* upregulation and TRM identity maintenance. This continuity reflects continuous antigen and cytokine-driven input to the local inflammatory niche.

**Fig. 8.**
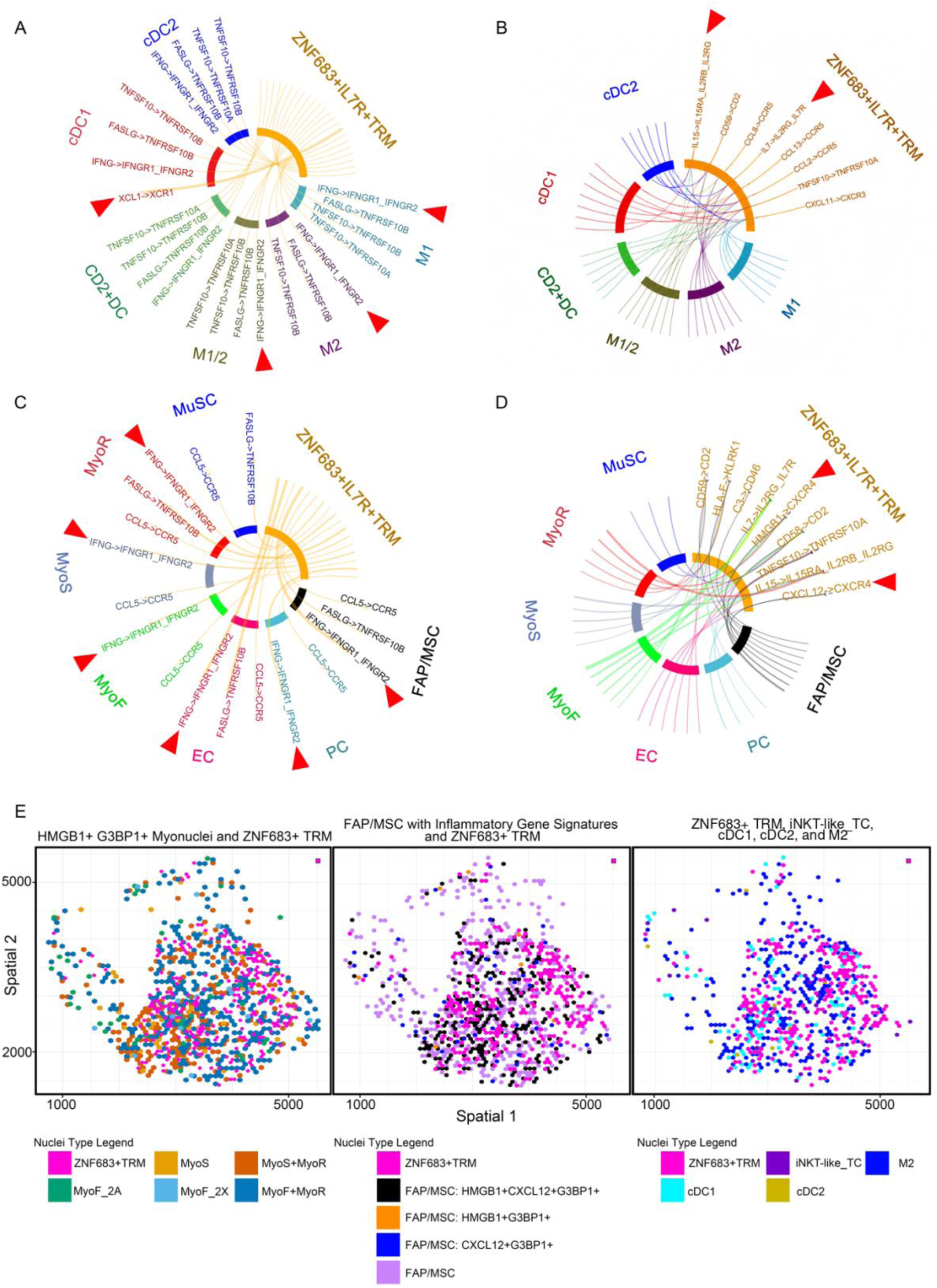
Ligand–receptor inference and spatial transcriptomics reveal the intercellular niche of ZNF683+IL7R+TRM in IBM muscle. **A,** Chord diagram of inferred ligand–receptor interactions with ZNF683+IL7R+TRM as senders and APC subtypes (M1, M2, M1/2, cDC1, cDC2, CD2⁺DC) as receivers. XCL1– XCR1 was uniquely detected in the TRM–cDC1 pairing. **B,** Reciprocal chord diagram with APC subtypes as senders and ZNF683+IL7R+TRM as receivers. Highlighted interactions include IL-15–IL15RA/IL2RB/IL2RG, IL-7–IL2RG/IL7R, and CXCL11–CXCR3. **C,** Inferred interactions with ZNF683+IL7R+TRM as senders and myonuclei and interstitial populations (MyoF, MyoS, MyoR, MuSC, FAP/MSC, EC, PC) as receivers. Highlighted interactions include IFNG–IFNGR1/IFNGR2, CCL5–CCR5, and FASLG–TNFRSF10B. **D,** Reciprocal interactions with myonuclei and interstitial cells as senders and ZNF683+IL7R+TRM as receivers. Highlighted interactions include HMGB1/CXCL12– CXCR4 and IL-15–IL15RA/IL2RB/IL2RG. Line thickness in **A–D** reflects relative interaction prioritization. Interactions in **A–D** were inferred using LIANA. Additional interactions retained through custom computation are reported as complementary analyses in tables S35 to S38. **E,** Spatial transcriptomics maps of IBM muscle showing proximity of ZNF683+TRM to stress-associated myonuclei (HMGB1+G3BP1+), inflammatory FAP/MSC subsets (HMGB1+, CXCL12+, and/or G3BP1+), and immune populations including iNKT-like_TC, cDC1, cDC2, and M2 macrophages.

Inferred interactions from ZNF683+IL7R+ TRM to non-immune cell populations (Fig. 8C; table S33) included IFNG–IFNGR1/IFNGR2, CCL5–CCR5, and FASLG–TNFRSF10B directed toward myofibers and interstitial cells (97–102). IBM myofibers showed transcriptional enrichment of IFN-γ response and endogenous antigen processing and presentation pathways (fig. S8, A and B), consistent with the overall bulk RNA-seq profile (fig. S7C). A reciprocal myogenic and interstitial-to-TRM CCL5–CCR5 axis was additionally retained through custom computation and is reported as a complementary analysis (table S38).

Convergent snRNA-seq and bulk RNA-seq identified upregulation of *GM2A* (a lysosomal lipid-transfer protein supporting CD1d loading), *PLA2G4A* (a phospholipase generating CD1d-accessible lysophospholipid and arachidonic acid-derived substrates), and *ANXA1* (associated with phagocytic uptake signaling) (table S39). This suggests that IBM fast-type myofibers are transcriptionally biased toward generating endogenous lipid antigens loadable onto CD1d and provides a candidate antigenic basis for potential engagement of the ZBTB16+ iNKT-like compartment. Reciprocal signals from myonuclei and interstitial cells to TRM (Fig. 8D) included a HMGB1/CXCL12–CXCR4 axis, in which HMGB1, expressed by stressed myonuclei as a damage-associated molecular pattern, reportedly facilitates CXCL12-mediated CXCR4 signaling through heterocomplex formation, with CXCL12 predominantly expressed by FAP/MSC (103). Additional inferred retention signals included IL-15 and IL-7 from myofibers and stromal populations (97, 98), and HLA-E–KLRK1, a network-level interaction reported in the Supplementary Information. HLA-E upregulation in IBM is consistent with stress-induced non-classical MHC-I surface expression downstream of the IFN-γ response (fig. S7C) and with modulation of KLRK1-associated function in ZNF683+ TRM within a broader feedback response (fig. S7D, table S39).

Together, these bidirectional inferred interactions are consistent with a feed-forward circuit in which TRM-derived IFN-γ contributes to myofiber stress and immune amplification, while stressed myonuclei and FAP/MSC reciprocally support TRM retention through HMGB1/CXCL12–CXCR4.

Analysis of public-domain spatial transcriptomics data comparing IBM, IMNM, and adult controls (8) (Fig. 8E; fig. S9, A to C; tables S40 to S44) showed that *ZNF683*-expressing TRM were specific to IBM in this dataset. *ZNF683* was selected as the defining TRM spatial marker for this population because it was the top differentially expressed gene in IBM TC by snRNA-seq. Transcription factor transcripts are captured with greater efficiency in spatial transcriptomics than surface receptor-encoding genes, making *ZNF683* the more sensitive and specific marker for subcluster-level spatial detection. TRM localized near myonuclei expressing *HMGB1* and *G3BP1* (104), including regeneration-associated myonuclear subtypes (Fig. 8E, left), and near FAP/MSC expressing *HMGB1*, *CXCL12*, and *G3BP1* (Fig. 8E, center).

Spatial proximity of TRM to cDC1, cDC2, and M2 macrophages was additionally observed (Fig. 8E, right), consistent with the inferred TRM–APC interaction networks in Fig. 8, A and B. Quantitative assessment of spatially defined microdomains (table S44) identified discrete tissue regions in which ZNF683+ TRM co-occurred with stress-associated nuclei and stromal support populations. Of 165 TRM-containing microdomains (total area 0.93 mm²), 81 (49.1%; 0.45 mm²) fell within the TRM hotspot zone. The stress myonuclear zone overlapped substantially with this hotspot (Jaccard index=0.640), with 74.9% of the stress KDE zone falling within TRM territory. Stress-containing microdomains accounted for 80.6% (133 of 165) of all TRM-containing microdomains. The FAP/MSC inflammatory zone showed partial overlap (Jaccard index=0.415), and microdomains in which stress-associated nuclei and FAP/MSC were simultaneously present (n=50) defined a triple-niche convergence zone of 0.28 mm². These spatial data indicate that ZNF683+ TRM occupy discrete tissue niches organized around zones of myonuclear stress and stromal CXCL12+ support.

## DISCUSSION

This study describes a spatially organized IBM inflammatory circuit centered on ZNF683+IL7R+CD8+ TRM co-localized with APC, stressed myofibers, and stromal populations. Interaction and spatial analyses suggest it may organize immune persistence (fig. S10). Three observations extend immune-centric descriptions. First, a *ZNF683*/HOBIT-associated stress-resistant effector state may explain the paradox of functionally active T cells co-expressing terminal differentiation markers. Second, CD1d lipid-loading machinery is upregulated in IBM APC and fast-type myofibers, suggesting an endogenous lipid substrate for pathogen-independent engagement of T cells with an innate cytotoxicity profile. Third, the inferred network suggests a feed-forward retention loop: TRM-derived IFN-γ contributes to myofiber stress, whereas stressed myonuclei and FAP/MSC support TRM localization via HMGB1/CXCL12–CXCR4, consistent with, though not demonstrated to account for, IBM’s treatment-refractory chronicity.

Predominant ZNF683+IL7R+TRM in IBM TC may reconcile KLRG1-expressing T cells (12, 13) with sustained IFN-γ production (16, 17), despite KLRG1’s canonical association with dampened effector responses (14, 15). In Jurkat cells, overexpression of the TRM/NKT tissue-residency master transcriptional regulator *ZNF683*/HOBIT (59) was associated with stronger memory and NK-like cytotoxic signatures but weaker glucocorticoid- and stress-response signatures. KLRG1 co-expression may thus reflect chronic stimulation, not exhaustion, consistent with *ZNF683*/HOBIT-associated retention without effector loss elsewhere (34, 35, 50). Progressive *ZNF683* expression across four independent platforms is consistent with expansion under sustained inflammation. Retained *IL7R* distinguishes it from terminally differentiated states, consistent with a memory survival signature supporting long-term tissue persistence (35, 51). *ZNF683*/HOBIT-associated glucocorticoid-response attenuation raises the possibility of a transcriptional basis for IBM immunotherapy resistance, an association not previously proposed or tested here.

The iNKT-like_TC–ZNF683+IL7R+TRM–NK-like_TC continuum is more consistent with chronic antigen exposure than discrete developmental boundaries. In progressive inflammatory disease, T cell states reflect stimulation duration and intensity rather than naïve lymphoid developmental sequences. Accordingly, a probabilistic continuum better fits these cross-sectional data than a strict lineage model and accords with plasticity in chronically stimulated T cells (53, 54). Protein-level iNKT-like_TC enrichment at earlier inferred states raises the possibility that iNKT-like cells represent an antigen-encounter phase preceding the consolidated tissue-resident effector state.

Data suggest convergent CD1d-associated lipid loading across APC and myofiber compartments. IBM M2 macrophages and cDC2 upregulated *CD1C*, *CD1D*, *IL15*, *IL7*, and TRM-positioning chemokines, a profile more consistent with maintaining resident cytotoxic T cells than new priming and distinct from classical APC-driven priming. IBM fast-type myofibers independently upregulated *GM2A*, *PLA2G4A*, and *ANXA1*, components associated with CD1d lipid loading, raising the possibility that they generate candidate endogenous lipid antigens accessible to ZBTB16+ T cells. This capacity across both cell types is compatible with non-classical presentation of endogenous stress-related lipid antigens independently of exogenous pathogens (18, 19). CD1d restriction was not demonstrated and requires direct testing. If supported, CD1d lipid antigen presentation could also be a therapeutic target.

Interaction and spatial analyses support reframing IBM as a tissue-niche disease. TRM-derived IFN-γ contributes to myofiber stress and HMGB1 from stressed myonuclei facilitates CXCL12–CXCR4-dependent TRM retention, with FAP/MSC the dominant CXCL12 source. The TRM-containing microdomains that contained stress-associated myonuclei and the stress zone that overlapped the TRM hotspot. This organization distinguishes IBM from IMNM and controls, which lack ZNF683+ TRM and niche components.

Ligand–receptor inference is probabilistic, not protein-demonstrated, and spatial proximity does not establish direct contact. However, snRNA-seq captures nuclear transcriptional states in native tissue without dissociation or ex vivo confounding, advantages over protein-based tissue interaction assays lacking single-nucleus spatial resolution. Convergence of inferred interactions, spatial niche organization, and bulk RNA-seq across independent cohorts provides triangulated support, though each line remains correlative.

Limitations include cross-sectional sampling precluding definitive lineage inference, Jurkat overexpression data from a cell-line model rather than in vivo evidence, unconfirmed CD1d restriction in iNKT-like_TC, small protein-validation subgroups (n=2 for ZNF683 immunofluorescence), and public-domain spatial transcriptomics from two IBM samples. Disease-duration subgroup analyses therefore require validation in larger longitudinal cohorts.

Causal perturbation studies remain the next step in testing this model. These findings describe IBM having a spatially organized inflammatory niche centered on *ZNF683*-expressing TRM and nominate IL-15 for TRM survival, the CXCR4–CXCL12/HMGB1 axis for niche retention, and XCL1 within the proposed feed-forward loop as candidate therapeutic targets. *ZNF683* may warrant evaluation as a biomarker of progression or treatment response. Convergence within spatially defined niches makes the model testable and supports evaluating existing agents in a disease that has resisted conventional immunosuppression.

## MATERIALS AND METHODS

### Study Design

This study was designed to define the cellular basis of treatment-refractory inflammation in IBM and to test whether the identified features are IBM-specific relative to other AM subtypes. Frozen diagnostic muscle biopsies from patients with IBM and comparator AM subtypes, diagnosed by muscle pathology, serology, and established classification criteria, were studied alongside adult nondisease controls. All specimens were obtained for diagnostic evaluation at the National Center of Neurology and Psychiatry (NCNP). Written informed consent for research use was obtained from all patients or their legal guardians after the nature and possible consequences of the study were explained, and research use was approved by the NCNP Ethics Committee (approval number B2024-130). Complete materials and methods, protocols, reagents, software versions, and analytical parameters are in the Supplementary Materials and Methods. Antibodies and primers are listed in tables S45 and S46.

## List of Supplementary Materials

Materials and Methods

Figs. S1 to S10

Tables S1 to S46

## Acknowledgments

We would like to thank all of the patients and their attending physician for submitting their samples to the NCNP Medical Genome Center for their muscle pathology diagnosis and their consent for research participation.

## Author contributions

Conceptualization: FVR, SH, SN, IN; Methodology: FVR, SH, MK, JT, SN, IN; Investigation: FVR, SH, MK, JT, SN, IN; Formal analysis: FVR, SH, MK; Data curation: FVR, SH, MK, JT; Visualization: FVR, SH, MK, SN, IN; Funding acquisition: SH, SN, IN; Resources: SH, SN, IN; Project administration: SH, IN; Supervision: SH, IN; Writing – original draft: FVR, SH, MK, JT, SN, IN; Writing – review & editing: FVR, SH, MK, JT, SN, IN.

## Ethics

All patients provided informed consent for using their samples for research after the diagnosis. This study was approved by the ethical committees of the NCNP (approval number: B2024-130).

## Data and materials availability

Repository deposition of the single-nucleus and bulk RNA-seq data generated in this study and the analysis code is planned following approval for journal submission. Accession numbers and code links will be added after deposition is complete. Third-party datasets analyzed in this study are available from GEO (GSE241091) for the Jurkat *ZNF683*-overexpression experiment and from the associated interactive cell browser (https://muscle-ibm.cells.ucsc.edu) for the inflammatory muscle disease single-nucleus and spatial transcriptomics data, deposited by the original authors at the European Genome-phenome Archive (accession EGAS50000000310) and the Human Cell Atlas Data Coordination Platform (project UUID d5c91e92-2e7f-473d-8cf3-ab03bbae21c2). Any additional information is available from the corresponding author upon reasonable request.

