## Supplementary Materials for "ZNF683-positive tissue-resident memory T cells organize treatment-refractory niches with targetable circuits in inclusion body myositis"

**This document includes:**

Materials and Methods

Figs. S1 to S10

Tables S1 to S46

### SUPPLEMENTARY MATERIALS AND METHODS

#### Study design

This study characterized the cellular and molecular basis of immune persistence in inclusion body myositis (IBM) by integrating single-nucleus RNA sequencing (snRNA-seq), bulk RNA-seq, reverse transcription quantitative polymerase chain reaction (RT-qPCR), RNA in situ hybridization, immunofluorescence, analysis of publicly available *ZNF683*-overexpression RNA-seq, and spatial transcriptomics. Patients with IBM and comparator autoimmune myositis (AM) subtypes were diagnosed by muscle pathology, serology, and established classification criteria. Adult nondisease controls were included. Frozen diagnostic muscle biopsies were used for tissue-based assays. The sections below provide the complete cohort, ethics, experimental, computational, statistical, reagent, and data-source specifications supporting the abbreviated study-design summary in the main manuscript.

#### Human tissue preparation and ethics

All human muscle biopsy specimens were obtained for diagnostic evaluation at the National Center of Neurology and Psychiatry (NCNP). Tissues were immediately snap-frozen according to the standardized institutional protocol. Samples collected at the NCNP and those referred from other institutions followed NCNP-recommended procedures for biopsy, freezing, and transport. Written informed consent for use of patient data and biopsy material in diagnostic and research applications was obtained from all participants or their legal guardians. Research use was approved by the NCNP Ethics Committee (approval number B2024-130), and all procedures adhered to institutional and ethical standards for human sample use.

#### snRNA-seq

snRNA-seq was performed as previously described (106). Muscle tissues stored at −80°C were sectioned (50 µm, 20 to 30 slices) and homogenized on ice in 4 ml of cold lysis buffer containing Tris-HCl (pH 7.4), NaCl, $MgCl_2$, NP-40, and nuclease-free water and 9 ml of cold BSA wash buffer containing BSA, RNase inhibitor (TOYOBO, SIN-201), and PBS. Homogenization used a Dounce homogenizer attached to a Wheaton Overhead Stirrer (10 strokes, speed 3), followed by sequential filtration through 100- and 40-µm strainers. Nuclei were labeled with 7-AAD, incubated on ice for 15 min, resuspended in 350 µl of cold wash buffer, and sorted using an ARIA III sorter with BD FACSDiva Software 8.0.1. Libraries were prepared with the Chromium Next GEM Single Cell 3′ Reagent Kit v3.1, Dual Index (10x Genomics), according to the manufacturer's protocol. cDNA quality and quantity were assessed with the Agilent High Sensitivity DNA Kit and 2100 Bioanalyzer. Fragment sizes ranged from 200 to 9000 base pairs and peaked at approximately 450 to 500 base pairs. Paired-end, dual-index sequencing was performed on the NovaSeq 6000, and data were processed with Cell Ranger 7.0.1 against GRCh38.

**Single-nucleus computational analysis**

Analyses were conducted in R (v4.5.1) within RStudio (v2023.12.1). Clustering, differential expression, and visualization used Seurat v5.0 (107) with 30 principal components at resolution 1.0. T cell (TC), antigen-presenting cell (APC), fast-type myofiber (MyoF), slow-type myofiber (MyoS), regenerating myofiber (MyoR), muscle stem cell (MuSC), fibro-adipogenic precursor/mesenchymal stromal cell (FAP/MSC), endothelial cell (EC), and pericyte (PC) compartments were subclustered after high-resolution reclustering, and gene signatures were examined across clustering resolutions and dataset subsets. Differential expression was performed by nuclei type across conditions. Pathway enrichment used Metascape (108). Output figures were reillustrated for layout consistency without changing the underlying enrichment results. Trajectory analysis used diffusion component analysis (109) and Monocle3 pseudotime inference (110), and co-expression visualization used dittoSeq (111). Comparative expression plots used weighted, scaled expression per gene per subcluster to reduce distortion from rare populations. Volcano plots were generated with ggplot2 (112), plotting $\log_2$ fold change against $-\log_{10}$(adjusted $p$ value), with thresholds of $|\log_2$ fold change$| > 1$ and adjusted $p < 0.05$. Prespecified genes of interest were labeled with ggrepel. Figures were exported as high-resolution TIFF files.

**Ligand–receptor interaction inference**

Ligand–receptor interactions were inferred with LIANA (113). Interactions were retained when ligand and receptor were expressed in at least 5% of the respective sender and receiver nuclei, the pair was within the top 50% of ranked interactions, the aggregate linkage score was >0.75, and $p < 0.05$ across LIANA inference methods. Sender–receiver directionality was defined a priori, and immune–immune and immune–tissue interactions were analyzed separately. Results were interpreted as inferred communication potential rather than direct evidence of functional signaling.

Certain experimentally established chemokine–receptor axes involved in cytotoxic or effector T cell recruitment were not returned by LIANA in the sparse single-nucleus data: the APC-to-TRM axes CXCL9–CXCR3, CXCL10–CXCR3, and CCL18–CCR8 and the myogenic/interstitial-to-TRM axis CCL5–CCR5. The reciprocal ZNF683+IL7R+TRM-to-myogenic/interstitial CCL5–CCR5 interaction was returned directly by LIANA and is reported as an inferred interaction in table S33. For axes not returned by LIANA, interaction weights were calculated as the product of normalized ligand expression in the sender and receptor expression in the receiver from the Seurat object and scaled to the range of LIANA-inferred weights using IFNG–IFNGR1/2 and FASLG–TNFRSF10A/B as reference interactions. Supplemented interactions were flagged throughout filtering and visualization and were interpreted only as complementary analyses supporting biological interpretation. Experimental support for each supplemented pair is provided in tables S35 to S38.

**Bulk RNA-seq**

Approximately 100 frozen muscle sections of 10 μm thickness were used for total RNA extraction with either the PureLink RNA Mini Kit with TRIzol (Thermo Fisher Scientific) or the RNeasy Mini Kit (Qiagen). The two procedures yielded comparable results. RNA concentration was measured with a NanoDrop spectrophotometer. Library preparation and sequencing were performed by BGI. Total RNA underwent ribosomal RNA depletion or oligo(dT) selection, followed by reverse transcription, end repair, adapter ligation, and polymerase chain reaction amplification. Sequencing used DNBSEQ, BGISEQ, or Illumina HiSeq platforms. FASTQ quality control used FastQC. Reads were aligned to GRCh38.p13 with comprehensive annotation using Rsubread (114), counted with featureCounts (115), and analyzed for differential expression with DESeq2 (116) using apeglm shrinkage (117). Variance-stabilizing transformation was applied before visualization. The within-IBM disease-duration comparison used *ZNF683* counts per million (CPM), grouping samples as $\leq 24$ versus $>24$ months (Fig. 4G; table S15).

**RT-qPCR and statistical analysis**

Total RNA was extracted with the RNeasy Mini Kit and reverse transcribed with SuperScript IV VILO Master Mix. Relative expression was calculated by the $-\Delta\Delta$Ct method, corresponding to $\log_2$ fold-change values. Because these values were nonnormally distributed, comparisons across conditions used the Kruskal–Wallis rank-sum test followed by pairwise Mann–Whitney (Wilcoxon rank-sum) tests with Benjamini–Hochberg correction (table S17). For the IBM disease-duration comparison, the two-sided Welch t-test was recalculated as a consistency check directly from the 11 $log_2FC$ values supplied in table S18, using the shorter-minus-longer group contrast (≤24 versus >24 months).

ROC–AUC analysis compared IBM (n = 11) with all non-IBM samples in table S16 (n = 54: 48 other AM, 4 VCP-M, and 2 nondisease adult controls). Figure 4J and table S19 also include every other condition, each compared with all remaining samples. Membership in the named condition defined the outcome for each curve. RT-qPCR $log_2FC$ values were coded as 0 for ≤200 and 1 for >200, with the higher score treated as positive throughout. Mild (>50), moderate (>100), and marked (>200) thresholds remained descriptive expression labels. AUC was calculated from case–comparator sample pairs for each condition, assigning half credit to tied scores. The DeLong variance was used for normal-approximation 95% confidence intervals, with bounds constrained to 0 to 1. Calculations used NumPy 2.3.5 and SciPy 1.17.0, and AUC was independently verified with scikit-learn 1.8.0. The IBM binary analysis yielded AUC = 0.900, 95% DeLong CI = 0.779–1.000 (upper bound capped at 1), sensitivity = 81.818% (9/11), and specificity = 98.148% (53/54) (table S19). No classifier fitting, feature selection, or training/test split was used. Primer sequences are provided in table S46.

**Immunofluorescence**

Frozen muscle sections (8 μm) were fixed in 4% paraformaldehyde in PBS (pH 7.6) or acetone/methanol at −20°C, blocked with 3% BSA-PBS for 1 hour, incubated with primary antibodies for 12 to 16 hours at 4°C, followed by secondary antibodies for 1 hour at room temperature, and counterstained with DAPI (1:1000, FUJIFILM Wako Pure Chemical Corporation) for 5 min. Images were acquired with a Keyence fluorescence microscope. Antibody information is provided in table S45.

**RNA in situ hybridization**

RNA in situ hybridization used the RNAscope Multiplex Fluorescent Reagent Kit v2 (Advanced Cell Diagnostics) according to the standard protocol. Pretreatment used the HybEZ Humidifying System with the ACD EZ-Batch Slide Holder and Wash Tray. A *ZNF683* C2 probe diluted with Probe Diluent (catalog no. 30041) was detected with TSA Vivid Fluorophore 520 (catalog no. 323271, 1:750 in TSA buffer), followed by CD8, p62, and DAPI counterstaining. Images were acquired with a Keyence fluorescence microscope. Antibody information is provided in table S45.

**Jurkat cell line analysis**

RNA-seq data from *ZNF683*-overexpressing Jurkat cells were obtained from GEO (GSE241091) (35). Raw gene-level read counts were analyzed as provided. Differential expression used a Poisson generalized linear model with library size as an offset to account for sequencing-depth differences. Coefficients for the condition term were used to estimate $\log_2$ fold changes. Genes with $|\log_2 \text{ fold change}| \geq 1$ and false discovery rate $<0.05$ were retained for pathway enrichment with Metascape (108). For predefined memory T cell, natural killer cell, and stress-response gene sets, z-score scaling with $|z| \geq 1.5$ was applied after differential-expression analysis to display consistent expression shifts. Z-scores were not used to define differential expression.

**Spatial transcriptomics analysis**

Publicly available single-nucleus and spatial transcriptomics data from IBM, immune-mediated necrotizing myopathy, and adult control muscle were obtained from the associated interactive cell browser (https://muscle-ibm.cells.ucsc.edu) (8). Processed data were analyzed directly to preserve the original authors' cell-type assignments and analytical parameters. Cell and nuclear states were mapped to spatial coordinates using snRNA-seq-derived expression signatures without unsupervised reclustering. TRM were identified by *ZNF683* and *CD247* co-expression. Stress-associated myonuclei were identified by *HMGB1* and *G3BP1* co-expression. Inflammatory stromal populations were identified by *PDGFRA* co-expressed with *HMGB1* and/or *CXCL12* (table S41).

Spatial coordinates were aggregated into hexagonal microdomains with a fixed bin size held constant across samples. Kernel density estimation (KDE) was applied to microdomain coordinates weighted by population abundance, using a Gaussian kernel with Silverman bandwidth on a uniform grid spanning the tissue coordinate range. Enrichment hotspot zones were defined at the 90th percentile of positive KDE density and core zones at the 95th percentile. Spatial overlap was quantified with the Jaccard index. Analyses assessed co-occurrence and proximity of transcriptionally defined populations and did not infer direct cell–cell contact, ligand–receptor signaling, or causal interactions. Spatial analyses used Seurat v5.0 (107) in R (v4.5.2).

**Computational analysis specifications**

Analyses were conducted at multiple time points, and R versions were updated when required by specific pipelines. Stable release versions of the corresponding packages available at the time of each analysis were used. The planned availability of study-generated data and analysis code is described in the Data and materials availability statement in the main manuscript. Repository deposition is deferred until approval for journal submission.

### SUPPLEMENTARY FIGURES

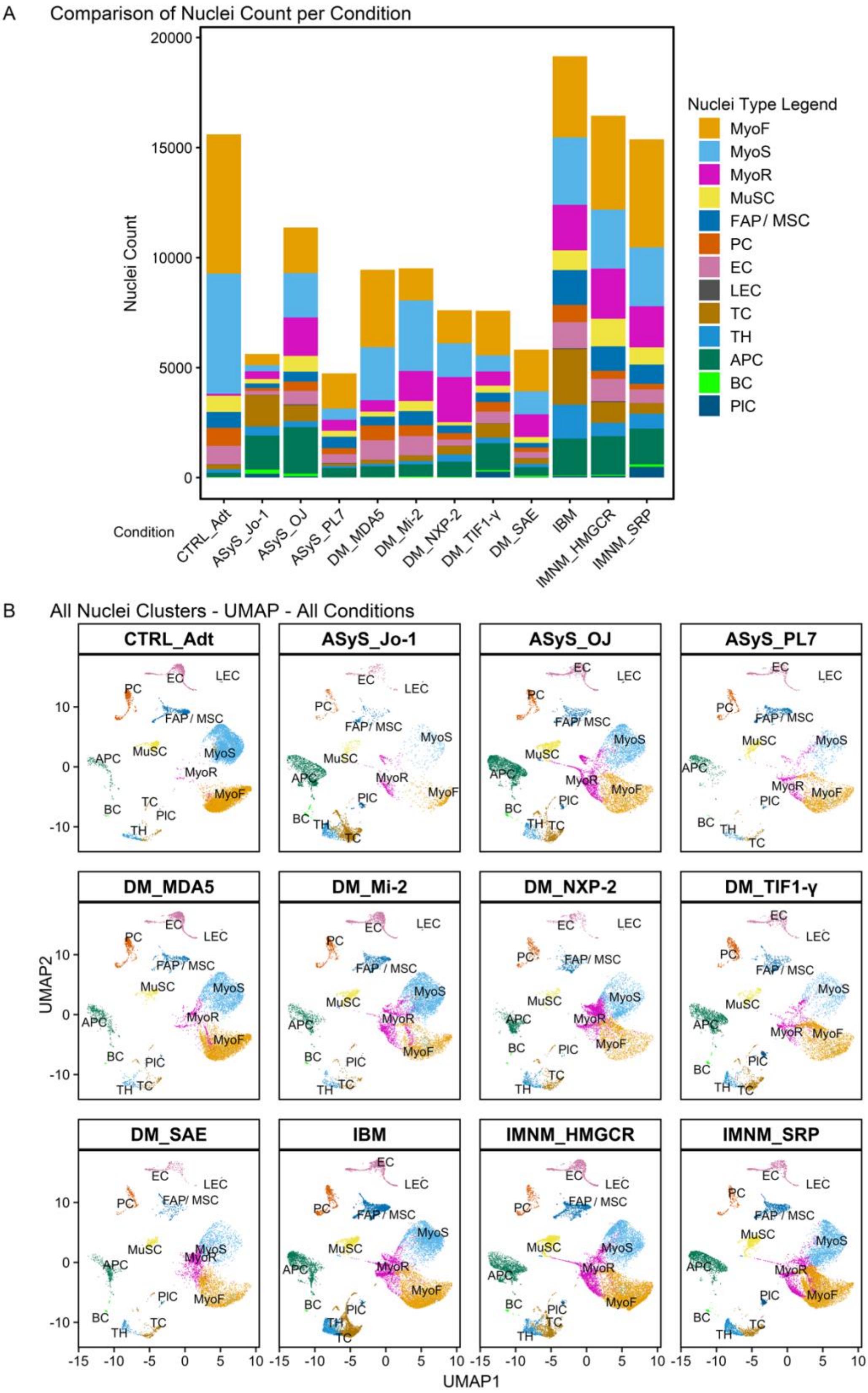

**Fig. S1. Nuclei composition and global clustering across conditions.**

**(A)** Stacked bar plot showing the total number of nuclei assigned to each major cell and nuclear type across all conditions, illustrating differences in cellular composition between autoimmune myopathy subtypes and adult control muscle. **(B)** UMAP visualization of all nuclei clusters separated by condition, showing the global distribution and relative organization of major muscle, immune, endothelial, and interstitial cell populations across conditions.

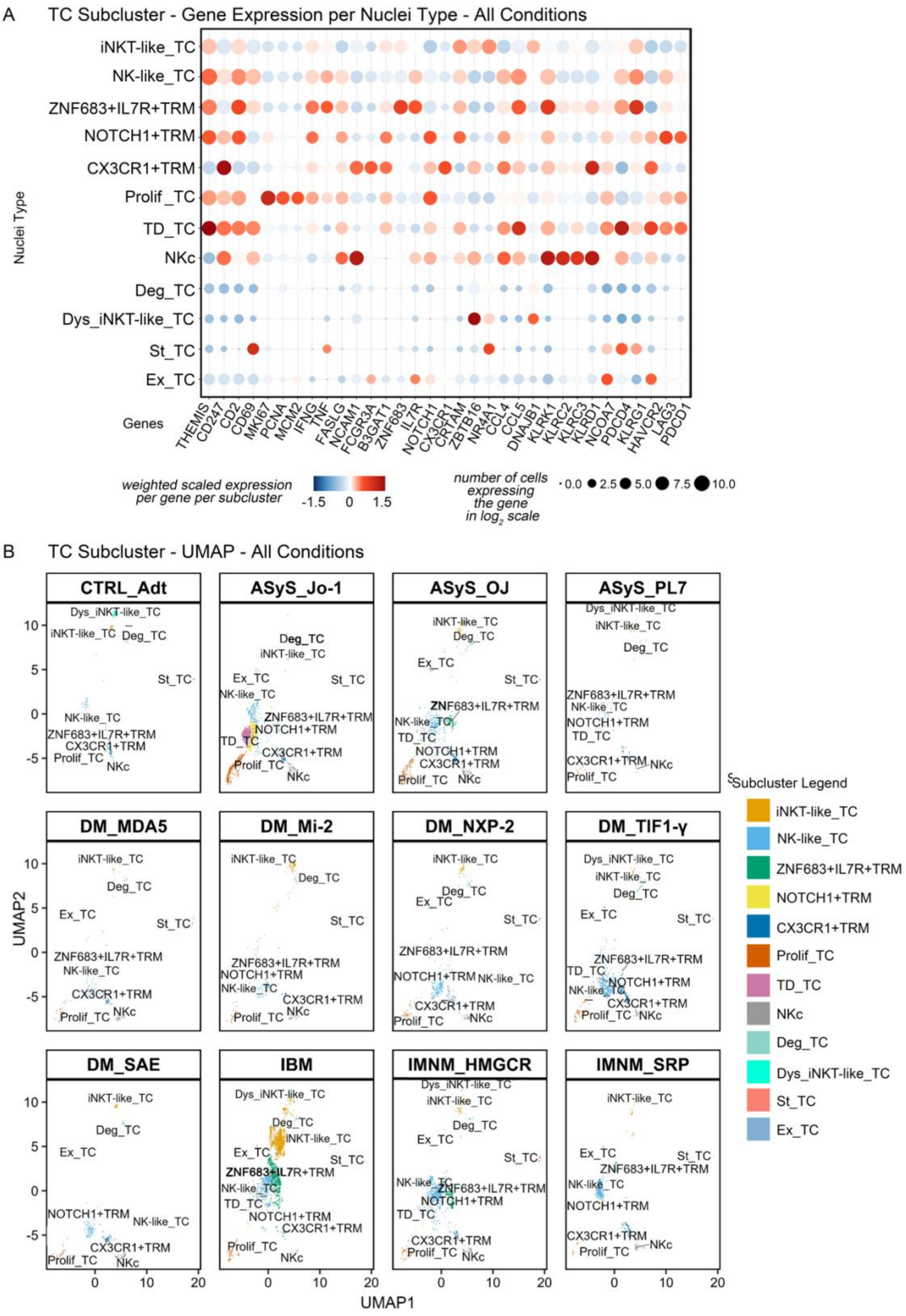
A
TC Subcluster - Gene Expression per Nuclei Type - All Conditions
Nuclei Type
iNKT-like_TC
NK-like_TC
ZNF683+IL7R+TRM
NOTCH1+TRM
CX3CR1+TRM
Prolif_TC
TD_TC
NKc
Deg_TC
Dys_iNKT-like_TC
St_TC
Ex_TC
Genes
THEMIS CD247 CD2 CD69 MKI67 PCNA MCM2 IFNG TNF FASLG NCAM1 FCGR3A B3GAT1 ZNF683 IL7R NOTCH1 CX3CR1 CRTAM ZBTB16 NR4A1 CCL4 CCL5 DNAJB1 KLRK1 KLRC2 KLRC3 KLRD1 NCOA7 PDCD4 KLRG1 HAVCR2 LAG3 PDCD1
weighted scaled expression per gene per subcluster
-1.5 0 1.5
number of cells expressing the gene in log2 scale
0.0 2.5 5.0 7.5 10.0
B
TC Subcluster - UMAP - All Conditions
CTRL_Adt
ASyS_Jo-1
ASyS_OJ
ASyS_PL7
DM_MDA5
DM_Mi-2
DM_NXP-2
DM_TIF1-γ
DM_SAE
IBM
IMNM_HMGCR
IMNM_SRP
UMAP2
UMAP1
Subcluster Legend
iNKT-like_TC
NK-like_TC
ZNF683+IL7R+TRM
NOTCH1+TRM
CX3CR1+TRM
Prolif_TC
TD_TC
NKc
Deg_TC
Dys_iNKT-like_TC
St_TC
Ex_TC

**Fig. S2. Transcriptional features and distribution of TC subclusters across autoimmune myopathies and adult control.**

**(A)** Dot plot showing weighted, scaled expression of selected T cell-associated genes across transcriptionally defined TC subclusters. Dot color indicates relative expression level and dot size reflects the proportion of nuclei expressing each gene ($log_2$ scale). **(B)** UMAP visualization of TC subclusters separated by condition, illustrating the relative distribution and abundance of iNKT-like_TC, NK-like_TC, ZNF683+IL7R+TRM, and additional TC states across AM subtypes and adult control.

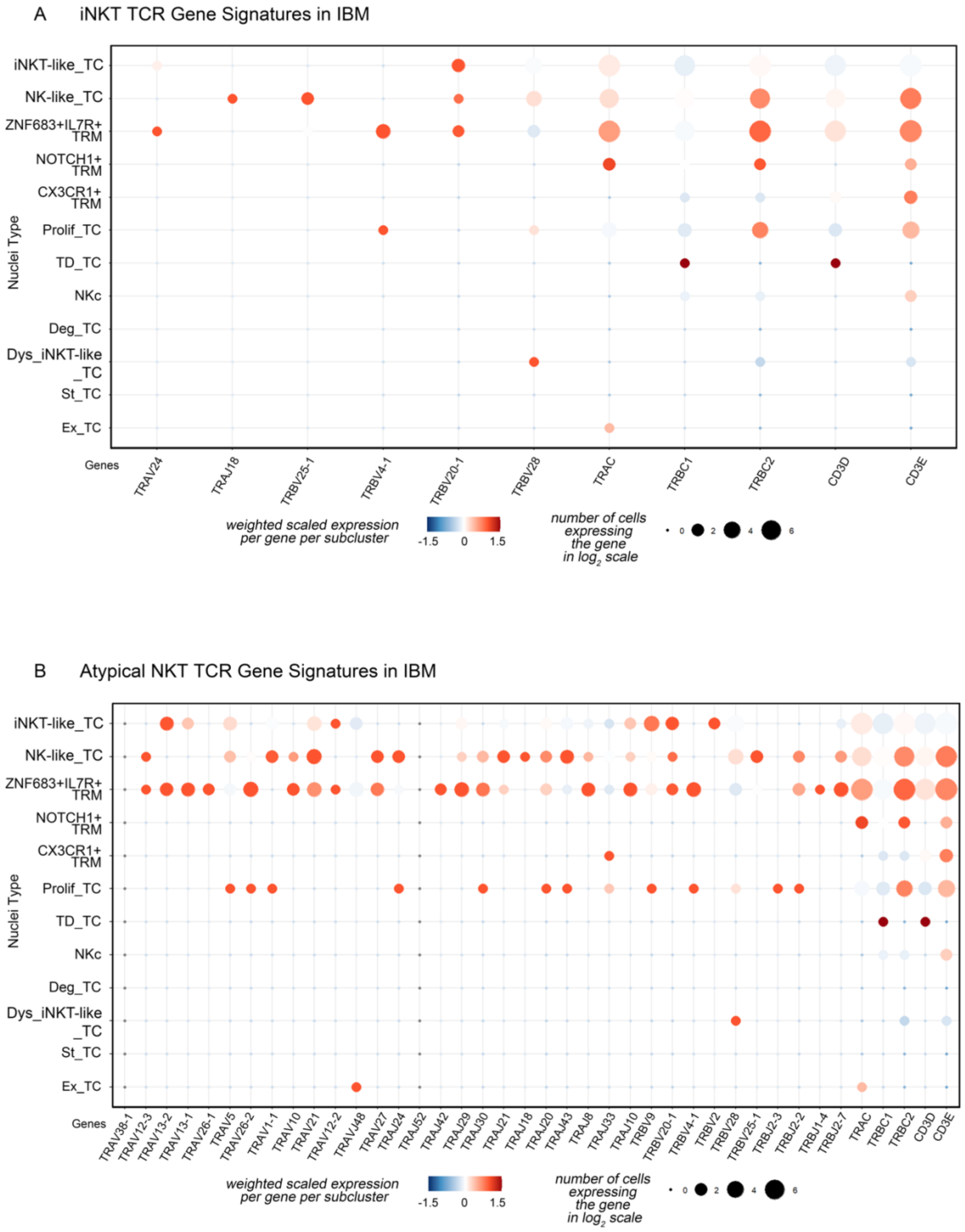
A iNKT TCR Gene Signatures in IBM
iNKT-like_TC
NK-like_TC
ZNF683+IL7R+ TRM
NOTCH1+ TRM
CX3CR1+ TRM
Prolif_TC
TD_TC
NKc
Deg_TC
Dys_iNKT-like _TC
St_TC
Ex_TC
Nuclei Type
Genes
TRAV24
TRAJ18
TRBV25-1
TRBV4-1
TRBV20-1
TRBV28
TRAC
TRBC1
TRBC2
CD3D
CD3E
weighted scaled expression per gene per subcluster
-1.5 0 1.5
number of cells expressing the gene in $log_2$ scale
0 2 4 6
B Atypical NKT TCR Gene Signatures in IBM
iNKT-like_TC
NK-like_TC
ZNF683+IL7R+ TRM
NOTCH1+ TRM
CX3CR1+ TRM
Prolif_TC
TD_TC
NKc
Deg_TC
Dys_iNKT-like _TC
St_TC
Ex_TC
Nuclei Type
Genes
TRAV38-1
TRAV12-3
TRAV13-2
TRAV13-1
TRAV26-1
TRAV5
TRAV26-2
TRAV1-1
TRAV10
TRAV21
TRAV12-2
TRAVJ48
TRAJ27
TRAJ24
TRAJ52
TRAJ42
TRAJ29
TRAJ30
TRAJ21
TRAJ18
TRAJ20
TRAJ43
TRAJ8
TRAJ33
TRAJ10
TRBV9
TRBV20-1
TRBV4-1
TRBV2
TRBV28
TRBV25-1
TRBJ2-3
TRBJ2-2
TRBJ1-4
TRBJ2-7
TRAC
TRBC1
TRBC2
CD3D
CD3E
weighted scaled expression per gene per subcluster
-1.5 0 1.5
number of cells expressing the gene in $log_2$ scale
0 2 4 6

**Fig. S3. TCR-associated gene signatures for NKT lineages in IBM.**

**(A)** Dot plot of weighted, scaled expression of T cell receptor genes from canonical iNKT gene sets across TC subclusters, showing enrichment of iNKT-associated *TRAV* and *TRBV* segments in iNKT-like_TC and ZNF683+IL7R+TRM. (**B)** Dot plot of weighted, scaled expression of T cell receptor genes from atypical NKT gene sets across TC subclusters. ZNF683+IL7R+TRM and NK-like_TC show the broadest expression across atypical NKT TCR segments, supporting shared NKT repertoire features across the transcriptional continuum. Dot color indicates weighted scaled expression per gene per subcluster. Dot size reflects the proportion of nuclei expressing each gene ($\log_2$ scale).

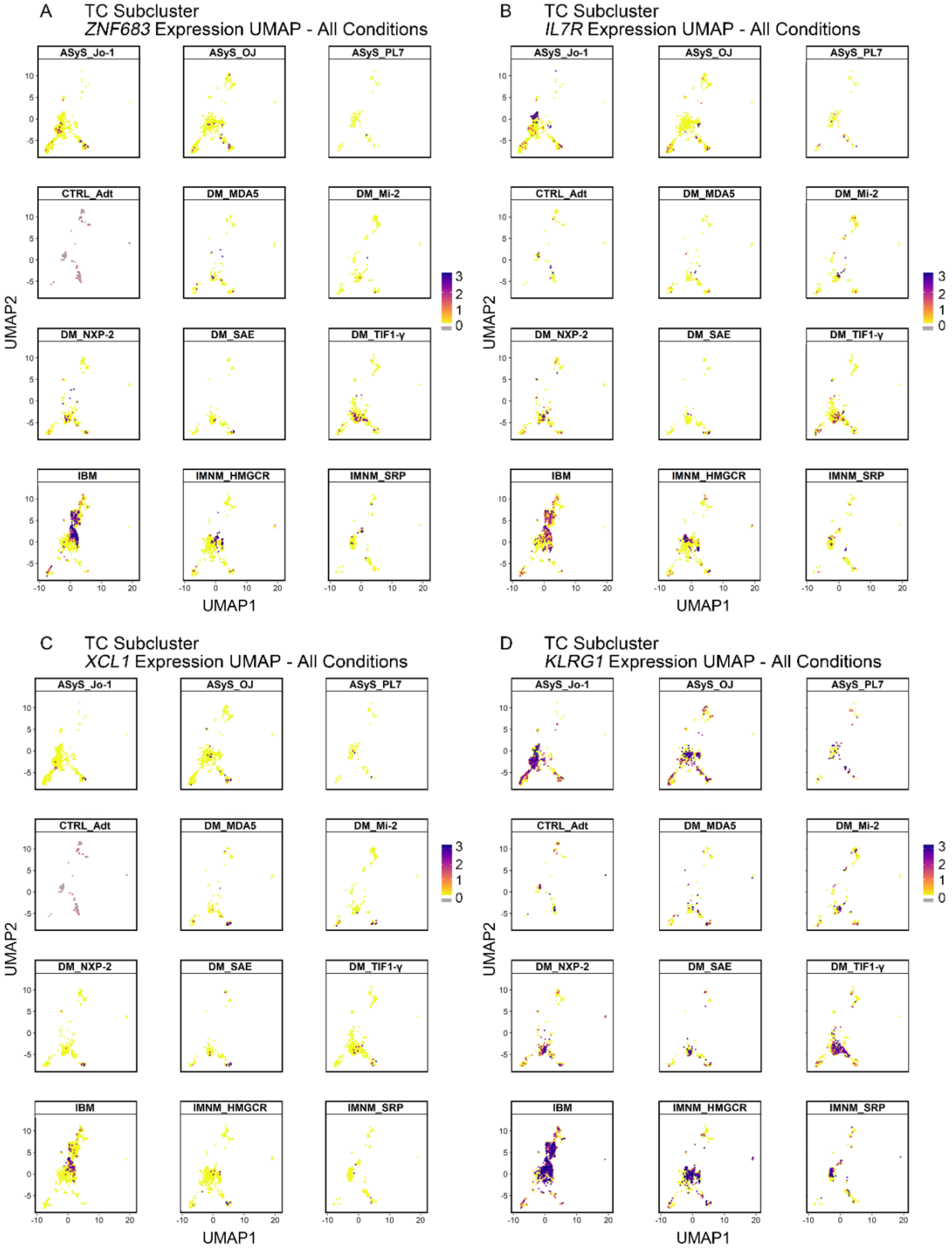
A
TC Subcluster
*ZNF683* Expression UMAP - All Conditions
B
TC Subcluster
*IL7R* Expression UMAP - All Conditions
C
TC Subcluster
*XCL1* Expression UMAP - All Conditions
D
TC Subcluster
*KLRG1* Expression UMAP - All Conditions
ASyS_Jo-1
ASyS_OJ
ASyS_PL7
CTRL_Adt
DM_MDA5
DM_Mi-2
DM_NXP-2
DM_SAE
DM_TIF1-γ
IBM
IMNM_HMGCR
IMNM_SRP
UMAP1
UMAP2

**Fig. S4. TC subcluster key gene expression across conditions.**

**(A–C)** UMAP feature plots demonstrating increased density and expression intensity of *ZNF683*, *IL7R*, and *XCL1* in IBM relative to all other conditions. **(D)** *KLRG1* expression was broadly distributed across conditions without IBM-specific enrichment.

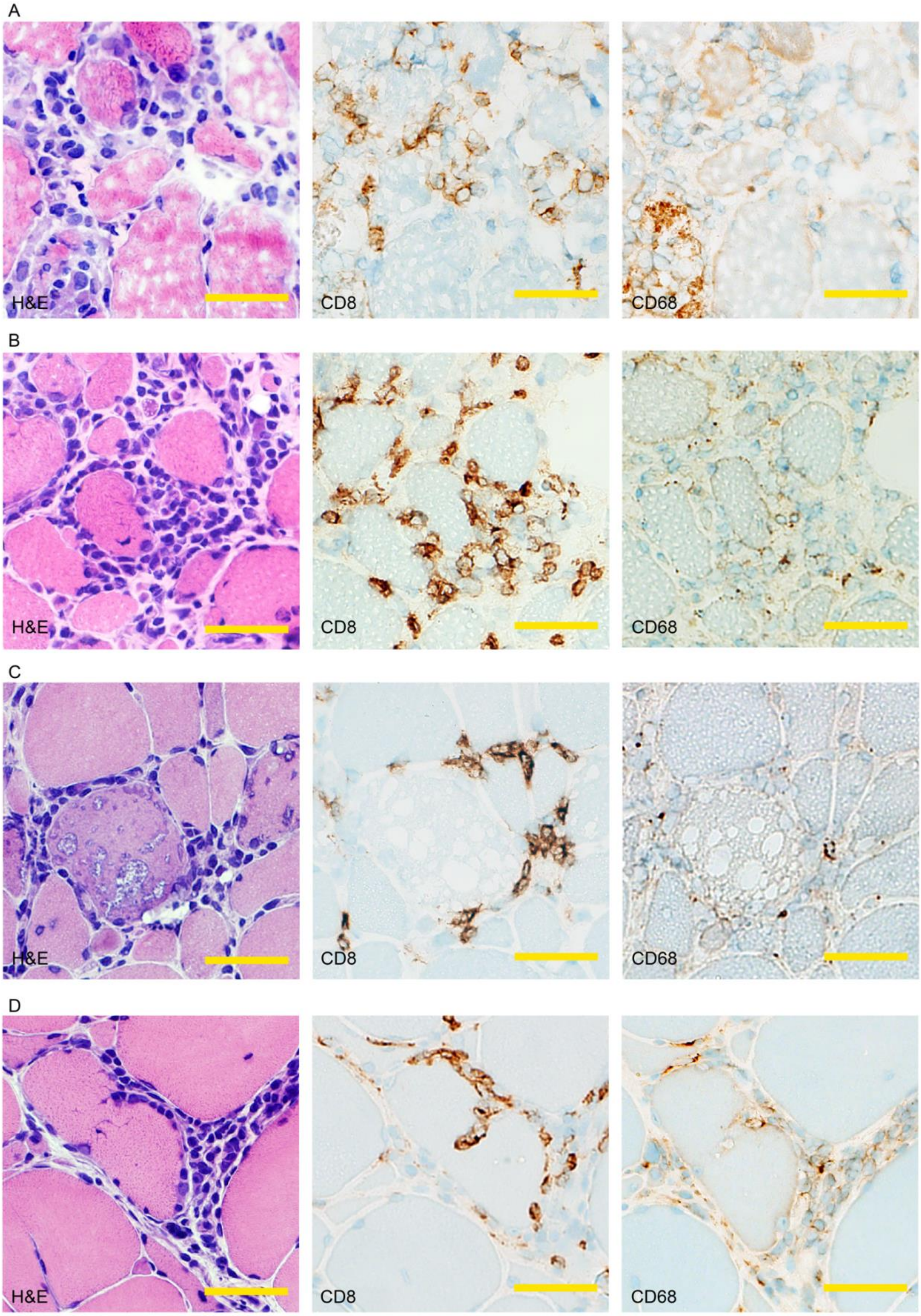


yellow scale bar - 50 µm

**Fig. S5. Cytotoxic T cell infiltration persists across disease duration in IBM.**

**(A–D)** Representative H&E, CD8, and CD68 immunohistochemistry of IBM muscle biopsies at disease durations of 11 months (A), 60 months (B), 96 months (C), and 108 months (D). Scale bar, 50 μm.

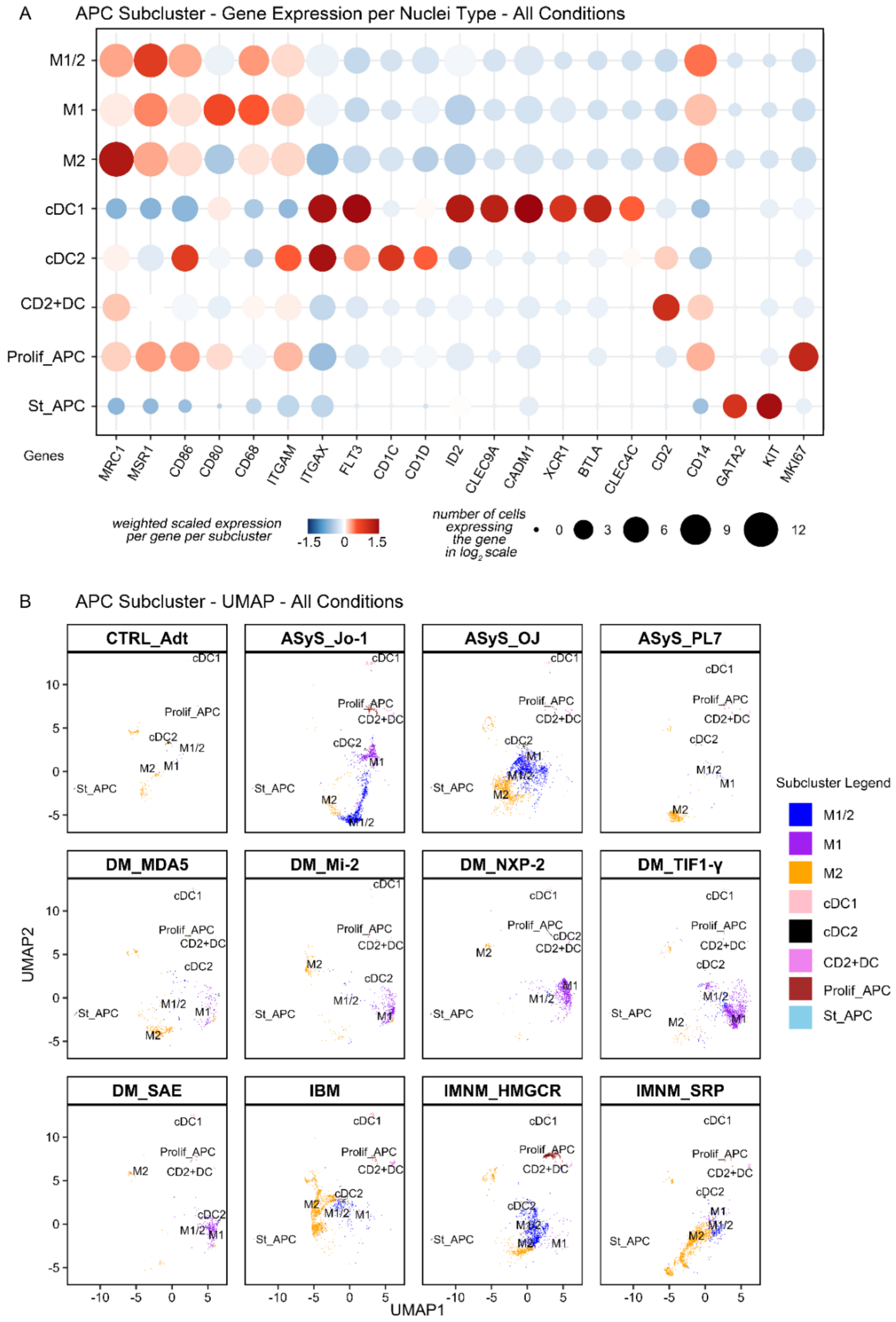
A
APC Subcluster - Gene Expression per Nuclei Type - All Conditions
M1/2
M1
M2
cDC1
cDC2
CD2+DC
Prolif_APC
St_APC
Genes
MRC1
MSR1
CD86
CD80
CD68
ITGAM
ITGAX
FLT3
CD1C
CD1D
ID2
CLEC9A
CADM1
XCR1
BTLA
CLEC4C
CD2
CD14
GATA2
KIT
MKI67
weighted scaled expression per gene per subcluster
-1.5 0 1.5
number of cells expressing the gene in $log_2$ scale
0 3 6 9 12
B
APC Subcluster - UMAP - All Conditions
CTRL_Adt
ASyS_Jo-1
ASyS_OJ
ASyS_PL7
DM_MDA5
DM_Mi-2
DM_NXP-2
DM_TIF1-γ
DM_SAE
IBM
IMNM_HMGCR
IMNM_SRP
UMAP2
UMAP1
Subcluster Legend
M1/2
M1
M2
cDC1
cDC2
CD2+DC
Prolif_APC
St_APC

**Fig. S6. APC subcluster composition across conditions.**

**(A)** Gene signature-based comparison of APC nuclei across 11 autoimmune myopathy subtypes and adult controls following dataset integration, confirming *XCR1* restriction to cDC1 across all conditions. (**B)** UMAP visualization of APC subclusters stratified by condition after integration.

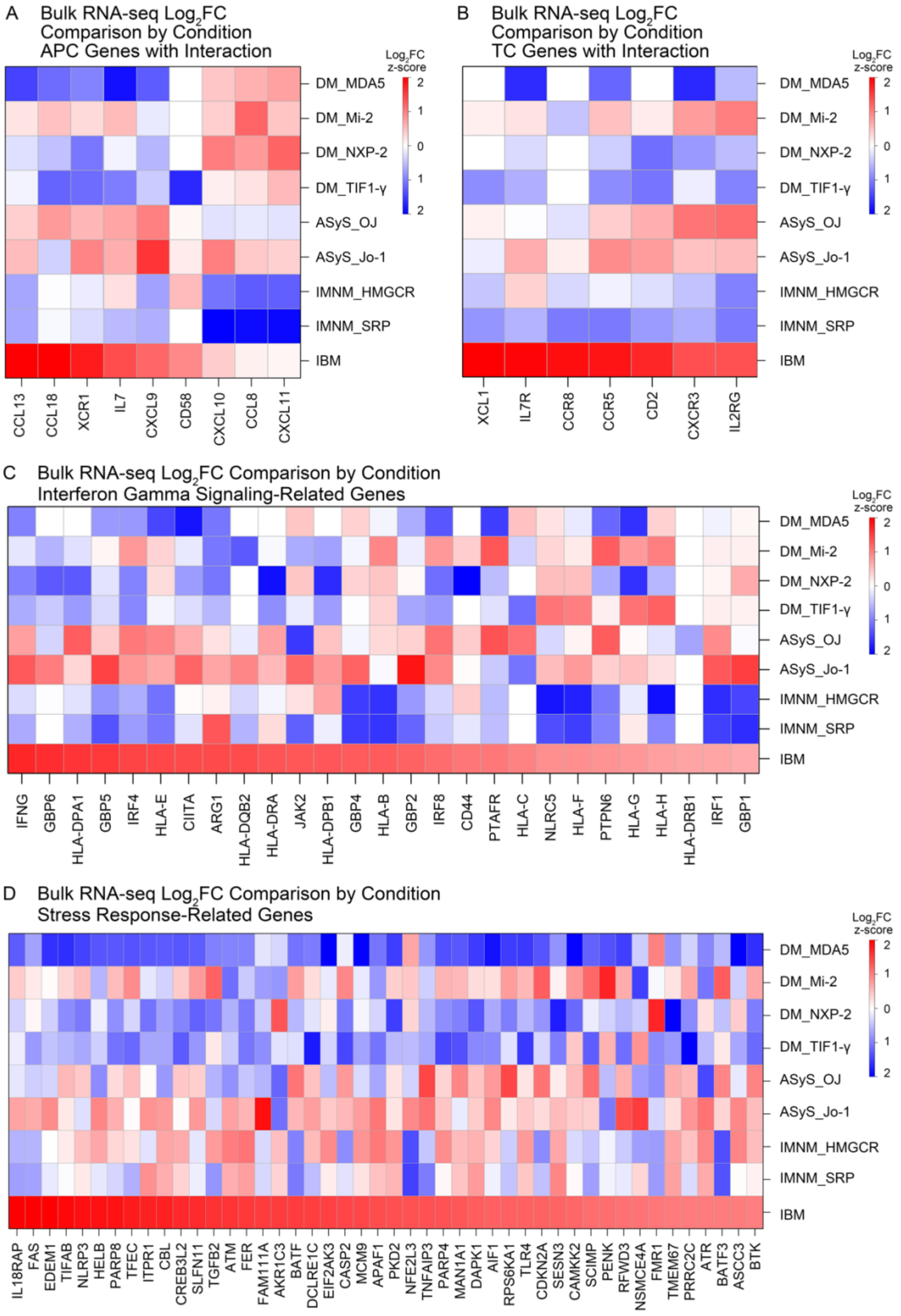

A
Bulk RNA-seq Log2FC Comparison by Condition APC Genes with Interaction
B
Bulk RNA-seq Log2FC Comparison by Condition TC Genes with Interaction
C
Bulk RNA-seq Log2FC Comparison by Condition Interferon Gamma Signaling-Related Genes
D
Bulk RNA-seq Log2FC Comparison by Condition Stress Response-Related Genes
Log2FC z-score
2
1
0
1
2
DM_MDA5
DM_Mi-2
DM_NXP-2
DM_TIF1-γ
ASyS_OJ
ASyS_Jo-1
IMNM_HMGCR
IMNM_SRP
IBM
CCL13 CCL18 XCR1 IL7 CXCL9 CD58 CXCL10 CCL8 CXCL11
XCL1 IL7R CCR8 CCR5 CD2 CXCR3 IL2RG
IFNG GBP6 HLA-DPA1 GBP5 IRF4 HLA-E CIITA ARG1 HLA-DQB2 HLA-DRA JAK2 HLA-DPB1 GBP4 HLA-B GBP2 IRF8 CD44 PTAFR HLA-C NLRC5 HLA-F PTPN6 HLA-G HLA-H HLA-DRB1 IRF1 GBP1
IL18RAP FAS EDEM1 TIFAB NLRP3 HELB PARP8 TFEC ITPR1 CBL CREB3L2 SLFN11 TGFB2 ATM FER FAM111A AKR1C3 BATF DCLRE1C EIF2AK3 CASP2 MCM9 APAF1 PKD2 NFE2L3 TNFAIP3 PARP4 MAN1A1 DAPK1 AIF1 RPS6KA1 TLR4 CDKN2A SESN3 CAMKK2 SCIMP PENK RFWD3 NSMCE4A FMR1 TMEM67 PRRC2C ATR BATF3 ASCC3 BTK

**Fig. S7. Interaction-associated gene expression patterns in bulk RNA-seq across conditions.**

**(A)** Bulk RNA-seq $log_2FC$ comparison across conditions for APC-associated interaction genes derived from snRNA-seq ligand–receptor analyses. IBM shows the strongest and most consistent upregulation across all nine genes shown, including the chemokines *CCL13*, *CCL18*, and *CXCL9–11*, the dendritic cell marker *CD58*, the T cell survival cytokine *IL7*, and the cDC1-restricted chemokine receptor ligand *XCR1*, relative to all other AM subtypes and controls. IMNM_SRP shows selective upregulation of *CCL8* and *CXCL10* but lacks the breadth of IBM upregulation across the full gene set. **(B)** Bulk RNA-seq $log_2FC$ comparison across conditions for TC-associated interaction genes derived from snRNA-seq ligand–receptor analyses. IBM shows marked upregulation of all seven genes shown (*XCL1*, *IL7R*, *CCR8*, *CCR5*, *CD2*, *CXCR3*, and *IL2RG*) relative to all other conditions, confirming at the bulk tissue level the TRM-associated receptor and ligand expression identified by snRNA-seq. The breadth and magnitude of upregulation across this gene set is specific to IBM and not observed in any other AM subtype examined. **(C)** Bulk RNA-seq $log_2FC$ comparison across conditions for IFN-γ signaling-related genes. IBM demonstrates broad and coordinated upregulation of canonical IFN-γ response genes including *IFNG*, *GBP5*, *GBP6*, *IRF4*, *HLA-E*, *CIITA*, *ARG1*, *HLA-DQB2*, *HLA-DRA*, *JAK2*, *HLA-DPB1*, *GBP4*, *HLA-B*, *GBP2*, *IRF8*, *CD44*, *PTAFR*, *HLA-C*, *NLRC5*, *HLA-F*, *PTPN6*, *HLA-G*, *HLA-H*, *HLA-DRB1*, *IRF1*, and *GBP1*. The magnitude and breadth of IFN-γ pathway upregulation in IBM substantially exceeds that of all other AM subtypes, consistent with sustained IFN-γ-mediated immune amplification driven by ZNF683+ TRM. ASyS_Jo-1 shows selective upregulation of a subset of HLA class II and JAK-STAT pathway genes, while DM and IMNM subtypes show predominantly downregulated or neutral expression across this gene set.

**(D)** Bulk RNA-seq $\log_2$FC comparison across conditions for stress response-related genes. IBM shows broad upregulation across a diverse set of stress-associated genes including *IL18RAP*, *FAS*, *EDEM1*, *TIFAB*, *NLRP3*, *HELB*, *PARP8*, *TFEC*, *ITPR1*, *CBL*, *CREB3L2*, *SLFN11*, *TGFB2*, *ATM*, *FER*, *FAM111A*, *AKR1C3*, *BATF*, *DCLRE1C*, *EIF2AK3*, *RPS6KA1*, *MCM9*, *PKD2*, *APAF1*, *NFE2L3*, *TNFAIP3*, *PARP4*, *DAPK1*, *MAN1A1*, *AIF1*, *CASP2*, *TLR4*, *CDKN2A*, *SESN3*, *CAMKK2*, *SCIMP*, *PENK*, *NSMCE4A*, *FMR1*, *TMEM67*, *PRRC2C*, *ATR*, *BATF3*, *ASCC3*, *REM1*, and *BTK*, indicating activation of DNA damage response, apoptotic signaling, inflammasome, and cellular stress pathways. This stress-associated transcriptional signature is IBM-specific and is not observed at comparable magnitude in any other AM subtype, consistent with the myofiber stress–TRM feed-forward circuit described in the main text.

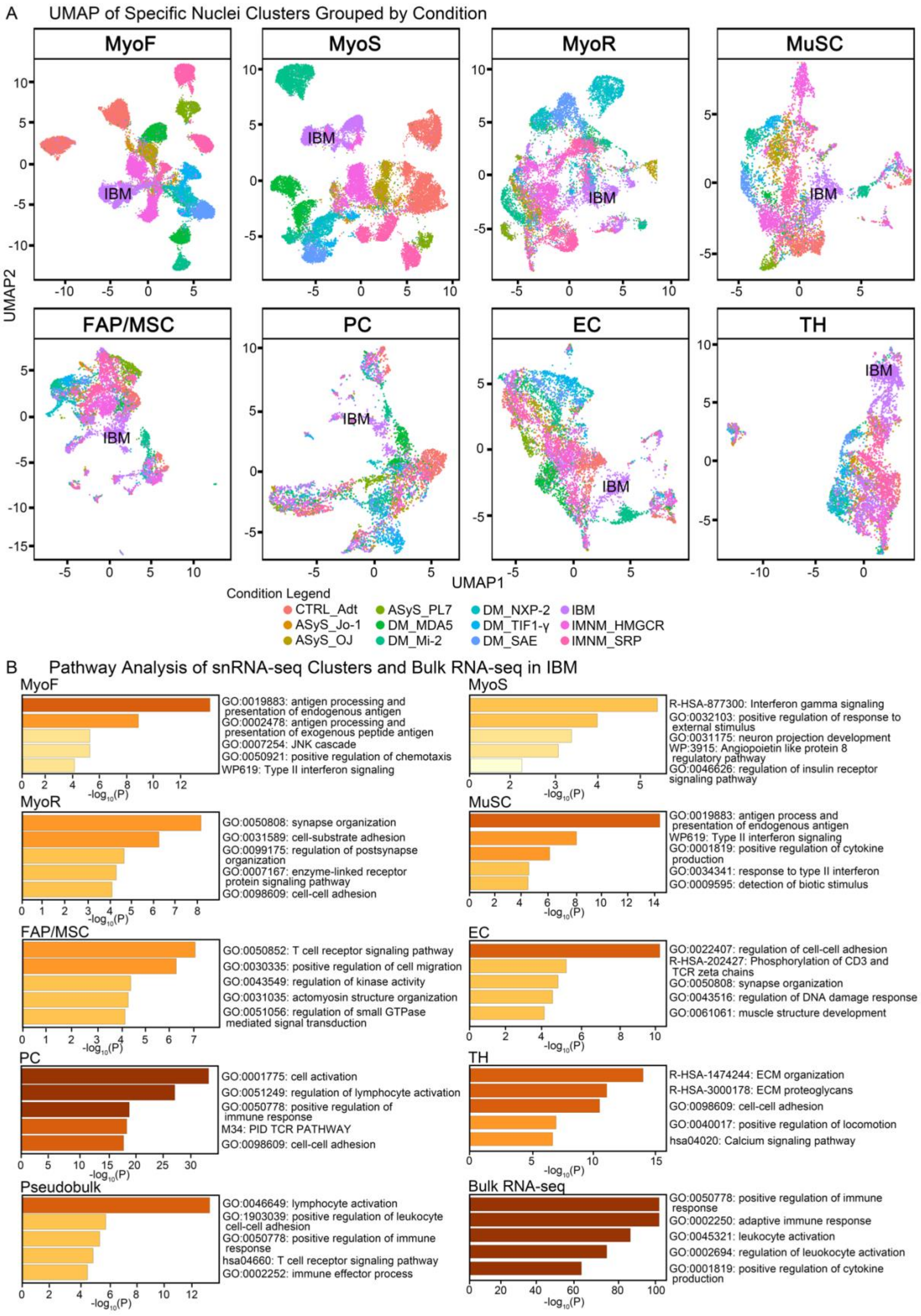
A UMAP of Specific Nuclei Clusters Grouped by Condition
MyoF
MyoS
MyoR
MuSC
FAP/MSC
PC
EC
TH
IBM
UMAP1
UMAP2
Condition Legend
CTRL_Adt
ASyS_Jo-1
ASyS_OJ
ASyS_PL7
DM_MDA5
DM_Mi-2
DM_NXP-2
DM_TIF1-γ
DM_SAE
IBM
IMNM_HMGCR
IMNM_SRP
B Pathway Analysis of snRNA-seq Clusters and Bulk RNA-seq in IBM
MyoF
GO:0019883: antigen processing and presentation of endogenous antigen
GO:0002478: antigen processing and presentation of exogenous peptide antigen
GO:0007254: JNK cascade
GO:0050921: positive regulation of chemotaxis
WP619: Type II interferon signaling
MyoS
R-HSA-877300: Interferon gamma signaling
GO:0032103: positive regulation of response to external stimulus
GO:0031175: neuron projection development
WP:3915: Angiopoietin like protein 8 regulatory pathway
GO:0046626: regulation of insulin receptor signaling pathway
MyoR
GO:0050808: synapse organization
GO:0031589: cell-substrate adhesion
GO:0099175: regulation of postsynapse organization
GO:0007167: enzyme-linked receptor protein signaling pathway
GO:0098609: cell-cell adhesion
MuSC
GO:0019883: antigen process and presentation of endogenous antigen
WP619: Type II interferon signaling
GO:0001819: positive regulation of cytokine production
GO:0034341: response to type II interferon
GO:0009595: detection of biotic stimulus
FAP/MSC
GO:0050852: T cell receptor signaling pathway
GO:0030335: positive regulation of cell migration
GO:0043549: regulation of kinase activity
GO:0031035: actomyosin structure organization
GO:0051056: regulation of small GTPase mediated signal transduction
EC
GO:0022407: regulation of cell-cell adhesion
R-HSA-202427: Phosphorylation of CD3 and TCR zeta chains
GO:0050808: synapse organization
GO:0043516: regulation of DNA damage response
GO:0061061: muscle structure development
PC
GO:0001775: cell activation
GO:0051249: regulation of lymphocyte activation
GO:0050778: positive regulation of immune response
M34: PID TCR PATHWAY
GO:0098609: cell-cell adhesion
TH
R-HSA-1474244: ECM organization
R-HSA-3000178: ECM proteoglycans
GO:0098609: cell-cell adhesion
GO:0040017: positive regulation of locomotion
hsa04020: Calcium signaling pathway
Pseudobulk
GO:0046649: lymphocyte activation
GO:1903039: positive regulation of leukocyte cell-cell adhesion
GO:0050778: positive regulation of immune response
hsa04660: T cell receptor signaling pathway
GO:0002252: immune effector process
Bulk RNA-seq
GO:0050778: positive regulation of immune response
GO:0002250: adaptive immune response
GO:0045321: leukocyte activation
GO:0002694: regulation of leukocyte activation
GO:0001819: positive regulation of cytokine production
-log10(P)

**Fig. S8. Condition-stratified UMAP distribution and pathway enrichment across nuclei clusters.**

**A,** UMAP visualization of eight nuclei clusters stratified by condition, highlighting IBM-specific transcriptional segregation across non-immune and stromal cell types. IBM nuclei form discrete subclusters within MyoF, MyoS, MyoR, and MuSC, consistent with disease-associated transcriptional remodeling of the myofiber and muscle stem cell compartments. IBM nuclei similarly segregate within FAP/MSC, PC, EC, and TH clusters, indicating that IBM-associated transcriptional alterations extend beyond the immune compartment to encompass stromal, vascular, and helper T cell populations.

**B,** Pathway enrichment analysis of IBM snRNA-seq clusters and bulk RNA-seq pseudobulk. MyoF nuclei are enriched for antigen processing and presentation of endogenous and exogenous antigens, JNK cascade, positive regulation of chemotaxis, and type II interferon signaling, consistent with active participation of fast-type myofibers in the IBM inflammatory circuit. MyoS nuclei show enrichment for interferon gamma signaling, positive regulation of response to external stimulus, and insulin receptor signaling pathway. MyoR nuclei are enriched for synapse organization, cell-substrate adhesion, regulation of postsynapse organization, and cell-cell adhesion pathways, reflecting the regenerative transcriptional state of this subpopulation. MuSC nuclei show enrichment for antigen processing and presentation of endogenous antigen, type II interferon signaling, positive regulation of cytokine production, and detection of biotic stimulus, indicating immune activation of the muscle stem cell compartment in IBM. FAP/MSC nuclei are enriched for T cell receptor signaling, positive regulation of cell migration, kinase activity, actomyosin structure organization, and small GTPase-mediated signal transduction. EC nuclei show enrichment for regulation of cell-cell adhesion, phosphorylation of CD3 and TCR zeta chains, synapse organization, and

DNA damage response. PC nuclei demonstrate the strongest enrichment signal overall, with cell activation, regulation of lymphocyte activation, positive regulation of immune response, TCR pathway, and cell-cell adhesion as top terms. TH nuclei are enriched for ECM organization, ECM proteoglycans, cell-cell adhesion, and calcium signaling. Pseudobulk analysis confirms lymphocyte activation, leukocyte cell-cell adhesion, T cell receptor signaling, and immune effector processes as IBM-wide transcriptional features. Bulk RNA-seq independently corroborates positive regulation of immune response, adaptive immune response, leukocyte activation, regulation of leukocyte activation, and positive regulation of cytokine production as the dominant IBM-associated pathways.

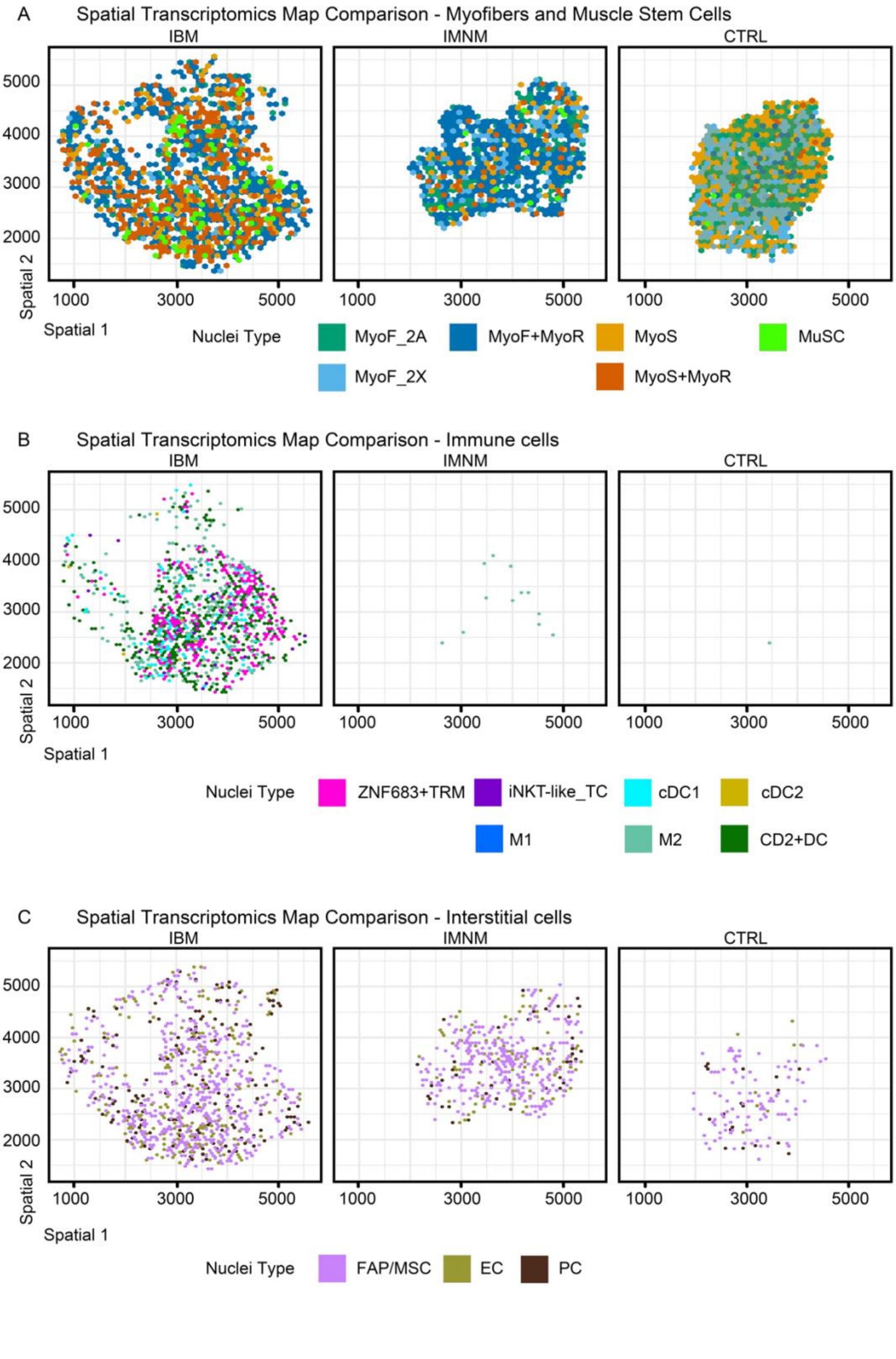
A
Spatial Transcriptomics Map Comparison - Myofibers and Muscle Stem Cells
IBM
IMNM
CTRL
Spatial 2
Spatial 1
1000
3000
5000
2000
3000
4000
5000
Nuclei Type
MyoF_2A
MyoF+MyoR
MyoS
MuSC
MyoF_2X
MyoS+MyoR
B
Spatial Transcriptomics Map Comparison - Immune cells
Nuclei Type
ZNF683+TRM
iNKT-like_TC
cDC1
cDC2
M1
M2
CD2+DC
C
Spatial Transcriptomics Map Comparison - Interstitial cells
Nuclei Type
FAP/MSC
EC
PC

334

**Fig. S9. Spatial transcriptomics comparison of IBM with IMNM and adult control muscle.**

**A,** Spatial distribution of myofiber and muscle stem cell-associated nuclei, including MyoF_2A, MyoF_2X, mixed MyoF+MyoR regions, MyoS, mixed MyoS+MyoR regions, and MuSC. IBM exhibited greater spatial heterogeneity and intermixing of myofiber-associated states compared with IMNM and adult control muscle. B, Spatial distribution of immune cell populations. IBM showed heterogeneous localization of ZNF683+ TRM, iNKT-like_TC, macrophage subsets (M1 and M2), and dendritic cell populations (cDC1, cDC2, and CD2+DC), whereas IMNM and adult control muscle showed sparse immune cell representation dominated primarily by M2 macrophages. C, Spatial distribution of interstitial cell populations, including FAP/MSC, endothelial cells (EC), and pericytes (PC), which showed broadly comparable spatial patterns across conditions proportionate to the underlying myofiber distribution.

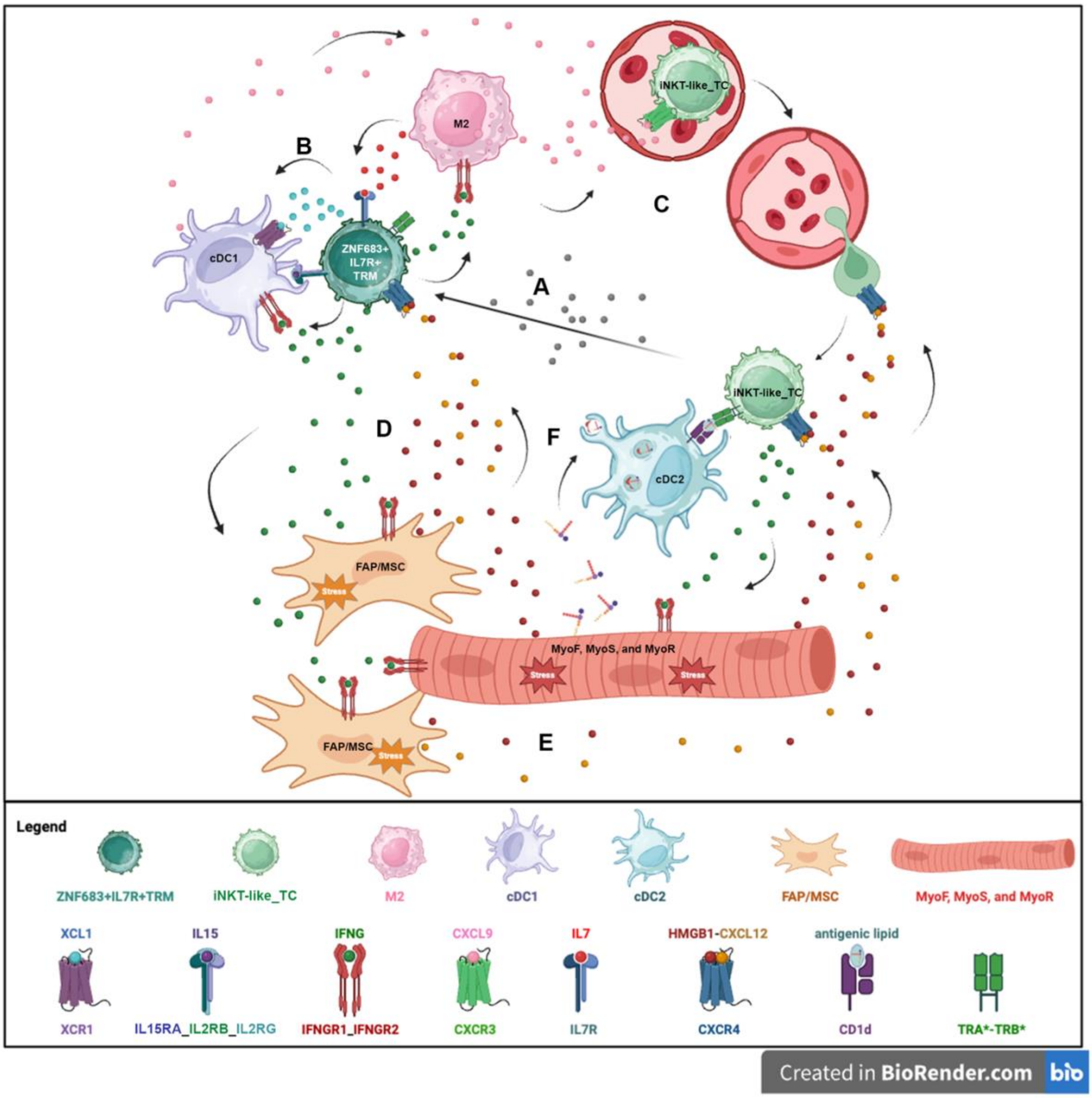

M2
iNKT-like_TC
B
C
cDC1
ZNF683+ IL7R+ TRM
A
iNKT-like_TC
D
F
cDC2
FAP/MSC
Stress
MyoF, MyoS, and MyoR
Stress
Stress
FAP/MSC
Stress
E
Legend
ZNF683+IL7R+TRM
iNKT-like_TC
M2
cDC1
cDC2
FAP/MSC
MyoF, MyoS, and MyoR
XCL1
XCR1
IL15
IL15RA_IL2RB_IL2RG
IFNG
IFNGR1_IFNGR2
CXCL9
CXCR3
IL7
IL7R
HMGB1-CXCL12
CXCR4
antigenic lipid
CD1d
TRA*-TRB*
Created in BioRender.com

**Fig. S10. Proposed model of the myofiber–inflammatory–interstitial axis in inclusion body myositis.**

This model combines observations from the present study with previously established biological mechanisms to summarize a proposed myofiber–inflammatory–interstitial axis in inclusion body myositis. The steps shown are inferred from transcriptional, interaction, and spatial data and have not been experimentally demonstrated in this study.

**A,** iNKT-like T cells (iNKT-like_TC), upon exposure to cytokines and chemokines associated with memory T cell formation, acquire *ZNF683* and *IL7R* expression, giving rise to ZNF683+IL7R+ tissue-resident memory T cells (ZNF683+IL7R+TRM).

**B,** ZNF683+IL7R+TRM secrete XCL1, which binds XCR1 on type 1 dendritic cells (cDC1). cDC1 in turn produce IL15 and trans-present it to ZNF683+IL7R+TRM through IL15RA_IL2RB_IL2RG, a route capable of activating ZNF683+IL7R+TRM independently of the T-cell receptor (TCR).

**C,** ZNF683+IL7R+TRM secrete IFNG, which engages IFNGR1_IFNGR2 on M2 macrophages (M2) and cDC1. These populations reciprocally secrete CXCL9, which may recruit iNKT-like_TC to muscle through CXCR3, and IL7, associated with long-lived memory characteristics in ZNF683+IL7R+TRM.

**D,** IFNG derived from both ZNF683+IL7R+TRM and iNKT-like_TC engages IFNGR1_IFNGR2 on myofibers and stromal cells, inducing HMGB1 expression in fast-type (MyoF), slow-type (MyoS), and regenerating (MyoR) myofibers, and HMGB1 together with CXCL12 in fibro-adipogenic precursors / mesenchymal stromal cells (FAP/MSC).

**E,** HMGB1–CXCL12 heterodimers engage CXCR4 on both ZNF683+IL7R+TRM and iNKT-like_TC, a mechanism proposed to retain ZNF683+IL7R+TRM within muscle, direct iNKT-like_TC toward areas of inflammation, and sustain cytokine and chemokine production supporting a persistent pro-inflammatory state.

**F,** MyoF subjected to IFNG-associated stress express genes that may lead to antigenic lipid secretion. These lipids are taken up by type 2 dendritic cells (cDC2), where presentation of CD1d-bound lipid antigen to the NKT-associated TCR may activate iNKT-like_TC and drive secretion of IFNG and other cytokines. CD1d restriction was not demonstrated in this study.

Table S1. Single Nuclei RNA-seq Samples

| Diagnosis | Number of samples | Age Group | Male / Female Distribution | CK (U/L) | Biopsy Site Frequency |
|---|---|---|---|---|---|
| ASyS_Jo-1 | 2 | 41-50 to 51-60 | 1 / 1 | 703-9992 | Sm, BB |
| ASyS_OJ | 2 | 51-60 to 71-80 | 0 / 2 | ND | BB |
| ASyS_PL7 | 1 | 71-80 | 0 / 1 | 2094 | QF |
| DM_MDA5 | 2 | 41-50 | 1 / 1 | 451 | VL, Del |
| DM_Mi-2 | 2 | 51-60 to 61-70 | 0 / 2 | 1408-1738 | Del |
| DM_NXP-2 | 2 | 61-70 | 0 / 2 | 2910-3759 | BB |
| DM_TIF1-γ | 2 | 41-50 to 61-70 | 1 / 1 | 1166-18775 | RF, Del |
| DM_SAE | 1 | 61-70 | 0 / 1 | 232 | TB |
| IBM | 4 | 61-70 to 71-80 | 4 / 0 | 301-1145 | BB>RF |
| IMNM_HMGCR | 2 | 31-40 to 41-50 | 1 / 1 | 1441-4995 | BB |
| IMNM_SRP | 3 | 61-70 to 71-80 | 1 / 2 | 3165-3723 | BB |
| Non-disease controls | 2 | 21-30 to 31-40 | 1 / 1 | ND | BB |

ND – no data; Sm – Semimembranosus; BB – biceps brachii; VL – vastus lateralis; Del – deltoid; RF – rectus femoris; TB – triceps brachii

Table S2. All Nuclei - Single Nuclei RNA-seq Nuclei Distribution

All Conditions – Raw Counts

| Raw Counts by Condition | CTRL _Adt | ASyS _Jo-1 | ASyS _OJ | ASyS _PL7 | DM _MDA5 | DM _Mi-2 | DM _NXP-2 | DM _TIF1-γ | DM _SAE | IBM | IMNM _HMGCR | IMNM _SRP |
|---|---|---|---|---|---|---|---|---|---|---|---|---|
| MyoF | 6329 | 509 | 2076 | 1599 | 3517 | 1462 | 1510 | 2026 | 1898 | 3687 | 4270 | 4916 |
| MyoS | 5459 | 281 | 2013 | 513 | 2405 | 3202 | 1528 | 728 | 1043 | 3064 | 2683 | 2667 |
| TC | 219 | 1434 | 722 | 101 | 185 | 249 | 390 | 646 | 254 | 2489 | 908 | 482 |
| APC | 190 | 1532 | 2070 | 403 | 477 | 543 | 693 | 1195 | 385 | 1599 | 1706 | 1591 |
| MuSC | 739 | 201 | 715 | 275 | 231 | 457 | 146 | 321 | 273 | 899 | 1259 | 783 |
| TH | 165 | 414 | 272 | 96 | 105 | 150 | 326 | 254 | 151 | 1541 | 603 | 679 |
| EC | 844 | 178 | 612 | 401 | 894 | 879 | 277 | 527 | 275 | 1182 | 1012 | 608 |
| FAP/ MSC | 724 | 209 | 453 | 517 | 397 | 647 | 352 | 424 | 212 | 1584 | 1107 | 873 |
| MyoR | 89 | 351 | 1744 | 494 | 517 | 1366 | 2051 | 648 | 1029 | 2067 | 2274 | 1873 |
| PC | 812 | 129 | 423 | 280 | 673 | 483 | 294 | 435 | 199 | 783 | 368 | 257 |
| PlC | 4 | 169 | 56 | 19 | 9 | 3 | 7 | 267 | 21 | 71 | 77 | 488 |
| BC | 14 | 194 | 119 | 6 | 19 | 43 | 20 | 72 | 62 | 29 | 48 | 117 |
| LEC | 12 | 5 | 44 | 25 | 8 | 10 | 10 | 12 | 7 | 44 | 44 | 3 |
| Total Nuclei Count | 15600 | 5606 | 11319 | 4729 | 9437 | 9494 | 7604 | 7555 | 5809 | 19039 | 16359 | 15337 |

Table S3. All Nuclei - Single Nuclei RNA-seq Nuclei Distribution

All Conditions – Proportion

| % by Condition | CTRL _Adt | ASyS _Jo-1 | ASyS _OJ | ASyS _PL7 | DM _MDA5 | DM _Mi-2 | DM _NXP-2 | DM _TIF1-γ | DM _SAE | IBM | IMNM _HMGCR | IMNM _SRP |
|---|---|---|---|---|---|---|---|---|---|---|---|---|
| MyoF | 40.57 | 9.08 | 18.34 | 33.81 | 37.27 | 15.40 | 19.86 | 26.82 | 32.67 | 19.37 | 26.10 | 32.05 |
| MyoS | 34.99 | 5.01 | 17.78 | 10.85 | 25.48 | 33.73 | 20.09 | 9.64 | 17.95 | 16.09 | 16.40 | 17.39 |
| TC | 1.40 | 25.58 | 6.38 | 2.14 | 1.96 | 2.62 | 5.13 | 8.55 | 4.37 | 13.07 | 5.55 | 3.14 |
| APC | 1.22 | 27.33 | 18.29 | 8.52 | 5.05 | 5.72 | 9.11 | 15.82 | 6.63 | 8.40 | 10.43 | 10.37 |
| MuSC | 4.74 | 3.59 | 6.32 | 5.82 | 2.45 | 4.81 | 1.92 | 4.25 | 4.70 | 4.72 | 7.70 | 5.11 |
| TH | 1.06 | 7.38 | 2.40 | 2.03 | 1.11 | 1.58 | 4.29 | 3.36 | 2.60 | 8.09 | 3.69 | 4.43 |
| EC | 5.41 | 3.18 | 5.41 | 8.48 | 9.47 | 9.26 | 3.64 | 6.98 | 4.73 | 6.21 | 6.19 | 3.96 |
| FAP/ MSC | 4.64 | 3.73 | 4.00 | 10.93 | 4.21 | 6.81 | 4.63 | 5.61 | 3.65 | 8.32 | 6.77 | 5.69 |
| MyoR | 0.57 | 6.26 | 15.41 | 10.45 | 5.48 | 14.39 | 26.97 | 8.58 | 17.71 | 10.86 | 13.90 | 12.21 |
| PC | 5.21 | 2.30 | 3.74 | 5.92 | 7.13 | 5.09 | 3.87 | 5.76 | 3.43 | 4.11 | 2.25 | 1.68 |
| PlC | 0.03 | 3.01 | 0.49 | 0.40 | 0.10 | 0.03 | 0.09 | 3.53 | 0.36 | 0.37 | 0.47 | 3.18 |
| BC | 0.09 | 3.46 | 1.05 | 0.13 | 0.20 | 0.45 | 0.26 | 0.95 | 1.07 | 0.15 | 0.29 | 0.76 |
| LEC | 0.08 | 0.09 | 0.39 | 0.53 | 0.08 | 0.11 | 0.13 | 0.16 | 0.12 | 0.23 | 0.27 | 0.02 |
| Total Nuclei Count | 100 | 100 | 100 | 100 | 100 | 100 | 100 | 100 | 100 | 100 | 100 | 100 |

Table S4. Differential Expression of Genes of IBM TC vs. other Myositis and Controls

Upregulated Genes - $Log_2FC > 1$ , adjusted $p$ value < 0.05 , pct.2 < 0.05

| | | | |
|---|---|---|---|
| *CNBD1* | *MMP16* | *LINC00343* | *ITGA11* |
| *AC002331.1* | *FXYD2* | *VSTM2L* | *AC105180.2* |
| *LINC01446* | *AC087878.1* | *VIT* | *AC011370.1* |
| *LINC02195* | *KCNMB2* | *AC211486.2* | *AL121672.1* |
| *AL591686.2* | *KCNC2* | *AC012645.2* | *AC011483.1* |
| *LINC00461* | *LINC02836* | *TBC1D9* | *WNT9A* |
| *LONRF2* | *CNTNAP3* | *LINC01098* | *AC023509.2* |
| *AF121898.1* | *AC068987.3* | *AC008571.2* | *MEGF10* |
| *AC117453.1* | *PAMR1* | *ASIC1* | *AC009560.4* |
| *RGS18* | *AL513128.3* | *AL023755.1* | *RYR2* |
| *AC013451.1* | *LINC00943* | *CNTNAP3B* | *MYH4* |
| *IL5RA* | *AC244213.1* | *AL136171.2* | *SRPX2* |
| *AC013460.1* | *AL591518.1* | *AC139100.1* | *AL035634.1* |
| *AJ011931.2* | *DSCAML1* | *PDGFB* | *TNC* |
| *LINC02357* | *AC022929.2* | *AP000561.1* | *SOX5* |
| *GEM* | *AC007218.3* | *GRID1* | *SLC24A2* |
| *AC005599.1* | *ITGBL1* | *SLC1A1* | *PTGER3* |
| *LINC00102* | *SATL1* | *AC073091.4* | *LINC02614* |
| *CNTNAP3C* | *ZNF683* | *HS3ST3A1* | *AL355490.2* |
| *TBL1Y* | *LINC02381* | *AL355501.1* | *HRH1* |
| *ADPGK-AS1* | *AC079336.4* | *SLC22A16* | *ABCA9-AS1* |
| *AC022929.1* | *AC139768.1* | *AC004917.1* | *AL121672.2* |
| *CCL18* | *AC048382.1* | *AL109930.1* | *SCN9A* |
| *LINC02109* | *AP003721.2* | *LINC02343* | *AC098848.1* |
| *AC090376.1* | *KLHL4* | *PLXNA2* | *ZBTB8B* |
| *NGB* | *AC024230.1* | *NTN4* | *AC026803.3* |
| *PCBP3* | *UPB1* | *AC009264.1* | *ADORA2A-AS1* |
| *AC119150.1* | *RNF17* | *ANGPTL5* | *SMOC2* |
| *ELAVL4* | *COL17A1* | *DIXDC1* | *SPEF2* |
| *ETV1* | *AC079203.2* | *ZBED2* | *AC078777.1* |
| *AC009102.4* | *TVP23A* | *SLC47A2* | *GNAO1* |
| *AL136985.3* | *AC099520.1* | *PPARG* | *AL353801.1* |
| *AL139294.1* | *C14orf132* | *LINC01479* | *EBF2* |

| | | | |
|---|---|---|---|
| *ZFY-AS1* | *NLGN4X* | *SYT17* | *GDPD4* |
| *FRMPD3-AS1* | *SFTPD* | *AC004039.1* | *AC019197.1* |
| *TGFBI* | *LINC01435* | *AL162414.1* | *PLA2R1* |
| *AC113615.2* | *ELN* | *AF130417.1* | *AP001372.1* |
| *AL662884.5* | *DSCAM* | *AL031846.2* | *PKD1L3* |
| *BTNL9* | *AL445231.1* | *HMCN1* | *SOX10* |
| *TRPA1* | *AC009975.1* | *AC026462.4* | *AP005131.5* |
| *ZBTB46-AS1* | *PCA3* | *AC211486.5* | *AC008728.1* |
| *LINC02824* | *AL353586.1* | *AC097534.2* | *AL133268.3* |
| *AC109472.1* | *KIF5C* | *BMPER* | *JDP2* |
| *LINC02725* | *CLSTN2* | *LINC01877* | *PLCH1* |
| *AC002306.1* | *ESRP1* | *PXT1* | *AL158011.1* |
| *AL110292.1* | *LINC00892* | *GFRA2* | *KCNK12* |
| *AC093607.1* | *ZBTB46* | *AC145124.1* | *IL26* |
| *AC007686.2* | *SLC7A14-AS1* | *AC006972.1* | *GREM1* |
| *PCDH11Y* | *TBX20* | *LINC00707* | *POTEG* |
| *SEMA4G* | *PAPSS2* | *AL133284.1* | *CFAP100* |
| *PPFIA2* | *TUBB1* | *NRG1* | *GRM7* |
| *FGF6* | *AC096577.1* | *AC009478.1* | *LINC01322* |
| *AC105402.4* | *ITGA9-AS1* | *AK7* | *STAC* |
| *SCRG1* | *EPHA3* | *CDH18* | *ZRANB2-AS1* |
| *AC006145.1* | *LPAR1* | *F13A1* | *GLIPR1L1* |
| *ABCA8* | *DNAJB7* | *NTSR1* | *GALNT15* |
| *PRB1* | *AC006581.1* | *AC093425.1* | *MCOLN3* |
| *SNAP25-AS1* | *PKHD1* | *AL670729.2* | *MGAT4C* |
| *GJB7* | *ZBTB20-AS3* | *AC114316.1* | *SLIT3* |
| *LINC00970* | *NOVA1* | *AC004542.2* | *TRPC5* |
| *PRDM16* | *MEGF11* | *JAKMIP2-AS1* | *ROBO2* |
| *PAK5* | *AC069437.1* | *HTRA1* | *CNTN4* |
| *AC015914.1* | *AC005050.3* | *SEMA5A* | *CCDC3* |
| *Z99774.1* | *AC137810.1* | *LINC02511* | *ZBTB7C* |
| *NLGN4Y* | *MIR4300HG* | *ZFP28* | *DNAH9* |
| *LRRTM4* | *TENM2* | *NRIP2* | *AC139769.2* |
| *ATCAY* | *SYT2* | *AC087190.1* | *AL590705.1* |
| *SPDYE2B* | *AC090541.1* | *AKAP12* | *PERP* |
| *PPP1R42* | *RTN1* | *AC007347.1* | *SEZ6* |

| | | | |
|---|---|---|---|
| *AL928970.1* | *AC007569.1* | *AL603840.1* | *AC092436.3* |
| *SLC24A5* | *LAMA3* | *AL583824.1* | *AC108159.1* |
| *AC123512.1* | *NRAV* | *AC006460.1* | *ANO2* |
| *MFAP5* | *RIPOR3* | *AC008945.2* | *AGBL5* |
| *LINC01948* | *AC013437.1* | *AC008680.1* | *HEATR4* |
| *AC018410.2* | *AC109492.1* | *AC009060.1* | *AL049646.1* |
| *AC046195.1* | *ZNF665* | *MMP26* | *FAM189A1* |
| *LINC02742* | *AC005856.1* | *LINC01500* | *ROR1-AS1* |
| *AC073569.1* | *LINC01609* | *LSAMP* | *EMP1* |
| *COL25A1* | *IGFN1* | *CADPS2* | *SLC5A10* |
| *SLC26A3* | *EGFR* | *PSTK* | *PTPRT* |
| *ZSCAN31* | *DANT2* | *EBF1* | *Z99758.1* |
| *AL161734.2* | *AC104078.1* | *AL583785.1* | *IQCJ-SCHIP1* |
| *P2RY14* | *PLAUR* | *LINC01814* | *WNT2B* |
| *LOXL1* | *COL15A1* | *FOXN3-AS2* | *AC123912.1* |
| *AC104664.1* | *AC061958.1* | *NR2F2-AS1* | *TANC1* |
| *AC116534.1* | *AL390728.5* | *SLC3A1* | *AGMO* |
| *EVA1A* | *SLC48A1* | *SMCO4* | *GALNTL6* |
| *CRISPLD2* | *AC008050.1* | *PDZRN4* | *ANTXR1* |
| *ZBTB20-AS4* | *CEMIP* | *CDH12* | *MEG3* |
| *CCR5AS* | *MMP25* | *HMCN2* | *AL138930.1* |
| *SAMD5* | *BEND7* | *CFAP299* | *AP006284.1* |
| *AQP11* | *AP001528.1* | *MYO3B* | *AL589990.1* |
| *AL035401.1* | *PCDH12* | *Z82188.2* | *LINC00558* |
| *C5orf58* | *PARD6G-AS1* | *AL021155.5* | *COL19A1* |
| *CORO2B* | *NTRK2* | *NUGGC* | *SEMA3E* |
| *AL391117.1* | *AL358473.2* | *ANGPTL2* | *EDDM13* |
| *AC108863.2* | *AC015971.1* | *TENM4* | *ARHGEF26* |
| *AC127521.1* | *AC010978.1* | *INO80B* | *TOM1L1* |
| *LINC01678* | *AC037487.2* | *AC018541.1* | *GPX2* |
| *LARGE-AS1* | *NGF* | *GARNL3* | *SYN3* |
| *AC092490.2* | *AC093690.1* | *AC069228.1* | *ERC2* |
| *AC013643.2* | *AL590807.1* | *LINC01090* | *AC009686.1* |
| *STMN3* | *PLXDC1* | *ZDHHC11B* | *ZNF213* |
| *LINC01320* | *SRSF12* | *AL031602.2* | *AC105031.2* |
| *AC002546.1* | *LINC01331* | *AC002066.1* | *AL137139.2* |

| | | | |
|---|---|---|---|
| *AC025287.4* | *REC114* | *DNAJC6* | *CYSLTR1* |
| *MEG8* | *SARDH* | *AC002451.1* | *TTTY14* |
| *AL049840.3* | *OSBPL10* | *CAMK1* | *AC016910.1* |
| *ASAP2* | *CSRNP3* | *WNK2* | *AP001267.3* |
| *AC096570.2* | *AC244102.4* | *SLIT2* | *AP000753.2* |
| *DENND2A* | *PAPLN* | *SCN8A* | *AC005355.1* |
| *AC046130.1* | *LINCMD1* | *CLDN20* | *WNT5B* |
| *TRIM17* | *MAMDC2* | *ZFPM2-AS1* | *SCUBE3* |
| *FP700111.1* | *ADNP-AS1* | *LINC00882* | *AC134772.1* |
| *AC007622.2* | *IL21-AS1* | *PLCB4* | *AC018946.1* |
| *EIF1AY* | *ABCA9* | *SLC5A4-AS1* | *CTTNBP2NL* |
| *AC007319.1* | *AC034268.2* | *ZNF880* | *DLGAP2* |
| *DPYSL3* | *PPL* | *CPNE4* | *AC084816.1* |
| *AC044839.1* | *LINC02388* | *ADAMTSL3* | *GPC5-AS1* |
| *AFAP1L2* | *SMIM18* | *AC009365.4* | *AC116158.2* |
| *MEOX2* | *AC099535.2* | *ALK* | *AC093879.1* |
| *AL161430.1* | *TNFRSF19* | *AC022872.1* | *AP002075.1* |
| *AC018557.3* | *AC005746.2* | *PREX2* | *AC093495.1* |
| *PP2D1* | *AL365295.2* | *RNF212B* | *ITPRIPL2* |
| *AC008906.1* | *AL050344.1* | *AL589693.1* | *AC016493.1* |
| *AC010680.5* | *RADX* | *LRRC4C* | *AP000721.2* |
| *LINC02112* | *ABCA6* | *FGFR2* | *GPR82* |
| *CDKN2B* | *AC018557.1* | *EVC* | *LUZP2* |
| *SYT1* | *EPHA5* | *IGF1* | *AC107068.2* |
| *DGKG* | *AL139120.1* | *C21orf62* | *AC010320.2* |
| *SLC10A1* | *NTN1* | *TERB1* | *C17orf97* |
| *STEAP1B* | *AC067969.1* | *FGL2* | *SMCR5* |
| *CLCA4-AS1* | *BMP5* | *AL117329.1* | *ITIH5* |
| *AC005323.2* | *AC060834.2* | *AC005884.1* | *EPHA6* |
| *AC023906.4* | *KCNC1* | *CARMN* | *ZDHHC11* |
| *PLEKHN1* | *CALCRL* | *DCLK1* | *AC007098.1* |
| *MRC1* | *SREBF2-AS1* | *AC023421.2* | *ZNF528-AS1* |
| *NR5A2* | *G0S2* | *APBB2* | *AP001767.3* |
| *AL035071.2* | *AC008554.1* | *CBR3* | *AL078581.4* |
| *ITIH6* | *AC016590.1* | *ABLIM3* | *CALD1* |
| *MAMDC2-AS1* | *NCOA7-AS1* | *AC024267.4* | *AC009226.1* |

| | | | |
|---|---|---|---|
| *AC012404.1* | *VWF* | *TTC9* | *PTPRN2* |
| *CRMP1* | *AP005329.2* | *AC005253.1* | *AC008731.1* |
| *AC078923.1* | *MXI1* | *CP* | *RAB31* |
| *KCNC4* | *ADGRV1* | *ABCA3* | *AL445928.2* |
| *L1CAM* | *AL022323.1* | *AC083870.1* | *CCDC80* |
| *AC022034.3* | *ADGRL3* | *NPAS3* | *FAM25G* |
| *MUC12* | *ELF3-AS1* | *NEUROD1* | *AC008537.4* |
| *AL035446.2* | *SVEP1* | *NUAK1* | *COL4A2* |
| *RBMS3-AS2* | *KCNMB2-AS1* | *AL035427.1* | *KIAA1324* |
| *AC004494.1* | *RCAN3AS* | *AC104532.2* | *SSTR2* |
| *MAPK10* | *TTTY10* | *AL022238.2* | *KCNJ3* |
| *PLEKHG6* | *EMILIN2* | *SPNS3* | *PARVA* |
| *AC016644.1* | *FAM20A* | *CDC14B* | *ZFHX2-AS1* |
| *ARG1* | *GNG12-AS1* | *IL6R* | *AC017101.1* |
| *COL4A1* | *RARB* | *PDGFC* | *AP000943.2* |
| *AC012184.4* | *AL031670.1* | *EPAS1* | *FIGN* |
| *AP002026.1* | *RAPGEF4* | *CCDC28B* | *AL136114.1* |
| *ARHGAP26-AS1* | *SNRK-AS1* | *TAC4* | *TBC1D16* |
| *AP003392.1* | *ARHGAP22* | *PRR16* | *KLHL1* |
| *CR1L* | *AL589740.1* | *ECT2L* | |

Table S5. Differential Expression of Genes of IBM APC vs. other Myositis and Controls

Upregulated Genes - $Log_2FC > 1$ , adjusted $p$ value < 0.05 , pct.2 < 0.01

| | | | |
|---|---|---|---|
| *CD1E* | *AC104827.1* | *AC004054.1* | *AC006369.1* |
| *AC112907.1* | *TBL1Y* | *AF228727.1* | *AC007091.1* |
| *ALOX15* | *DUSP15* | *AC108879.1* | *AL353593.2* |
| *OR7D4* | *ASTN1* | *AC025434.1* | *AL132642.1* |
| *AC117480.1* | *DTHD1* | *MACROD2-AS1* | *PRB2* |
| *RNF17* | *PLA2G2D* | *HHLA2* | *LONRF2* |
| *AC113615.2* | *GPR174* | *SLC12A5-AS1* | *AC067945.3* |
| *IFNG* | *AMBP* | *AC110760.1* | *AL358473.2* |
| *LINC02312* | *AC079328.2* | *AP003464.1* | *GNAO1* |
| *ARSJ* | *UBASH3A* | *AL157829.1* | *ADAM21* |
| *AC239860.4* | *BSN-DT* | *AL591178.1* | *AC124312.5* |
| *CTLA4* | *LINC01836* | *AL590705.1* | *GPR55* |
| *LINC02520* | *RXRG* | *AL359436.1* | *AP002505.1* |
| *AC074124.1* | *SLC1A2* | *LINC00377* | *AC097459.1* |
| *GRM5-AS1* | *AP000857.2* | *LINC01239* | *CYP4F11* |
| *AC113418.1* | *AC104938.1* | *FGF14* | *NAV2-AS5* |
| *U91324.1* | *SLC29A4* | *OR2G6* | *LRRC18* |
| *THRA1/BTR* | *ARL4D* | *AL450338.2* | *AC002472.1* |
| *AC104078.2* | *LINC02343* | *VIT* | *IZUMO3* |
| *AC024028.1* | *TRAT1* | *AC073091.4* | *LINC01871* |
| *KCNK1* | *PAMR1* | *AC008914.1* | *SLC2A10* |
| *AC010609.1* | *SIRPG* | *AC068987.3* | *CYP4X1* |
| *AC007743.1* | *TSBP1* | *AL354977.1* | *AC138904.1* |
| *AL138899.1* | *AL391069.1* | *CLEC4F* | *AC007952.2* |
| *AL590006.1* | *AL731575.1* | *CD1C* | *AC135782.1* |
| *TRPA1* | *AL049874.3* | *AC023095.1* | *GFI1B* |
| *TUSC7* | *AC078955.1* | *LINC01446* | *HLA-DQB1-AS1* |
| *SPX* | *STYK1* | *AC007036.1* | *LINC02725* |
| *AL445493.2* | *AC011239.1* | *AC021242.2* | *AC087428.1* |
| *AP001893.3* | *LINC00298* | *AC004594.1* | *AC243829.2* |
| *LINC00051* | *AC092378.1* | *AL355835.1* | *C4BPA* |
| *AL355597.1* | *AC119150.1* | *AC133961.1* | *AC006145.1* |
| *SCARA5* | *LINC02446* | *INPP5J* | *LINC01679* |

| | | | |
|---|---|---|---|
| *AC239799.1* | *AL079307.1* | *LEF1* | *LINC02204* |
| *TBX21* | *FAM153B* | *AWAT1* | *PPP1R1C* |
| *MSX1* | *AL356608.3* | *AC010615.2* | *AL512283.2* |
| *LINC01802* | *LINC00536* | *AC091946.1* | *AC244213.1* |
| *AC008571.2* | *AC100830.2* | *TIFAB* | *IGSF21-AS1* |
| *AL035078.1* | *AC060834.2* | *LINC01956* | *BOC* |
| *AC116634.1* | *LINC02086* | *LINC01697* | *LINC01625* |
| *AC067945.4* | *AC105450.1* | *KCNK2* | *NCMAP-DT* |
| *TSPAN2* | *WIF1* | *FRMPD3* | *AC234031.1* |
| *AC004690.2* | *NALCN* | *GLS2* | *IGDCC4* |
| *APCDD1* | *ADRA1B* | *ASTL* | *NAALADL2-AS3* |
| *AL024497.2* | *SPON1-AS1* | *LINC02054* | *ELN* |
| *AL022323.4* | *PCAT7* | *SLC14A1* | *AL592430.1* |
| *AL365394.1* | *LINC01198* | *IDI2* | *ZIC1* |
| *AC023794.1* | *GNAS-AS1* | *OTOGL* | *BCAR1* |
| *AC011483.1* | *MGAT3* | *ENPP5* | *LINC02471* |
| *AC244517.11* | *LINC02066* | *MIR548A1HG* | *AL049869.2* |
| *TMSB4Y* | *NTN4* | *AL109930.1* | *AL603840.1* |
| *AC005081.1* | *AC009478.1* | *CD27* | *VIPR2* |
| *NAALADL2-AS2* | *CD3D* | *LRRIQ1* | *AL356113.1* |
| *AC002377.1* | *AC108860.2* | *ISM1* | *AL592431.1* |
| *GATA3* | *LINC00944* | *NPR2* | *LINC01563* |
| *C6* | *LINC02069* | *AC090617.5* | *APBA2* |
| *AC006372.3* | *AC005232.1* | *SOX5-AS1* | *MMP26* |
| *MEOX2* | *ADGRF5* | *IL2RB* | *AC139019.1* |
| *PPP1R3G* | *AC008067.1* | *EGFLAM* | *CXCR6* |
| *ANO1* | *IQSEC3* | *CYYR1-AS1* | *EDIL3* |
| *SSUH2* | *AC087620.1* | *ANGPT4* | *MMRN2* |
| *CD3E* | *XXYLT1-AS2* | *AC005089.1* | *AC006504.2* |
| *NOL4* | *SDR42E2* | *CXXC4-AS1* | *AL023755.1* |
| *AC025419.1* | *LINC02821* | *TPM1-AS* | *SRPX2* |
| *DUSP4* | *CDHR2* | *AC044839.1* | *NFIA-AS1* |
| *AC008703.1* | *C17orf100* | *CDH20* | *P2RY10* |
| *PDGFD* | *MFAP5* | *AL713852.1* | *PHYHIPL* |
| *AC090018.2* | *MTUS2* | *GPR176* | *Z98949.1* |
| *AC093010.2* | *ACSBG2* | *AC022034.3* | *NECTIN3* |

| | | | |
|---|---|---|---|
| *SUSD4* | *SNX22* | *GP1BA* | *AC124290.1* |
| *AC010245.1* | *TRBC2* | *AL096794.1* | *DBN1* |
| *AC093110.1* | *IL5RA* | *NECAB1* | *BRINP1* |
| *BTNL9* | *SHISAL2A* | *RAB3C* | *AC079089.1* |
| *GRB14* | *AP000561.1* | *EBF4* | *AC134312.5* |
| *AL136119.1* | *FAM245A* | *AC007679.1* | *CFAP58* |
| *SPTBN2* | *MYH4* | *AC002072.1* | *ZNF536* |
| *CCL14* | *ANGPTL7* | *ZFHX4-AS1* | *LINC01117* |
| *AC008574.1* | *C10orf105* | *CDRT1* | *LURAP1L-AS1* |
| *EPHX2* | *AC107463.1* | *AC024361.3* | *AL021707.2* |
| *LINC02421* | *EXOC3L4* | *SELENBP1* | *FAT1* |
| *SYNPR* | *AL670729.3* | *AC245164.2* | *FOXO6* |
| *TDRD1* | *RORC* | *ICOS* | *AC008415.1* |
| *AC022087.1* | *AC096577.1* | *AP006565.1* | *LINC02848* |
| *ZNF711* | *LINC01331* | *SLC9A2* | *LINC02465* |
| *Z98949.3* | *FRMD6-AS1* | *CD24* | *LINC01550* |
| *CST7* | *AC098851.1* | *AC091646.1* | *AC108058.1* |
| *FBP2* | *AC090376.1* | *CLEC6A* | *RGN* |
| *FBXO17* | *TMEM200A* | *SPOCK2* | *PXDN* |
| *LINC00595* | *ALDH1A3* | *BCAM* | *MAP1B* |
| *GPD1* | *AC011416.4* | *DUSP26* | *AC022929.2* |
| *CNTNAP3C* | *AL445465.1* | *NGF* | *RBPMS2* |
| *TPO* | *AC009152.3* | *AC006148.1* | *PRDM8* |
| *AC026124.1* | *ITGBL1* | *SNTG2* | *KLRG2* |
| *LPAR4* | *AC007216.1* | *SMAD9* | *AC090709.1* |
| *LINC02511* | *EMX2OS* | *AC091534.1* | *NFIA-AS2* |
| *AC009063.3* | *OXTR* | *FAM135B* | *AC016987.1* |
| *AP000350.6* | *AL353593.3* | *GNA14-AS1* | *PTGDR* |
| *TSPAN8* | *AJ003147.2* | *FAM25G* | *HSD11B1* |
| *AL121929.2* | *KCNJ6* | *SLC12A3* | *C14orf132* |
| *AC008440.2* | *CXCL17* | *DNAJB13* | *FZD4* |
| *AL670729.2* | *AC008443.4* | *AC015849.1* | *AL136298.1* |
| *AC093607.1* | *AL359220.1* | *C1QTNF2* | *ROS1* |
| *AL137026.3* | *ASPA* | *EPHA4* | *AC135166.1* |
| *PSG8* | *NEUROD1* | *AP003059.1* | *EFHB* |
| *AC125793.1* | *AC010336.6* | *AL133230.1* | *PLAAT1* |

| | | | |
|---|---|---|---|
| *AC018541.1* | *AL158195.1* | *BHMT2* | *KIAA0408* |
| *AC022272.1* | *RAI1-AS1* | *AP001363.2* | *MFAP3L* |
| *TIGIT* | *TRAC* | *LINC01547* | *GLI2* |
| *SPIB* | *ADAM18* | *AC104574.2* | *PPM1J* |
| *MYADML2* | *KY* | *AL355581.1* | *SLC14A2* |
| *LINC00519* | *WNT9A* | *SLC12A1* | *AC007601.1* |
| *AL390195.2* | *FGF6* | *AL355816.1* | *CFAP69* |
| *PPFIA2* | *SERPINE3* | *ZAP70* | *DOK7* |
| *RGPD1* | *AP000525.1* | *ACOX2* | *CRPPA-AS1* |
| *LGI1* | *AC021723.2* | *SIK1* | *HHIP* |
| *AL512380.2* | *AL139300.2* | *ZBTB8B* | *AC004053.1* |
| *AC079148.3* | *FAM149A* | *NUTM1* | *NLGN4X* |
| *CD40LG* | *AC069437.1* | *HIF3A* | *TTC9* |
| *TNIP3* | *CARMN* | *LINC00426* | *AC092839.1* |
| *WTIP* | *ANO3* | *ZNF32-AS3* | *AL353586.1* |
| *LINC02585* | *CARNS1* | *GZMA* | *F5* |
| *CCR2* | *CCDC63* | *BX284613.2* | |
| *AQP7* | *AC093866.1* | *LDLRAD2* | |
| *WSCD1* | *CATSPERB* | *AC025280.3* | |
| *LINC01840* | *HNF1A-AS1* | *ABCA13* | |

Table S6. TC - Single Nuclei RNA-seq Nuclei Distribution

All Conditions – Raw Counts

| Raw Counts after Reclustering | CTRL _Adt | ASyS _Jo-1 | ASyS _OJ | ASyS _PL7 | DM _MDA5 | DM _Mi-2 | DM _NXP-2 | DM _TIF1-γ | DM _SAE | IBM | IMNM _HMGCR | IMNM _SRP |
|---|---|---|---|---|---|---|---|---|---|---|---|---|
| iNKT-like_TC | 28 | 6 | 64 | 7 | 15 | 78 | 36 | 17 | 18 | 1256 | 48 | 26 |
| ZNF683+IL7R +TRM | 4 | 13 | 86 | 2 | 8 | 7 | 11 | 12 | 0 | 673 | 161 | 28 |
| NK-like_TC | 58 | 216 | 257 | 43 | 37 | 76 | 200 | 382 | 102 | 419 | 526 | 311 |
| Prolif_TC | 1 | 291 | 103 | 11 | 26 | 22 | 37 | 49 | 23 | 64 | 35 | 11 |
| Deg_TC | 11 | 2 | 23 | 5 | 12 | 24 | 25 | 38 | 17 | 28 | 16 | 0 |
| CX3CR1+ TRM | 21 | 49 | 63 | 23 | 31 | 8 | 30 | 85 | 36 | 14 | 28 | 57 |
| Dys_iNKT-like_TC | 81 | 0 | 0 | 1 | 0 | 0 | 0 | 1 | 0 | 10 | 2 | 0 |
| NOTCH1+ TRM | 0 | 315 | 13 | 4 | 0 | 2 | 2 | 9 | 1 | 9 | 14 | 13 |
| NKc | 1 | 67 | 75 | 4 | 40 | 30 | 32 | 34 | 56 | 8 | 41 | 31 |
| Ex_TC | 0 | 6 | 22 | 0 | 1 | 0 | 11 | 6 | 1 | 6 | 4 | 4 |
| TD_TC | 0 | 468 | 11 | 1 | 0 | 0 | 0 | 11 | 0 | 1 | 11 | 0 |
| St_TC | 14 | 1 | 5 | 0 | 15 | 2 | 6 | 2 | 0 | 1 | 22 | 1 |
| Total TC | 219 | 1434 | 722 | 101 | 185 | 249 | 390 | 646 | 254 | 2489 | 908 | 482 |

Table S7. TC - Single Nuclei RNA-seq Nuclei Distribution

All Conditions – Proportion

| % after Reclustering | CTRL _Adt | ASyS _Jo-1 | ASyS _OJ | ASyS _PL7 | DM _MDA5 | DM _Mi-2 | DM _NXP-2 | DM _TIF1-γ | DM _SAE | IBM | IMNM _HMGCR | IMNM _SRP |
|---|---|---|---|---|---|---|---|---|---|---|---|---|
| iNKT-like_TC | 12.79 | 0.42 | 8.87 | 6.93 | 8.11 | 31.33 | 9.23 | 2.63 | 7.09 | 50.46 | 5.29 | 5.39 |
| ZNF683+IL7R+ TRM | 1.83 | 0.91 | 11.91 | 1.98 | 4.32 | 2.81 | 2.82 | 1.86 | 0.00 | 27.04 | 17.73 | 5.81 |
| NK-like_TC | 26.48 | 15.06 | 35.60 | 42.57 | 20.00 | 30.52 | 51.28 | 59.13 | 40.16 | 16.83 | 57.93 | 64.52 |
| Prolif_TC | 0.46 | 20.29 | 14.27 | 10.89 | 14.05 | 8.84 | 9.49 | 7.59 | 9.06 | 2.57 | 3.85 | 2.28 |
| Deg_TC | 5.02 | 0.14 | 3.19 | 4.95 | 6.49 | 9.64 | 6.41 | 5.88 | 6.69 | 1.12 | 1.76 | 0.00 |
| CX3CR1+ TRM | 9.59 | 3.42 | 8.73 | 22.77 | 16.76 | 3.21 | 7.69 | 13.16 | 14.17 | 0.56 | 3.08 | 11.83 |
| Dys_iNKT-like_TC | 36.99 | 0.00 | 0.00 | 0.99 | 0.00 | 0.00 | 0.00 | 0.15 | 0.00 | 0.40 | 0.22 | 0.00 |
| NOTCH1+ TRM | 0.00 | 21.97 | 1.80 | 3.96 | 0.00 | 0.80 | 0.51 | 1.39 | 0.39 | 0.36 | 1.54 | 2.70 |
| NKc | 0.46 | 4.67 | 10.39 | 3.96 | 21.62 | 12.05 | 8.21 | 5.26 | 22.05 | 0.32 | 4.52 | 6.43 |
| Ex_TC | 0.00 | 0.42 | 3.05 | 0.00 | 0.54 | 0.00 | 2.82 | 0.93 | 0.39 | 0.24 | 0.44 | 0.83 |
| TD_TC | 0.00 | 32.64 | 1.52 | 0.99 | 0.00 | 0.00 | 0.00 | 1.70 | 0.00 | 0.04 | 1.21 | 0.00 |
| St_TC | 6.39 | 0.07 | 0.69 | 0.00 | 8.11 | 0.80 | 1.54 | 0.31 | 0.00 | 0.04 | 2.42 | 0.21 |
| Total TC | 100 | 100 | 100 | 100 | 100 | 100 | 100 | 100 | 100 | 100 | 100 | 100 |

Table S8. Differential Expression of Genes of IBM iNKT-like_TC vs. other Myositis and Controls. Upregulated Genes - Log2FC > 1 , adjusted *p* value < 0.05 , pct.2 < 0.05

| | | | |
|---|---|---|---|
| *AC002331.1* | *AC022929.2* | *AC013437.1* | *DSCAML1* |
| *CNBD1* | *AC113615.2* | *TNC* | *BATF* |
| *CTLA4* | *LINC00944* | *LINC02725* | *MPZL3* |
| *AC005599.1* | *GNAO1* | *HLA-DQA1* | *KLRC4* |
| *STYK1* | *BCAS4* | *DDX3Y* | *MRC1* |
| *AC079209.2* | *PLXDC1* | *RPS4Y1* | *GALNT6* |
| *ETV1* | *ADCY3* | *LINC01871* | *AL162493.1* |
| *NLGN4Y* | *P2RY10* | *AC024028.1* | *SMOC2* |
| *LINC00278* | *CRTAM* | *ATP10D* | *PPARG* |
| *LINC02446* | *KDM5D* | *AC090772.2* | *KLRC3* |
| *LY9* | *IL5RA* | *TSPAN2* | *MYADM* |
| *P2RY14* | *PARD6G* | *F13A1* | *SEMA3E* |
| *ZNF683* | *DUSP4* | *AL137779.1* | |
| *NUGGC* | *GPR132* | *ANKRD49* | |

Table S9. Differential Expression of Genes of IBM ZNF683+IL7R+TRM vs. other Myositis and Controls. Upregulated Genes - $Log_2FC > 1$ , adjusted $p$ value < 0.05

| | | | |
|---|---|---|---|
| *SLC27A2* | *LINC02446* | *RCAN2* | *MAP4K1* |
| *NGB* | *GPM6A* | *MMP25-AS1* | *ARSG* |
| *RGS18* | *EID1* | *COL10A1* | *OTUD5* |
| *LINC02195* | *AC022075.1* | *LINC00243* | *AC073342.1* |
| *MYO3B* | *AC113615.2* | *GNAO1* | *USP20* |
| *PTGER3* | *MFSD10* | *ZKSCAN7-AS1* | *PRTFDC1* |
| *XCL1* | *SLC24A3* | *SLC16A4* | *CD302* |
| *AC002331.1* | *TRNT1* | *D2HGDH* | *PIN4* |
| *AC004917.1* | *AL096794.1* | *KLRD1* | *AL133480.1* |
| *SPDYE5* | *LMO4* | *AC099063.1* | *COLGALT2* |
| *DUOX1* | *FGD5-AS1* | *ZNF683* | *MXD4* |
| *AC005599.1* | *AC018557.3* | *SPEF2* | *AGBL5* |
| *AL022238.2* | *PLXDC1* | *LINC01871* | *AC013652.1* |
| *IFNG* | *LINC02725* | *AL390957.1* | *GPSM2* |
| *IL5RA* | *AL591368.1* | *A2M* | *PKD2L2* |
| *FRMD5* | *PSTPIP2* | *CTSW* | *LINC02312* |
| *LINC02614* | *WDR63* | *CLNK* | *AL136171.2* |
| *CHPT1* | *NUGGC* | *MIAT* | *YPEL1* |
| *DALRD3* | *CERS4* | *PPM1M* | *AC025171.1* |
| *AC092902.4* | *TBXAS1* | *PZP* | *FCRL6* |
| *KLRC2* | *NHLRC3* | *SLC16A6* | *PCBP3* |
| *LONRF2* | *AC104078.2* | *AC011933.4* | *LY9* |
| *STYK1* | *ANKS3* | *GRIP1* | *GABPB2* |
| *MMP25* | *AC005856.1* | *CASS4* | *SPDYE2* |
| *LINC01479* | *CCZ1B* | *CARNMT1-AS1* | *SETBP1* |
| *LINC00892* | *AC022146.2* | *IFNG-AS1* | *AC017002.5* |
| *AC004825.3* | *AC004797.1* | *DSCAML1* | *LINC00623* |
| *AL158071.1* | *AC022929.2* | *RPF1* | *ACBD5* |
| *ETV1* | *BATF* | *DPYD-AS1* | *DTHD1* |
| *STMN3* | *GFI1* | *CYSLTR1* | *ZMYM5* |
| *ADORA2A-AS1* | *NLGN4Y* | *DAPK2* | *PTPRK* |
| *LINC00486* | *SPAG1* | *CP* | *MVB12B* |
| *PLXNA2* | *SLC24A1* | *AP000787.1* | *TG* |

| | | | |
|---|---|---|---|
| *ST8SIA1* | *KCTD7* | *CMTM1* | *OGFR* |
| *C12orf4* | *ZNF808* | *MOGS* | *PIK3CD-AS2* |
| *ACTA2* | *ID2* | *GSAP* | |
| *AC053513.1* | *CC2D1B* | *STOM* | |
| *BMERB1* | *AL365356.1* | *LINC00612* | |

Table S10. IBM vs. Controls only TC , Individual Sample Comparison – Raw Counts

| Nuclei Type | CTRL_1 | CTRL_2 | IBM_1 | IBM_2 | IBM_3 | IBM_4 |
|---|---|---|---|---|---|---|
| iNKT-like_TC | 14 | 14 | 424 | 752 | 10 | 70 |
| NK-like_TC | 15 | 43 | 29 | 10 | 137 | 243 |
| Prolif_TC | 0 | 1 | 23 | 12 | 20 | 9 |
| Deg_TC | 7 | 4 | 9 | 4 | 1 | 14 |
| ZNF683+IL7R+TRM | 3 | 1 | 6 | 7 | 469 | 191 |
| CX3CR1+TRM | 15 | 6 | 5 | 0 | 6 | 3 |
| Dys_iNKT-like_TC | 57 | 24 | 4 | 3 | 0 | 3 |
| NKc | 1 | 0 | 3 | 0 | 3 | 2 |
| NOTCH1+TRM | 0 | 0 | 2 | 0 | 6 | 1 |
| Ex_TC | 0 | 0 | 1 | 0 | 2 | 3 |
| St_TC | 14 | 0 | 0 | 1 | 0 | 0 |
| TD_TC | 0 | 0 | 0 | 0 | 1 | 0 |

Table S11. Individual Sample Information of Fig. 4A

| ID Number | Age Group | Sex | Duration | Biopsy Site |
|---|---|---|---|---|
| IBM_RISH1 | 71-80 | Male | >24 months | L BB |

L – left, BB – biceps brachii

Table S12. Individual Sample Information of Bulk RNA-seq patients

| Bulk RNAseq Label | Age Group | Sex | CK | Biopsy Site |
|---|---|---|---|---|
| CTRL_Adt_1 | 41-50 | Male | 116 | L BB |
| CTRL_Adt_2 | 21-30 | Male | 77 | L BB |
| CTRL_Adt_3 | 41-50 | Male | 320 | L BB |
| CTRL_Adt_4 | 31-40 | Male | 387 | L BB |
| ASyS_Jo-1_1 | 61-70 | Female | ND | L BB |
| ASyS_Jo-1_2 | 61-70 | Female | ND | L VL |
| ASyS_Jo-1_3 | 71-80 | Female | 5807 | R BB |
| ASyS_Jo-1_4 | 61-70 | Female | ND | L RF |
| ASyS_Jo-1_5 | 61-70 | Female | ND | L TA |
| ASyS_Jo-1_6 | 61-70 | Male | ND | L BB |
| ASyS_Jo-1_7 | 71-80 | Female | ND | RF |
| ASyS_Jo-1_8 | 31-40 | Female | ND | R BB |
| ASyS_Jo-1_9 | 51-60 | Male | ND | L QF |
| ASyS_Jo-1_10 | 51-60 | Male | 2956 | L RF |
| ASyS_Jo-1_11 | 71-80 | Female | 1757 | L BB |
| ASyS_Jo-1_12 | 51-60 | Female | 1245 | L QF |
| ASyS_Jo-1_13 | 41-50 | Male | 4511 | L VL |
| ASyS_OJ_1 | 71-80 | Male | ND | L BB |
| ASyS_OJ_2 | 31-40 | Male | ND | L QF |
| ASyS_OJ_3 | 71-80 | Female | ND | R BB |
| ASyS_OJ_4 | 51-60 | Female | ND | L BB |
| ASyS_OJ_5 | 61-70 | Male | ND | L BB |
| ASyS_OJ_6 | 71-80 | Male | ND | R RF |
| ASyS_OJ_7 | 71-80 | Male | ND | R Del |
| ASyS_OJ_8 | 61-70 | Male | ND | L RF |

| | | | | |
|---|---|---|---|---|
| ASyS_OJ_9 | 61-70 | Female | ND | L BB |
| ASyS_OJ_10 | 61-70 | Male | ND | L BB |
| ASyS_OJ_11 | 61-70 | Female | 2805 | L QF |
| ASyS_OJ_12 | 71-80 | Female | 9202 | R RF |
| ASyS_OJ_13 | 81-90 | Female | 1236 | L BB |
| DM_MDA5_1 | 51-60 | Female | 402 | L BB |
| DM_MDA5_2 | 51-60 | Female | 656 | L Del |
| DM_MDA5_3 | 61-70 | Female | 77 | R QF |
| DM_MDA5_4 | 61-70 | Female | 407 | L BB |
| DM_MDA5_5 | 61-70 | Male | 759 | R VL |
| DM_MDA5_6 | 41-50 | Male | 586 | L BB |
| DM_MDA5_7 | 41-50 | Male | 519 | R VL |
| DM_MDA5_8 | 41-50 | Female | ND | R VL |
| DM_MDA5_9 | 61-70 | Female | ND | L BB |
| DM_MDA5_10 | 71-80 | Female | ND | L BB |
| DM_MDA5_11 | 41-50 | Male | 899 | L QF |
| DM_MDA5_12 | 41-50 | Male | ND | L BB |
| DM_Mi-2_1 | 51-60 | Female | ND | L BB |
| DM_Mi-2_2 | 21-30 | Male | 8131 | R BB |
| DM_Mi-2_3 | 51-60 | Female | ND | L QF |
| DM_Mi-2_4 | 61-70 | Male | 2434 | L BB |
| DM_Mi-2_5 | 41-50 | Male | ND | R BB |
| DM_Mi-2_6 | 51-60 | Male | ND | L TB |
| DM_Mi-2_7 | 61-70 | Male | ND | R VL |
| DM_Mi-2_8 | 21-30 | Female | ND | R BB |
| DM_Mi-2_9 | 71-80 | Male | 3343 | L BB |
| DM_Mi-2_10 | 61-70 | Female | 5454 | L RF |
| DM_Mi-2_11 | 61-70 | Female | 5080 | L BB |
| DM_Mi-2_12 | 41-50 | Male | 7844 | R BB |

| | | | | |
|---|---|---|---|---|
| DM_Mi-2_13 | 51-60 | Male | 13355 | R BB |
| DM_NXP-2_1 | 31-40 | Male | ND | L BB |
| DM_NXP-2_2 | 61-70 | Male | ND | R BB |
| DM_NXP-2_3 | 31-40 | Male | 195 | L Del |
| DM_NXP-2_4 | 61-70 | Female | 584 | L Del |
| DM_NXP-2_5 | 31-40 | Male | 23244 | R QF |
| DM_NXP-2_6 | 51-60 | Female | 433 | R BB |
| DM_NXP-2_7 | 61-70 | Female | 14365 | R VL |
| DM_NXP-2_8 | 31-40 | Male | 417 | R BB |
| DM_NXP-2_9 | 51-60 | Female | 250 | R VL |
| DM_NXP-2_10 | 51-60 | Female | 2202 | L BB |
| DM_NXP-2_11 | 41-50 | Female | 1492 | R TB |
| DM_NXP-2_12 | 31-40 | Male | 45093 | L BB |
| DM_TIF1-γ_1 | 71-80 | Female | 271 | L BB |
| DM_TIF1-γ_2 | 71-80 | Female | 179 | R QF |
| DM_TIF1-γ_3 | 51-60 | Male | 298 | R QF |
| DM_TIF1-γ_4 | 71-80 | Female | 96 | L BB |
| DM_TIF1-γ_5 | 61-70 | Male | 1581 | L VL |
| DM_TIF1-γ_6 | 71-80 | Male | 5007 | L BB |
| DM_TIF1-γ_7 | 41-50 | Female | 713 | R TB |
| DM_TIF1-γ_8 | 61-70 | Female | 309 | L VL |
| DM_TIF1-γ_9 | 61-70 | Female | 234 | L BB |
| DM_TIF1-γ_10 | 61-70 | Female | ND | L BB |
| DM_TIF1-γ_11 | 61-70 | Male | ND | L Del |
| DM_TIF1-γ_12 | 41-50 | Female | 122 | L VL |
| DM_TIF1-γ_13 | 61-70 | Male | 377 | L BB |
| DM_TIF1-γ_14 | 71-80 | Female | 1102 | R Del |
| DM_TIF1-γ_15 | 81-90 | Male | 447 | R BB |
| DM_TIF1-γ_16 | 21-30 | F | 317 | L RF |
| IBM_1 | 71-80 | Male | 476 | R VL |

| | | | | |
|---|---|---|---|---|
| IBM_2 | 51-60 | Male | 2917 | L BB |
| IBM_3 | 61-70 | Female | 544 | L RF |
| IBM_4 | 71-80 | Female | 277 | L BB |
| IBM_5 | 61-70 | Male | 1138 | L QF |
| IBM_6 | 61-70 | Female | 364 | R RF |
| IBM_7 | 61-70 | Male | 1969 | L Gc |
| IBM_8 | 51-60 | Male | 405 | L VL |
| IBM_9 | 71-80 | Male | 722 | R BB |
| IBM_10 | 71-80 | Male | 90 | L BB |
| IBM_11 | 71-80 | Male | 534 | R QF |
| IBM_12 | 61-70 | Male | 746 | R QF |
| IMNM_HMGCR_1 | 51-60 | Male | 10061 | L BB |
| IMNM_HMGCR_2 | 51-60 | Female | 14910 | L BB |
| IMNM_HMGCR_3 | 71-80 | Female | 1801 | L BB |
| IMNM_HMGCR_4 | 61-70 | Female | 5942 | L BB |
| IMNM_HMGCR_5 | 61-70 | Male | 1208 | L SM |
| IMNM_HMGCR_6 | 71-80 | Male | 10022 | L BB |
| IMNM_HMGCR_7 | 51-60 | Male | 2723 | L TB |
| IMNM_HMGCR_8 | 61-70 | Female | 4267 | L BB |
| IMNM_HMGCR_9 | 51-60 | Female | 7613 | L BB |
| IMNM_HMGCR_10 | 21-30 | Female | 2104 | L BB |
| IMNM_HMGCR_11 | 71-80 | Male | 3283 | L BB |
| IMNM_SRP_1 | 51-60 | Male | 5624 | R QF |
| IMNM_SRP_2 | 71-80 | Male | 8656 | L RF |
| IMNM_SRP_3 | 41-50 | Male | 1428 | L Del |
| IMNM_SRP_4 | 51-60 | Male | 3497 | L VL |
| IMNM_SRP_5 | 71-80 | Female | 7600 | L BB |
| IMNM_SRP_6 | 71-80 | Female | 6794 | L VL |
| IMNM_SRP_7 | 61-70 | Female | 67740 | BB |
| IMNM_SRP_8 | 41-50 | Male | 2083 | L Del |

| | | | | |
|---|---|---|---|---|
| IMNM_SRP_9 | 51-60 | Female | 2387 | R AM R St |
| IMNM_SRP_10 | 61-70 | Female | 304 | R QF |
| IMNM_SRP_11 | 61-70 | Male | 3297 | L QF |

ND – no data; R – right; L- left; AM – adductor magnus; BB – biceps brachii; Del – deltoid; Gc – gastrocnemius; QF – quadriceps femoris; RF – rectus femoris; Sm – semimembranosus; St – semitendinosus; TA – tibialis anterior; TB – triceps brachii; VL – vastus lateralis

Table S13. Bulk RNA-seq $Log_2FC$ vs. Condition CD8 T Cells Gene Signature

| GENE SYMBOL | DM_MDA5. $Log_2FC$ | DM_MDA5. padj | DM_Mi-2. $Log_2FC$ | DM_Mi-2. padj | DM_NXP-2. $Log_2FC$ | DM_NXP-2. padj | DM_TIF1-γ. $Log_2FC$ | DM_TIF1-γ. padj |
|---|---|---|---|---|---|---|---|---|
| *ZNF683* | 2.01E-06 | 0.04557235 | 5.832239462 | 6.29E-05 | 1.81E-06 | 0.065145 | 3.072 | 0.0029107 |
| *CD8A* | 1.08E-06 | 0.38177315 | 2.959400679 | 6.62E-06 | 3.18E-06 | 0.044078 | 1.004 | 0.0571449 |
| *CD247* | 1.64E-06 | 0.70188827 | 1.913174104 | 1.49E-07 | 3.32E-06 | 0.138268 | 1.097 | 0.0082395 |
| *CD28* | 1.8588884 | 2.88E-06 | 4.222606104 | 6.27E-27 | 2.7368634 | 6.40E-06 | 2.318 | 4.13E-08 |
| *KLRC4-KLRK1* | 1.72E-06 | 0.42810353 | 1.33900115 | 0.0717309 | 6.47E-07 | 0.616542 | 0.468 | 0.388496 |
| *KLRC3* | 4.09E-07 | NA | 2.028608504 | 0.0218632 | 5.27E-07 | NA | 0.587 | 0.2540697 |
| *KLRC2* | 4.00E-07 | NA | 1.130003459 | 0.13737 | 1.24E-06 | 0.347234 | 1.024 | 0.1279665 |
| *KLRC4* | -9.94E-07 | NA | 0.747513382 | 0.2570633 | -1.21E-07 | 0.940402 | -0.103 | 0.8504023 |
| *KLRC1* | 2.82E-06 | 0.0438524 | 3.894477628 | 3.12E-06 | 3.2512605 | 0.000795 | 2.901 | 0.0014348 |
| *KLRD1* | 0.6799036 | 0.02421557 | 2.280512565 | 9.51E-09 | 1.4932099 | 0.000842 | 1.727 | 6.64E-07 |
| *KLRB1* | -3.05E-06 | 0.45036335 | 1.032350873 | 0.0410265 | 1.83E-06 | 0.426087 | 0.147 | 0.7761351 |
| *IFNG* | 1.62E-06 | 0.49331426 | 1.470719834 | 0.0351982 | 1.81E-06 | 0.137707 | 0.617 | 0.2323463 |
| *XCL1* | 2.97E-06 | 0.10717001 | 2.650150933 | 0.0018173 | 2.42E-06 | 0.07062 | 1.341 | 0.0408021 |
| *CCL4* | 3.57E-06 | 0.05887 | 2.378075302 | 4.75E-05 | 1.5179874 | 0.008835 | 1.015 | 0.0372489 |
| *CCL5* | 3.2347607 | 0.00011298 | 4.182559806 | 1.52E-15 | 3.1941422 | 5.64E-06 | 3.277 | 1.63E-11 |
| *GZMK* | 2.57E-06 | 0.20827401 | 2.878342456 | 3.40E-05 | 3.46E-06 | 0.020745 | 1.201 | 0.0473856 |
| *GZMA* | 4.59E-06 | 0.12153366 | 2.640675438 | 1.05E-06 | 1.7452566 | 0.003112 | 1.565 | 0.0044273 |
| *GZMH* | 1.87E-06 | 0.43362587 | 2.421342489 | 2.06E-05 | 4.08E-06 | 0.03476 | 1.93 | 0.0001331 |
| *GZMB* | 2.6055443 | 0.0001488 | 3.699528676 | 1.28E-17 | 3.0193445 | 4.14E-06 | 3.159 | 2.47E-08 |
| *GZMM* | 6.49E-07 | 0.76520045 | 2.207279775 | 9.07E-06 | 1.47E-06 | 0.429261 | 1.703 | 0.0015294 |
| *PRF1* | 4.08E-06 | 0.03271206 | 2.615850702 | 1.27E-18 | 1.4625591 | 0.003174 | 2.339 | 1.89E-08 |

| GENE SYMBOL | ASyS_OJ. Log2FC | ASyS_OJ. padj | ASyS _Jo-1. Log2FC | ASyS _Jo-1. padj | IMNM _HMGCR. Log2FC | IMNM _HMGCR. padj | IMNM _SRP. Log2FC | IMNM _SRP. padj | IBM. Log2FC | IBM. padj |
|---|---|---|---|---|---|---|---|---|---|---|
| *ZNF683* | 5.27945 | 1.73E-05 | 2.79892 | 0.009434 | 2.65900 | 0.009435 | 2.811 | 0.0642 | 8.4844 | 1.68E-09 |
| *CD8A* | 4.05057 | 6.95E-16 | 4.11787 | 4.86E-17 | 1.91784 | 0.000898 | 1.229 | 0.0396 | 5.1028 | 3.71E-22 |
| *CD247* | 1.95261 | 1.83E-07 | 1.95478 | 7.71E-06 | 0.45813 | 0.202289 | 0.475 | 0.2633 | 2.5547 | 1.26E-08 |
| *CD28* | 4.84858 | 1.26E-19 | 4.73412 | 2.80E-14 | 4.36127 | 6.50E-17 | 4.402 | 1.47E-15 | 4.4011 | 1.42E-22 |
| *KLRC4-KLRK1* | 1.47872 | 0.033621 | 2.61598 | 0.001693 | 2.10454 | 0.008148 | 1.6 | 0.0284 | 4.2582 | 8.39E-08 |
| *KLRC3* | 1.96887 | 0.021228 | 1.45623 | 0.064793 | 0.59916 | 0.213303 | 0.786 | 0.1566 | 3.888 | 0.0001 |
| *KLRC2* | 1.53483 | 0.052514 | 1.65326 | 0.051107 | 0.63403 | 0.213117 | 0.354 | 0.4485 | 3.3553 | 0.0006 |
| *KLRC4* | 1.35973 | 0.034362 | 0.97558 | 0.123720 | 0.17178 | 0.720181 | 0.269 | 0.5882 | 2.8199 | 9.28E-05 |
| *KLRC1* | 4.10009 | 2.10E-05 | 4.66309 | 1.77E-05 | 2.47772 | 0.004809 | 2.442 | 0.0048 | 4.6368 | 4.30E-06 |
| *KLRD1* | 2.36819 | 1.16E-09 | 2.47595 | 4.40E-07 | 1.26840 | 0.005574 | 1.248 | 0.0078 | 2.7065 | 1.61E-08 |
| *KLRB1* | 1.60954 | 0.000183 | 1.55244 | 0.001592 | 1.11117 | 0.023461 | 0.748 | 0.1096 | 2.8094 | 1.92E-08 |
| *IFNG* | 3.20563 | 1.58E-05 | 4.21888 | 2.29E-09 | 1.31335 | 0.032691 | 0.594 | 0.2458 | 4.9599 | 4.68E-13 |
| *XCL1* | 2.62970 | 0.000159 | 2.31800 | 0.002427 | 1.91184 | 0.013481 | 1.412 | 0.0436 | 5.1848 | 1.67E-09 |
| *CCL4* | 3.37828 | 1.39E-10 | 3.53476 | 2.36E-09 | 2.32359 | 0.000205 | 1.783 | 0.0036 | 3.65 | 1.83E-09 |
| *CCL5* | 4.91921 | 3.14E-26 | 4.61509 | 1.89E-19 | 2.89913 | 4.17E-07 | 2.408 | 0.0002 | 5.8295 | 5.18E-26 |
| *GZMK* | 3.91096 | 5.86E-13 | 4.37978 | 4.28E-14 | 2.74390 | 2.35E-05 | 1.827 | 0.0060 | 5.0399 | 9.07E-24 |
| *GZMA* | 3.57602 | 1.32E-13 | 4.31164 | 1.35E-17 | 2.48679 | 5.82E-05 | 1.712 | 0.0038 | 4.882 | 7.25E-28 |
| *GZMH* | 3.57996 | 4.23E-16 | 3.33042 | 1.40E-11 | 1.59840 | 0.001831 | 1.207 | 0.0187 | 4.6967 | 3.84E-17 |
| *GZMB* | 3.68197 | 8.61E-14 | 3.99426 | 2.19E-14 | 0.70182 | 0.123478 | 1.128 | 0.0237 | 4.0149 | 1.77E-10 |
| *GZMM* | 2.45657 | 1.62E-07 | 1.75779 | 0.01213 | 0.02323 | 0.966865 | 0.177 | 0.3917 | 3.3275 | 4.35E-05 |
| *PRF1* | 2.45290 | 1.03E-12 | 2.28117 | 2.11E-08 | 0.20680 | 0.565175 | 0.346 | 0.4372 | 2.5465 | 4.77E-06 |

Table S14. Grouped Kruskal-Wallis Comparison per CD8 T Cell Gene Set per Condition

Method - Kruskal-Wallis rank sum test, $\chi2$ =51.89, Statistic df = 8, $p$-value =1.77× $10^{-8}$

Post-hoc Wilcoxon Rank-sum Test

| Condition1 | Condition2 | W_stat | p_value | adj_p_value |
|---|---|---|---|---|
| IBM | ASyS_Jo-1 | 270 | 0.02334 | 0.02334 |
| IBM | ASyS_OJ | 307 | 0.011839 | 0.01353 |
| IBM | DM_MDA5 | 119 | 0.001209 | 0.001612 |
| IBM | DM_Mi-2 | 305 | 0.001139 | 0.001612 |
| IBM | DM_NXP-2 | 213 | 0.000119 | 0.000238 |
| IBM | DM_TIF1-γ | 316 | 6.13E-06 | 4.90E-05 |
| IBM | IMNM_HMGCR | 273 | 2.38E-05 | 6.35E-05 |
| IBM | IMNM_SRP | 257 | 2.11E-05 | 6.35E-05 |

Table S15. Comparison of *ZNF683* Counts per Million (CPM) in IBM Bulk RNA-seq Samples ≤24 months versus > 24 months.

Method - Wilcoxon rank sum test with continuity correction, W = 6, *p*-value = 0.048.

| ID Number | Age Group | Sex | Duration | CPM |
|---|---|---|---|---|
| IBM_B1 | 61-70 | Male | ≤24 months | 53.53 |
| IBM_B2 | 71-80 | Male | ≤24 months | 1.99 |
| IBM_B3 | 61-70 | Male | ≤24 months | 23.66 |
| IBM_B4 | 51-60 | Male | ≤24 months | 54.37 |
| IBM_B5 | 51-60 | Male | ≤24 months | 766.03 |
| IBM_B6 | 61-70 | Female | > 24 months | 269.92 |
| IBM_B7 | 61-70 | Male | > 24 months | 871.88 |
| IBM_B8 | 71-80 | Female | > 24 months | 69.35 |
| IBM_B9 | 71-80 | Male | > 24 months | 72.84 |
| IBM_B10 | 61-70 | Male | > 24 months | 379.72 |
| IBM_B11 | 71-80 | Male | > 24 months | 272.46 |
| IBM_B12 | 71-80 | Female | > 24 months | 875.90 |
| IBM_B13 | 71-80 | Male | > 24 months | 164.72 |

Shapiro-Wilk normality test $p < 0.05$ for ≤24 months and > 24 months

Table S16. Comparison of *ZNF683* $Log_2FC$ in RT-PCR Samples between IBM versus other conditions

| Condition | RT-PCR Label | Age Group | Sex | CK (U/L) | Log2FC |
|---|---|---|---|---|---|
| Nondisease_control | CTRL_Adt_1 | 41-50 | Male | 103 | 0.97056055 |
| Nondisease_control | CTRL_Adt_2 | 21-30 | Male | 163 | 0.00010854 |
| Disease_control VCP-M | VCP-M _1 | 51-60 | Female | 128 | 13.485507 |
| Disease_control VCP-M | VCP-M _2 | 51-60 | Male | 310 | 19.6623422 |
| Disease_control VCP-M | VCP-M _3 | 71-80 | Female | 103 | 34.2167326 |
| Disease_control VCP-M | VCP-M _4 | 71-80 | Female | 9373 | 16.8792953 |
| ASyS_Jo-1 | ASyS_Jo-1_1 | 51-60 | Female | 1303 | 1.57577667 |
| ASyS_Jo-1 | ASyS_Jo-1_2 | 71-80 | Male | 729 | 3.13615803 |
| ASyS_Jo-1 | ASyS_Jo-1_3 | 51-60 | Male | 3012 | 1.47119621 |
| ASyS_Jo-1 | ASyS_Jo-1_4 | 51-60 | Female | 878 | 5.92469934 |
| ASyS_Jo-1 | ASyS_Jo-1_5 | 41-50 | Female | 9992 | 6.50430556 |
| ASyS_Jo-1 | ASyS_Jo-1_6 | 11-20 | Female | 3092 | 8.96208008 |
| ASyS_Jo-1 | ASyS_Jo-1_7 | 31-40 | Female | 615 | 2.30483926 |
| ASyS_Jo-1 | ASyS_Jo-1_8 | 31-40 | Male | 3780 | 5.70114403 |
| ASyS_Jo-1 | ASyS_Jo-1_9 | 41-50 | Male | 10823 | 21.0664231 |
| ASyS_Jo-1 | ASyS_Jo-1_10 | 71-80 | Male | 2336 | 21.3818664 |
| ASyS_OJ | ASyS_OJ_1 | 71-80 | Female | 1331 | 10.6646054 |
| ASyS_OJ | ASyS_OJ_2 | 41-50 | Female | 3029 | 9.45456227 |
| ASyS_OJ | ASyS_OJ_3 | 71-80 | Male | 4204 | 0.88635371 |
| ASyS_OJ | ASyS_OJ_4 | 41-50 | Male | 20015 | 2.76975253 |
| ASyS_OJ | ASyS_OJ_5 | 71-80 | Female | 5813 | 16.3263346 |
| ASyS_OJ | ASyS_OJ_6 | 71-80 | Female | 1510 | 7.35849021 |
| ASyS_OJ | ASyS_OJ_7 | 71-80 | Female | 1202 | 3.03270467 |
| ASyS_OJ | ASyS_OJ_8 | 41-50 | Male | 1678 | 1.21282695 |
| ASyS_OJ | ASyS_OJ_9 | 91-100 | Male | 2240 | 3.80873961 |
| ASyS_OJ | ASyS_OJ_10 | 71-80 | Male | 3073 | 7.54839605 |
| DM_Mi-2 | DM_Mi-2_1 | 81-90 | Male | 7842 | 64.6333083 |
| DM_Mi-2 | DM_Mi-2_2 | 11-20 | Female | 8132 | 7.38552652 |

| | | | | | |
|---|---|---|---|---|---|
| DM_Mi-2 | DM_Mi-2_3 | 21-30 | Female | 7718 | 19.706844 |
| DM_Mi-2 | DM_Mi-2_4 | 71-80 | Female | 5830 | 7.18874219 |
| DM_Mi-2 | DM_Mi-2_5 | 21-30 | Male | 11164 | 9.83023532 |
| DM_Mi-2 | DM_Mi-2_6 | 41-50 | Male | 843 | 19.3376708 |
| DM_Mi-2 | DM_Mi-2_7 | 81-90 | Male | 692 | 415.067101 |
| DM_Mi-2 | DM_Mi-2_8 | 11-20 | Female | 233 | 136.936613 |
| DM_Mi-2 | DM_Mi-2_9 | 51-60 | Male | 654 | 4.26932352 |
| DM_NXP-2 | DM_NXP-2_1 | 1-10 | Female | 725 | 20.5989937 |
| DM_NXP-2 | DM_NXP-2_2 | 31-40 | Male | 966 | 21.5672011 |
| DM_NXP-2 | DM_NXP-2_3 | 61-70 | Female | 8602 | 35.7466149 |
| DM_NXP-2 | DM_NXP-2_4 | 31-40 | Female | 654 | 126.390214 |
| DM_NXP-2 | DM_NXP-2_5 | 51-60 | Female | 261 | 21.9006798 |
| DM_SAE | DM_SAE_1 | 61-70 | Female | 232 | 5.68287273 |
| DM_SAE | DM_SAE_2 | 71-80 | Female | 226 | 87.6589086 |
| DM_SAE | DM_SAE_3 | 51-60 | Female | 514 | 9.65871666 |
| DM_SAE | DM_SAE_4 | 61-70 | Female | 240 | 14.2405035 |
| DM_SAE | DM_SAE_5 | 71-80 | Male | 696 | 6.92093973 |
| DM_SAE | DM_SAE_6 | 81-90 | Female | 248 | 2.28012912 |
| DM_SAE | DM_SAE_7 | 71-80 | Female | 181 | 10.5013422 |
| DM_TIF1-γ | DM_TIF1-γ_1 | 61-70 | Female | 880 | 23.2825901 |
| DM_TIF1-γ | DM_TIF1-γ_2 | 51-60 | Male | 14293 | 14.178791 |
| DM_TIF1-γ | DM_TIF1-γ_3 | 41-50 | Female | 695 | 28.2053296 |
| IBM | IBM_1 | 51-60 | Female | 934 | 1242.261585 |
| IBM | IBM_2 | 71-80 | Female | 406 | 1939.238035 |
| IBM | IBM_3 | 61-70 | Female | 157 | 2075.903436 |
| IBM | IBM_4 | 71-80 | Female | 346 | 859.4060943 |
| IBM | IBM_5 | 71-80 | Female | 630 | 180.2737162 |
| IBM | IBM_6 | 71-80 | Female | 1033 | 163.8473809 |
| IBM | IBM_7 | 71-80 | Male | 90 | 208.5412977 |
| IBM | IBM_8 | 71-80 | Female | 579 | 295.1243634 |
| IBM | IBM_9 | 61-70 | Male | 743 | 224.2179209 |
| IBM | IBM_10 | 51-60 | Female | 254 | 229.2912912 |
| IBM | IBM_11 | 51-60 | Male | 2024 | 241.8349763 |
| IMNM_HMGCR | IMNM_HMGCR_1 | 31-40 | Female | 2771 | 11.9095665 |
| IMNM_HMGCR | IMNM_HMGCR_2 | 41-50 | Female | 3375 | 5.24637591 |

| | | | | | |
|---|---|---|---|---|---|
| IMNM_HMGCR | IMNM_HMGCR_3 | 61-70 | Female | 4401 | 19.23494 |
| IMNM_HMGCR | IMNM_HMGCR_4 | 71-80 | Female | 4118 | 21.3861071 |

VCP-M – valosin-containing protein associated myopathy

Table S17. Statistical Analysis - Comparison of *ZNF683* $Log_2FC$ in RT-PCR Samples between IBM versus other conditions

Method - Kruskal-Wallis rank sum test, $\chi2$= 44.258 , Statistic df= 9, $p = 1.27 \times 10^{-6}$

Post-hoc Pairwise Comparison

| Condition1 | Condition2 | W_stat | p_value | padj |
|---|---|---|---|---|
| IBM | VCP | 44 | 0.005 | 0.006429 |
| IBM | DM_NXP-2 | 55 | 0.002222 | 0.004 |
| IBM | DM_TIF1-γ | 33 | 0.012731 | 0.014323 |
| IBM | DM_Mi-2 | 92 | 0.001418 | 0.003191 |
| IBM | DM_SAE | 77 | 0.000578 | 0.001735 |
| IBM | ASyS_Jo-1 | 110 | 0.000124 | 0.000559 |
| IBM | IMNM_HMGCR | 44 | 0.005 | 0.006429 |
| IBM | ASyS_OJ | 110 | 0.000124 | 0.000559 |
| IBM | CTRL_Adt | 22 | 0.038214 | 0.038214 |

Table S18. Comparison of *ZNF683* Log2FC in IBM RT-PCR Samples ≤24 months versus > 24 months

Method - Welch Two Sample t-test, t = −2.508, df = 8.016, *p*-value = 0.036, 95% CI = −1229.882 to −51.734

| Condition | RT-PCR Label | Duration | Age Group | Sex | CK (U/L) | $Log_2FC$ |
|---|---|---|---|---|---|---|
| IBM | IBM_1 | > 24 months | 51-60 | Female | 934 | 1242.261585 |
| IBM | IBM_2 | > 24 months | 71-80 | Female | 406 | 1939.238035 |
| IBM | IBM_3 | > 24 months | 61-70 | Female | 157 | 2075.903436 |
| IBM | IBM_4 | > 24 months | 71-80 | Female | 346 | 859.4060943 |
| IBM | IBM_5 | ≤24 months | 71-80 | Female | 630 | 180.2737162 |
| IBM | IBM_6 | ≤24 months | 71-80 | Female | 1033 | 163.8473809 |
| IBM | IBM_7 | > 24 months | 71-80 | Male | 90 | 208.5412977 |
| IBM | IBM_8 | > 24 months | 71-80 | Female | 579 | 295.1243634 |
| IBM | IBM_9 | > 24 months | 61-70 | Male | 743 | 224.2179209 |
| IBM | IBM_10 | > 24 months | 51-60 | Female | 254 | 229.2912912 |
| IBM | IBM_11 | > 24 months | 51-60 | Male | 2024 | 241.8349763 |

Table S19. Binary ROC analysis of *ZNF683* RT-qPCR $log_2FC$ across conditions

RT-qPCR $log_2FC$ scores were coded as 0 for ≤200 and 1 for >200. Each condition was compared with all remaining samples in table S16 (total n = 65), with higher scores treated as positive for every comparison. For IBM, the 54 non-IBM comparators comprise 48 other AM, 4 VCP-M, and 2 nondisease adult controls. Expression thresholds remain mild (>50), moderate (>100), and marked (>200).

| Condition (n) vs. remaining (n) | AUC | 95% DeLong CI | Sensitivity | Specificity |
|---|---|---|---|---|
| IBM (11) vs. remaining (54) | 0.900 | 0.779–1.000 | 81.818% (9/11) | 98.148% (53/54) |
| VCP-M (4) vs. remaining (61) | 0.418 | 0.371–0.465 | 0.000% (0/4) | 83.607% (51/61) |
| IMNM_HMGCR (4) vs. remaining (61) | 0.418 | 0.371–0.465 | 0.000% (0/4) | 83.607% (51/61) |
| ASyS_OJ (10) vs. remaining (55) | 0.409 | 0.358–0.461 | 0.000% (0/10) | 81.818% (45/55) |
| ASyS_Jo-1 (10) vs. remaining (55) | 0.409 | 0.358–0.461 | 0.000% (0/10) | 81.818% (45/55) |
| DM_SAE (7) vs. remaining (58) | 0.414 | 0.365–0.463 | 0.000% (0/7) | 82.759% (48/58) |
| DM_TIF1-γ (3) vs. remaining (62) | 0.419 | 0.373–0.466 | 0.000% (0/3) | 83.871% (52/62) |
| DM_NXP-2 (5) vs. remaining (60) | 0.417 | 0.369–0.464 | 0.000% (0/5) | 83.333% (50/60) |
| DM_Mi-2 (9) vs. remaining (56) | 0.475 | 0.356–0.594 | 11.111% (1/9) | 83.929% (47/56) |
| Nondisease controls (2) vs. remaining (63) | 0.421 | 0.375–0.466 | 0.000% (0/2) | 84.127% (53/63) |

For the IBM comparison at the >200 cutoff, 9 IBM and 1 non-IBM samples were positive, and 2 IBM and 53 non-IBM samples were negative. Confidence intervals use the DeLong normal approximation and are constrained to 0–1. The IBM upper bound was capped at 1. The fixed high-score-positive direction is retained even when an AUC is below 0.5. Overlapping curves in Figure 4J are listed separately here.

Table S20. Protein Expression Comparison for IBM Samples only

| Antibody Tested | Disease Duration | Number of areas (20x magnification)/ samples | Age Group | Male / Female Distribution | CK (U/L) | Biopsy Site Frequency |
|---|---|---|---|---|---|---|
| PLZF / CD8A ratio | ≤24 months | 5/4 | 51-60 to 71-80 | 0/4 | 145-6364 | L GC, L RF, R QF, L VL |
| PLZF / CD8A ratio | > 24 months | 5/4 | 61-70 to 71-80 | 4/0 | 294-895 | R VL > L QF, L RF |
| KLRK1 / CD8A ratio | ≤24 months | 5/4 | 51-60 to 71-80 | 0/4 | 145-6364 | R QF > L RF, L GC |
| KLRK1 / CD8A ratio | > 24 months | 5/3 | 71-80 | 1/2 | 294-750 | R VL, L VL, L RF |
| ZNF683 / CD8A ratio | ≤24 months | 5/2 | 51-60 to 61-70 | 0/2 | 145-786 | L GC, L RF |
| ZNF683/ CD8A ratio | > 24 months | 5/2 | 51-60 to 71-80 | 1/1 | 90-1784 | L BB, L VL |

L – left, R – right, Gc– gastrocnemius; RF – rectus femoris; QF – quadriceps femoris; VL – vastus lateralis; BB – biceps brachii

Table S21. SRA Run Number and *ZNF683* Status Public Dataset from Jurkat Cell *ZNF683* Overexpression Experiment

GEO Accession: GSM7713745. Series: GSE241091

| SRA Run Number | *ZNF683* Status |
| --- | --- |
| SRR25658158 | DOX + (overexpression) |
| SRR25658160 | DOX + (overexpression) |
| SRR25658161 | DOX + (overexpression) |
| SRR25658156 | DOX – (no overexpression) |
| SRR25658159 | DOX – (no overexpression) |

Table S22. Upregulated Genes from DOX+ Jurkat Cells - Public Dataset from Jurkat Cell *ZNF683* Overexpression Experiment

GEO Accession: GSM7713745. Series: GSE241091. Upregulated Genes from DOX+ Jurkat Cells. $Log_2FC > 1$ , adjusted $p$ value $< 0.05$

| | | | |
|---|---|---|---|
| ENSG00000139292.13 | ENSG00000304693.1 | ENSG00000249881.2 | ENSG00000243970.6 |
| ENSG00000259332.3 | ENSG00000234311.2 | ENSG00000294776.1 | ENSG00000304201.1 |
| ENSG00000143816.8 | ENSG00000179546.5 | ENSG00000252919.1 | ENSG00000297638.1 |
| ENSG00000232598.2 | ENSG00000212402.1 | ENSG00000276696.1 | ENSG00000287335.3 |
| ENSG00000132692.19 | ENSG00000255526.6 | ENSG00000307085.1 | ENSG00000293509.2 |
| ENSG00000297413.1 | ENSG00000264607.1 | ENSG00000119535.20 | ENSG00000272989.1 |
| ENSG00000299259.1 | ENSG00000236327.2 | ENSG00000202360.1 | ENSG00000278202.1 |
| ENSG00000173083.16 | ENSG00000298384.1 | ENSG00000205832.8 | ENSG00000236188.1 |
| ENSG00000295604.1 | ENSG00000287493.1 | ENSG00000167711.14 | ENSG00000290840.2 |
| ENSG00000176040.14 | ENSG00000242551.2 | ENSG00000307621.1 | ENSG00000294751.1 |
| ENSG00000187796.16 | ENSG00000293795.1 | ENSG00000254781.1 | ENSG00000253238.4 |
| ENSG00000303098.1 | ENSG00000188223.9 | ENSG00000301487.1 | ENSG00000227445.1 |
| ENSG00000304637.1 | ENSG00000307994.1 | ENSG00000255503.1 | ENSG00000244380.2 |
| ENSG00000298875.1 | ENSG00000242282.7 | ENSG00000295200.1 | ENSG00000196600.13 |
| ENSG00000227834.1 | ENSG00000106236.4 | ENSG00000238829.1 | ENSG00000262624.2 |
| ENSG00000286088.1 | ENSG00000287467.2 | ENSG00000271741.1 | ENSG00000267986.1 |
| ENSG00000273682.1 | ENSG00000251455.1 | ENSG00000252615.1 | ENSG00000307882.1 |
| ENSG00000236039.4 | ENSG00000234880.1 | ENSG00000205085.13 | ENSG00000268144.1 |
| ENSG00000288539.1 | ENSG00000288875.3 | ENSG00000187783.12 | ENSG00000278869.3 |
| ENSG00000256128.8 | ENSG00000268994.3 | ENSG00000286981.1 | ENSG00000268447.6 |
| ENSG00000250644.3 | ENSG00000237751.3 | ENSG00000206589.1 | ENSG00000249240.2 |
| ENSG00000143512.13 | ENSG00000307822.1 | ENSG00000264070.1 | ENSG00000175318.12 |
| ENSG00000273189.1 | ENSG00000285122.1 | ENSG00000138172.11 | ENSG00000231952.3 |
| ENSG00000137960.6 | ENSG00000245008.5 | ENSG00000223648.4 | ENSG00000200683.1 |
| ENSG00000310447.1 | ENSG00000295195.1 | ENSG00000286867.1 | ENSG00000169071.15 |
| ENSG00000279484.3 | ENSG00000306104.1 | ENSG00000224025.2 | ENSG00000303454.1 |
| ENSG00000288067.1 | ENSG00000128285.5 | ENSG00000293459.1 | ENSG00000308689.1 |
| ENSG00000186094.18 | ENSG00000235079.2 | ENSG00000295769.1 | ENSG00000149403.13 |
| ENSG00000270571.2 | ENSG00000258256.2 | ENSG00000299504.1 | ENSG00000261104.1 |
| ENSG00000248302.4 | ENSG00000227992.1 | ENSG00000231967.1 | ENSG00000286607.2 |

| | | | |
|---|---|---|---|
| ENSG00000108691.10 | ENSG00000144057.16 | ENSG00000289080.2 | ENSG00000173198.6 |
| ENSG00000203995.11 | ENSG00000274351.1 | ENSG00000249624.10 | ENSG00000120324.9 |
| ENSG00000300548.1 | ENSG00000229996.1 | ENSG00000272968.5 | ENSG00000276418.5 |
| ENSG00000302756.1 | ENSG00000260342.2 | ENSG00000223806.9 | ENSG00000263924.1 |
| ENSG00000296368.1 | ENSG00000299124.1 | ENSG00000263015.2 | ENSG00000305749.1 |
| ENSG00000273056.1 | ENSG00000225937.3 | ENSG00000294323.1 | ENSG00000280502.1 |
| ENSG00000234857.2 | ENSG00000188162.12 | ENSG00000230323.7 | ENSG00000207313.1 |
| ENSG00000187242.6 | ENSG00000264176.2 | ENSG00000306385.1 | ENSG00000297966.1 |
| ENSG00000158258.16 | ENSG00000129845.6 | ENSG00000260246.1 | ENSG00000213316.10 |
| ENSG00000248973.4 | ENSG00000294422.1 | ENSG00000254760.1 | ENSG00000304750.1 |
| ENSG00000259075.6 | ENSG00000302088.1 | ENSG00000186451.2 | ENSG00000302563.1 |
| ENSG00000064787.14 | ENSG00000301490.1 | ENSG00000252061.1 | ENSG00000144550.14 |
| ENSG00000239494.2 | ENSG00000261722.2 | ENSG00000199237.1 | ENSG00000306119.1 |
| ENSG00000164188.9 | ENSG00000208018.1 | ENSG00000296674.1 | ENSG00000301513.1 |
| ENSG00000306683.1 | ENSG00000222051.1 | ENSG00000223701.3 | ENSG00000227456.10 |
| ENSG00000264876.3 | ENSG00000250796.1 | ENSG00000139970.18 | ENSG00000278661.1 |
| ENSG00000204850.4 | ENSG00000223826.2 | ENSG00000266801.1 | ENSG00000297762.1 |
| ENSG00000196954.14 | ENSG00000176358.16 | ENSG00000164850.16 | ENSG00000148411.8 |
| ENSG00000276302.1 | ENSG00000007908.16 | ENSG00000286064.1 | ENSG00000266869.3 |
| ENSG00000306310.1 | ENSG00000278131.1 | ENSG00000125618.18 | ENSG00000279294.1 |
| ENSG00000200220.1 | ENSG00000236107.12 | ENSG00000293446.2 | ENSG00000226061.1 |
| ENSG00000262662.1 | ENSG00000163705.13 | ENSG00000287823.2 | ENSG00000309205.1 |
| ENSG00000288978.4 | ENSG00000252503.1 | ENSG00000260955.2 | ENSG00000233705.10 |
| ENSG00000233421.6 | ENSG00000270560.1 | ENSG00000306355.1 | ENSG00000202533.1 |
| ENSG00000265396.1 | ENSG00000294892.1 | ENSG00000270175.2 | ENSG00000303713.1 |
| ENSG00000303264.1 | ENSG00000227463.4 | ENSG00000303307.1 | ENSG00000115353.11 |
| ENSG00000300608.1 | ENSG00000295217.1 | ENSG00000288736.3 | ENSG00000091106.19 |
| ENSG00000299446.1 | ENSG00000305445.1 | ENSG00000299987.1 | ENSG00000306484.1 |
| ENSG00000259780.3 | ENSG00000196273.9 | ENSG00000231787.4 | ENSG00000235447.2 |
| ENSG00000220008.4 | ENSG00000259261.2 | ENSG00000187944.5 | ENSG00000181072.12 |
| ENSG00000286669.1 | ENSG00000301745.1 | ENSG00000170122.6 | ENSG00000283141.2 |
| ENSG00000125900.13 | ENSG00000303272.1 | ENSG00000169224.13 | ENSG00000283515.1 |
| ENSG00000241333.3 | ENSG00000249288.1 | ENSG00000273599.1 | ENSG00000276063.1 |
| ENSG00000149798.5 | ENSG00000256442.3 | ENSG00000211978.2 | ENSG00000118729.15 |
| ENSG00000226312.7 | ENSG00000162458.13 | ENSG00000183979.8 | ENSG00000282097.2 |
| ENSG00000227790.8 | ENSG00000053328.9 | ENSG00000277277.4 | ENSG00000305919.1 |

| | | | |
|---|---|---|---|
| ENSG00000294987.1 | ENSG00000212329.1 | ENSG00000277692.1 | ENSG00000229635.1 |
| ENSG00000268133.1 | ENSG00000279608.1 | ENSG00000279421.1 | ENSG00000287698.2 |
| ENSG00000254203.1 | ENSG00000289352.3 | ENSG00000231742.7 | ENSG00000295916.1 |
| ENSG00000305306.1 | ENSG00000100628.12 | ENSG00000248629.1 | ENSG00000185736.16 |
| ENSG00000002726.21 | ENSG00000269091.6 | ENSG00000113396.13 | ENSG00000275191.1 |
| ENSG00000136694.9 | ENSG00000308151.1 | ENSG00000293734.1 | ENSG00000289169.2 |
| ENSG00000176956.13 | ENSG00000259219.2 | ENSG00000271547.1 | ENSG00000235686.1 |
| ENSG00000305209.1 | ENSG00000207344.1 | ENSG00000274576.2 | ENSG00000186603.7 |
| ENSG00000299456.1 | ENSG00000308693.1 | ENSG00000295107.1 | ENSG00000296359.1 |
| ENSG00000224629.1 | ENSG00000303697.1 | ENSG00000277873.1 | ENSG00000299066.1 |
| ENSG00000295750.1 | ENSG00000001036.14 | ENSG00000111834.13 | ENSG00000165588.19 |
| ENSG00000297134.1 | ENSG00000287114.2 | ENSG00000197046.12 | ENSG00000310494.1 |
| ENSG00000301336.1 | ENSG00000198125.13 | ENSG00000272911.2 | ENSG00000286191.2 |
| ENSG00000290093.1 | ENSG00000294698.1 | ENSG00000245719.2 | ENSG00000248717.2 |
| ENSG00000162641.20 | ENSG00000198842.10 | ENSG00000106328.10 | ENSG00000248791.1 |
| ENSG00000182667.15 | ENSG00000293924.1 | ENSG00000238098.10 | ENSG00000298439.1 |
| ENSG00000166492.9 | ENSG00000261219.1 | ENSG00000229536.4 | ENSG00000270112.6 |
| ENSG00000274944.4 | ENSG00000270804.2 | ENSG00000288751.2 | ENSG00000220891.1 |
| ENSG00000174473.17 | ENSG00000299974.1 | ENSG00000269151.5 | ENSG00000236431.1 |
| ENSG00000224093.8 | ENSG00000232487.1 | ENSG00000101280.8 | ENSG00000301406.1 |
| ENSG00000309430.1 | ENSG00000296083.1 | ENSG00000300638.1 | ENSG00000250506.9 |
| ENSG00000302284.1 | ENSG00000296850.1 | ENSG00000226180.3 | ENSG00000228630.8 |
| ENSG00000285035.3 | ENSG00000308283.1 | ENSG00000283782.2 | ENSG00000101347.11 |
| ENSG00000120903.13 | ENSG00000174460.4 | ENSG00000183066.15 | ENSG00000108702.4 |
| ENSG00000270460.2 | ENSG00000263627.2 | ENSG00000271967.1 | ENSG00000309272.1 |
| ENSG00000220960.1 | ENSG00000132915.12 | ENSG00000293058.1 | ENSG00000237164.1 |
| ENSG00000225431.2 | ENSG00000196553.15 | ENSG00000266744.1 | ENSG00000307936.1 |
| ENSG00000267275.2 | ENSG00000251372.9 | ENSG00000163218.15 | ENSG00000290107.1 |
| ENSG00000305100.1 | ENSG00000267472.1 | ENSG00000250899.3 | ENSG00000299096.1 |
| ENSG00000205978.6 | ENSG00000287853.1 | ENSG00000269886.1 | ENSG00000176083.18 |
| ENSG00000270449.1 | ENSG00000200156.2 | ENSG00000074803.21 | ENSG00000277022.1 |
| ENSG00000278962.1 | ENSG00000134817.12 | ENSG00000081148.12 | ENSG00000249252.6 |
| ENSG00000223462.3 | ENSG00000290642.1 | ENSG00000309664.1 | ENSG00000310160.1 |
| ENSG00000080947.16 | ENSG00000253167.1 | ENSG00000288051.2 | ENSG00000164078.15 |
| ENSG00000250015.1 | ENSG00000173699.17 | ENSG00000279177.1 | ENSG00000258082.2 |
| ENSG00000259399.1 | ENSG00000181785.3 | ENSG00000243649.10 | ENSG00000238122.2 |

| | | | |
|---|---|---|---|
| ENSG00000257515.1 | ENSG00000302830.1 | ENSG00000301157.1 | ENSG00000300503.1 |
| ENSG00000287236.2 | ENSG00000274685.2 | ENSG00000183690.13 | ENSG00000303147.1 |
| ENSG00000172159.16 | ENSG00000294824.1 | ENSG00000179855.9 | ENSG00000301019.1 |
| ENSG00000206640.1 | ENSG00000135773.13 | ENSG00000175505.11 | ENSG00000309800.1 |
| ENSG00000178297.15 | ENSG00000302963.1 | ENSG00000172938.3 | ENSG00000297040.1 |
| ENSG00000085871.9 | ENSG00000175564.13 | ENSG00000169064.13 | ENSG00000302724.1 |
| ENSG00000203258.3 | ENSG00000304187.1 | ENSG00000141748.13 | ENSG00000271288.1 |
| ENSG00000121380.12 | ENSG00000310577.1 | ENSG00000272799.1 | ENSG00000282572.5 |
| ENSG00000298281.1 | ENSG00000303802.1 | ENSG00000139574.9 | ENSG00000204851.7 |
| ENSG00000184635.17 | ENSG00000303755.1 | ENSG00000267758.1 | ENSG00000297250.1 |
| ENSG00000300168.1 | ENSG00000296370.1 | ENSG00000271367.1 | ENSG00000230790.2 |
| ENSG00000310059.1 | ENSG00000064489.23 | ENSG00000253910.3 | ENSG00000132622.11 |
| ENSG00000092969.12 | ENSG00000165731.22 | ENSG00000278013.1 | ENSG00000180549.8 |
| ENSG00000303883.1 | ENSG00000261465.8 | ENSG00000268790.5 | ENSG00000228300.15 |
| ENSG00000308776.1 | ENSG00000277587.1 | ENSG00000295501.1 | ENSG00000047617.18 |
| ENSG00000204779.2 | ENSG00000299324.1 | ENSG00000214189.10 | ENSG00000290423.2 |
| ENSG00000298240.1 | ENSG00000245975.3 | ENSG00000177679.16 | ENSG00000289928.2 |
| ENSG00000299430.1 | ENSG00000125810.11 | ENSG00000234423.3 | ENSG00000215812.5 |
| ENSG00000272542.1 | ENSG00000257596.2 | ENSG00000136535.15 | ENSG00000278455.1 |
| ENSG00000279074.1 | ENSG00000286732.2 | ENSG00000248758.1 | ENSG00000117834.13 |
| ENSG00000300726.1 | ENSG00000303469.1 | ENSG00000307023.1 | ENSG00000257842.7 |
| ENSG00000308122.1 | ENSG00000207973.1 | ENSG00000306196.1 | ENSG00000307009.1 |
| ENSG00000305089.1 | ENSG00000302029.1 | ENSG00000201830.1 | ENSG00000269026.2 |
| ENSG00000038427.16 | ENSG00000011590.14 | ENSG00000184985.17 | ENSG00000270823.1 |
| ENSG00000169583.13 | ENSG00000084110.11 | ENSG00000280163.1 | ENSG00000211747.3 |
| ENSG00000303745.1 | ENSG00000226516.8 | ENSG00000273387.1 | ENSG00000296342.1 |
| ENSG00000188389.12 | ENSG00000167759.13 | ENSG00000270550.1 | ENSG00000277498.1 |
| ENSG00000100385.15 | ENSG00000171903.17 | ENSG00000288602.1 | ENSG00000153446.16 |
| ENSG00000267800.1 | ENSG00000303292.1 | ENSG00000166143.10 | ENSG00000212855.6 |
| ENSG00000279908.1 | ENSG00000233579.1 | ENSG00000288843.3 | ENSG00000250413.2 |
| ENSG00000236953.1 | ENSG00000297045.1 | ENSG00000157303.11 | ENSG00000309683.1 |
| ENSG00000299018.1 | ENSG00000021488.13 | ENSG00000309684.1 | ENSG00000181781.10 |
| ENSG00000101017.15 | ENSG00000214102.7 | ENSG00000179044.16 | ENSG00000303291.1 |
| ENSG00000303650.1 | ENSG00000235779.9 | ENSG00000232973.15 | ENSG00000100985.7 |
| ENSG00000130768.15 | ENSG00000297571.1 | ENSG00000215838.5 | ENSG00000186810.8 |
| ENSG00000287076.2 | ENSG00000248740.9 | ENSG00000180708.6 | ENSG00000164304.16 |

| | | | |
|---|---|---|---|
| ENSG00000253651.1 | ENSG00000289755.2 | ENSG00000307021.1 | ENSG00000287032.2 |
| ENSG00000307201.1 | ENSG00000166359.11 | ENSG00000297815.1 | ENSG00000270839.1 |
| ENSG00000287035.2 | ENSG00000294454.1 | ENSG00000278505.5 | ENSG00000290044.2 |
| ENSG00000303186.1 | ENSG00000180644.8 | ENSG00000286188.1 | ENSG00000295235.1 |
| ENSG00000290075.2 | ENSG00000176029.14 | ENSG00000171217.5 | ENSG00000307059.1 |
| ENSG00000269416.8 | ENSG00000276272.1 | ENSG00000124780.14 | ENSG00000112378.12 |
| ENSG00000215595.3 | ENSG00000289488.1 | ENSG00000297460.1 | ENSG00000279951.1 |
| ENSG00000295746.1 | ENSG00000235848.4 | ENSG00000240211.2 | ENSG00000223387.7 |
| ENSG00000298617.1 | ENSG00000254978.2 | ENSG00000266189.1 | ENSG00000272941.1 |
| ENSG00000291062.2 | ENSG00000204475.10 | ENSG00000248866.2 | ENSG00000168282.6 |
| ENSG00000303695.1 | ENSG00000113525.10 | ENSG00000255036.6 | ENSG00000281358.2 |
| ENSG00000297119.1 | ENSG00000236317.1 | ENSG00000308416.1 | ENSG00000181885.18 |
| ENSG00000255351.1 | ENSG00000157765.13 | ENSG00000137473.19 | ENSG00000289412.3 |
| ENSG00000256596.2 | ENSG00000254289.2 | ENSG00000297157.1 | ENSG00000271886.1 |
| ENSG00000302210.1 | ENSG00000213014.3 | ENSG00000283317.1 | ENSG00000248356.1 |
| ENSG00000239335.4 | ENSG00000102935.12 | ENSG00000239519.1 | ENSG00000231702.2 |
| ENSG00000288637.1 | ENSG00000254592.1 | ENSG00000286268.1 | ENSG00000231585.2 |
| ENSG00000288534.1 | ENSG00000078900.16 | ENSG00000164746.14 | ENSG00000164403.15 |
| ENSG00000228133.3 | ENSG00000294376.1 | ENSG00000112619.8 | ENSG00000298570.1 |
| ENSG00000255639.3 | ENSG00000188763.5 | ENSG00000167858.13 | ENSG00000258400.1 |
| ENSG00000256852.1 | ENSG00000271986.2 | ENSG00000296538.1 | ENSG00000267353.1 |
| ENSG00000308658.1 | ENSG00000188517.17 | ENSG00000240403.6 | ENSG00000258707.2 |
| ENSG00000271155.1 | ENSG00000289127.2 | ENSG00000249650.2 | ENSG00000181284.3 |
| ENSG00000175868.14 | ENSG00000297935.1 | ENSG00000211955.2 | ENSG00000243227.3 |
| ENSG00000307343.1 | ENSG00000196866.3 | ENSG00000299043.1 | ENSG00000227776.1 |
| ENSG00000227237.2 | ENSG00000293239.2 | ENSG00000260452.1 | ENSG00000274769.1 |
| ENSG00000225886.3 | ENSG00000228915.3 | ENSG00000258694.2 | ENSG00000264860.1 |
| ENSG00000303600.1 | ENSG00000302488.1 | ENSG00000229368.1 | ENSG00000271228.1 |
| ENSG00000211947.2 | ENSG00000279925.1 | ENSG00000296886.1 | ENSG00000307533.1 |
| ENSG00000200815.1 | ENSG00000259499.1 | ENSG00000278665.1 | ENSG00000231840.3 |
| ENSG00000306441.1 | ENSG00000305251.1 | ENSG00000251461.4 | ENSG00000164049.14 |
| ENSG00000229162.1 | ENSG00000223519.9 | ENSG00000162711.19 | ENSG00000287339.1 |
| ENSG00000294465.1 | ENSG00000144668.12 | ENSG00000286001.1 | ENSG00000293405.2 |
| ENSG00000163833.8 | ENSG00000250410.1 | ENSG00000289140.2 | ENSG00000243319.7 |
| ENSG00000171931.13 | ENSG00000306686.1 | ENSG00000050628.21 | ENSG00000196420.8 |
| ENSG00000290034.2 | ENSG00000233124.3 | ENSG00000310289.1 | ENSG00000288766.2 |

| | | | |
|---|---|---|---|
| ENSG00000253440.1 | ENSG00000260306.1 | ENSG00000239577.3 | ENSG00000283122.1 |
| ENSG00000188199.11 | ENSG00000286905.1 | ENSG00000162542.14 | ENSG00000226660.2 |
| ENSG00000265739.1 | ENSG00000273816.1 | ENSG00000287502.1 | ENSG00000268231.1 |
| ENSG00000286703.2 | ENSG00000168802.14 | ENSG00000214870.12 | ENSG00000182795.13 |
| ENSG00000185523.6 | ENSG00000304724.1 | ENSG00000298067.1 | ENSG00000272425.1 |
| ENSG00000288551.2 | ENSG00000235358.3 | ENSG00000165905.18 | ENSG00000253305.3 |
| ENSG00000252641.2 | ENSG00000291017.2 | ENSG00000289981.2 | ENSG00000138835.24 |
| ENSG00000276775.1 | ENSG00000241546.1 | ENSG00000081985.14 | ENSG00000275793.1 |
| ENSG00000295993.1 | ENSG00000297156.1 | ENSG00000279558.2 | ENSG00000282418.2 |
| ENSG00000224950.2 | ENSG00000138311.18 | ENSG00000186334.10 | ENSG00000225556.2 |
| ENSG00000255330.9 | ENSG00000123130.17 | ENSG00000253522.7 | ENSG00000286940.2 |
| ENSG00000257264.5 | ENSG00000237641.1 | ENSG00000305446.1 | ENSG00000232479.2 |
| ENSG00000003436.16 | ENSG00000182489.9 | ENSG00000273821.1 | ENSG00000240051.1 |
| ENSG00000242352.2 | ENSG00000228748.3 | ENSG00000204179.11 | ENSG00000106538.10 |
| ENSG00000144642.22 | ENSG00000227630.5 | ENSG00000303043.1 | ENSG00000294756.1 |
| ENSG00000280348.1 | ENSG00000296942.1 | ENSG00000142347.21 | ENSG00000260810.1 |
| ENSG00000108950.12 | ENSG00000301671.1 | ENSG00000197279.5 | ENSG00000019505.8 |
| ENSG00000186471.13 | ENSG00000189120.5 | ENSG00000286920.2 | ENSG00000309885.1 |
| ENSG00000038295.8 | ENSG00000295412.1 | ENSG00000089692.9 | ENSG00000296162.1 |
| ENSG00000157601.15 | ENSG00000243480.8 | ENSG00000211956.2 | ENSG00000269559.2 |
| ENSG00000233079.4 | ENSG00000159403.18 | ENSG00000260063.2 | ENSG00000116157.6 |
| ENSG00000223935.4 | ENSG00000295507.1 | ENSG00000303629.1 | ENSG00000302384.1 |
| ENSG00000232457.4 | ENSG00000226450.2 | ENSG00000253196.4 | ENSG00000300349.1 |
| ENSG00000274995.1 | ENSG00000214369.2 | ENSG00000102265.12 | ENSG00000301402.1 |
| ENSG00000236723.2 | ENSG00000232422.1 | ENSG00000296999.1 | ENSG00000152475.7 |
| ENSG00000305432.1 | ENSG00000294464.1 | ENSG00000287090.2 | ENSG00000270084.2 |
| ENSG00000305856.1 | ENSG00000289553.2 | ENSG00000250685.7 | ENSG00000164185.7 |
| ENSG00000260253.2 | ENSG00000307354.1 | ENSG00000303516.1 | ENSG00000255339.6 |
| ENSG00000299529.1 | ENSG00000305547.1 | ENSG00000230747.1 | ENSG00000144452.15 |
| ENSG00000272674.4 | ENSG00000284194.3 | ENSG00000298401.1 | ENSG00000100427.16 |
| ENSG00000274750.3 | ENSG00000196569.14 | ENSG00000231050.4 | ENSG00000139044.12 |
| ENSG00000242798.2 | ENSG00000258915.1 | ENSG00000236040.1 | ENSG00000159784.18 |
| ENSG00000108798.9 | ENSG00000267291.2 | ENSG00000233131.1 | ENSG00000296270.1 |
| ENSG00000211942.3 | ENSG00000226862.1 | ENSG00000188383.8 | ENSG00000230013.3 |
| ENSG00000216906.2 | ENSG00000271717.1 | ENSG00000294235.1 | ENSG00000128268.12 |
| ENSG00000169575.5 | ENSG00000253598.3 | ENSG00000235072.3 | ENSG00000214562.15 |

| | | | |
|---|---|---|---|
| ENSG00000104524.14 | ENSG00000255079.2 | ENSG00000266126.1 | ENSG00000294389.1 |
| ENSG00000289990.2 | ENSG00000115252.19 | ENSG00000290054.2 | ENSG00000258867.9 |
| ENSG00000294377.1 | ENSG00000300583.1 | ENSG00000175899.15 | ENSG00000306976.1 |
| ENSG00000308410.1 | ENSG00000297301.1 | ENSG00000156206.14 | ENSG00000172380.6 |
| ENSG00000136002.21 | ENSG00000305390.1 | ENSG00000229983.1 | ENSG00000144230.17 |
| ENSG00000140682.19 | ENSG00000168065.16 | ENSG00000228397.4 | ENSG00000289716.1 |
| ENSG00000259598.1 | ENSG00000162039.19 | ENSG00000021355.13 | ENSG00000280623.2 |
| ENSG00000293063.1 | ENSG00000100228.14 | ENSG00000275183.2 | ENSG00000197057.10 |
| ENSG00000234969.1 | ENSG00000096264.14 | ENSG00000172426.16 | ENSG00000180042.5 |
| ENSG00000136197.14 | ENSG00000267595.1 | ENSG00000111490.15 | ENSG00000290714.2 |
| ENSG00000223731.4 | ENSG00000279452.1 | ENSG00000198133.9 | ENSG00000302489.1 |
| ENSG00000300846.1 | ENSG00000248559.1 | ENSG00000070526.15 | ENSG00000124313.19 |
| ENSG00000171388.12 | ENSG00000142973.16 | ENSG00000254061.1 | ENSG00000251442.9 |
| ENSG00000294333.1 | ENSG00000289499.2 | ENSG00000275778.4 | ENSG00000186594.17 |
| ENSG00000287055.1 | ENSG00000158865.13 | ENSG00000253759.1 | ENSG00000125533.6 |
| ENSG00000251396.10 | ENSG00000276651.1 | ENSG00000298184.1 | ENSG00000302010.1 |
| ENSG00000305995.1 | ENSG00000256263.1 | ENSG00000257159.1 | ENSG00000304628.1 |
| ENSG00000167194.8 | ENSG00000241316.9 | ENSG00000154556.20 | ENSG00000242888.1 |
| ENSG00000295678.1 | ENSG00000092068.21 | ENSG00000183246.8 | ENSG00000310441.1 |
| ENSG00000300728.1 | ENSG00000308631.1 | ENSG00000269940.1 | ENSG00000175202.6 |
| ENSG00000244302.3 | ENSG00000259529.2 | ENSG00000302593.1 | ENSG00000302876.1 |
| ENSG00000243749.2 | ENSG00000281832.2 | ENSG00000260439.1 | ENSG00000229921.8 |
| ENSG00000299254.1 | ENSG00000287262.2 | ENSG00000260742.1 | ENSG00000225075.2 |
| ENSG00000268750.7 | ENSG00000271554.1 | ENSG00000287918.3 | ENSG00000196878.15 |
| ENSG00000138771.16 | ENSG00000236449.3 | ENSG00000270194.2 | ENSG00000289052.2 |
| ENSG00000270505.1 | ENSG00000235584.4 | ENSG00000294996.1 | ENSG00000204532.9 |
| ENSG00000201182.1 | ENSG00000196932.12 | ENSG00000295846.1 | ENSG00000119686.10 |
| ENSG00000295897.1 | ENSG00000284948.1 | ENSG00000308311.1 | ENSG00000245025.4 |
| ENSG00000236987.2 | ENSG00000228817.4 | ENSG00000255314.5 | ENSG00000290803.2 |
| ENSG00000293569.1 | ENSG00000301915.1 | ENSG00000304960.1 | ENSG00000307028.1 |
| ENSG00000149489.9 | ENSG00000288600.2 | ENSG00000309627.1 | ENSG00000068001.14 |
| ENSG00000293448.2 | ENSG00000304493.1 | ENSG00000203758.4 | ENSG00000037897.17 |
| ENSG00000272223.1 | ENSG00000165973.19 | ENSG00000256043.5 | ENSG00000233025.1 |
| ENSG00000272646.1 | ENSG00000224650.2 | ENSG00000211935.3 | ENSG00000293260.1 |
| ENSG00000272057.1 | ENSG00000258101.3 | ENSG00000228081.2 | ENSG00000141837.23 |
| ENSG00000286271.4 | ENSG00000287287.2 | ENSG00000173285.5 | ENSG00000213977.8 |

| | | | |
|---|---|---|---|
| ENSG00000278607.4 | ENSG00000011052.21 | ENSG00000302043.1 | ENSG00000202415.1 |
| ENSG00000159792.10 | ENSG00000235119.1 | ENSG00000173638.19 | ENSG00000283994.1 |
| ENSG00000188993.4 | ENSG00000010379.16 | ENSG00000174807.4 | ENSG00000273551.1 |
| ENSG00000303568.1 | ENSG00000165168.9 | ENSG00000287935.2 | ENSG00000243433.1 |
| ENSG00000108405.4 | ENSG00000297686.1 | ENSG00000297455.1 | ENSG00000288844.2 |
| ENSG00000284677.1 | ENSG00000288033.1 | ENSG00000081692.13 | ENSG00000307566.1 |
| ENSG00000295221.1 | ENSG00000135842.17 | ENSG00000261519.3 | ENSG00000105967.16 |
| ENSG00000186281.13 | ENSG00000309390.1 | ENSG00000302371.1 | ENSG00000291071.2 |
| ENSG00000137672.13 | ENSG00000171051.10 | ENSG00000122223.13 | ENSG00000101306.11 |
| ENSG00000182057.6 | ENSG00000173548.9 | ENSG00000138356.14 | ENSG00000251226.3 |
| ENSG00000145936.9 | ENSG00000253240.1 | ENSG00000294763.1 | ENSG00000164933.13 |
| ENSG00000183230.18 | ENSG00000160401.15 | ENSG00000183117.20 | ENSG00000205209.8 |
| ENSG00000302324.1 | ENSG00000135525.19 | ENSG00000298587.1 | ENSG00000171121.17 |
| ENSG00000288684.1 | ENSG00000152784.16 | ENSG00000280327.1 | ENSG00000105122.13 |
| ENSG00000238961.1 | ENSG00000289057.1 | ENSG00000134508.13 | ENSG00000185883.12 |
| ENSG00000177839.7 | ENSG00000268375.1 | ENSG00000260646.1 | ENSG00000176125.7 |
| ENSG00000285677.3 | ENSG00000254777.6 | ENSG00000287654.1 | ENSG00000283948.1 |
| ENSG00000274275.1 | ENSG00000172955.17 | ENSG00000234769.7 | ENSG00000072163.20 |
| ENSG00000188032.10 | ENSG00000189419.9 | ENSG00000276255.2 | ENSG00000172179.13 |
| ENSG00000212747.5 | ENSG00000309702.1 | ENSG00000169026.13 | ENSG00000160307.10 |
| ENSG00000227868.7 | ENSG00000285932.1 | ENSG00000269924.1 | ENSG00000227356.3 |
| ENSG00000286579.1 | ENSG00000138823.14 | ENSG00000052749.15 | ENSG00000302468.1 |
| ENSG00000164929.17 | ENSG00000236398.2 | ENSG00000275549.1 | ENSG00000175206.11 |
| ENSG00000262185.4 | ENSG00000269910.1 | ENSG00000285171.1 | ENSG00000307676.1 |
| ENSG00000180015.12 | ENSG00000287894.2 | ENSG00000290994.2 | ENSG00000287420.2 |
| ENSG00000249978.1 | ENSG00000224837.1 | ENSG00000296702.1 | ENSG00000166341.9 |
| ENSG00000117560.8 | ENSG00000204934.11 | ENSG00000135318.12 | ENSG00000305962.1 |
| ENSG00000130720.13 | ENSG00000159961.3 | ENSG00000112541.19 | ENSG00000026508.21 |
| ENSG00000187952.9 | ENSG00000267666.3 | ENSG00000187987.10 | ENSG00000242689.3 |
| ENSG00000301622.1 | ENSG00000254419.1 | ENSG00000173227.14 | ENSG00000199509.1 |
| ENSG00000206739.1 | ENSG00000294647.1 | ENSG00000168952.16 | ENSG00000213171.3 |
| ENSG00000293107.1 | ENSG00000283288.2 | ENSG00000283562.1 | ENSG00000149506.12 |
| ENSG00000176108.9 | ENSG00000189002.10 | ENSG00000224256.1 | ENSG00000171502.15 |
| ENSG00000255561.7 | ENSG00000268107.6 | ENSG00000301761.1 | ENSG00000299483.1 |
| ENSG00000239705.3 | ENSG00000163554.15 | ENSG00000127588.5 | ENSG00000272002.1 |
| ENSG00000168453.16 | ENSG00000131097.7 | ENSG00000154975.14 | ENSG00000293628.1 |

| | | | |
|---|---|---|---|
| ENSG00000303980.1 | ENSG00000259218.6 | ENSG00000297343.1 | ENSG00000172748.14 |
| ENSG00000147082.18 | ENSG00000211974.3 | ENSG00000260325.2 | ENSG00000178921.14 |
| ENSG00000008853.18 | ENSG00000266017.1 | ENSG00000289446.2 | ENSG00000304830.1 |
| ENSG00000233483.5 | ENSG00000143869.7 | ENSG00000291077.2 | ENSG00000224988.1 |
| ENSG00000132382.15 | ENSG00000261373.1 | ENSG00000287863.1 | ENSG00000100055.21 |
| ENSG00000241679.3 | ENSG00000267288.3 | ENSG00000260238.6 | ENSG00000130711.5 |
| ENSG00000296209.1 | ENSG00000229950.2 | ENSG00000229589.1 | ENSG00000295178.1 |
| ENSG00000305208.1 | ENSG00000276116.3 | ENSG00000211765.1 | ENSG00000166401.15 |
| ENSG00000110104.13 | ENSG00000117115.13 | ENSG00000287672.2 | ENSG00000121807.7 |
| ENSG00000106991.15 | ENSG00000169896.18 | ENSG00000151079.8 | ENSG00000117322.19 |
| ENSG00000206762.1 | ENSG00000110665.12 | ENSG00000233246.2 | ENSG00000275367.1 |
| ENSG00000272148.1 | ENSG00000271646.2 | ENSG00000304746.1 | ENSG00000110931.19 |
| ENSG00000234476.6 | ENSG00000058866.15 | ENSG00000154330.13 | ENSG00000275454.1 |
| ENSG00000309595.1 | ENSG00000294840.1 | ENSG00000269892.1 | ENSG00000278619.5 |
| ENSG00000299601.1 | ENSG00000152689.19 | ENSG00000160791.14 | ENSG00000296598.1 |
| ENSG00000165985.12 | ENSG00000100413.17 | ENSG00000287362.2 | ENSG00000204929.14 |
| ENSG00000230613.2 | ENSG00000235837.2 | ENSG00000309662.1 | ENSG00000272914.1 |
| ENSG00000266962.4 | ENSG00000167549.19 | ENSG00000306543.1 | ENSG00000277142.1 |
| ENSG00000297558.1 | ENSG00000260979.2 | ENSG00000285382.1 | ENSG00000308804.1 |
| ENSG00000163485.18 | ENSG00000124160.12 | ENSG00000167769.5 | ENSG00000240990.10 |
| ENSG00000260219.5 | ENSG00000248590.2 | ENSG00000167676.5 | ENSG00000256646.8 |
| ENSG00000064763.12 | ENSG00000228672.4 | ENSG00000175746.8 | ENSG00000231858.5 |
| ENSG00000274600.2 | ENSG00000107831.14 | ENSG00000255428.1 | ENSG00000134539.17 |
| ENSG00000290374.2 | ENSG00000289003.1 | ENSG00000214706.12 | ENSG00000270248.1 |
| ENSG00000184178.17 | ENSG00000294575.1 | ENSG00000291186.2 | ENSG00000297970.1 |
| ENSG00000256663.1 | ENSG00000253133.2 | ENSG00000100304.13 | ENSG00000304925.1 |
| ENSG00000163519.14 | ENSG00000137843.12 | ENSG00000230105.2 | ENSG00000196872.12 |
| ENSG00000232837.1 | ENSG00000226938.2 | ENSG00000306456.1 | ENSG00000285644.2 |
| ENSG00000299358.1 | ENSG00000307322.1 | ENSG00000289723.1 | ENSG00000289235.2 |
| ENSG00000128617.3 | ENSG00000273419.1 | ENSG00000166126.11 | ENSG00000240143.1 |
| ENSG00000244155.1 | ENSG00000188051.7 | ENSG00000270277.1 | ENSG00000230778.2 |
| ENSG00000282034.2 | ENSG00000188175.10 | ENSG00000277245.1 | ENSG00000265790.1 |
| ENSG00000177096.9 | ENSG00000295329.1 | ENSG00000277270.3 | ENSG00000228570.8 |
| ENSG00000273669.1 | ENSG00000291163.1 | ENSG00000282904.3 | ENSG00000107796.14 |
| ENSG00000298167.1 | ENSG00000166189.8 | ENSG00000280187.2 | ENSG00000293992.1 |
| ENSG00000271806.1 | ENSG00000162390.18 | ENSG00000261587.3 | ENSG00000283304.2 |

| | | | |
|---|---|---|---|
| ENSG00000213246.7 | ENSG00000125089.18 | ENSG00000290957.2 | ENSG00000122122.10 |
| ENSG00000286691.2 | ENSG00000299798.1 | ENSG00000105784.16 | ENSG00000298716.1 |
| ENSG00000235173.7 | ENSG00000179869.15 | ENSG00000299982.1 | ENSG00000229324.1 |
| ENSG00000304574.1 | ENSG00000263843.1 | ENSG00000278922.1 | ENSG00000298316.1 |
| ENSG00000144681.11 | ENSG00000285521.2 | ENSG00000308336.1 | ENSG00000300626.1 |
| ENSG00000226954.1 | ENSG00000289570.2 | ENSG00000310416.1 | ENSG00000293515.2 |
| ENSG00000295772.1 | ENSG00000274292.3 | ENSG00000295019.1 | ENSG00000269421.1 |
| ENSG00000268480.4 | ENSG00000295519.1 | ENSG00000183250.14 | ENSG00000298504.1 |
| ENSG00000283635.1 | ENSG00000122783.17 | ENSG00000257813.2 | ENSG00000167969.13 |
| ENSG00000268889.3 | ENSG00000223513.1 | ENSG00000174448.9 | ENSG00000296385.1 |
| ENSG00000134575.13 | ENSG00000303001.1 | ENSG00000292345.1 | ENSG00000121281.13 |
| ENSG00000269054.3 | ENSG00000277449.2 | ENSG00000263528.8 | ENSG00000307661.1 |
| ENSG00000204110.8 | ENSG00000294279.1 | ENSG00000242540.3 | ENSG00000100351.17 |
| ENSG00000262921.1 | ENSG00000308318.1 | ENSG00000013016.16 | ENSG00000235888.6 |
| ENSG00000260876.7 | ENSG00000007312.14 | ENSG00000308586.1 | ENSG00000258798.2 |
| ENSG00000042832.12 | ENSG00000250474.2 | ENSG00000137275.16 | ENSG00000260388.2 |
| ENSG00000259356.1 | ENSG00000105953.16 | ENSG00000269951.1 | ENSG00000285712.2 |
| ENSG00000258728.2 | ENSG00000281005.2 | ENSG00000289398.2 | ENSG00000125968.10 |
| ENSG00000284626.1 | ENSG00000198121.15 | ENSG00000172260.15 | ENSG00000143167.12 |
| ENSG00000105676.14 | ENSG00000300869.1 | ENSG00000063176.16 | ENSG00000110876.11 |
| ENSG00000223547.10 | ENSG00000161653.11 | ENSG00000303877.1 | ENSG00000187688.15 |
| ENSG00000214248.3 | ENSG00000309737.1 | ENSG00000257940.2 | ENSG00000279348.1 |
| ENSG00000253720.2 | ENSG00000276633.1 | ENSG00000295204.1 | ENSG00000272282.2 |
| ENSG00000211766.1 | ENSG00000305148.2 | ENSG00000162461.8 | ENSG00000106305.10 |
| ENSG00000279923.1 | ENSG00000106948.17 | ENSG00000235413.3 | ENSG00000262319.1 |
| ENSG00000103742.12 | ENSG00000260855.1 | ENSG00000300491.1 | ENSG00000304651.1 |
| ENSG00000183751.16 | ENSG00000269749.1 | ENSG00000115129.14 | ENSG00000278175.7 |
| ENSG00000275591.5 | ENSG00000224609.9 | ENSG00000196388.9 | ENSG00000007376.8 |
| ENSG00000258644.6 | ENSG00000303567.1 | ENSG00000270587.1 | ENSG00000172936.18 |
| ENSG00000288045.1 | ENSG00000305020.1 | ENSG00000249328.4 | ENSG00000213089.4 |
| ENSG00000285142.1 | ENSG00000137225.13 | ENSG00000163017.14 | ENSG00000168806.8 |
| ENSG00000241627.3 | ENSG00000287254.2 | ENSG00000004864.14 | ENSG00000152207.8 |
| ENSG00000118894.15 | ENSG00000302958.1 | ENSG00000168004.10 | ENSG00000257495.6 |
| ENSG00000167984.18 | ENSG00000166166.13 | ENSG00000268006.2 | ENSG00000157911.11 |
| ENSG00000296783.2 | ENSG00000132881.12 | ENSG00000179397.18 | ENSG00000272405.1 |
| ENSG00000167600.14 | ENSG00000270959.3 | ENSG00000137486.17 | ENSG00000260643.2 |

| | | | |
|---|---|---|---|
| ENSG00000104043.15 | ENSG00000293210.2 | ENSG00000261147.1 | ENSG00000146834.16 |
| ENSG00000150455.15 | ENSG00000123416.15 | ENSG00000102349.18 | ENSG00000184574.10 |
| ENSG00000168461.13 | ENSG00000307919.1 | ENSG00000273812.3 | ENSG00000197632.9 |
| ENSG00000299332.1 | ENSG00000226777.10 | ENSG00000293475.2 | ENSG00000170364.13 |
| ENSG00000255381.1 | ENSG00000115705.22 | ENSG00000236297.2 | ENSG00000113273.17 |
| ENSG00000185261.16 | ENSG00000309428.1 | ENSG00000229870.1 | ENSG00000227942.1 |
| ENSG00000259719.7 | ENSG00000214960.11 | ENSG00000141994.16 | ENSG00000180190.13 |
| ENSG00000302983.1 | ENSG00000168961.17 | ENSG00000074370.19 | ENSG00000211968.3 |
| ENSG00000165409.18 | ENSG00000174004.6 | ENSG00000101986.12 | ENSG00000107815.10 |
| ENSG00000189420.9 | ENSG00000231991.4 | ENSG00000293476.1 | ENSG00000150990.9 |
| ENSG00000089012.14 | ENSG00000158473.8 | ENSG00000109667.12 | ENSG00000273154.3 |
| ENSG00000230682.1 | ENSG00000143851.17 | ENSG00000240449.2 | ENSG00000287885.2 |
| ENSG00000167131.17 | ENSG00000148468.17 | ENSG00000245688.1 | ENSG00000229291.3 |
| ENSG00000136514.3 | ENSG00000293412.2 | ENSG00000153404.15 | ENSG00000169314.15 |
| ENSG00000238207.3 | ENSG00000280231.1 | ENSG00000162494.6 | ENSG00000297451.1 |
| ENSG00000171777.17 | ENSG00000271664.1 | ENSG00000175093.5 | ENSG00000205089.8 |
| ENSG00000186517.14 | ENSG00000299781.1 | ENSG00000273540.5 | ENSG00000125454.12 |
| ENSG00000286912.1 | ENSG00000305150.1 | ENSG00000182162.11 | ENSG00000161618.10 |
| ENSG00000226508.2 | ENSG00000296862.1 | ENSG00000124334.19 | ENSG00000280537.2 |
| ENSG00000144712.13 | ENSG00000225230.1 | ENSG00000125630.16 | ENSG00000222046.3 |
| ENSG00000130943.7 | ENSG00000233627.2 | ENSG00000297869.1 | ENSG00000092929.13 |
| ENSG00000310152.1 | ENSG00000188512.6 | ENSG00000290913.2 | ENSG00000139192.12 |
| ENSG00000303578.1 | ENSG00000260172.3 | ENSG00000233337.1 | ENSG00000226913.3 |
| ENSG00000033100.17 | ENSG00000302071.1 | ENSG00000296560.1 | ENSG00000078589.13 |
| ENSG00000104356.11 | ENSG00000169946.14 | ENSG00000211970.3 | ENSG00000169715.15 |
| ENSG00000163568.17 | ENSG00000309307.1 | ENSG00000177628.16 | ENSG00000288869.3 |
| ENSG00000040633.13 | ENSG00000251229.1 | ENSG00000172543.9 | ENSG00000239620.3 |
| ENSG00000179409.11 | ENSG00000118898.16 | ENSG00000136010.14 | ENSG00000254215.1 |
| ENSG00000228157.5 | ENSG00000253965.4 | ENSG00000226942.2 | ENSG00000297688.1 |
| ENSG00000102967.12 | ENSG00000278948.1 | ENSG00000204519.11 | ENSG00000096996.16 |
| ENSG00000279667.1 | ENSG00000285602.1 | ENSG00000257488.5 | ENSG00000255299.6 |
| ENSG00000295537.1 | ENSG00000225406.1 | ENSG00000279296.1 | ENSG00000185085.2 |
| ENSG00000176318.8 | ENSG00000204947.9 | ENSG00000144120.13 | ENSG00000175040.6 |
| ENSG00000246095.3 | ENSG00000235214.2 | ENSG00000174343.6 | ENSG00000285830.1 |
| ENSG00000184811.4 | ENSG00000298889.1 | ENSG00000250778.1 | ENSG00000299278.1 |
| ENSG00000160959.8 | ENSG00000288804.2 | ENSG00000161395.14 | ENSG00000287925.2 |

| | | | |
|---|---|---|---|
| ENSG00000266524.3 | ENSG00000303333.1 | ENSG00000018869.18 | ENSG00000270062.1 |
| ENSG00000224418.2 | ENSG00000302681.1 | ENSG00000295869.1 | ENSG00000247774.10 |
| ENSG00000101265.16 | ENSG00000121764.12 | ENSG00000283010.2 | ENSG00000304828.1 |
| ENSG00000115998.8 | ENSG00000305299.1 | ENSG00000198736.13 | ENSG00000229278.5 |
| ENSG00000214717.12 | ENSG00000183134.5 | ENSG00000181004.10 | ENSG00000167394.13 |
| ENSG00000240286.1 | ENSG00000175283.8 | ENSG00000154760.14 | ENSG00000185347.19 |
| ENSG00000233081.2 | ENSG00000160703.17 | ENSG00000116096.7 | ENSG00000173599.15 |
| ENSG00000163449.11 | ENSG00000289257.3 | ENSG00000161267.12 | ENSG00000006744.19 |
| ENSG00000301095.1 | ENSG00000272602.6 | ENSG00000272711.1 | ENSG00000198440.9 |
| ENSG00000303252.1 | ENSG00000102245.9 | ENSG00000291300.2 | ENSG00000075399.14 |
| ENSG00000288856.3 | ENSG00000297999.1 | ENSG00000272604.1 | ENSG00000179627.10 |
| ENSG00000301691.1 | ENSG00000255794.13 | ENSG00000149634.5 | ENSG00000271643.3 |
| ENSG00000167996.16 | ENSG00000136279.21 | ENSG00000306694.1 | ENSG00000271868.1 |
| ENSG00000224186.8 | ENSG00000105447.13 | ENSG00000004809.14 | ENSG00000144648.16 |
| ENSG00000288040.2 | ENSG00000257872.1 | ENSG00000289388.1 | ENSG00000157184.7 |
| ENSG00000109436.8 | ENSG00000115221.13 | ENSG00000114491.14 | ENSG00000180596.9 |
| ENSG00000198771.11 | ENSG00000162909.18 | ENSG00000211797.2 | ENSG00000079999.14 |
| ENSG00000162227.8 | ENSG00000292363.1 | ENSG00000177352.10 | ENSG00000177406.6 |
| ENSG00000153930.13 | ENSG00000116824.5 | ENSG00000286548.1 | ENSG00000154016.14 |
| ENSG00000297346.1 | ENSG00000289102.1 | ENSG00000297896.1 | ENSG00000305874.1 |
| ENSG00000116701.16 | ENSG00000290713.2 | ENSG00000213964.3 | ENSG00000285816.1 |
| ENSG00000260698.1 | ENSG00000263482.3 | ENSG00000229562.1 | ENSG00000117971.12 |
| ENSG00000163814.8 | ENSG00000137474.23 | ENSG00000207547.1 | ENSG00000229036.8 |
| ENSG00000155666.12 | ENSG00000135392.19 | ENSG00000174175.17 | ENSG00000300948.1 |
| ENSG00000287475.1 | ENSG00000296732.1 | ENSG00000135521.9 | ENSG00000302646.1 |
| ENSG00000072952.21 | ENSG00000126353.4 | ENSG00000233555.2 | ENSG00000196096.4 |
| ENSG00000146859.6 | ENSG00000269235.5 | ENSG00000233058.4 | ENSG00000288021.2 |
| ENSG00000000938.13 | ENSG00000111335.14 | ENSG00000187244.12 | ENSG00000185504.17 |
| ENSG00000304244.1 | ENSG00000131781.13 | ENSG00000114395.11 | ENSG00000216740.2 |
| ENSG00000284693.3 | ENSG00000026751.17 | ENSG00000183597.16 | ENSG00000228162.3 |
| ENSG00000129194.8 | ENSG00000295488.1 | ENSG00000292333.1 | ENSG00000103042.9 |
| ENSG00000307305.1 | ENSG00000127084.19 | ENSG00000131408.15 | ENSG00000285640.2 |
| ENSG00000230310.1 | ENSG00000085117.13 | ENSG00000227933.2 | ENSG00000228172.6 |
| ENSG00000196155.13 | ENSG00000265688.2 | ENSG00000276718.1 | ENSG00000104883.8 |
| ENSG00000238186.1 | ENSG00000185614.7 | ENSG00000289110.2 | ENSG00000297212.1 |
| ENSG00000241697.5 | ENSG00000285895.1 | ENSG00000145835.6 | ENSG00000105699.17 |

| | | | |
|---|---|---|---|
| ENSG00000243725.7 | ENSG00000013503.10 | ENSG00000295685.1 | ENSG00000288956.3 |
| ENSG00000162739.14 | ENSG00000303096.1 | ENSG00000105889.16 | ENSG00000135763.10 |
| ENSG00000197483.11 | ENSG00000286033.2 | ENSG00000069812.11 | ENSG00000294236.1 |
| ENSG00000213888.4 | ENSG00000155926.14 | ENSG00000302136.1 | ENSG00000196366.3 |
| ENSG00000166979.13 | ENSG00000131848.10 | ENSG00000115159.17 | ENSG00000211959.2 |
| ENSG00000106006.6 | ENSG00000131981.16 | ENSG00000151117.9 | ENSG00000070413.21 |
| ENSG00000187676.8 | ENSG00000286388.1 | ENSG00000292991.1 | ENSG00000197362.15 |
| ENSG00000198890.9 | ENSG00000249346.9 | ENSG00000289827.2 | ENSG00000060709.17 |
| ENSG00000010539.12 | ENSG00000144749.14 | ENSG00000258300.2 | ENSG00000198246.10 |
| ENSG00000143365.20 | ENSG00000293610.1 | ENSG00000301754.1 | ENSG00000106608.17 |
| ENSG00000225111.4 | ENSG00000261584.1 | ENSG00000227431.7 | ENSG00000165804.16 |
| ENSG00000289073.2 | ENSG00000302769.1 | ENSG00000164485.15 | ENSG00000228544.1 |
| ENSG00000297865.1 | ENSG00000220875.1 | ENSG00000228559.3 | ENSG00000302611.1 |
| ENSG00000169439.12 | ENSG00000188158.17 | ENSG00000175294.6 | ENSG00000235546.2 |
| ENSG00000297953.1 | ENSG00000309941.1 | ENSG00000259002.3 | ENSG00000157110.16 |
| ENSG00000272647.3 | ENSG00000305799.1 | ENSG00000310446.1 | ENSG00000185345.25 |
| ENSG00000293600.2 | ENSG00000226349.2 | ENSG00000048162.22 | ENSG00000257135.9 |
| ENSG00000304837.1 | ENSG00000138449.11 | ENSG00000248122.1 | ENSG00000129749.3 |
| ENSG00000155714.14 | ENSG00000257405.1 | ENSG00000196810.6 | ENSG00000286480.1 |
| ENSG00000260871.1 | ENSG00000101846.9 | ENSG00000259207.10 | ENSG00000164077.15 |
| ENSG00000251988.1 | ENSG00000143786.8 | ENSG00000187862.13 | ENSG00000288990.3 |
| ENSG00000216809.1 | ENSG00000149474.15 | ENSG00000197951.11 | ENSG00000281357.6 |
| ENSG00000307217.1 | ENSG00000287222.2 | ENSG00000207399.1 | ENSG00000171766.17 |
| ENSG00000158941.17 | ENSG00000303518.1 | ENSG00000126246.10 | ENSG00000164530.15 |
| ENSG00000114993.17 | ENSG00000043462.13 | ENSG00000267454.6 | ENSG00000175221.16 |
| ENSG00000171115.4 | ENSG00000072832.15 | ENSG00000288955.2 | ENSG00000111679.17 |
| ENSG00000288988.2 | ENSG00000149781.14 | ENSG00000251408.1 | ENSG00000167778.9 |
| ENSG00000132109.10 | ENSG00000276043.5 | ENSG00000296998.1 | ENSG00000196812.6 |
| ENSG00000282173.1 | ENSG00000178445.10 | ENSG00000216781.2 | ENSG00000066230.12 |
| ENSG00000160226.16 | ENSG00000270149.5 | ENSG00000075624.17 | ENSG00000083817.9 |
| ENSG00000172116.23 | ENSG00000271151.1 | ENSG00000099256.19 | ENSG00000151576.10 |
| ENSG00000244509.4 | ENSG00000114378.17 | ENSG00000233901.9 | ENSG00000225632.2 |
| ENSG00000131467.12 | ENSG00000149646.12 | ENSG00000231638.1 | ENSG00000175928.6 |
| ENSG00000179994.12 | ENSG00000116455.14 | ENSG00000302040.1 | ENSG00000257446.4 |
| ENSG00000099957.17 | ENSG00000111331.14 | ENSG00000125637.16 | ENSG00000234807.10 |
| ENSG00000126391.14 | ENSG00000132932.19 | ENSG00000211957.2 | ENSG00000158882.15 |

| | | | |
|---|---|---|---|
| ENSG00000162073.14 | ENSG00000104142.11 | ENSG00000205959.4 | ENSG00000289129.3 |
| ENSG00000185811.21 | ENSG00000129465.16 | ENSG00000288043.2 | ENSG00000309154.1 |
| ENSG00000188596.11 | ENSG00000234614.2 | ENSG00000233003.4 | ENSG00000229941.7 |
| ENSG00000294658.1 | ENSG00000108679.13 | ENSG00000262412.2 | ENSG00000223476.1 |
| ENSG00000089639.11 | ENSG00000175567.12 | ENSG00000270917.1 | ENSG00000310576.1 |
| ENSG00000086061.16 | ENSG00000249345.9 | ENSG00000275070.1 | ENSG00000232118.2 |
| ENSG00000297383.1 | ENSG00000221882.5 | ENSG00000131043.13 | ENSG00000132793.12 |
| ENSG00000289296.3 | ENSG00000076258.10 | ENSG00000157895.11 | ENSG00000259477.1 |
| ENSG00000228613.2 | ENSG00000131943.20 | ENSG00000197863.9 | ENSG00000279259.1 |
| ENSG00000213471.10 | ENSG00000006042.12 | ENSG00000188176.12 | ENSG00000178252.19 |
| ENSG00000197768.11 | ENSG00000300321.1 | ENSG00000272374.1 | ENSG00000288568.2 |
| ENSG00000180509.13 | ENSG00000123643.14 | ENSG00000286359.2 | ENSG00000308132.1 |
| ENSG00000237222.5 | ENSG00000177646.20 | ENSG00000306611.1 | ENSG00000179242.16 |
| ENSG00000309252.1 | ENSG00000238062.7 | ENSG00000308454.1 | ENSG00000133313.15 |
| ENSG00000269535.1 | ENSG00000090612.22 | ENSG00000182871.16 | ENSG00000108312.16 |
| ENSG00000228034.1 | ENSG00000258539.1 | ENSG00000132405.19 | ENSG00000130347.13 |
| ENSG00000165025.15 | ENSG00000287077.2 | ENSG00000189184.12 | ENSG00000011009.12 |
| ENSG00000224043.10 | ENSG00000196632.12 | ENSG00000264269.2 | ENSG00000182718.18 |
| ENSG00000102931.8 | ENSG00000259710.1 | ENSG00000221887.6 | ENSG00000109113.20 |
| ENSG00000033011.14 | ENSG00000197451.12 | ENSG00000261693.1 | ENSG00000290916.2 |
| ENSG00000100557.10 | ENSG00000133561.15 | ENSG00000279792.1 | ENSG00000284052.2 |
| ENSG00000293426.2 | ENSG00000300014.1 | ENSG00000162396.6 | ENSG00000293473.2 |
| ENSG00000309338.1 | ENSG00000265763.4 | ENSG00000272610.1 | ENSG00000011376.12 |
| ENSG00000117595.13 | ENSG00000274290.3 | ENSG00000275417.1 | ENSG00000148057.16 |
| ENSG00000280356.2 | ENSG00000258072.2 | ENSG00000213920.9 | ENSG00000169955.8 |
| ENSG00000174292.13 | ENSG00000164818.16 | ENSG00000168040.5 | ENSG00000297011.1 |
| ENSG00000156521.14 | ENSG00000156097.13 | ENSG00000205220.12 | ENSG00000049769.14 |
| ENSG00000268858.3 | ENSG00000204576.12 | ENSG00000267432.6 | ENSG00000167880.8 |
| ENSG00000132004.13 | ENSG00000218052.8 | ENSG00000227459.1 | ENSG00000152128.13 |
| ENSG00000256294.8 | ENSG00000166133.18 | ENSG00000260572.2 | ENSG00000180257.14 |
| ENSG00000251201.8 | ENSG00000230487.10 | ENSG00000288809.2 | ENSG00000173894.11 |
| ENSG00000188229.6 | ENSG00000133195.12 | ENSG00000101082.14 | ENSG00000173218.15 |
| ENSG00000254065.1 | ENSG00000130545.16 | ENSG00000111667.14 | ENSG00000257727.6 |
| ENSG00000101187.16 | ENSG00000306624.1 | ENSG00000188191.16 | ENSG00000128218.8 |
| ENSG00000114867.22 | ENSG00000255378.1 | ENSG00000197548.13 | ENSG00000059377.19 |
| ENSG00000263327.7 | ENSG00000293420.2 | ENSG00000131634.14 | ENSG00000259856.1 |

| | | | |
|---|---|---|---|
| ENSG00000101222.12 | ENSG00000140691.18 | ENSG00000255441.1 | ENSG00000265451.2 |
| ENSG00000166444.19 | ENSG00000136371.11 | ENSG00000306471.1 | ENSG00000119574.13 |
| ENSG00000308645.1 | ENSG00000053372.5 | ENSG00000259236.1 | ENSG00000162367.12 |
| ENSG00000290891.1 | ENSG00000271978.3 | ENSG00000260053.2 | ENSG00000288838.2 |
| ENSG00000110851.12 | ENSG00000285427.1 | ENSG00000287292.2 | ENSG00000170365.10 |
| ENSG00000157637.13 | ENSG00000291206.2 | ENSG00000286070.2 | ENSG00000235750.10 |
| ENSG00000066294.15 | ENSG00000167083.7 | ENSG00000298933.1 | ENSG00000144455.14 |
| ENSG00000134824.14 | ENSG00000272195.2 | ENSG00000169031.21 | ENSG00000260464.1 |
| ENSG00000213380.16 | ENSG00000251598.3 | ENSG00000171056.8 | ENSG00000176723.10 |
| ENSG00000237423.4 | ENSG00000182551.14 | ENSG00000276576.1 | ENSG00000260339.3 |
| ENSG00000239920.2 | ENSG00000215270.3 | ENSG00000297944.1 | ENSG00000196116.8 |
| ENSG00000182329.14 | ENSG00000310107.1 | ENSG00000135549.16 | ENSG00000182318.7 |
| ENSG00000234568.3 | ENSG00000217702.4 | ENSG00000260273.3 | ENSG00000188636.4 |
| ENSG00000010165.20 | ENSG00000007923.17 | ENSG00000252552.1 | ENSG00000229002.1 |
| ENSG00000122121.12 | ENSG00000176024.18 | ENSG00000295969.1 | ENSG00000093217.11 |
| ENSG00000111875.8 | ENSG00000291225.2 | ENSG00000116981.4 | ENSG00000294538.1 |
| ENSG00000289234.3 | ENSG00000265100.3 | ENSG00000293643.1 | ENSG00000259024.7 |
| ENSG00000174684.7 | ENSG00000020129.16 | ENSG00000169994.20 | ENSG00000256628.5 |
| ENSG00000042286.15 | ENSG00000301228.1 | ENSG00000259169.1 | ENSG00000182704.8 |
| ENSG00000125877.13 | ENSG00000124104.19 | ENSG00000084090.14 | ENSG00000111254.8 |
| ENSG00000109062.12 | ENSG00000196405.13 | ENSG00000015676.18 | ENSG00000141040.15 |
| ENSG00000109743.12 | ENSG00000212123.4 | ENSG00000137713.16 | ENSG00000307406.1 |
| ENSG00000233093.9 | ENSG00000154930.15 | ENSG00000139211.7 | ENSG00000172794.20 |
| ENSG00000113648.17 | ENSG00000181830.9 | ENSG00000239322.1 | ENSG00000287784.2 |
| ENSG00000309312.1 | ENSG00000272849.1 | ENSG00000037474.15 | ENSG00000309290.1 |
| ENSG00000236824.2 | ENSG00000188549.13 | ENSG00000072422.17 | ENSG00000100239.16 |
| ENSG00000171174.15 | ENSG00000286681.1 | ENSG00000124459.12 | ENSG00000246375.6 |
| ENSG00000269296.2 | ENSG00000198853.12 | ENSG00000014216.16 | ENSG00000296294.1 |
| ENSG00000104833.12 | ENSG00000068654.17 | ENSG00000269343.9 | ENSG00000269161.1 |
| ENSG00000010671.18 | ENSG00000113845.10 | ENSG00000264968.2 | ENSG00000148719.16 |
| ENSG00000118855.21 | ENSG00000166272.18 | ENSG00000251867.4 | ENSG00000139372.15 |
| ENSG00000301220.1 | ENSG00000204659.5 | ENSG00000116990.12 | ENSG00000289617.1 |
| ENSG00000165271.17 | ENSG00000136877.16 | ENSG00000128973.13 | ENSG00000178977.5 |
| ENSG00000160193.12 | ENSG00000234776.5 | ENSG00000168496.4 | ENSG00000237788.1 |
| ENSG00000248015.7 | ENSG00000166819.12 | ENSG00000178685.14 | ENSG00000147586.10 |
| ENSG00000049283.19 | ENSG00000309151.1 | ENSG00000112578.11 | ENSG00000309455.1 |

| | | | |
|---|---|---|---|
| ENSG00000187624.9 | ENSG00000134013.16 | ENSG00000165164.14 | ENSG00000133246.12 |
| ENSG00000167740.10 | ENSG00000301063.1 | ENSG00000115325.14 | ENSG00000262700.1 |
| ENSG00000286084.4 | ENSG00000182810.7 | ENSG00000214286.2 | ENSG00000133895.19 |
| ENSG00000274849.1 | ENSG00000159650.9 | ENSG00000263164.1 | ENSG00000142303.14 |
| ENSG00000267260.4 | ENSG00000120820.13 | ENSG00000286810.2 | ENSG00000128039.13 |
| ENSG00000280431.1 | ENSG00000258701.1 | ENSG00000298409.1 | ENSG00000135482.7 |
| ENSG00000181577.16 | ENSG00000273473.1 | ENSG00000171608.18 | ENSG00000015285.12 |
| ENSG00000175137.11 | ENSG00000266153.1 | ENSG00000258429.2 | ENSG00000173171.15 |
| ENSG00000148840.11 | ENSG00000140104.14 | ENSG00000163933.11 | ENSG00000254369.7 |
| ENSG00000123146.20 | ENSG00000230438.10 | ENSG00000140854.13 | ENSG00000127580.17 |
| ENSG00000278383.1 | ENSG00000212556.1 | ENSG00000227799.1 | ENSG00000009790.15 |
| ENSG00000302966.1 | ENSG00000161920.11 | ENSG00000075790.12 | ENSG00000290613.2 |
| ENSG00000228285.2 | ENSG00000293211.1 | ENSG00000294462.1 | ENSG00000111199.12 |
| ENSG00000279641.1 | ENSG00000071539.14 | ENSG00000302786.1 | ENSG00000125266.8 |
| ENSG00000204718.3 | ENSG00000160991.17 | ENSG00000104518.11 | ENSG00000128606.13 |
| ENSG00000174903.16 | ENSG00000292349.2 | ENSG00000126903.16 | ENSG00000284084.2 |
| ENSG00000307273.1 | ENSG00000299391.1 | ENSG00000255517.9 | ENSG00000270870.1 |
| ENSG00000128191.17 | ENSG00000165972.13 | ENSG00000125510.18 | ENSG00000159339.13 |
| ENSG00000174327.7 | ENSG00000204540.11 | ENSG00000204993.3 | ENSG00000169403.12 |
| ENSG00000167114.13 | ENSG00000246145.2 | ENSG00000302338.1 | ENSG00000147100.11 |
| ENSG00000213047.13 | ENSG00000254389.4 | ENSG00000250878.6 | ENSG00000134057.15 |
| ENSG00000236151.1 | ENSG00000198951.12 | ENSG00000233456.2 | ENSG00000243485.6 |
| ENSG00000232917.5 | ENSG00000160051.12 | ENSG00000207181.1 | ENSG00000168329.14 |
| ENSG00000102547.19 | ENSG00000183346.9 | ENSG00000281852.2 | ENSG00000224424.9 |
| ENSG00000268883.2 | ENSG00000141469.18 | ENSG00000100664.11 | ENSG00000213190.4 |
| ENSG00000181929.13 | ENSG00000147206.17 | ENSG00000132386.11 | ENSG00000268870.1 |
| ENSG00000046653.15 | ENSG00000236519.3 | ENSG00000121406.9 | ENSG00000142512.15 |
| ENSG00000110321.19 | ENSG00000196839.14 | ENSG00000152969.21 | ENSG00000087266.18 |
| ENSG00000138829.14 | ENSG00000103274.11 | ENSG00000197043.15 | ENSG00000164010.15 |
| ENSG00000259224.3 | ENSG00000251414.1 | ENSG00000028116.18 | ENSG00000175130.7 |
| ENSG00000271816.1 | ENSG00000196345.13 | ENSG00000103043.15 | ENSG00000174527.11 |
| ENSG00000251417.3 | ENSG00000139055.7 | ENSG00000166676.17 | ENSG00000212237.1 |
| ENSG00000170915.9 | ENSG00000182636.8 | ENSG00000289866.2 | ENSG00000305839.1 |
| ENSG00000292373.2 | ENSG00000158806.14 | ENSG00000117593.12 | ENSG00000136932.16 |
| ENSG00000164011.18 | ENSG00000203734.13 | ENSG00000141524.19 | ENSG00000099783.13 |
| ENSG00000198624.13 | ENSG00000275219.1 | ENSG00000206535.8 | ENSG00000267546.3 |

| | | | |
|---|---|---|---|
| ENSG00000122986.14 | ENSG00000221949.6 | ENSG00000293229.1 | ENSG00000284707.3 |
| ENSG00000171450.6 | ENSG00000172831.14 | ENSG00000249241.3 | ENSG00000185875.13 |
| ENSG00000179115.11 | ENSG00000158234.13 | ENSG00000117013.17 | ENSG00000187186.15 |
| ENSG00000150456.11 | ENSG00000206652.1 | ENSG00000197665.8 | ENSG00000132591.12 |
| ENSG00000116133.13 | ENSG00000306882.1 | ENSG00000300791.1 | ENSG00000130037.5 |
| ENSG00000234964.4 | ENSG00000163823.4 | ENSG00000138131.4 | ENSG00000261151.1 |
| ENSG00000132768.14 | ENSG00000256124.7 | ENSG00000115526.11 | ENSG00000289606.1 |
| ENSG00000294638.1 | ENSG00000123444.14 | ENSG00000168077.14 | ENSG00000230679.2 |
| ENSG00000301230.1 | ENSG00000274922.4 | ENSG00000261342.1 | ENSG00000293338.1 |
| ENSG00000100599.16 | ENSG00000132589.16 | ENSG00000288709.1 | ENSG00000121210.16 |
| ENSG00000137098.14 | ENSG00000279539.1 | ENSG00000152270.10 | ENSG00000273124.2 |
| ENSG00000004866.22 | ENSG00000110934.13 | ENSG00000175536.7 | ENSG00000300261.1 |
| ENSG00000215113.7 | ENSG00000164100.9 | ENSG00000275405.1 | ENSG00000258818.4 |
| ENSG00000308355.1 | ENSG00000241852.11 | ENSG00000108958.4 | ENSG00000271477.1 |
| ENSG00000128815.20 | ENSG00000135632.12 | ENSG00000288722.1 | ENSG00000279406.1 |
| ENSG00000169442.9 | ENSG00000282780.1 | ENSG00000119487.17 | ENSG00000293192.2 |
| ENSG00000291223.1 | ENSG00000111684.11 | ENSG00000104951.16 | ENSG00000228966.2 |
| ENSG00000150637.9 | ENSG00000242474.4 | ENSG00000300374.1 | ENSG00000105639.21 |
| ENSG00000143390.18 | ENSG00000173327.9 | ENSG00000132481.7 | ENSG00000142528.16 |
| ENSG00000273568.1 | ENSG00000164961.16 | ENSG00000245954.11 | ENSG00000236643.2 |
| ENSG00000249375.9 | ENSG00000160602.14 | ENSG00000204581.3 | ENSG00000253483.1 |
| ENSG00000215472.10 | ENSG00000224152.3 | ENSG00000251602.8 | ENSG00000140932.10 |
| ENSG00000256862.6 | ENSG00000272455.1 | ENSG00000062282.15 | ENSG00000299671.1 |
| ENSG00000235488.1 | ENSG00000091137.14 | ENSG00000167208.16 | ENSG00000207554.1 |
| ENSG00000164002.12 | ENSG00000255468.8 | ENSG00000148248.14 | ENSG00000160818.17 |
| ENSG00000129667.13 | ENSG00000298791.1 | ENSG00000231718.2 | ENSG00000136827.12 |
| ENSG00000111203.13 | ENSG00000300674.1 | ENSG00000079785.16 | ENSG00000257108.2 |
| ENSG00000266094.8 | ENSG00000185090.15 | ENSG00000132182.13 | ENSG00000184436.12 |
| ENSG00000307470.1 | ENSG00000258881.6 | ENSG00000299346.1 | ENSG00000220201.8 |
| ENSG00000063244.14 | ENSG00000259431.6 | ENSG00000158488.16 | ENSG00000224452.1 |
| ENSG00000109066.14 | ENSG00000226238.1 | ENSG00000181026.15 | ENSG00000290729.2 |
| ENSG00000178031.18 | ENSG00000250764.1 | ENSG00000293143.1 | ENSG00000296859.1 |
| ENSG00000196724.13 | ENSG00000129480.14 | ENSG00000250896.1 | ENSG00000064601.21 |
| ENSG00000130775.16 | ENSG00000230658.3 | ENSG00000163157.15 | ENSG00000175806.15 |
| ENSG00000105607.14 | ENSG00000254788.7 | ENSG00000123080.12 | ENSG00000128604.21 |
| ENSG00000183091.21 | ENSG00000261620.1 | ENSG00000172349.19 | ENSG00000130511.16 |

| | | | |
|---|---|---|---|
| ENSG00000162913.10 | ENSG00000254409.4 | ENSG00000261236.8 | ENSG00000259314.1 |
| ENSG00000173585.18 | ENSG00000108883.13 | ENSG00000235106.11 | ENSG00000162144.10 |
| ENSG00000117682.18 | ENSG00000126709.16 | ENSG00000197142.11 | ENSG00000235008.1 |
| ENSG00000240204.3 | ENSG00000179761.12 | ENSG00000250030.2 | ENSG00000163344.6 |
| ENSG00000027075.18 | ENSG00000167112.10 | ENSG00000090316.16 | ENSG00000307411.1 |
| ENSG00000147912.13 | ENSG00000099985.4 | ENSG00000170473.17 | ENSG00000236144.10 |
| ENSG00000250053.1 | ENSG00000169105.8 | ENSG00000105556.12 | ENSG00000136997.22 |
| ENSG00000103266.11 | ENSG00000115649.16 | ENSG00000280037.1 | ENSG00000256683.7 |
| ENSG00000167705.12 | ENSG00000100294.13 | ENSG00000224698.2 | ENSG00000298243.1 |
| ENSG00000211958.2 | ENSG00000272509.1 | ENSG00000135454.14 | ENSG00000215246.7 |
| ENSG00000079691.19 | ENSG00000173456.5 | ENSG00000224051.7 | ENSG00000091732.18 |
| ENSG00000196924.19 | ENSG00000255506.1 | ENSG00000167094.16 | ENSG00000274349.5 |
| ENSG00000225822.5 | ENSG00000277161.2 | ENSG00000198899.2 | ENSG00000300498.1 |
| ENSG00000150768.18 | ENSG00000154767.16 | ENSG00000135636.16 | ENSG00000235863.4 |
| ENSG00000033050.9 | ENSG00000277491.1 | ENSG00000308927.1 | ENSG00000290292.10 |
| ENSG00000158164.7 | ENSG00000253409.1 | ENSG00000206582.1 | ENSG00000272825.1 |
| ENSG00000162650.17 | ENSG00000250273.1 | ENSG00000126088.14 | ENSG00000178150.10 |
| ENSG00000100987.15 | ENSG00000272733.1 | ENSG00000234928.4 | ENSG00000100029.18 |
| ENSG00000170873.19 | ENSG00000145685.14 | ENSG00000197024.9 | ENSG00000185813.11 |
| ENSG00000307990.1 | ENSG00000233170.4 | ENSG00000183317.17 | ENSG00000237980.3 |
| ENSG00000095319.14 | ENSG00000136250.13 | ENSG00000176473.14 | ENSG00000023191.17 |
| ENSG00000164347.18 | ENSG00000115232.14 | ENSG00000230870.2 | ENSG00000093072.19 |
| ENSG00000224848.5 | ENSG00000162510.6 | ENSG00000145916.19 | ENSG00000270361.2 |
| ENSG00000291095.2 | ENSG00000261335.2 | ENSG00000242485.6 | ENSG00000188690.16 |
| ENSG00000186919.13 | ENSG00000307341.1 | ENSG00000286276.2 | ENSG00000276809.1 |
| ENSG00000293636.1 | ENSG00000130303.14 | ENSG00000272798.1 | ENSG00000290866.2 |
| ENSG00000050730.16 | ENSG00000271361.1 | ENSG00000159496.15 | ENSG00000117245.13 |
| ENSG00000239978.1 | ENSG00000276744.1 | ENSG00000005882.12 | ENSG00000106025.9 |
| ENSG00000307387.1 | ENSG00000213513.3 | ENSG00000198298.13 | ENSG00000299214.1 |
| ENSG00000159842.16 | ENSG00000164053.22 | ENSG00000176049.16 | ENSG00000240571.1 |
| ENSG00000272031.3 | ENSG00000156966.8 | ENSG00000142039.4 | ENSG00000229043.4 |
| ENSG00000185238.13 | ENSG00000289888.2 | ENSG00000183048.12 | ENSG00000162341.18 |
| ENSG00000153395.11 | ENSG00000089472.18 | ENSG00000063127.16 | ENSG00000087303.19 |
| ENSG00000185475.11 | ENSG00000145022.5 | ENSG00000182993.5 | ENSG00000104140.7 |
| ENSG00000198700.10 | ENSG00000153563.17 | ENSG00000117525.14 | ENSG00000265511.1 |
| ENSG00000255344.3 | ENSG00000159231.6 | ENSG00000092330.19 | ENSG00000309658.1 |

| | | | |
|---|---|---|---|
| ENSG00000132530.17 | ENSG00000309654.1 | ENSG00000092820.19 | ENSG00000102743.16 |
| ENSG00000100350.16 | ENSG00000114013.16 | ENSG00000213024.13 | ENSG00000160285.15 |
| ENSG00000065183.16 | ENSG00000171492.14 | ENSG00000175711.9 | ENSG00000175220.12 |
| ENSG00000265828.1 | ENSG00000162194.13 | ENSG00000242616.4 | ENSG00000114757.19 |
| ENSG00000184719.13 | ENSG00000287780.2 | ENSG00000167552.16 | ENSG00000294101.1 |
| ENSG00000204580.14 | ENSG00000142513.6 | ENSG00000120913.24 | ENSG00000228084.4 |
| ENSG00000076248.12 | ENSG00000159063.14 | ENSG00000267533.1 | ENSG00000096080.12 |
| ENSG00000086289.13 | ENSG00000117395.13 | ENSG00000132361.18 | ENSG00000298390.1 |
| ENSG00000305018.1 | ENSG00000298887.1 | ENSG00000177303.10 | ENSG00000182141.11 |
| ENSG00000186377.8 | ENSG00000273361.2 | ENSG00000282939.1 | ENSG00000287133.2 |
| ENSG00000156398.13 | ENSG00000261573.2 | ENSG00000163154.6 | ENSG00000164627.18 |
| ENSG00000293332.2 | ENSG00000138614.16 | ENSG00000144043.13 | ENSG00000249035.8 |
| ENSG00000132128.18 | ENSG00000235655.3 | ENSG00000125863.20 | ENSG00000166704.13 |
| ENSG00000008226.20 | ENSG00000121900.19 | ENSG00000171552.14 | ENSG00000179222.18 |
| ENSG00000083838.17 | ENSG00000112559.15 | ENSG00000141642.9 | ENSG00000159267.17 |
| ENSG00000228549.5 | ENSG00000213859.8 | ENSG00000224870.10 | ENSG00000172339.10 |
| ENSG00000234286.4 | ENSG00000176422.14 | ENSG00000308597.1 | ENSG00000160214.13 |
| ENSG00000166938.13 | ENSG00000132600.18 | ENSG00000163803.13 | ENSG00000140853.17 |
| ENSG00000291137.2 | ENSG00000187792.5 | ENSG00000304642.1 | ENSG00000151067.23 |
| ENSG00000160325.15 | ENSG00000130377.14 | ENSG00000228076.2 | ENSG00000258824.2 |
| ENSG00000100461.18 | ENSG00000181019.13 | ENSG00000288933.3 | ENSG00000171385.11 |
| ENSG00000232699.3 | ENSG00000155660.11 | ENSG00000286086.2 | ENSG00000189292.17 |
| ENSG00000183715.15 | ENSG00000177426.22 | ENSG00000294920.1 | ENSG00000118246.14 |
| ENSG00000104147.9 | ENSG00000130479.11 | ENSG00000293560.3 | ENSG00000226711.10 |
| ENSG00000141076.18 | ENSG00000215045.9 | ENSG00000304841.1 | ENSG00000065268.11 |
| ENSG00000294501.1 | ENSG00000291045.2 | ENSG00000287511.1 | ENSG00000266947.4 |
| ENSG00000114455.15 | ENSG00000123472.13 | ENSG00000290839.2 | ENSG00000204438.11 |
| ENSG00000155275.19 | ENSG00000076928.20 | ENSG00000286312.2 | ENSG00000160117.16 |
| ENSG00000136715.19 | ENSG00000120949.15 | ENSG00000306566.1 | ENSG00000267498.3 |
| ENSG00000138794.10 | ENSG00000105058.12 | ENSG00000309388.1 | ENSG00000166261.11 |
| ENSG00000198055.11 | ENSG00000103356.18 | ENSG00000237440.9 | ENSG00000170425.4 |
| ENSG00000113504.21 | ENSG00000233247.3 | ENSG00000179101.5 | ENSG00000029993.15 |
| ENSG00000141219.16 | ENSG00000302539.1 | ENSG00000115207.15 | ENSG00000024422.12 |
| ENSG00000235806.2 | ENSG00000160404.18 | ENSG00000023902.14 | ENSG00000143569.20 |
| ENSG00000123213.23 | ENSG00000103174.13 | ENSG00000272764.1 | ENSG00000189233.12 |
| ENSG00000293468.2 | ENSG00000249014.2 | ENSG00000265354.4 | ENSG00000308980.1 |

| | | | |
|---|---|---|---|
| ENSG00000111671.10 | ENSG00000068976.14 | ENSG00000300544.1 | ENSG00000177192.14 |
| ENSG00000167613.16 | ENSG00000230299.1 | ENSG00000250403.1 | ENSG00000173540.14 |
| ENSG00000211699.2 | ENSG00000289421.2 | ENSG00000188931.4 | ENSG00000063322.15 |
| ENSG00000286281.1 | ENSG00000110777.12 | ENSG00000115155.19 | ENSG00000308909.1 |
| ENSG00000280789.2 | ENSG00000254008.2 | ENSG00000279082.6 | ENSG00000254614.2 |
| ENSG00000064201.16 | ENSG00000186704.9 | ENSG00000289841.1 | ENSG00000279794.1 |
| ENSG00000279464.1 | ENSG00000277196.4 | ENSG00000213470.4 | ENSG00000256771.5 |
| ENSG00000106868.17 | ENSG00000188277.10 | ENSG00000255446.1 | ENSG00000250853.1 |
| ENSG00000027001.10 | ENSG00000103707.10 | ENSG00000258500.2 | ENSG00000303092.1 |
| ENSG00000163219.12 | ENSG00000158477.7 | ENSG00000197841.15 | ENSG00000070159.14 |
| ENSG00000143061.18 | ENSG00000187801.15 | ENSG00000305313.1 | ENSG00000138074.15 |
| ENSG00000155115.7 | ENSG00000178814.17 | ENSG00000150510.17 | ENSG00000235109.8 |
| ENSG00000171103.11 | ENSG00000236656.2 | ENSG00000106336.13 | ENSG00000165102.15 |
| ENSG00000174028.6 | ENSG00000175305.18 | ENSG00000286652.2 | ENSG00000179144.5 |
| ENSG00000228852.6 | ENSG00000136870.11 | ENSG00000223799.4 | ENSG00000298775.1 |
| ENSG00000174586.11 | ENSG00000090432.7 | ENSG00000174177.13 | ENSG00000169371.14 |
| ENSG00000268575.1 | ENSG00000135637.15 | ENSG00000263731.1 | ENSG00000178971.17 |
| ENSG00000080608.10 | ENSG00000232090.2 | ENSG00000127399.15 | ENSG00000100297.16 |
| ENSG00000048140.18 | ENSG00000134297.9 | ENSG00000073111.15 | ENSG00000143368.10 |
| ENSG00000115539.15 | ENSG00000294928.1 | ENSG00000151690.15 | ENSG00000124608.5 |
| ENSG00000169410.11 | ENSG00000304066.1 | ENSG00000157653.12 | ENSG00000215835.2 |
| ENSG00000185252.19 | ENSG00000198954.9 | ENSG00000254986.8 | ENSG00000258744.4 |
| ENSG00000206337.13 | ENSG00000162066.16 | ENSG00000224165.9 | ENSG00000171611.10 |
| ENSG00000173898.16 | ENSG00000274523.5 | ENSG00000170468.8 | ENSG00000185220.12 |
| ENSG00000177932.7 | ENSG00000076662.11 | ENSG00000129535.13 | ENSG00000229809.9 |
| ENSG00000233296.3 | ENSG00000145907.16 | ENSG00000012232.10 | ENSG00000286132.1 |
| ENSG00000116353.16 | ENSG00000166192.15 | ENSG00000176371.14 | ENSG00000183918.18 |
| ENSG00000105171.10 | ENSG00000040531.16 | ENSG00000289494.3 | ENSG00000308729.1 |
| ENSG00000231856.5 | ENSG00000179256.2 | ENSG00000295124.1 | ENSG00000197798.9 |
| ENSG00000205913.8 | ENSG00000185730.8 | ENSG00000174680.10 | ENSG00000049089.15 |
| ENSG00000145860.13 | ENSG00000116922.15 | ENSG00000226753.1 | ENSG00000165730.16 |
| ENSG00000117899.11 | ENSG00000300428.1 | ENSG00000272631.2 | ENSG00000126062.4 |
| ENSG00000102181.22 | ENSG00000142961.16 | ENSG00000132938.22 | ENSG00000157992.12 |
| ENSG00000135083.17 | ENSG00000099910.17 | ENSG00000288187.2 | ENSG00000259953.3 |
| ENSG00000277782.2 | ENSG00000132854.19 | ENSG00000109586.12 | ENSG00000178927.19 |
| ENSG00000171649.12 | ENSG00000309304.1 | ENSG00000248569.1 | ENSG00000131037.15 |

| | | | |
|---|---|---|---|
| ENSG00000160233.8 | ENSG00000227741.2 | ENSG00000180198.17 | ENSG00000168421.14 |
| ENSG00000111845.5 | ENSG00000156265.17 | ENSG00000188917.15 | ENSG00000100473.18 |
| ENSG00000167716.19 | ENSG00000184508.12 | ENSG00000237868.1 | ENSG00000197150.13 |
| ENSG00000281538.1 | ENSG00000167528.13 | ENSG00000002549.13 | ENSG00000163520.14 |
| ENSG00000185340.16 | ENSG00000278709.2 | ENSG00000132394.11 | ENSG00000139719.12 |
| ENSG00000166526.19 | ENSG00000307565.1 | ENSG00000239392.2 | ENSG00000115041.14 |
| ENSG00000137965.11 | ENSG00000197054.12 | ENSG00000165816.13 | ENSG00000005156.12 |
| ENSG00000143799.14 | ENSG00000136573.16 | ENSG00000167851.16 | ENSG00000250220.2 |
| ENSG00000225806.9 | ENSG00000274554.1 | ENSG00000310066.1 | ENSG00000008283.16 |
| ENSG00000138795.10 | ENSG00000042062.13 | ENSG00000297240.1 | ENSG00000286918.1 |
| ENSG00000015171.21 | ENSG00000159873.10 | ENSG00000289883.2 | ENSG00000083290.20 |
| ENSG00000105248.16 | ENSG00000084073.10 | ENSG00000227028.7 | ENSG00000213117.4 |
| ENSG00000169188.5 | ENSG00000101945.17 | ENSG00000286408.1 | ENSG00000230366.11 |
| ENSG00000288829.1 | ENSG00000275318.1 | ENSG00000132153.15 | ENSG00000295889.1 |
| ENSG00000136518.17 | ENSG00000112195.9 | ENSG00000254027.2 | ENSG00000105063.19 |
| ENSG00000288945.1 | ENSG00000117280.13 | ENSG00000259032.2 | ENSG00000143418.20 |
| ENSG00000167553.17 | ENSG00000137806.10 | ENSG00000167695.15 | ENSG00000286311.2 |
| ENSG00000106477.20 | ENSG00000162931.12 | ENSG00000160691.19 | ENSG00000303694.1 |
| ENSG00000303764.1 | ENSG00000132274.17 | ENSG00000261338.2 | ENSG00000222033.1 |
| ENSG00000184489.13 | ENSG00000260648.3 | ENSG00000160221.18 | ENSG00000145982.13 |
| ENSG00000258056.4 | ENSG00000125378.16 | ENSG00000213638.6 | ENSG00000150873.12 |
| ENSG00000164068.16 | ENSG00000159184.8 | ENSG00000261839.1 | ENSG00000162377.6 |
| ENSG00000177599.13 | ENSG00000230513.5 | ENSG00000220771.2 | ENSG00000260174.1 |
| ENSG00000138080.14 | ENSG00000100218.12 | ENSG00000131473.17 | ENSG00000006468.15 |
| ENSG00000222018.2 | ENSG00000071246.11 | ENSG00000089335.22 | ENSG00000298669.1 |
| ENSG00000303329.1 | ENSG00000232060.3 | ENSG00000237594.2 | ENSG00000175792.12 |
| ENSG00000115946.8 | ENSG00000117586.12 | ENSG00000013725.14 | ENSG00000129255.16 |
| ENSG00000145703.17 | ENSG00000177889.10 | ENSG00000070614.15 | ENSG00000186666.6 |
| ENSG00000185482.8 | ENSG00000181396.13 | ENSG00000157927.17 | ENSG00000197124.12 |
| ENSG00000137841.12 | ENSG00000065457.11 | ENSG00000204767.4 | ENSG00000261371.6 |
| ENSG00000245848.3 | ENSG00000135437.10 | ENSG00000273447.1 | ENSG00000296581.1 |
| ENSG00000186111.11 | ENSG00000162894.12 | ENSG00000224034.2 | ENSG00000289511.2 |
| ENSG00000068079.8 | ENSG00000100271.17 | ENSG00000251667.1 | ENSG00000186575.20 |
| ENSG00000289555.2 | ENSG00000184828.10 | ENSG00000164062.13 | ENSG00000132434.10 |
| ENSG00000111602.12 | ENSG00000248712.8 | ENSG00000177076.6 | ENSG00000073849.16 |
| ENSG00000014164.7 | ENSG00000113088.6 | ENSG00000157613.11 | ENSG00000169710.9 |

| | | | |
|---|---|---|---|
| ENSG00000256988.2 | ENSG00000184361.13 | ENSG00000204261.9 | ENSG00000101307.16 |
| ENSG00000144744.18 | ENSG00000289596.2 | ENSG00000180581.7 | ENSG00000274209.7 |
| ENSG00000213626.13 | ENSG00000221886.4 | ENSG00000170819.5 | ENSG00000225025.2 |
| ENSG00000125843.12 | ENSG00000124251.11 | ENSG00000140382.15 | ENSG00000265996.1 |
| ENSG00000247728.3 | ENSG00000302636.1 | ENSG00000182508.14 | ENSG00000241945.8 |
| ENSG00000111206.13 | ENSG00000294552.1 | ENSG00000182473.22 | ENSG00000301866.3 |
| ENSG00000110719.11 | ENSG00000055130.18 | ENSG00000277156.1 | ENSG00000229237.2 |
| ENSG00000187266.14 | ENSG00000111110.12 | ENSG00000223723.1 | ENSG00000101246.20 |
| ENSG00000147439.14 | ENSG00000205060.11 | ENSG00000180626.10 | ENSG00000267549.7 |
| ENSG00000167554.15 | ENSG00000204682.10 | ENSG00000235316.1 | ENSG00000130518.18 |
| ENSG00000204604.13 | ENSG00000090674.16 | ENSG00000189152.12 | ENSG00000176894.10 |
| ENSG00000143753.14 | ENSG00000139508.15 | ENSG00000225725.5 | ENSG00000174206.13 |
| ENSG00000196659.10 | ENSG00000083454.23 | ENSG00000131477.11 | ENSG00000103047.8 |
| ENSG00000149182.15 | ENSG00000042088.14 | ENSG00000250999.3 | ENSG00000143337.19 |
| ENSG00000241839.10 | ENSG00000172995.17 | ENSG00000228661.2 | ENSG00000102893.18 |
| ENSG00000110074.12 | ENSG00000177854.8 | ENSG00000129226.14 | ENSG00000100243.22 |
| ENSG00000196663.16 | ENSG00000119965.13 | ENSG00000283667.1 | ENSG00000129675.16 |
| ENSG00000170854.18 | ENSG00000198551.10 | ENSG00000178458.5 | ENSG00000280013.1 |
| ENSG00000158481.13 | ENSG00000100462.16 | ENSG00000196372.13 | ENSG00000298052.1 |
| ENSG00000302261.1 | ENSG00000167264.18 | ENSG00000162600.12 | ENSG00000273901.1 |
| ENSG00000213536.2 | ENSG00000130734.10 | ENSG00000165406.16 | ENSG00000304289.1 |
| ENSG00000308900.1 | ENSG00000197937.13 | ENSG00000272858.1 | ENSG00000273674.7 |
| ENSG00000299474.1 | ENSG00000187164.20 | ENSG00000117461.15 | ENSG00000234558.1 |
| ENSG00000286113.3 | ENSG00000281856.1 | ENSG00000184730.12 | ENSG00000226268.3 |
| ENSG00000259460.2 | ENSG00000299687.1 | ENSG00000163704.13 | ENSG00000187325.5 |
| ENSG00000261886.2 | ENSG00000250584.4 | ENSG00000101888.12 | ENSG00000178947.9 |
| ENSG00000252274.1 | ENSG00000286430.1 | ENSG00000255725.2 | ENSG00000288877.2 |
| ENSG00000100399.16 | ENSG00000214773.1 | ENSG00000297847.1 | ENSG00000137767.14 |
| ENSG00000159720.13 | ENSG00000135439.12 | ENSG00000150783.10 | ENSG00000135144.8 |
| ENSG00000129195.16 | ENSG00000176248.9 | ENSG00000289389.2 | ENSG00000291230.2 |
| ENSG00000122390.19 | ENSG00000304745.1 | ENSG00000224023.11 | ENSG00000171860.5 |
| ENSG00000290808.2 | ENSG00000289956.2 | ENSG00000114745.15 | ENSG00000309328.1 |
| ENSG00000145431.11 | ENSG00000101940.18 | ENSG00000289406.2 | ENSG00000125445.11 |
| ENSG00000167768.4 | ENSG00000131061.15 | ENSG00000128322.7 | ENSG00000168237.18 |
| ENSG00000218350.1 | ENSG00000155629.15 | ENSG00000102699.6 | ENSG00000231579.3 |
| ENSG00000123329.20 | ENSG00000102178.13 | ENSG00000105662.16 | ENSG00000165055.16 |

| | | | |
|---|---|---|---|
| ENSG00000038358.15 | ENSG00000254473.3 | ENSG00000132507.18 | ENSG00000260608.2 |
| ENSG00000167720.13 | ENSG00000155034.21 | ENSG00000290767.2 | ENSG00000171223.6 |
| ENSG00000289056.3 | ENSG00000168792.5 | ENSG00000118407.15 | ENSG00000089723.10 |
| ENSG00000165474.8 | ENSG00000176635.18 | ENSG00000115317.13 | ENSG00000171161.13 |
| ENSG00000232882.1 | ENSG00000119333.12 | ENSG00000171793.17 | ENSG00000233010.1 |
| ENSG00000171970.14 | ENSG00000140406.4 | ENSG00000136243.18 | ENSG00000289399.3 |
| ENSG00000143942.5 | ENSG00000162804.15 | ENSG00000100395.15 | ENSG00000265458.4 |
| ENSG00000128408.9 | ENSG00000104880.19 | ENSG00000160219.12 | ENSG00000143494.16 |
| ENSG00000261408.7 | ENSG00000295880.1 | ENSG00000254858.10 | ENSG00000079435.11 |
| ENSG00000163508.13 | ENSG00000171790.16 | ENSG00000244462.8 | ENSG00000229056.3 |
| ENSG00000126106.14 | ENSG00000122642.11 | ENSG00000122971.9 | ENSG00000123815.13 |
| ENSG00000160688.19 | ENSG00000240889.2 | ENSG00000128833.13 | ENSG00000184967.7 |
| ENSG00000138442.11 | ENSG00000276136.1 | ENSG00000304084.1 | ENSG00000227191.8 |
| ENSG00000196290.17 | ENSG00000095596.12 | ENSG00000150457.9 | ENSG00000197933.13 |
| ENSG00000134326.12 | ENSG00000227591.7 | ENSG00000011485.15 | ENSG00000229666.3 |
| ENSG00000172508.11 | ENSG00000186897.6 | ENSG00000144579.9 | ENSG00000126773.13 |
| ENSG00000104368.19 | ENSG00000227914.3 | ENSG00000123609.11 | ENSG00000269983.1 |
| ENSG00000137274.14 | ENSG00000304302.1 | ENSG00000196743.9 | ENSG00000142185.18 |
| ENSG00000144649.10 | ENSG00000220685.3 | ENSG00000184923.12 | ENSG00000141522.13 |
| ENSG00000198324.14 | ENSG00000172731.15 | ENSG00000169427.8 | ENSG00000138867.17 |
| ENSG00000139641.13 | ENSG00000148331.12 | ENSG00000108219.17 | ENSG00000134308.14 |
| ENSG00000234043.3 | ENSG00000103005.13 | ENSG00000176410.8 | ENSG00000157017.16 |
| ENSG00000198431.18 | ENSG00000253917.5 | ENSG00000248527.1 | ENSG00000161999.13 |
| ENSG00000306927.1 | ENSG00000136213.10 | ENSG00000223612.3 | ENSG00000293615.1 |
| ENSG00000275464.6 | ENSG00000189306.12 | ENSG00000283005.1 | ENSG00000100994.12 |
| ENSG00000100726.15 | ENSG00000163673.8 | ENSG00000184939.16 | ENSG00000294163.1 |
| ENSG00000100982.12 | ENSG00000035681.10 | ENSG00000157657.15 | ENSG00000128311.14 |
| ENSG00000117597.18 | ENSG00000267106.10 | ENSG00000178665.16 | ENSG00000130717.13 |
| ENSG00000050130.18 | ENSG00000138496.17 | ENSG00000254224.3 | ENSG00000168936.12 |
| ENSG00000164116.17 | ENSG00000293546.1 | ENSG00000148459.16 | ENSG00000149527.18 |
| ENSG00000154734.16 | ENSG00000101076.20 | ENSG00000179151.13 | ENSG00000267583.6 |
| ENSG00000141449.16 | ENSG00000249471.8 | ENSG00000106624.11 | ENSG00000251303.1 |
| ENSG00000278156.4 | ENSG00000108423.16 | ENSG00000115758.14 | ENSG00000261645.8 |
| ENSG00000269802.2 | ENSG00000112584.14 | ENSG00000181638.20 | ENSG00000293982.1 |
| ENSG00000238141.3 | ENSG00000205744.10 | ENSG00000149476.16 | ENSG00000013523.12 |
| ENSG00000124232.11 | ENSG00000144504.16 | ENSG00000256269.11 | ENSG00000141655.18 |

| | | | |
|---|---|---|---|
| ENSG00000142149.9 | ENSG00000153066.13 | ENSG00000184385.2 | ENSG00000204569.10 |
| ENSG00000130590.14 | ENSG00000189298.14 | ENSG00000157870.17 | ENSG00000213965.4 |
| ENSG00000183978.8 | ENSG00000243667.7 | ENSG00000268603.3 | ENSG00000087589.17 |
| ENSG00000143106.13 | ENSG00000124422.12 | ENSG00000298585.1 | ENSG00000153406.14 |
| ENSG00000082781.12 | ENSG00000140750.17 | ENSG00000197858.12 | ENSG00000283183.5 |
| ENSG00000307584.1 | ENSG00000180096.13 | ENSG00000204394.13 | ENSG00000232485.5 |
| ENSG00000063241.8 | ENSG00000224790.4 | ENSG00000276931.1 | ENSG00000218725.2 |
| ENSG00000213949.10 | ENSG00000277283.2 | ENSG00000211973.2 | ENSG00000215912.13 |
| ENSG00000113838.13 | ENSG00000180867.12 | ENSG00000273783.3 | ENSG00000123810.9 |
| ENSG00000198870.8 | ENSG00000143850.17 | ENSG00000136925.15 | ENSG00000130038.10 |
| ENSG00000089820.17 | ENSG00000289962.2 | ENSG00000123453.19 | ENSG00000149809.17 |
| ENSG00000159921.20 | ENSG00000305425.1 | ENSG00000089195.15 | ENSG00000108179.14 |
| ENSG00000204025.8 | ENSG00000132773.12 | ENSG00000257179.1 | ENSG00000301697.1 |
| ENSG00000122694.16 | ENSG00000143570.19 | ENSG00000276093.1 | ENSG00000198356.12 |
| ENSG00000303898.1 | ENSG00000163576.18 | ENSG00000136271.11 | ENSG00000163755.9 |
| ENSG00000250286.2 | ENSG00000167986.15 | ENSG00000130684.14 | ENSG00000033170.17 |
| ENSG00000178082.6 | ENSG00000115297.11 | ENSG00000213406.3 | ENSG00000154511.12 |
| ENSG00000137094.16 | ENSG00000165837.12 | ENSG00000182180.14 | ENSG00000141012.13 |
| ENSG00000226885.1 | ENSG00000213699.9 | ENSG00000288784.1 | ENSG00000083720.13 |
| ENSG00000099203.7 | ENSG00000233020.1 | ENSG00000259039.3 | ENSG00000213213.14 |
| ENSG00000136848.19 | ENSG00000258735.2 | ENSG00000271947.1 | ENSG00000165125.22 |
| ENSG00000164647.9 | ENSG00000078596.11 | ENSG00000165874.14 | ENSG00000160803.8 |
| ENSG00000284428.1 | ENSG00000153029.16 | ENSG00000134802.18 | ENSG00000101003.12 |
| ENSG00000175463.12 | ENSG00000133265.11 | ENSG00000164308.18 | ENSG00000060069.18 |
| ENSG00000068120.15 | ENSG00000234155.2 | ENSG00000160410.15 | ENSG00000188404.10 |
| ENSG00000289549.1 | ENSG00000100311.17 | ENSG00000094914.14 | ENSG00000235335.4 |
| ENSG00000074706.14 | ENSG00000113721.14 | ENSG00000291179.2 | ENSG00000060642.12 |
| ENSG00000213189.5 | ENSG00000131899.13 | ENSG00000145050.19 | ENSG00000184500.16 |
| ENSG00000228801.9 | ENSG00000273156.2 | ENSG00000179253.3 | ENSG00000280071.5 |
| ENSG00000140632.17 | ENSG00000085491.17 | ENSG00000184302.7 | ENSG00000299029.1 |
| ENSG00000124233.12 | ENSG00000198301.12 | ENSG00000291081.2 | ENSG00000147457.14 |
| ENSG00000022976.16 | ENSG00000246898.3 | ENSG00000085982.16 | ENSG00000168913.7 |
| ENSG00000255893.1 | ENSG00000187778.14 | ENSG00000258682.1 | ENSG00000301717.1 |
| ENSG00000110200.9 | ENSG00000263846.1 | ENSG00000167703.15 | ENSG00000182866.18 |
| ENSG00000214803.3 | ENSG00000131711.15 | ENSG00000156853.13 | ENSG00000116649.10 |
| ENSG00000121486.12 | ENSG00000163521.16 | ENSG00000197852.12 | ENSG00000197943.13 |

| | | | |
|---|---|---|---|
| ENSG00000140675.13 | ENSG00000131504.18 | ENSG00000188368.11 | ENSG00000130204.13 |
| ENSG00000105991.10 | ENSG00000232774.8 | ENSG00000165506.15 | ENSG00000040608.14 |
| ENSG00000148803.13 | ENSG00000263326.1 | ENSG00000171169.9 | ENSG00000226445.3 |
| ENSG00000198053.12 | ENSG00000158006.14 | ENSG00000133316.16 | ENSG00000107651.15 |
| ENSG00000108439.11 | ENSG00000227627.6 | ENSG00000146109.5 | ENSG00000151062.15 |
| ENSG00000154845.16 | ENSG00000280832.4 | ENSG00000168675.19 | ENSG00000272752.7 |
| ENSG00000225077.4 | ENSG00000307433.1 | ENSG00000185924.7 | ENSG00000107937.19 |
| ENSG00000110711.11 | ENSG00000304199.1 | ENSG00000198538.11 | ENSG00000087586.18 |
| ENSG00000260583.1 | ENSG00000277851.3 | ENSG00000162302.13 | ENSG00000140395.9 |
| ENSG00000160207.9 | ENSG00000302463.1 | ENSG00000294231.1 | ENSG00000136280.17 |
| ENSG00000272221.2 | ENSG00000167182.16 | ENSG00000253439.1 | ENSG00000035687.10 |
| ENSG00000135750.15 | ENSG00000234664.1 | ENSG00000140450.9 | ENSG00000167461.12 |
| ENSG00000260804.4 | ENSG00000100522.10 | ENSG00000027847.14 | ENSG00000031081.11 |
| ENSG00000117480.16 | ENSG00000277150.2 | ENSG00000067221.14 | ENSG00000173825.7 |
| ENSG00000276961.1 | ENSG00000167700.9 | ENSG00000210156.1 | ENSG00000187554.15 |
| ENSG00000126217.22 | ENSG00000108187.16 | ENSG00000152556.17 | ENSG00000063660.9 |
| ENSG00000112576.13 | ENSG00000226432.3 | ENSG00000134321.13 | ENSG00000146267.12 |
| ENSG00000162972.11 | ENSG00000059691.12 | ENSG00000117419.16 | ENSG00000235172.10 |
| ENSG00000119977.22 | ENSG00000243479.4 | ENSG00000177427.13 | ENSG00000211697.4 |
| ENSG00000110148.10 | ENSG00000249244.1 | ENSG00000132825.7 | ENSG00000176927.16 |
| ENSG00000072958.9 | ENSG00000132694.20 | ENSG00000107551.21 | ENSG00000184209.15 |
| ENSG00000183208.13 | ENSG00000213588.6 | ENSG00000108469.15 | ENSG00000021826.18 |
| ENSG00000293044.2 | ENSG00000140157.16 | ENSG00000304875.1 | ENSG00000148297.16 |
| ENSG00000003400.16 | ENSG00000274270.2 | ENSG00000273024.7 | ENSG00000198406.7 |
| ENSG00000159210.10 | ENSG00000115145.10 | ENSG00000148606.15 | ENSG00000122376.12 |
| ENSG00000111652.10 | ENSG00000281189.1 | ENSG00000196227.11 | ENSG00000105364.14 |
| ENSG00000263366.2 | ENSG00000264278.2 | ENSG00000015475.19 | ENSG00000099956.20 |
| ENSG00000298852.1 | ENSG00000136878.14 | ENSG00000229953.2 | ENSG00000203880.13 |
| ENSG00000087263.17 | ENSG00000198855.7 | ENSG00000107021.17 | ENSG00000113719.17 |
| ENSG00000163995.22 | ENSG00000106638.18 | ENSG00000126749.17 | ENSG00000204193.12 |
| ENSG00000101407.13 | ENSG00000294692.1 | ENSG00000170876.9 | ENSG00000188779.12 |
| ENSG00000178718.7 | ENSG00000216915.2 | ENSG00000177398.19 | |

Table S23. Downregulated Genes from DOX+ Jurkat Cells - Public Dataset from Jurkat Cell *ZNF683* Overexpression Experiment

GEO Accession: GSM7713745. Series: GSE241091. Downregulated Genes from DOX+ Jurkat Cells. $Log_2FC < -1$ , adjusted $p$ value $< 0.05$

| | | | |
|---|---|---|---|
| ENSG00000173110.8 | ENSG00000293614.1 | ENSG00000284138.1 | ENSG00000204390.10 |
| ENSG00000109846.9 | ENSG00000234203.2 | ENSG00000225195.2 | ENSG00000188004.11 |
| ENSG00000200879.1 | ENSG00000223305.1 | ENSG00000294644.1 | ENSG00000299947.1 |
| ENSG00000124253.11 | ENSG00000120937.9 | ENSG00000204389.11 | ENSG00000260170.1 |
| ENSG00000310471.1 | ENSG00000244998.2 | ENSG00000204388.7 | ENSG00000141431.14 |
| ENSG00000171804.11 | ENSG00000199732.1 | ENSG00000259967.1 | ENSG00000101180.17 |
| ENSG00000298649.1 | ENSG00000279137.1 | ENSG00000198597.10 | ENSG00000254649.1 |
| ENSG00000142871.18 | ENSG00000201343.1 | ENSG00000305265.1 | ENSG00000295515.1 |
| ENSG00000228677.1 | ENSG00000295692.1 | ENSG00000251056.1 | ENSG00000233783.9 |
| ENSG00000294609.1 | ENSG00000267703.1 | ENSG00000304463.1 | ENSG00000298926.1 |
| ENSG00000296167.1 | ENSG00000105370.8 | ENSG00000286514.2 | ENSG00000279217.1 |
| ENSG00000309804.1 | ENSG00000243314.1 | ENSG00000201616.1 | ENSG00000202184.1 |
| ENSG00000276097.1 | ENSG00000306702.1 | ENSG00000277382.2 | ENSG00000212296.1 |
| ENSG00000070985.14 | ENSG00000141579.7 | ENSG00000258779.9 | ENSG00000272330.1 |
| ENSG00000289340.3 | ENSG00000308465.1 | ENSG00000294623.1 | ENSG00000139219.19 |
| ENSG00000123095.6 | ENSG00000295381.1 | ENSG00000260740.2 | ENSG00000240454.1 |
| ENSG00000264966.1 | ENSG00000187546.14 | ENSG00000308662.1 | ENSG00000130222.11 |
| ENSG00000299729.1 | ENSG00000266618.1 | ENSG00000301737.1 | ENSG00000287330.1 |
| ENSG00000260550.3 | ENSG00000136928.7 | ENSG00000173530.6 | ENSG00000007933.13 |
| ENSG00000304760.1 | ENSG00000274183.2 | ENSG00000230515.2 | ENSG00000300285.1 |
| ENSG00000293328.2 | ENSG00000236426.6 | ENSG00000308762.1 | ENSG00000257057.3 |
| ENSG00000104755.16 | ENSG00000189056.15 | ENSG00000304298.1 | ENSG00000170426.2 |
| ENSG00000206816.1 | ENSG00000296096.1 | ENSG00000259134.8 | ENSG00000267339.9 |
| ENSG00000307007.1 | ENSG00000241246.3 | ENSG00000095917.15 | ENSG00000262097.4 |
| ENSG00000286643.1 | ENSG00000241484.10 | ENSG00000266573.7 | ENSG00000295424.1 |
| ENSG00000231689.4 | ENSG00000301646.1 | ENSG00000266541.1 | ENSG00000271410.1 |
| ENSG00000250592.1 | ENSG00000137441.8 | ENSG00000276845.1 | ENSG00000260596.5 |
| ENSG00000130052.14 | ENSG00000300008.1 | ENSG00000239840.1 | ENSG00000139973.18 |
| ENSG00000082293.13 | ENSG00000206172.8 | ENSG00000294576.1 | ENSG00000230661.1 |

| | | | |
|---|---|---|---|
| ENSG00000303379.1 | ENSG00000305423.1 | ENSG00000264324.1 | ENSG00000271681.1 |
| ENSG00000225126.2 | ENSG00000273213.4 | ENSG00000252768.1 | ENSG00000211749.1 |
| ENSG00000224318.7 | ENSG00000308463.1 | ENSG00000230910.7 | ENSG00000308267.1 |
| ENSG00000303175.1 | ENSG00000293905.1 | ENSG00000262120.1 | ENSG00000100095.19 |
| ENSG00000235535.10 | ENSG00000148926.10 | ENSG00000295613.1 | ENSG00000235038.1 |
| ENSG00000253154.5 | ENSG00000290021.1 | ENSG00000230046.1 | ENSG00000301958.1 |
| ENSG00000222302.1 | ENSG00000226658.1 | ENSG00000279485.1 | ENSG00000237738.1 |
| ENSG00000205396.12 | ENSG00000292347.1 | ENSG00000310049.1 | ENSG00000232600.3 |
| ENSG00000296836.1 | ENSG00000152208.13 | ENSG00000302394.1 | ENSG00000277246.1 |
| ENSG00000231729.1 | ENSG00000302790.1 | ENSG00000305283.1 | ENSG00000261436.7 |
| ENSG00000253342.1 | ENSG00000251421.2 | ENSG00000307499.1 | ENSG00000296960.1 |
| ENSG00000229853.1 | ENSG00000199855.1 | ENSG00000182348.7 | ENSG00000235385.4 |
| ENSG00000261430.1 | ENSG00000258632.2 | ENSG00000295687.1 | ENSG00000294418.1 |
| ENSG00000269959.1 | ENSG00000254843.1 | ENSG00000260230.5 | ENSG00000296731.1 |
| ENSG00000293016.1 | ENSG00000226620.9 | ENSG00000295051.1 | ENSG00000299101.1 |
| ENSG00000227043.4 | ENSG00000295919.1 | ENSG00000148671.15 | ENSG00000199672.1 |
| ENSG00000289331.2 | ENSG00000227881.1 | ENSG00000172987.14 | ENSG00000254560.7 |
| ENSG00000308828.1 | ENSG00000279513.1 | ENSG00000305052.1 | ENSG00000229613.4 |
| ENSG00000143333.7 | ENSG00000272491.1 | ENSG00000303473.1 | ENSG00000260865.1 |
| ENSG00000229305.1 | ENSG00000302141.1 | ENSG00000288768.2 | ENSG00000172362.4 |
| ENSG00000198758.11 | ENSG00000278909.1 | ENSG00000229246.2 | ENSG00000237373.1 |
| ENSG00000305776.1 | ENSG00000283201.1 | ENSG00000140015.20 | ENSG00000305635.1 |
| ENSG00000270190.1 | ENSG00000222067.1 | ENSG00000260658.8 | ENSG00000279354.1 |
| ENSG00000197506.8 | ENSG00000225450.1 | ENSG00000309888.1 | ENSG00000287530.2 |
| ENSG00000199411.1 | ENSG00000234683.1 | ENSG00000140832.11 | ENSG00000271437.1 |
| ENSG00000232373.2 | ENSG00000287591.2 | ENSG00000213304.3 | ENSG00000304899.1 |
| ENSG00000287109.1 | ENSG00000295026.1 | ENSG00000304571.1 | ENSG00000174226.9 |
| ENSG00000305134.1 | ENSG00000289459.2 | ENSG00000233162.1 | ENSG00000244383.2 |
| ENSG00000302462.1 | ENSG00000238288.1 | ENSG00000307179.1 | ENSG00000302839.1 |
| ENSG00000252590.1 | ENSG00000177791.12 | ENSG00000235244.7 | ENSG00000301775.1 |
| ENSG00000231824.4 | ENSG00000249961.10 | ENSG00000308592.1 | ENSG00000225391.5 |
| ENSG00000278057.4 | ENSG00000260011.3 | ENSG00000177464.5 | ENSG00000230109.2 |
| ENSG00000130822.16 | ENSG00000266602.3 | ENSG00000304850.1 | ENSG00000286575.2 |
| ENSG00000199381.1 | ENSG00000269320.1 | ENSG00000201942.1 | ENSG00000259415.1 |
| ENSG00000290106.2 | ENSG00000232778.1 | ENSG00000240760.1 | ENSG00000207083.1 |
| ENSG00000234361.2 | ENSG00000232163.2 | ENSG00000304665.1 | ENSG00000267748.4 |

| | | | |
|---|---|---|---|
| ENSG00000202150.1 | ENSG00000240553.1 | ENSG00000298693.1 | ENSG00000118492.19 |
| ENSG00000297263.1 | ENSG00000229010.1 | ENSG00000259695.2 | ENSG00000106536.21 |
| ENSG00000280408.1 | ENSG00000225087.3 | ENSG00000285829.2 | ENSG00000199627.1 |
| ENSG00000258323.1 | ENSG00000287039.2 | ENSG00000200013.1 | ENSG00000226899.1 |
| ENSG00000290783.1 | ENSG00000289159.3 | ENSG00000302007.1 | ENSG00000300967.1 |
| ENSG00000255237.1 | ENSG00000266407.1 | ENSG00000147255.19 | ENSG00000296360.1 |
| ENSG00000235606.3 | ENSG00000230928.1 | ENSG00000217455.9 | ENSG00000266644.1 |
| ENSG00000237480.4 | ENSG00000254865.1 | ENSG00000307937.1 | ENSG00000228835.1 |
| ENSG00000303559.1 | ENSG00000111058.8 | ENSG00000303542.1 | ENSG00000286201.2 |
| ENSG00000253056.1 | ENSG00000258866.3 | ENSG00000185631.7 | ENSG00000200882.1 |
| ENSG00000205038.12 | ENSG00000260128.7 | ENSG00000225832.1 | ENSG00000185070.12 |
| ENSG00000234502.2 | ENSG00000260541.1 | ENSG00000219274.1 | ENSG00000309281.1 |
| ENSG00000265190.7 | ENSG00000207042.1 | ENSG00000256315.2 | ENSG00000198995.3 |
| ENSG00000299052.1 | ENSG00000170579.19 | ENSG00000243313.2 | ENSG00000224477.6 |
| ENSG00000296955.1 | ENSG00000295778.1 | ENSG00000301662.1 | ENSG00000239572.4 |
| ENSG00000243116.1 | ENSG00000300315.1 | ENSG00000243945.1 | ENSG00000230831.2 |
| ENSG00000199536.1 | ENSG00000297457.1 | ENSG00000243033.2 | ENSG00000227512.1 |
| ENSG00000117983.17 | ENSG00000262294.2 | ENSG00000304678.1 | ENSG00000290420.2 |
| ENSG00000139220.17 | ENSG00000188536.13 | ENSG00000274766.3 | ENSG00000287432.2 |
| ENSG00000235573.2 | ENSG00000297593.1 | ENSG00000308431.1 | ENSG00000179818.18 |
| ENSG00000233928.8 | ENSG00000241156.3 | ENSG00000173917.11 | ENSG00000302926.1 |
| ENSG00000143199.18 | ENSG00000275139.1 | ENSG00000207047.2 | ENSG00000109047.8 |
| ENSG00000310110.1 | ENSG00000223113.1 | ENSG00000228939.1 | ENSG00000287889.2 |
| ENSG00000280162.1 | ENSG00000207326.1 | ENSG00000301014.1 | ENSG00000102243.13 |
| ENSG00000277027.2 | ENSG00000199545.1 | ENSG00000105219.10 | ENSG00000239946.1 |
| ENSG00000207008.1 | ENSG00000295148.1 | ENSG00000237387.1 | ENSG00000216519.2 |
| ENSG00000160870.15 | ENSG00000299238.1 | ENSG00000235262.1 | ENSG00000183888.4 |
| ENSG00000309126.1 | ENSG00000283296.1 | ENSG00000196240.6 | ENSG00000233478.1 |
| ENSG00000256250.1 | ENSG00000179342.4 | ENSG00000287003.2 | ENSG00000294297.1 |
| ENSG00000231966.2 | ENSG00000308510.1 | ENSG00000306409.1 | ENSG00000235002.1 |
| ENSG00000297825.1 | ENSG00000233290.5 | ENSG00000304163.1 | ENSG00000308284.1 |
| ENSG00000310076.1 | ENSG00000229742.1 | ENSG00000260035.1 | ENSG00000164023.15 |
| ENSG00000167749.12 | ENSG00000305212.1 | ENSG00000267098.2 | ENSG00000249690.1 |
| ENSG00000278340.1 | ENSG00000166351.11 | ENSG00000177993.4 | ENSG00000113430.11 |
| ENSG00000105675.9 | ENSG00000239805.1 | ENSG00000297637.1 | ENSG00000250328.8 |
| ENSG00000204478.11 | ENSG00000248701.1 | ENSG00000215124.2 | ENSG00000249996.2 |

| | | | |
|---|---|---|---|
| ENSG00000294166.1 | ENSG00000236564.1 | ENSG00000178795.10 | ENSG00000196263.8 |
| ENSG00000087085.16 | ENSG00000264932.3 | ENSG00000255929.7 | ENSG00000188800.6 |
| ENSG00000240015.2 | ENSG00000225331.3 | ENSG00000255483.1 | ENSG00000225521.2 |
| ENSG00000293877.1 | ENSG00000279652.1 | ENSG00000255580.1 | ENSG00000231668.2 |
| ENSG00000286295.1 | ENSG00000207525.1 | ENSG00000221491.2 | ENSG00000299193.1 |
| ENSG00000279286.2 | ENSG00000276337.1 | ENSG00000259125.2 | ENSG00000158517.16 |
| ENSG00000259654.1 | ENSG00000261267.1 | ENSG00000228573.1 | ENSG00000270682.1 |
| ENSG00000197476.3 | ENSG00000231208.5 | ENSG00000304780.1 | ENSG00000215165.3 |
| ENSG00000299202.1 | ENSG00000301732.1 | ENSG00000296913.1 | ENSG00000304157.1 |
| ENSG00000238231.2 | ENSG00000305285.1 | ENSG00000039987.7 | ENSG00000258739.1 |
| ENSG00000219150.2 | ENSG00000214748.2 | ENSG00000303209.1 | ENSG00000070729.14 |
| ENSG00000301463.1 | ENSG00000289201.2 | ENSG00000298665.1 | ENSG00000296554.1 |
| ENSG00000294388.1 | ENSG00000249992.2 | ENSG00000222810.1 | ENSG00000234030.1 |
| ENSG00000170345.11 | ENSG00000295445.1 | ENSG00000206638.1 | ENSG00000233541.4 |
| ENSG00000230623.9 | ENSG00000232825.4 | ENSG00000253665.3 | ENSG00000279214.1 |
| ENSG00000293851.1 | ENSG00000293080.2 | ENSG00000294889.1 | ENSG00000277545.1 |
| ENSG00000206848.1 | ENSG00000302457.1 | ENSG00000235224.1 | ENSG00000305037.1 |
| ENSG00000232628.8 | ENSG00000298207.1 | ENSG00000229014.3 | ENSG00000162992.5 |
| ENSG00000199477.1 | ENSG00000252414.1 | ENSG00000235226.1 | ENSG00000228877.3 |
| ENSG00000249234.1 | ENSG00000198203.10 | ENSG00000018625.15 | ENSG00000289864.2 |
| ENSG00000153707.20 | ENSG00000231534.1 | ENSG00000207450.1 | ENSG00000272620.2 |
| ENSG00000293824.1 | ENSG00000294113.1 | ENSG00000306073.1 | ENSG00000227714.1 |
| ENSG00000247595.5 | ENSG00000109255.11 | ENSG00000286675.2 | ENSG00000227117.7 |
| ENSG00000212615.1 | ENSG00000264678.1 | ENSG00000310307.1 | ENSG00000006283.18 |
| ENSG00000170689.10 | ENSG00000305691.1 | ENSG00000301396.1 | ENSG00000302856.1 |
| ENSG00000287393.2 | ENSG00000200275.1 | ENSG00000268289.1 | ENSG00000309599.1 |
| ENSG00000249669.13 | ENSG00000185002.10 | ENSG00000295578.1 | ENSG00000272337.1 |
| ENSG00000233381.4 | ENSG00000219409.2 | ENSG00000283509.1 | ENSG00000279658.2 |
| ENSG00000230647.4 | ENSG00000294978.1 | ENSG00000248587.9 | ENSG00000201724.1 |
| ENSG00000305049.1 | ENSG00000254319.7 | ENSG00000305437.1 | ENSG00000235597.4 |
| ENSG00000303790.1 | ENSG00000309479.1 | ENSG00000287092.2 | ENSG00000269068.2 |
| ENSG00000258490.1 | ENSG00000297850.1 | ENSG00000228742.12 | ENSG00000147509.14 |
| ENSG00000242493.3 | ENSG00000234534.1 | ENSG00000173535.15 | ENSG00000287272.2 |
| ENSG00000294304.1 | ENSG00000224245.1 | ENSG00000288761.3 | ENSG00000279085.1 |
| ENSG00000200488.1 | ENSG00000275024.1 | ENSG00000258933.2 | ENSG00000297780.1 |
| ENSG00000207109.1 | ENSG00000183378.13 | ENSG00000201034.1 | ENSG00000297562.1 |

| | | | |
|---|---|---|---|
| ENSG00000239686.1 | ENSG00000308262.1 | ENSG00000222585.1 | ENSG00000226496.4 |
| ENSG00000257759.2 | ENSG00000295248.1 | ENSG00000257604.1 | ENSG00000198576.4 |
| ENSG00000206937.1 | ENSG00000275924.1 | ENSG00000022556.16 | ENSG00000267476.2 |
| ENSG00000248469.2 | ENSG00000272104.1 | ENSG00000295337.1 | ENSG00000306493.1 |
| ENSG00000212207.1 | ENSG00000224797.1 | ENSG00000162981.14 | ENSG00000266467.2 |
| ENSG00000302080.1 | ENSG00000293176.2 | ENSG00000230408.4 | ENSG00000299150.1 |
| ENSG00000213937.4 | ENSG00000294204.1 | ENSG00000248416.1 | ENSG00000294546.1 |
| ENSG00000294948.1 | ENSG00000267764.2 | ENSG00000229949.1 | ENSG00000269506.2 |
| ENSG00000241593.6 | ENSG00000288964.2 | ENSG00000301886.1 | ENSG00000116741.8 |
| ENSG00000264630.5 | ENSG00000254629.1 | ENSG00000300073.1 | ENSG00000279691.1 |
| ENSG00000288928.1 | ENSG00000259921.1 | ENSG00000251381.8 | ENSG00000282022.1 |
| ENSG00000290674.2 | ENSG00000114115.11 | ENSG00000307203.1 | ENSG00000255321.1 |
| ENSG00000268707.1 | ENSG00000261357.1 | ENSG00000235347.1 | ENSG00000143801.18 |
| ENSG00000304805.1 | ENSG00000124216.4 | ENSG00000226878.2 | ENSG00000241067.2 |
| ENSG00000305860.1 | ENSG00000087074.9 | ENSG00000187957.8 | ENSG00000162069.16 |
| ENSG00000005243.10 | ENSG00000287001.4 | ENSG00000258337.1 | ENSG00000248100.2 |
| ENSG00000272533.1 | ENSG00000228386.2 | ENSG00000306330.1 | ENSG00000302098.1 |
| ENSG00000232684.1 | ENSG00000250033.6 | ENSG00000301484.1 | ENSG00000227560.1 |
| ENSG00000163273.4 | ENSG00000168309.18 | ENSG00000293846.1 | ENSG00000153993.14 |
| ENSG00000099860.10 | ENSG00000275607.1 | ENSG00000296912.1 | ENSG00000237768.2 |
| ENSG00000260744.1 | ENSG00000173627.8 | ENSG00000284966.3 | ENSG00000268100.1 |
| ENSG00000229954.1 | ENSG00000228411.1 | ENSG00000308778.1 | ENSG00000179862.7 |
| ENSG00000298718.1 | ENSG00000309414.1 | ENSG00000308288.1 | ENSG00000298593.1 |
| ENSG00000206650.1 | ENSG00000283538.4 | ENSG00000201428.1 | ENSG00000296078.1 |
| ENSG00000276926.1 | ENSG00000185046.21 | ENSG00000305552.1 | ENSG00000244310.3 |
| ENSG00000262074.7 | ENSG00000240498.10 | ENSG00000224529.1 | ENSG00000234521.2 |
| ENSG00000289829.2 | ENSG00000253112.1 | ENSG00000302293.1 | ENSG00000249001.7 |
| ENSG00000226476.8 | ENSG00000172940.13 | ENSG00000227619.4 | ENSG00000301290.1 |
| ENSG00000229452.2 | ENSG00000266579.1 | ENSG00000166268.11 | ENSG00000112246.10 |
| ENSG00000279044.1 | ENSG00000289461.3 | ENSG00000258603.6 | ENSG00000228467.2 |
| ENSG00000224943.2 | ENSG00000289373.2 | ENSG00000275092.1 | ENSG00000293799.1 |
| ENSG00000297456.1 | ENSG00000207233.1 | ENSG00000285863.1 | ENSG00000304388.1 |
| ENSG00000230499.1 | ENSG00000201749.1 | ENSG00000227528.7 | ENSG00000273172.2 |
| ENSG00000291007.1 | ENSG00000265942.2 | ENSG00000240423.4 | ENSG00000243813.1 |
| ENSG00000270661.1 | ENSG00000139832.5 | ENSG00000232978.2 | ENSG00000201109.1 |
| ENSG00000198488.10 | ENSG00000272256.1 | ENSG00000298112.1 | ENSG00000303824.1 |

| | | | |
|---|---|---|---|
| ENSG00000205181.9 | ENSG00000238728.1 | ENSG00000273524.1 | ENSG00000302580.1 |
| ENSG00000217195.1 | ENSG00000201185.1 | ENSG00000306300.1 | ENSG00000233998.1 |
| ENSG00000228210.1 | ENSG00000229558.3 | ENSG00000216471.3 | ENSG00000070669.17 |
| ENSG00000302198.1 | ENSG00000297779.1 | ENSG00000233622.4 | ENSG00000292997.1 |
| ENSG00000154252.12 | ENSG00000274747.1 | ENSG00000260918.1 | ENSG00000104972.16 |
| ENSG00000100292.18 | ENSG00000306015.1 | ENSG00000266245.1 | ENSG00000181847.12 |
| ENSG00000002745.13 | ENSG00000276712.1 | ENSG00000260438.1 | ENSG00000234068.7 |
| ENSG00000238290.2 | ENSG00000285737.2 | ENSG00000164283.13 | ENSG00000242170.3 |
| ENSG00000254105.1 | ENSG00000217653.1 | ENSG00000263301.1 | ENSG00000307714.1 |
| ENSG00000275586.1 | ENSG00000293294.2 | ENSG00000200291.1 | ENSG00000257964.1 |
| ENSG00000294255.1 | ENSG00000294450.1 | ENSG00000201882.1 | ENSG00000303569.1 |
| ENSG00000201592.1 | ENSG00000279685.2 | ENSG00000265185.6 | ENSG00000222259.1 |
| ENSG00000304565.1 | ENSG00000276102.1 | ENSG00000280035.1 | ENSG00000266498.1 |
| ENSG00000167105.8 | ENSG00000241334.2 | ENSG00000308515.1 | ENSG00000253190.5 |
| ENSG00000230806.2 | ENSG00000108551.5 | ENSG00000253596.2 | ENSG00000263120.1 |
| ENSG00000286155.1 | ENSG00000252759.1 | ENSG00000302057.1 | ENSG00000162438.12 |
| ENSG00000262869.2 | ENSG00000230916.2 | ENSG00000309202.1 | ENSG00000251253.1 |
| ENSG00000306426.1 | ENSG00000151882.11 | ENSG00000271418.1 | ENSG00000296131.1 |
| ENSG00000293826.1 | ENSG00000240069.1 | ENSG00000307046.1 | ENSG00000260840.3 |
| ENSG00000263813.1 | ENSG00000203971.3 | ENSG00000270610.1 | ENSG00000207231.1 |
| ENSG00000183921.8 | ENSG00000244040.8 | ENSG00000267393.1 | ENSG00000287269.2 |
| ENSG00000165617.16 | ENSG00000243532.3 | ENSG00000234222.7 | ENSG00000262141.1 |
| ENSG00000200731.1 | ENSG00000227236.2 | ENSG00000200164.1 | ENSG00000254846.1 |
| ENSG00000295556.1 | ENSG00000236427.3 | ENSG00000295652.1 | ENSG00000137875.4 |
| ENSG00000234428.2 | ENSG00000286870.1 | ENSG00000297914.1 | ENSG00000162433.15 |
| ENSG00000153292.16 | ENSG00000233656.3 | ENSG00000250129.6 | ENSG00000273521.1 |
| ENSG00000231605.6 | ENSG00000222432.1 | ENSG00000197410.14 | ENSG00000242028.9 |
| ENSG00000134115.13 | ENSG00000272915.1 | ENSG00000176697.20 | ENSG00000169551.13 |
| ENSG00000299864.1 | ENSG00000224038.2 | ENSG00000269300.1 | ENSG00000148053.18 |
| ENSG00000296268.1 | ENSG00000224761.5 | ENSG00000251614.1 | ENSG00000116703.14 |
| ENSG00000304024.1 | ENSG00000221042.1 | ENSG00000308309.1 | ENSG00000270789.1 |
| ENSG00000131126.19 | ENSG00000276951.1 | ENSG00000287970.1 | ENSG00000307885.1 |
| ENSG00000237913.1 | ENSG00000286402.1 | ENSG00000196586.17 | ENSG00000212370.1 |
| ENSG00000232048.2 | ENSG00000259043.1 | ENSG00000161640.15 | ENSG00000259589.2 |
| ENSG00000107159.14 | ENSG00000272372.1 | ENSG00000303180.1 | ENSG00000173080.6 |
| ENSG00000254450.1 | ENSG00000278941.1 | ENSG00000250786.5 | ENSG00000280023.1 |

| | | | |
|---|---|---|---|
| ENSG00000273999.1 | ENSG00000160856.21 | ENSG00000241745.3 | ENSG00000270098.1 |
| ENSG00000234848.1 | ENSG00000272254.1 | ENSG00000226876.4 | ENSG00000284638.2 |
| ENSG00000255491.5 | ENSG00000242498.8 | ENSG00000199695.1 | ENSG00000232545.1 |
| ENSG00000300410.1 | ENSG00000256981.1 | ENSG00000269099.2 | ENSG00000240328.1 |
| ENSG00000156218.13 | ENSG00000272084.1 | ENSG00000215184.2 | ENSG00000224895.1 |
| ENSG00000239659.2 | ENSG00000298452.1 | ENSG00000231334.3 | ENSG00000198910.14 |
| ENSG00000251774.1 | ENSG00000111729.15 | ENSG00000270230.1 | ENSG00000254205.2 |
| ENSG00000296009.1 | ENSG00000199436.1 | ENSG00000257316.1 | ENSG00000303479.1 |
| ENSG00000290077.1 | ENSG00000200926.1 | ENSG00000304890.1 | ENSG00000229054.1 |
| ENSG00000247925.3 | ENSG00000304733.1 | ENSG00000185133.15 | ENSG00000279971.1 |
| ENSG00000254898.1 | ENSG00000268119.9 | ENSG00000260910.3 | ENSG00000255804.2 |
| ENSG00000310433.1 | ENSG00000213946.3 | ENSG00000248498.5 | ENSG00000304763.1 |
| ENSG00000275830.5 | ENSG00000305438.1 | ENSG00000286938.2 | ENSG00000265733.1 |
| ENSG00000275162.1 | ENSG00000226216.1 | ENSG00000306189.1 | ENSG00000231310.4 |
| ENSG00000209702.1 | ENSG00000283036.4 | ENSG00000268598.1 | ENSG00000153495.11 |
| ENSG00000288837.2 | ENSG00000297830.1 | ENSG00000300952.1 | ENSG00000290001.2 |
| ENSG00000233613.6 | ENSG00000304225.1 | ENSG00000272834.1 | ENSG00000286349.1 |
| ENSG00000174990.8 | ENSG00000298116.1 | ENSG00000297998.1 | ENSG00000201680.1 |
| ENSG00000223418.1 | ENSG00000263177.1 | ENSG00000221514.1 | ENSG00000254442.1 |
| ENSG00000286048.1 | ENSG00000253633.3 | ENSG00000126856.16 | ENSG00000278896.1 |
| ENSG00000240184.7 | ENSG00000266708.1 | ENSG00000294284.1 | ENSG00000270379.6 |
| ENSG00000200496.1 | ENSG00000259367.1 | ENSG00000253269.1 | ENSG00000267690.2 |
| ENSG00000303560.1 | ENSG00000248268.1 | ENSG00000178460.18 | ENSG00000301607.1 |
| ENSG00000270036.1 | ENSG00000231764.12 | ENSG00000231491.2 | ENSG00000251791.1 |
| ENSG00000207320.1 | ENSG00000224451.2 | ENSG00000225236.1 | ENSG00000277882.1 |
| ENSG00000307396.1 | ENSG00000244332.1 | ENSG00000308280.1 | ENSG00000286020.2 |
| ENSG00000289166.2 | ENSG00000081479.15 | ENSG00000287002.1 | ENSG00000299634.1 |
| ENSG00000107562.18 | ENSG00000279020.1 | ENSG00000285190.2 | ENSG00000288831.3 |
| ENSG00000293390.1 | ENSG00000196431.4 | ENSG00000291155.2 | ENSG00000250359.1 |
| ENSG00000303642.1 | ENSG00000015413.10 | ENSG00000237853.7 | ENSG00000299743.1 |
| ENSG00000238585.1 | ENSG00000289528.1 | ENSG00000303299.1 | ENSG00000221400.2 |
| ENSG00000267717.1 | ENSG00000253613.3 | ENSG00000200312.1 | ENSG00000226159.3 |
| ENSG00000306608.1 | ENSG00000277572.1 | ENSG00000239129.1 | ENSG00000305090.1 |
| ENSG00000164904.18 | ENSG00000227195.12 | ENSG00000151715.8 | ENSG00000262154.1 |
| ENSG00000297452.1 | ENSG00000275515.1 | ENSG00000233511.1 | ENSG00000278214.2 |
| ENSG00000153551.14 | ENSG00000253385.1 | ENSG00000135406.15 | ENSG00000260750.7 |

| | | | |
|---|---|---|---|
| ENSG00000304181.1 | ENSG00000289869.2 | ENSG00000300939.1 | ENSG00000255348.2 |
| ENSG00000267333.1 | ENSG00000205696.6 | ENSG00000168938.6 | ENSG00000196890.5 |
| ENSG00000232884.12 | ENSG00000150045.12 | ENSG00000271825.1 | ENSG00000290864.1 |
| ENSG00000230899.2 | ENSG00000111057.11 | ENSG00000201229.1 | ENSG00000236893.1 |
| ENSG00000306602.1 | ENSG00000241183.1 | ENSG00000162630.6 | ENSG00000244671.3 |
| ENSG00000268681.1 | ENSG00000285519.1 | ENSG00000224977.3 | ENSG00000213994.4 |
| ENSG00000226744.1 | ENSG00000232144.1 | ENSG00000308339.1 | ENSG00000239615.1 |
| ENSG00000297151.1 | ENSG00000201044.1 | ENSG00000287934.1 | ENSG00000293182.1 |
| ENSG00000256433.3 | ENSG00000290845.2 | ENSG00000222511.2 | ENSG00000249192.1 |
| ENSG00000224259.10 | ENSG00000229875.1 | ENSG00000161551.15 | ENSG00000294211.1 |
| ENSG00000293946.1 | ENSG00000271503.7 | ENSG00000267563.1 | ENSG00000208028.1 |
| ENSG00000253047.1 | ENSG00000293093.1 | ENSG00000286418.2 | ENSG00000168135.4 |
| ENSG00000250472.1 | ENSG00000233072.1 | ENSG00000187905.11 | ENSG00000199933.1 |
| ENSG00000263883.1 | ENSG00000301082.1 | ENSG00000266371.1 | ENSG00000174951.12 |
| ENSG00000204091.9 | ENSG00000277235.1 | ENSG00000266086.3 | ENSG00000270990.1 |
| ENSG00000227123.1 | ENSG00000261216.1 | ENSG00000231104.9 | ENSG00000252577.1 |
| ENSG00000202078.1 | ENSG00000239570.1 | ENSG00000289050.2 | ENSG00000300517.1 |
| ENSG00000300394.1 | ENSG00000309940.1 | ENSG00000286957.2 | ENSG00000206636.1 |
| ENSG00000298331.1 | ENSG00000258274.2 | ENSG00000305533.1 | ENSG00000286444.1 |
| ENSG00000279463.1 | ENSG00000141979.4 | ENSG00000180712.6 | ENSG00000113389.16 |
| ENSG00000227676.5 | ENSG00000295663.1 | ENSG00000294980.1 | ENSG00000298322.1 |
| ENSG00000295473.1 | ENSG00000171657.6 | ENSG00000237187.10 | ENSG00000251733.1 |
| ENSG00000159167.12 | ENSG00000273483.1 | ENSG00000109471.5 | ENSG00000258572.2 |
| ENSG00000247473.2 | ENSG00000232526.1 | ENSG00000223721.3 | ENSG00000137819.14 |
| ENSG00000259033.3 | ENSG00000310296.1 | ENSG00000199683.1 | ENSG00000290721.2 |
| ENSG00000217767.2 | ENSG00000270640.1 | ENSG00000235145.2 | ENSG00000181143.17 |
| ENSG00000310465.1 | ENSG00000253516.1 | ENSG00000310385.1 | ENSG00000256642.1 |
| ENSG00000259196.1 | ENSG00000263535.2 | ENSG00000199697.1 | ENSG00000201581.1 |
| ENSG00000255666.8 | ENSG00000230818.1 | ENSG00000198932.13 | ENSG00000256115.7 |
| ENSG00000307272.1 | ENSG00000283274.1 | ENSG00000242256.3 | ENSG00000263155.7 |
| ENSG00000297753.1 | ENSG00000293710.1 | ENSG00000289567.3 | ENSG00000260395.1 |
| ENSG00000298318.1 | ENSG00000304189.1 | ENSG00000287351.1 | ENSG00000262966.2 |
| ENSG00000279345.1 | ENSG00000250672.1 | ENSG00000310392.1 | ENSG00000253058.1 |
| ENSG00000305793.1 | ENSG00000259905.7 | ENSG00000287715.1 | ENSG00000267496.5 |
| ENSG00000242290.2 | ENSG00000251273.5 | ENSG00000205955.4 | ENSG00000232448.3 |
| ENSG00000151490.15 | ENSG00000225479.2 | ENSG00000269321.1 | ENSG00000252695.1 |

| | | | |
|---|---|---|---|
| ENSG00000138892.12 | ENSG00000227945.1 | ENSG00000289208.1 | ENSG00000199630.1 |
| ENSG00000250371.1 | ENSG00000252657.1 | ENSG00000234231.4 | ENSG00000149531.15 |
| ENSG00000309745.1 | ENSG00000254208.1 | ENSG00000296677.1 | ENSG00000100558.9 |
| ENSG00000294437.1 | ENSG00000258471.2 | ENSG00000235303.2 | ENSG00000265745.2 |
| ENSG00000258091.1 | ENSG00000270690.1 | ENSG00000223188.1 | ENSG00000200086.1 |
| ENSG00000104432.15 | ENSG00000270000.1 | ENSG00000224333.1 | ENSG00000236681.1 |
| ENSG00000176490.5 | ENSG00000306404.1 | ENSG00000132170.24 | ENSG00000302237.1 |
| ENSG00000204113.6 | ENSG00000168418.8 | ENSG00000145864.14 | ENSG00000235278.2 |
| ENSG00000309402.1 | ENSG00000289609.3 | ENSG00000233388.2 | ENSG00000278819.1 |
| ENSG00000301617.1 | ENSG00000259933.6 | ENSG00000110675.13 | ENSG00000290005.2 |
| ENSG00000253577.3 | ENSG00000200332.1 | ENSG00000285601.2 | ENSG00000299897.1 |
| ENSG00000256913.1 | ENSG00000207031.1 | ENSG00000152672.8 | ENSG00000253532.1 |
| ENSG00000273407.1 | ENSG00000232655.2 | ENSG00000109321.11 | ENSG00000241744.1 |
| ENSG00000106366.9 | ENSG00000234481.3 | ENSG00000274111.1 | ENSG00000236133.3 |
| ENSG00000141434.12 | ENSG00000255504.2 | ENSG00000278959.1 | ENSG00000310144.1 |
| ENSG00000303332.1 | ENSG00000303808.1 | ENSG00000307729.1 | ENSG00000072315.4 |
| ENSG00000257531.2 | ENSG00000205129.9 | ENSG00000296354.1 | ENSG00000289287.2 |
| ENSG00000301902.1 | ENSG00000209082.1 | ENSG00000305592.1 | ENSG00000236928.3 |
| ENSG00000301649.1 | ENSG00000256482.1 | ENSG00000248290.1 | ENSG00000271996.1 |
| ENSG00000176641.11 | ENSG00000228446.2 | ENSG00000289587.3 | ENSG00000295782.1 |
| ENSG00000289534.1 | ENSG00000094755.17 | ENSG00000285857.1 | ENSG00000275358.2 |
| ENSG00000242278.1 | ENSG00000297071.1 | ENSG00000167355.8 | ENSG00000261873.3 |
| ENSG00000289741.1 | ENSG00000217786.1 | ENSG00000273802.3 | ENSG00000297286.1 |
| ENSG00000284670.1 | ENSG00000244036.4 | ENSG00000279684.1 | ENSG00000236986.6 |
| ENSG00000286533.2 | ENSG00000130958.13 | ENSG00000279532.1 | ENSG00000285204.1 |
| ENSG00000152154.12 | ENSG00000259810.2 | ENSG00000305869.1 | ENSG00000225434.6 |
| ENSG00000306939.1 | ENSG00000253623.1 | ENSG00000305337.1 | ENSG00000279265.1 |
| ENSG00000143341.12 | ENSG00000207117.1 | ENSG00000199357.1 | ENSG00000252906.1 |
| ENSG00000240167.1 | ENSG00000308272.1 | ENSG00000223573.7 | ENSG00000252010.1 |
| ENSG00000243988.1 | ENSG00000294230.1 | ENSG00000257568.1 | ENSG00000155265.12 |
| ENSG00000228414.9 | ENSG00000188959.10 | ENSG00000265442.1 | ENSG00000254484.1 |
| ENSG00000240270.1 | ENSG00000249210.1 | ENSG00000228988.1 | ENSG00000229044.2 |
| ENSG00000285603.2 | ENSG00000293890.1 | ENSG00000265097.1 | ENSG00000229120.2 |
| ENSG00000223044.1 | ENSG00000177098.9 | ENSG00000307446.1 | ENSG00000287212.1 |
| ENSG00000263934.5 | ENSG00000231672.9 | ENSG00000122386.11 | ENSG00000184678.11 |
| ENSG00000261764.1 | ENSG00000287509.1 | ENSG00000198844.12 | ENSG00000200350.1 |

| | | | |
|---|---|---|---|
| ENSG00000228238.1 | ENSG00000144481.17 | ENSG00000305185.1 | ENSG00000228412.10 |
| ENSG00000171855.7 | ENSG00000267557.2 | ENSG00000273675.1 | ENSG00000211713.3 |
| ENSG00000252929.1 | ENSG00000216813.1 | ENSG00000308589.1 | ENSG00000169418.10 |
| ENSG00000297516.1 | ENSG00000309103.1 | ENSG00000269480.1 | ENSG00000234665.10 |
| ENSG00000205644.5 | ENSG00000298931.1 | ENSG00000235868.3 | ENSG00000260851.7 |
| ENSG00000309729.1 | ENSG00000226772.1 | ENSG00000267026.6 | ENSG00000205863.12 |
| ENSG00000217791.4 | ENSG00000225657.3 | ENSG00000293032.1 | ENSG00000235964.1 |
| ENSG00000223271.1 | ENSG00000280422.1 | ENSG00000202523.1 | ENSG00000283913.2 |
| ENSG00000261055.2 | ENSG00000290730.2 | ENSG00000149257.16 | ENSG00000252164.1 |
| ENSG00000153246.13 | ENSG00000278920.1 | ENSG00000108821.14 | ENSG00000265874.1 |
| ENSG00000293334.1 | ENSG00000206645.1 | ENSG00000225455.3 | ENSG00000261442.1 |
| ENSG00000299283.1 | ENSG00000206892.1 | ENSG00000271049.1 | ENSG00000229167.3 |
| ENSG00000206781.1 | ENSG00000243260.3 | ENSG00000298591.1 | ENSG00000197099.9 |
| ENSG00000230684.1 | ENSG00000111405.9 | ENSG00000238793.1 | ENSG00000268416.1 |
| ENSG00000242971.3 | ENSG00000224743.8 | ENSG00000287201.2 | ENSG00000230889.2 |
| ENSG00000279162.1 | ENSG00000109819.10 | ENSG00000256314.1 | ENSG00000296700.1 |
| ENSG00000289085.1 | ENSG00000293153.1 | ENSG00000200897.1 | ENSG00000184545.11 |
| ENSG00000235772.1 | ENSG00000149596.7 | ENSG00000279026.1 | ENSG00000304806.1 |
| ENSG00000241560.8 | ENSG00000105550.10 | ENSG00000272170.2 | ENSG00000310549.1 |
| ENSG00000279066.1 | ENSG00000258667.5 | ENSG00000264462.1 | ENSG00000277373.1 |
| ENSG00000068137.15 | ENSG00000257663.2 | ENSG00000283599.3 | ENSG00000258285.2 |
| ENSG00000259639.7 | ENSG00000233293.2 | ENSG00000206698.1 | ENSG00000252712.1 |
| ENSG00000309670.1 | ENSG00000242510.1 | ENSG00000290095.2 | ENSG00000306165.1 |
| ENSG00000308068.1 | ENSG00000261481.2 | ENSG00000289523.1 | ENSG00000226473.2 |
| ENSG00000274064.1 | ENSG00000135925.9 | ENSG00000267587.1 | ENSG00000214886.4 |
| ENSG00000301214.1 | ENSG00000232139.7 | ENSG00000308650.1 | ENSG00000155011.9 |
| ENSG00000201793.1 | ENSG00000050165.19 | ENSG00000309083.1 | ENSG00000259786.8 |
| ENSG00000222012.2 | ENSG00000267150.2 | ENSG00000265123.2 | ENSG00000293806.1 |
| ENSG00000226483.2 | ENSG00000299492.1 | ENSG00000112379.9 | ENSG00000251018.2 |
| ENSG00000237953.1 | ENSG00000300577.1 | ENSG00000266916.10 | ENSG00000289637.2 |
| ENSG00000226645.1 | ENSG00000201563.1 | ENSG00000162105.21 | ENSG00000303420.1 |
| ENSG00000165702.15 | ENSG00000302363.1 | ENSG00000229622.1 | ENSG00000228546.2 |
| ENSG00000293760.1 | ENSG00000279174.1 | ENSG00000283045.1 | ENSG00000215372.6 |
| ENSG00000259682.1 | ENSG00000255038.2 | ENSG00000173166.18 | ENSG00000305102.1 |
| ENSG00000301183.1 | ENSG00000069188.17 | ENSG00000221230.1 | ENSG00000305084.1 |
| ENSG00000231907.2 | ENSG00000197467.17 | ENSG00000285879.1 | ENSG00000151631.8 |

| | | | |
|---|---|---|---|
| ENSG00000075891.24 | ENSG00000106688.12 | ENSG00000293728.1 | ENSG00000304717.1 |
| ENSG00000300981.1 | ENSG00000112715.26 | ENSG00000134107.5 | ENSG00000239884.3 |
| ENSG00000298548.1 | ENSG00000279713.1 | ENSG00000285627.1 | ENSG00000096696.17 |
| ENSG00000236117.2 | ENSG00000239602.1 | ENSG00000264196.1 | ENSG00000286334.1 |
| ENSG00000257494.1 | ENSG00000267457.1 | ENSG00000221493.1 | ENSG00000091138.13 |
| ENSG00000232918.2 | ENSG00000213421.4 | ENSG00000304732.1 | ENSG00000303688.1 |
| ENSG00000229087.1 | ENSG00000200241.1 | ENSG00000301184.1 | ENSG00000249593.7 |
| ENSG00000301547.1 | ENSG00000281404.2 | ENSG00000207003.1 | ENSG00000131096.11 |
| ENSG00000250330.1 | ENSG00000167995.17 | ENSG00000305566.1 | ENSG00000222607.1 |
| ENSG00000251330.3 | ENSG00000134398.15 | ENSG00000295830.1 | ENSG00000280216.1 |
| ENSG00000304237.1 | ENSG00000294192.1 | ENSG00000230137.1 | ENSG00000235090.1 |
| ENSG00000308810.1 | ENSG00000271065.2 | ENSG00000205879.5 | ENSG00000227535.2 |
| ENSG00000255650.6 | ENSG00000258602.3 | ENSG00000295410.1 | ENSG00000132002.10 |
| ENSG00000307967.1 | ENSG00000309667.1 | ENSG00000300331.1 | ENSG00000249236.3 |
| ENSG00000253491.2 | ENSG00000298708.1 | ENSG00000262228.3 | ENSG00000254907.2 |
| ENSG00000299838.1 | ENSG00000230081.2 | ENSG00000278668.1 | ENSG00000227811.4 |
| ENSG00000229541.2 | ENSG00000240767.3 | ENSG00000230433.1 | ENSG00000264940.5 |
| ENSG00000260558.2 | ENSG00000304538.1 | ENSG00000207294.1 | ENSG00000263657.1 |
| ENSG00000254388.1 | ENSG00000197320.5 | ENSG00000260965.1 | ENSG00000307750.1 |
| ENSG00000285664.3 | ENSG00000136931.10 | ENSG00000231494.1 | ENSG00000078114.19 |
| ENSG00000245970.2 | ENSG00000101958.14 | ENSG00000293190.2 | ENSG00000189060.6 |
| ENSG00000250869.2 | ENSG00000086696.11 | ENSG00000226957.1 | ENSG00000200142.1 |
| ENSG00000236375.3 | ENSG00000178404.11 | ENSG00000167414.5 | ENSG00000293823.1 |
| ENSG00000206899.1 | ENSG00000156486.8 | ENSG00000252892.1 | ENSG00000295046.1 |
| ENSG00000243094.1 | ENSG00000232568.2 | ENSG00000266805.1 | ENSG00000291262.2 |
| ENSG00000307752.1 | ENSG00000285978.1 | ENSG00000270606.1 | ENSG00000305326.1 |
| ENSG00000132000.14 | ENSG00000220643.1 | ENSG00000260571.1 | ENSG00000254132.1 |
| ENSG00000275429.1 | ENSG00000264404.4 | ENSG00000229955.1 | ENSG00000296557.1 |
| ENSG00000264540.2 | ENSG00000286223.2 | ENSG00000258837.1 | ENSG00000205100.3 |
| ENSG00000233460.1 | ENSG00000206913.1 | ENSG00000248387.1 | ENSG00000267265.7 |
| ENSG00000228107.1 | ENSG00000212743.2 | ENSG00000235267.1 | ENSG00000279476.1 |
| ENSG00000285827.1 | ENSG00000202512.1 | ENSG00000228737.3 | ENSG00000233176.3 |
| ENSG00000175197.13 | ENSG00000259017.1 | ENSG00000226205.1 | ENSG00000301313.1 |
| ENSG00000294181.1 | ENSG00000196218.15 | ENSG00000226242.1 | ENSG00000294582.1 |
| ENSG00000165186.12 | ENSG00000301163.1 | ENSG00000239280.1 | ENSG00000213221.5 |
| ENSG00000128594.8 | ENSG00000261341.7 | ENSG00000299515.1 | ENSG00000309647.1 |

| | | | |
|---|---|---|---|
| ENSG00000201264.1 | ENSG00000258955.3 | ENSG00000259563.1 | ENSG00000277453.1 |
| ENSG00000244712.1 | ENSG00000251418.2 | ENSG00000306329.1 | ENSG00000219384.1 |
| ENSG00000260545.1 | ENSG00000181333.12 | ENSG00000188242.4 | ENSG00000290090.1 |
| ENSG00000165507.9 | ENSG00000300081.1 | ENSG00000222679.1 | ENSG00000238086.5 |
| ENSG00000102802.10 | ENSG00000294184.1 | ENSG00000248769.1 | ENSG00000293174.1 |
| ENSG00000216490.4 | ENSG00000225499.1 | ENSG00000261101.2 | ENSG00000228000.1 |
| ENSG00000270333.1 | ENSG00000196735.13 | ENSG00000260109.1 | ENSG00000118971.9 |
| ENSG00000179564.4 | ENSG00000270992.1 | ENSG00000259113.1 | ENSG00000242816.3 |
| ENSG00000146147.15 | ENSG00000237207.2 | ENSG00000267048.1 | ENSG00000279283.1 |
| ENSG00000186652.10 | ENSG00000224919.2 | ENSG00000009724.18 | ENSG00000272400.1 |
| ENSG00000307473.1 | ENSG00000211727.3 | ENSG00000227782.2 | ENSG00000296649.1 |
| ENSG00000227343.1 | ENSG00000248690.9 | ENSG00000237382.2 | ENSG00000230194.1 |
| ENSG00000260012.1 | ENSG00000258847.2 | ENSG00000279786.1 | ENSG00000260428.3 |
| ENSG00000248843.1 | ENSG00000265801.1 | ENSG00000257674.1 | ENSG00000287931.2 |
| ENSG00000279544.1 | ENSG00000270679.1 | ENSG00000267222.1 | ENSG00000299015.1 |
| ENSG00000248669.1 | ENSG00000207234.1 | ENSG00000288965.1 | ENSG00000213449.2 |
| ENSG00000299923.1 | ENSG00000250253.3 | ENSG00000287093.2 | ENSG00000286242.1 |
| ENSG00000304638.1 | ENSG00000284713.2 | ENSG00000280274.1 | ENSG00000298307.1 |
| ENSG00000166592.13 | ENSG00000232718.1 | ENSG00000130208.10 | ENSG00000232564.4 |
| ENSG00000277324.2 | ENSG00000293095.1 | ENSG00000237402.2 | ENSG00000294584.1 |
| ENSG00000121552.4 | ENSG00000302533.1 | ENSG00000229780.2 | ENSG00000263300.1 |
| ENSG00000278705.4 | ENSG00000227717.5 | ENSG00000233186.2 | ENSG00000239948.2 |
| ENSG00000141934.10 | ENSG00000298219.1 | ENSG00000112796.10 | ENSG00000279819.1 |
| ENSG00000287695.1 | ENSG00000233339.1 | ENSG00000233979.1 | ENSG00000202347.1 |
| ENSG00000229206.3 | ENSG00000274066.1 | ENSG00000242986.2 | ENSG00000305631.1 |
| ENSG00000281097.1 | ENSG00000230988.3 | ENSG00000303342.1 | ENSG00000272054.1 |
| ENSG00000275708.1 | ENSG00000223724.1 | ENSG00000253475.1 | ENSG00000212525.1 |
| ENSG00000299115.1 | ENSG00000296620.1 | ENSG00000243697.1 | ENSG00000250170.1 |
| ENSG00000162374.18 | ENSG00000236662.1 | ENSG00000207393.1 | ENSG00000262445.3 |
| ENSG00000230184.1 | ENSG00000255150.2 | ENSG00000177606.9 | ENSG00000095777.17 |
| ENSG00000297761.1 | ENSG00000268678.2 | ENSG00000235908.1 | ENSG00000158482.10 |
| ENSG00000242641.7 | ENSG00000207392.1 | ENSG00000202314.1 | ENSG00000298698.1 |
| ENSG00000156234.8 | ENSG00000122756.15 | ENSG00000167619.13 | ENSG00000262115.1 |
| ENSG00000307287.1 | ENSG00000286000.1 | ENSG00000187634.13 | ENSG00000244640.1 |
| ENSG00000250102.7 | ENSG00000266946.1 | ENSG00000294929.1 | ENSG00000134376.17 |
| ENSG00000309490.1 | ENSG00000287403.1 | ENSG00000302812.1 | ENSG00000235917.1 |

| | | | |
|---|---|---|---|
| ENSG00000249899.7 | ENSG00000235236.2 | ENSG00000124260.12 | ENSG00000244203.3 |
| ENSG00000251574.9 | ENSG00000228808.1 | ENSG00000286379.1 | ENSG00000300358.1 |
| ENSG00000306183.1 | ENSG00000213174.3 | ENSG00000298824.1 | ENSG00000250600.3 |
| ENSG00000236782.8 | ENSG00000234997.4 | ENSG00000218749.1 | ENSG00000183663.6 |
| ENSG00000271557.1 | ENSG00000069702.11 | ENSG00000298226.1 | ENSG00000262135.1 |
| ENSG00000295320.1 | ENSG00000294381.1 | ENSG00000212396.1 | ENSG00000232612.1 |
| ENSG00000299440.1 | ENSG00000165802.23 | ENSG00000301789.1 | ENSG00000243423.1 |
| ENSG00000263982.1 | ENSG00000260031.1 | ENSG00000289946.1 | ENSG00000308598.1 |
| ENSG00000307685.1 | ENSG00000285770.1 | ENSG00000253528.2 | ENSG00000006071.16 |
| ENSG00000232344.2 | ENSG00000294729.1 | ENSG00000280150.1 | ENSG00000241961.1 |
| ENSG00000199562.1 | ENSG00000252363.1 | ENSG00000259429.6 | ENSG00000302863.1 |
| ENSG00000208308.1 | ENSG00000293202.2 | ENSG00000301787.1 | ENSG00000234724.1 |
| ENSG00000297355.1 | ENSG00000297002.1 | ENSG00000280138.1 | ENSG00000273555.1 |
| ENSG00000264273.1 | ENSG00000266965.1 | ENSG00000109107.14 | ENSG00000254884.1 |
| ENSG00000259192.1 | ENSG00000237413.6 | ENSG00000261270.1 | ENSG00000234019.1 |
| ENSG00000287643.1 | ENSG00000214067.2 | ENSG00000267592.1 | ENSG00000105146.14 |
| ENSG00000249515.2 | ENSG00000207297.1 | ENSG00000255353.1 | ENSG00000238061.2 |
| ENSG00000297241.1 | ENSG00000100336.18 | ENSG00000203262.3 | ENSG00000246422.2 |
| ENSG00000299573.1 | ENSG00000259635.1 | ENSG00000101951.18 | ENSG00000283204.1 |
| ENSG00000299926.1 | ENSG00000230262.10 | ENSG00000256603.1 | ENSG00000170950.6 |
| ENSG00000302084.1 | ENSG00000235602.5 | ENSG00000228930.1 | ENSG00000288994.1 |
| ENSG00000280109.4 | ENSG00000295624.1 | ENSG00000300189.1 | ENSG00000290792.1 |
| ENSG00000130513.7 | ENSG00000293786.1 | ENSG00000304102.1 | ENSG00000301211.1 |
| ENSG00000236889.1 | ENSG00000256987.1 | ENSG00000265625.1 | ENSG00000226443.3 |
| ENSG00000307662.1 | ENSG00000239490.1 | ENSG00000204657.4 | ENSG00000295251.1 |
| ENSG00000261634.5 | ENSG00000293245.2 | ENSG00000252821.1 | ENSG00000224967.1 |
| ENSG00000257087.1 | ENSG00000225063.1 | ENSG00000202279.1 | ENSG00000290738.2 |
| ENSG00000238297.2 | ENSG00000163915.7 | ENSG00000240174.1 | ENSG00000270739.1 |
| ENSG00000295872.1 | ENSG00000237705.1 | ENSG00000298419.1 | ENSG00000253218.1 |
| ENSG00000255843.2 | ENSG00000266776.1 | ENSG00000302821.1 | ENSG00000302031.1 |
| ENSG00000237249.1 | ENSG00000206784.1 | ENSG00000290559.2 | ENSG00000224331.2 |
| ENSG00000302290.1 | ENSG00000151778.11 | ENSG00000255440.3 | ENSG00000225343.1 |
| ENSG00000271052.1 | ENSG00000279764.1 | ENSG00000256361.1 | ENSG00000299414.1 |
| ENSG00000217896.2 | ENSG00000299802.1 | ENSG00000255973.1 | ENSG00000237166.5 |
| ENSG00000080824.19 | ENSG00000278917.1 | ENSG00000146005.4 | ENSG00000293504.2 |
| ENSG00000258951.1 | ENSG00000226985.8 | ENSG00000254909.1 | ENSG00000232170.7 |

| | | | |
|---|---|---|---|
| ENSG00000253807.7 | ENSG00000286214.1 | ENSG00000226144.2 | ENSG00000273096.1 |
| ENSG00000288032.1 | ENSG00000231562.1 | ENSG00000177359.23 | ENSG00000290590.2 |
| ENSG00000234478.2 | ENSG00000295839.1 | ENSG00000239527.1 | ENSG00000265462.1 |
| ENSG00000186472.20 | ENSG00000309075.1 | ENSG00000283503.5 | ENSG00000306343.1 |
| ENSG00000198561.16 | ENSG00000308538.1 | ENSG00000200534.1 | ENSG00000287437.1 |
| ENSG00000295348.1 | ENSG00000237049.1 | ENSG00000275491.2 | ENSG00000269172.1 |
| ENSG00000278963.1 | ENSG00000138152.10 | ENSG00000264102.1 | ENSG00000266936.1 |
| ENSG00000276727.2 | ENSG00000262961.1 | ENSG00000308255.1 | ENSG00000235029.1 |
| ENSG00000285938.2 | ENSG00000273687.1 | ENSG00000213046.4 | ENSG00000241088.2 |
| ENSG00000250069.1 | ENSG00000207187.1 | ENSG00000254791.1 | ENSG00000259083.2 |
| ENSG00000294500.1 | ENSG00000306879.1 | ENSG00000200502.1 | ENSG00000239503.1 |
| ENSG00000290581.1 | ENSG00000225349.2 | ENSG00000259713.1 | ENSG00000213495.3 |
| ENSG00000239253.1 | ENSG00000277268.3 | ENSG00000275339.1 | ENSG00000175356.14 |
| ENSG00000296260.1 | ENSG00000104059.5 | ENSG00000297826.1 | ENSG00000229502.5 |
| ENSG00000274427.1 | ENSG00000224678.3 | ENSG00000275110.1 | ENSG00000298426.1 |
| ENSG00000306806.1 | ENSG00000242849.2 | ENSG00000277586.4 | ENSG00000233737.3 |
| ENSG00000255081.1 | ENSG00000249318.1 | ENSG00000176349.12 | ENSG00000188365.4 |
| ENSG00000279474.1 | ENSG00000254554.2 | ENSG00000270846.1 | ENSG00000287657.2 |
| ENSG00000236476.2 | ENSG00000230138.2 | ENSG00000285366.1 | ENSG00000284484.1 |
| ENSG00000252122.1 | ENSG00000241654.1 | ENSG00000244756.1 | ENSG00000198729.5 |
| ENSG00000287451.2 | ENSG00000290409.2 | ENSG00000288915.2 | ENSG00000303850.1 |
| ENSG00000253395.2 | ENSG00000214860.5 | ENSG00000243655.2 | ENSG00000230171.1 |
| ENSG00000236576.1 | ENSG00000202031.1 | ENSG00000143847.16 | ENSG00000167178.17 |
| ENSG00000310198.1 | ENSG00000218682.1 | ENSG00000294665.1 | ENSG00000102001.13 |
| ENSG00000222078.1 | ENSG00000170075.10 | ENSG00000306369.1 | ENSG00000234883.8 |
| ENSG00000300446.1 | ENSG00000285611.3 | ENSG00000238390.1 | ENSG00000231586.2 |
| ENSG00000300685.1 | ENSG00000256762.1 | ENSG00000225578.1 | ENSG00000251794.1 |
| ENSG00000249786.8 | ENSG00000309534.1 | ENSG00000308148.1 | ENSG00000273035.1 |
| ENSG00000309364.1 | ENSG00000274591.1 | ENSG00000214657.4 | ENSG00000217776.1 |
| ENSG00000272402.1 | ENSG00000229623.1 | ENSG00000137962.13 | ENSG00000301334.1 |
| ENSG00000163624.6 | ENSG00000169067.4 | ENSG00000223824.1 | ENSG00000252824.1 |
| ENSG00000233454.1 | ENSG00000236132.1 | ENSG00000305788.1 | ENSG00000118515.12 |
| ENSG00000214604.3 | ENSG00000263343.1 | ENSG00000205307.12 | ENSG00000225721.5 |
| ENSG00000279957.1 | ENSG00000285658.1 | ENSG00000228804.6 | ENSG00000267016.1 |
| ENSG00000212607.1 | ENSG00000309113.1 | ENSG00000124134.9 | ENSG00000214142.2 |
| ENSG00000260799.1 | ENSG00000224085.3 | ENSG00000235440.1 | ENSG00000170236.16 |

| | | | |
|---|---|---|---|
| ENSG00000181631.7 | ENSG00000104888.10 | ENSG00000082397.18 | ENSG00000228992.2 |
| ENSG00000164684.13 | ENSG00000265706.1 | ENSG00000141448.11 | ENSG00000214563.2 |
| ENSG00000273041.1 | ENSG00000266100.1 | ENSG00000266538.1 | ENSG00000206786.1 |
| ENSG00000160282.15 | ENSG00000197405.8 | ENSG00000215278.4 | ENSG00000221216.1 |
| ENSG00000305699.1 | ENSG00000201519.1 | ENSG00000254091.2 | ENSG00000223646.3 |
| ENSG00000229891.5 | ENSG00000105143.13 | ENSG00000299196.1 | ENSG00000280387.1 |
| ENSG00000298809.1 | ENSG00000305753.1 | ENSG00000213917.2 | ENSG00000237851.1 |
| ENSG00000274215.1 | ENSG00000285704.1 | ENSG00000219253.1 | ENSG00000278385.1 |
| ENSG00000277959.1 | ENSG00000080572.14 | ENSG00000219758.6 | ENSG00000273855.1 |
| ENSG00000294063.1 | ENSG00000296222.1 | ENSG00000163534.16 | ENSG00000231112.1 |
| ENSG00000248979.2 | ENSG00000289141.3 | ENSG00000302225.1 | ENSG00000287838.2 |
| ENSG00000297034.1 | ENSG00000228433.2 | ENSG00000303710.1 | ENSG00000270104.1 |
| ENSG00000289019.2 | ENSG00000298966.1 | ENSG00000135248.16 | ENSG00000273706.5 |
| ENSG00000099840.14 | ENSG00000307064.1 | ENSG00000255115.2 | ENSG00000219712.1 |
| ENSG00000224121.1 | ENSG00000290164.1 | ENSG00000118785.15 | ENSG00000164683.19 |
| ENSG00000219433.2 | ENSG00000293608.1 | ENSG00000101842.14 | ENSG00000304541.1 |
| ENSG00000234995.1 | ENSG00000302705.1 | ENSG00000295491.1 | ENSG00000230734.1 |
| ENSG00000309068.1 | ENSG00000274414.1 | ENSG00000261832.6 | ENSG00000157856.12 |
| ENSG00000270832.1 | ENSG00000273245.1 | ENSG00000139278.10 | ENSG00000273925.1 |
| ENSG00000273353.1 | ENSG00000273196.1 | ENSG00000206077.14 | ENSG00000259873.1 |
| ENSG00000125895.6 | ENSG00000239207.2 | ENSG00000295309.1 | ENSG00000243095.1 |
| ENSG00000295242.1 | ENSG00000277224.3 | ENSG00000261369.2 | ENSG00000269246.3 |
| ENSG00000302521.1 | ENSG00000299367.1 | ENSG00000305059.1 | ENSG00000258618.2 |
| ENSG00000304007.1 | ENSG00000229190.2 | ENSG00000235328.1 | ENSG00000252060.1 |
| ENSG00000293142.1 | ENSG00000233933.2 | ENSG00000178596.10 | ENSG00000270240.3 |
| ENSG00000254338.3 | ENSG00000264044.2 | ENSG00000235656.1 | ENSG00000125084.13 |
| ENSG00000239861.1 | ENSG00000284669.2 | ENSG00000202358.1 | ENSG00000301123.1 |
| ENSG00000298772.1 | ENSG00000242995.1 | ENSG00000230799.1 | ENSG00000243927.7 |
| ENSG00000226455.1 | ENSG00000228086.2 | ENSG00000305868.1 | ENSG00000229326.3 |
| ENSG00000308725.1 | ENSG00000102239.5 | ENSG00000254577.1 | ENSG00000210077.1 |
| ENSG00000263624.1 | ENSG00000155659.15 | ENSG00000255229.2 | ENSG00000228363.3 |
| ENSG00000254631.5 | ENSG00000284431.1 | ENSG00000293264.2 | ENSG00000224511.3 |
| ENSG00000183018.9 | ENSG00000286588.1 | ENSG00000213744.3 | ENSG00000295065.1 |
| ENSG00000134460.19 | ENSG00000230063.1 | ENSG00000309946.1 | ENSG00000237971.1 |
| ENSG00000272870.5 | ENSG00000197980.14 | ENSG00000273076.1 | ENSG00000100346.18 |
| ENSG00000229990.3 | ENSG00000229800.1 | ENSG00000270894.1 | ENSG00000301612.1 |

| | | | |
|---|---|---|---|
| ENSG00000275811.1 | ENSG00000279057.1 | ENSG00000229169.1 | ENSG00000230355.1 |
| ENSG00000295575.1 | ENSG00000229111.2 | ENSG00000215218.4 | ENSG00000273365.1 |
| ENSG00000272975.3 | ENSG00000286314.1 | ENSG00000285605.1 | ENSG00000260082.1 |
| ENSG00000235685.2 | ENSG00000249840.2 | ENSG00000261838.6 | ENSG00000229658.1 |
| ENSG00000250787.1 | ENSG00000228681.1 | ENSG00000301682.1 | ENSG00000142611.17 |
| ENSG00000250045.2 | ENSG00000265883.1 | ENSG00000230470.1 | ENSG00000283199.3 |
| ENSG00000307020.1 | ENSG00000290119.2 | ENSG00000107593.17 | ENSG00000095627.10 |
| ENSG00000254730.1 | ENSG00000289564.2 | ENSG00000304967.1 | ENSG00000202538.2 |
| ENSG00000279595.1 | ENSG00000117643.16 | ENSG00000235700.1 | ENSG00000234147.5 |
| ENSG00000302877.1 | ENSG00000300101.1 | ENSG00000290830.1 | ENSG00000227082.4 |
| ENSG00000226356.2 | ENSG00000234709.2 | ENSG00000264546.1 | ENSG00000237101.2 |
| ENSG00000187772.8 | ENSG00000226636.1 | ENSG00000278828.2 | ENSG00000172209.6 |
| ENSG00000166743.10 | ENSG00000296617.1 | ENSG00000231434.1 | ENSG00000115884.11 |
| ENSG00000294744.1 | ENSG00000277007.1 | ENSG00000286688.1 | ENSG00000128917.8 |
| ENSG00000260277.3 | ENSG00000234943.2 | ENSG00000303101.1 | ENSG00000225308.2 |
| ENSG00000232970.1 | ENSG00000234586.2 | ENSG00000298979.1 | ENSG00000231017.1 |
| ENSG00000223138.1 | ENSG00000273976.2 | ENSG00000279161.1 | ENSG00000266989.1 |
| ENSG00000134548.11 | ENSG00000133138.20 | ENSG00000212443.1 | ENSG00000246695.10 |
| ENSG00000230789.2 | ENSG00000276925.1 | ENSG00000242094.1 | ENSG00000298118.1 |
| ENSG00000270760.1 | ENSG00000290040.1 | ENSG00000308189.1 | ENSG00000223884.8 |
| ENSG00000296036.1 | ENSG00000229814.1 | ENSG00000275954.6 | ENSG00000288235.1 |
| ENSG00000308033.1 | ENSG00000261072.2 | ENSG00000303049.1 | ENSG00000240459.2 |
| ENSG00000199424.1 | ENSG00000171189.18 | ENSG00000053438.11 | ENSG00000269534.5 |
| ENSG00000128253.16 | ENSG00000307218.1 | ENSG00000239412.1 | ENSG00000252206.1 |
| ENSG00000101850.13 | ENSG00000164949.8 | ENSG00000299648.1 | ENSG00000232878.3 |
| ENSG00000282977.1 | ENSG00000181218.6 | ENSG00000281780.1 | ENSG00000238311.1 |
| ENSG00000230417.13 | ENSG00000252427.1 | ENSG00000252096.1 | ENSG00000301425.1 |
| ENSG00000143878.10 | ENSG00000102452.18 | ENSG00000228016.2 | ENSG00000115738.10 |
| ENSG00000237409.2 | ENSG00000153234.16 | ENSG00000285881.2 | ENSG00000229570.2 |
| ENSG00000305746.1 | ENSG00000238317.2 | ENSG00000226647.2 | ENSG00000279416.1 |
| ENSG00000213594.4 | ENSG00000244411.4 | ENSG00000239291.1 | ENSG00000292328.1 |
| ENSG00000287665.1 | ENSG00000217718.1 | ENSG00000235833.1 | ENSG00000213338.3 |
| ENSG00000264853.1 | ENSG00000253107.1 | ENSG00000276550.6 | ENSG00000271714.1 |
| ENSG00000260618.1 | ENSG00000303467.1 | ENSG00000227999.1 | ENSG00000111341.10 |
| ENSG00000230188.1 | ENSG00000226801.2 | ENSG00000240881.1 | ENSG00000274231.1 |
| ENSG00000259712.1 | ENSG00000288545.2 | ENSG00000285970.1 | ENSG00000224746.3 |

| | | | |
|---|---|---|---|
| ENSG00000286302.1 | ENSG00000123358.20 | ENSG00000164692.19 | ENSG00000296794.1 |
| ENSG00000237432.2 | ENSG00000277763.2 | ENSG00000296798.1 | ENSG00000217527.1 |
| ENSG00000237329.2 | ENSG00000236811.1 | ENSG00000020577.14 | ENSG00000277172.1 |
| ENSG00000256056.1 | ENSG00000238180.1 | ENSG00000287965.1 | ENSG00000236299.9 |
| ENSG00000309772.1 | ENSG00000224855.5 | ENSG00000260436.1 | ENSG00000236234.1 |
| ENSG00000227573.2 | ENSG00000277144.1 | ENSG00000253273.2 | ENSG00000275215.1 |
| ENSG00000245080.8 | ENSG00000240808.1 | ENSG00000199638.1 | ENSG00000242041.1 |
| ENSG00000168874.14 | ENSG00000244740.1 | ENSG00000200059.1 | ENSG00000289727.1 |
| ENSG00000259398.1 | ENSG00000270165.1 | ENSG00000271697.1 | ENSG00000259103.3 |
| ENSG00000256079.1 | ENSG00000223460.1 | ENSG00000294514.1 | ENSG00000260808.1 |
| ENSG00000251869.1 | ENSG00000225840.2 | ENSG00000238069.1 | ENSG00000233133.1 |
| ENSG00000237109.1 | ENSG00000228825.1 | ENSG00000232627.1 | ENSG00000120075.6 |
| ENSG00000258640.3 | ENSG00000295987.1 | ENSG00000091831.25 | ENSG00000134363.12 |
| ENSG00000224987.1 | ENSG00000302586.1 | ENSG00000287422.1 | ENSG00000074964.17 |
| ENSG00000232455.3 | ENSG00000271400.1 | ENSG00000295218.1 | ENSG00000256950.2 |
| ENSG00000295861.1 | ENSG00000258546.1 | ENSG00000184111.6 | ENSG00000259109.1 |
| ENSG00000165300.8 | ENSG00000243954.3 | ENSG00000229743.7 | ENSG00000265399.1 |
| ENSG00000247372.3 | ENSG00000287532.1 | ENSG00000229230.3 | ENSG00000307703.1 |
| ENSG00000230284.1 | ENSG00000225442.2 | ENSG00000294022.1 | ENSG00000272483.1 |
| ENSG00000110092.4 | ENSG00000241739.1 | ENSG00000235323.2 | ENSG00000303800.1 |
| ENSG00000267734.1 | ENSG00000213376.4 | ENSG00000267508.6 | ENSG00000295471.1 |
| ENSG00000103313.14 | ENSG00000267255.1 | ENSG00000293500.2 | ENSG00000308625.1 |
| ENSG00000241738.1 | ENSG00000300322.1 | ENSG00000236320.4 | ENSG00000198049.7 |
| ENSG00000277358.1 | ENSG00000184384.15 | ENSG00000271215.1 | ENSG00000249270.1 |
| ENSG00000079156.17 | ENSG00000242522.2 | ENSG00000306181.1 | ENSG00000288658.2 |
| ENSG00000215704.10 | ENSG00000213361.2 | ENSG00000273271.1 | ENSG00000283526.3 |
| ENSG00000280078.1 | ENSG00000235674.2 | ENSG00000262265.2 | ENSG00000268469.1 |
| ENSG00000233231.1 | ENSG00000241535.1 | ENSG00000233924.1 | ENSG00000309574.1 |
| ENSG00000294185.1 | ENSG00000280376.1 | ENSG00000162783.11 | ENSG00000254734.1 |
| ENSG00000278979.1 | ENSG00000273433.1 | ENSG00000289355.3 | ENSG00000196408.12 |
| ENSG00000232998.4 | ENSG00000229312.3 | ENSG00000258479.7 | ENSG00000297330.1 |
| ENSG00000252213.1 | ENSG00000305936.1 | ENSG00000239268.6 | ENSG00000243609.1 |
| ENSG00000307603.1 | ENSG00000309778.1 | ENSG00000231439.5 | ENSG00000289518.2 |
| ENSG00000224308.1 | ENSG00000231760.5 | ENSG00000234136.1 | ENSG00000159208.16 |
| ENSG00000263740.2 | ENSG00000218754.4 | ENSG00000158050.6 | ENSG00000303728.1 |
| ENSG00000266190.1 | ENSG00000152086.9 | ENSG00000277597.3 | ENSG00000287644.2 |

| | | | |
|---|---|---|---|
| ENSG00000243193.5 | ENSG00000282100.1 | ENSG00000297284.1 | ENSG00000255850.3 |
| ENSG00000109424.4 | ENSG00000279236.1 | ENSG00000237540.1 | ENSG00000073792.16 |
| ENSG00000211899.10 | ENSG00000259783.7 | ENSG00000258725.2 | ENSG00000287789.1 |
| ENSG00000288859.2 | ENSG00000198435.4 | ENSG00000214457.3 | ENSG00000199875.1 |
| ENSG00000226338.1 | ENSG00000258552.1 | ENSG00000295152.1 | ENSG00000217227.1 |
| ENSG00000257752.1 | ENSG00000272034.1 | ENSG00000276756.5 | ENSG00000302420.1 |
| ENSG00000232611.1 | ENSG00000305684.1 | ENSG00000215186.7 | ENSG00000224789.2 |
| ENSG00000242978.1 | ENSG00000240219.1 | ENSG00000223968.1 | ENSG00000231871.5 |
| ENSG00000132554.21 | ENSG00000283409.1 | ENSG00000278517.1 | ENSG00000169884.14 |
| ENSG00000299387.1 | ENSG00000228019.1 | ENSG00000291041.2 | ENSG00000250362.2 |
| ENSG00000211707.3 | ENSG00000114854.8 | ENSG00000234735.1 | ENSG00000268105.1 |
| ENSG00000301008.1 | ENSG00000287724.1 | ENSG00000267203.1 | ENSG00000200397.1 |
| ENSG00000251308.1 | ENSG00000285942.1 | ENSG00000158458.21 | ENSG00000286458.2 |
| ENSG00000279862.1 | ENSG00000304761.1 | ENSG00000220131.1 | ENSG00000200913.1 |
| ENSG00000280809.6 | ENSG00000261588.1 | ENSG00000212163.6 | ENSG00000225420.2 |
| ENSG00000275409.1 | ENSG00000286364.1 | ENSG00000305875.1 | ENSG00000254968.9 |
| ENSG00000303097.1 | ENSG00000225284.1 | ENSG00000300742.1 | ENSG00000267207.2 |
| ENSG00000145623.13 | ENSG00000167195.8 | ENSG00000267056.2 | ENSG00000206846.1 |
| ENSG00000278637.3 | ENSG00000226026.8 | ENSG00000218537.1 | ENSG00000226540.2 |
| ENSG00000269540.1 | ENSG00000249930.1 | ENSG00000218313.1 | ENSG00000236977.2 |
| ENSG00000280402.1 | ENSG00000279766.1 | ENSG00000295464.1 | ENSG00000236194.3 |
| ENSG00000235748.1 | ENSG00000158445.10 | ENSG00000279281.1 | ENSG00000289380.1 |
| ENSG00000263011.4 | ENSG00000259135.2 | ENSG00000244113.1 | ENSG00000188000.6 |
| ENSG00000261813.2 | ENSG00000273448.1 | ENSG00000225400.1 | ENSG00000293235.1 |
| ENSG00000293150.1 | ENSG00000286478.2 | ENSG00000141314.13 | ENSG00000285948.2 |
| ENSG00000288839.1 | ENSG00000281469.1 | ENSG00000272663.2 | ENSG00000259045.1 |
| ENSG00000259121.2 | ENSG00000232486.1 | ENSG00000237080.3 | ENSG00000290879.2 |
| ENSG00000184221.13 | ENSG00000242527.1 | ENSG00000261872.1 | ENSG00000175820.4 |
| ENSG00000279562.1 | ENSG00000308164.1 | ENSG00000182379.10 | ENSG00000149564.12 |
| ENSG00000222588.1 | ENSG00000259535.1 | ENSG00000200075.1 | ENSG00000270083.1 |
| ENSG00000270799.1 | ENSG00000172638.13 | ENSG00000250295.7 | ENSG00000248954.2 |
| ENSG00000273554.4 | ENSG00000174562.14 | ENSG00000274383.1 | ENSG00000310354.1 |
| ENSG00000215498.10 | ENSG00000294331.1 | ENSG00000228634.1 | ENSG00000256632.3 |
| ENSG00000238365.3 | ENSG00000272320.1 | ENSG00000249488.1 | ENSG00000294875.1 |
| ENSG00000249963.1 | ENSG00000224476.3 | ENSG00000310286.1 | ENSG00000252620.1 |
| ENSG00000007174.18 | ENSG00000298795.1 | ENSG00000243225.2 | ENSG00000289014.2 |

| | | | |
|---|---|---|---|
| ENSG00000223901.2 | ENSG00000242853.3 | ENSG00000287767.2 | ENSG00000279568.1 |
| ENSG00000213403.2 | ENSG00000200922.1 | ENSG00000301589.1 | ENSG00000279554.1 |
| ENSG00000245748.2 | ENSG00000279433.1 | ENSG00000310508.1 | ENSG00000286071.4 |
| ENSG00000214988.4 | ENSG00000118903.6 | ENSG00000187600.15 | ENSG00000277759.1 |
| ENSG00000226072.1 | ENSG00000303913.1 | ENSG00000260190.1 | ENSG00000256325.1 |
| ENSG00000128242.13 | ENSG00000243537.1 | ENSG00000307831.1 | ENSG00000230162.1 |
| ENSG00000183722.9 | ENSG00000295844.1 | ENSG00000224519.1 | ENSG00000289569.2 |
| ENSG00000294853.1 | ENSG00000303737.1 | ENSG00000213302.3 | ENSG00000275759.1 |
| ENSG00000302700.1 | ENSG00000287917.4 | ENSG00000207336.1 | ENSG00000228108.1 |
| ENSG00000130561.17 | ENSG00000253829.1 | ENSG00000179580.10 | ENSG00000272812.1 |
| ENSG00000176046.9 | ENSG00000230630.6 | ENSG00000236191.3 | ENSG00000240083.2 |
| ENSG00000231340.1 | ENSG00000296924.1 | ENSG00000171246.7 | ENSG00000243761.1 |
| ENSG00000273165.1 | ENSG00000296756.1 | ENSG00000253708.1 | ENSG00000198526.7 |
| ENSG00000299857.1 | ENSG00000271933.1 | ENSG00000235051.3 | ENSG00000243455.3 |
| ENSG00000242431.2 | ENSG00000231139.1 | ENSG00000232142.1 | ENSG00000287319.1 |
| ENSG00000224655.8 | ENSG00000232378.1 | ENSG00000300555.1 | ENSG00000280269.1 |
| ENSG00000216902.3 | ENSG00000302265.1 | ENSG00000225024.1 | ENSG00000268204.1 |
| ENSG00000271687.1 | ENSG00000229423.2 | ENSG00000266980.1 | ENSG00000267127.7 |
| ENSG00000135678.12 | ENSG00000239040.1 | ENSG00000287216.1 | ENSG00000213574.2 |
| ENSG00000180525.15 | ENSG00000263603.1 | ENSG00000240093.2 | ENSG00000276393.1 |
| ENSG00000234119.1 | ENSG00000239256.1 | ENSG00000231072.1 | ENSG00000263311.1 |
| ENSG00000242829.1 | ENSG00000301099.1 | ENSG00000125657.5 | ENSG00000271980.1 |
| ENSG00000272767.2 | ENSG00000243705.1 | ENSG00000285976.4 | ENSG00000101115.13 |
| ENSG00000293981.1 | ENSG00000279320.1 | ENSG00000274422.1 | ENSG00000235583.2 |
| ENSG00000231956.1 | ENSG00000234429.1 | ENSG00000302819.1 | ENSG00000237924.1 |
| ENSG00000238259.1 | ENSG00000269897.5 | ENSG00000168298.7 | ENSG00000233565.1 |
| ENSG00000214280.3 | ENSG00000244242.2 | ENSG00000239524.2 | ENSG00000267551.3 |
| ENSG00000279570.1 | ENSG00000255838.1 | ENSG00000236844.1 | ENSG00000178750.3 |
| ENSG00000288863.1 | ENSG00000228262.12 | ENSG00000247095.6 | ENSG00000295071.1 |
| ENSG00000255959.2 | ENSG00000253237.2 | ENSG00000274678.1 | ENSG00000073598.6 |
| ENSG00000289506.3 | ENSG00000101311.17 | ENSG00000277290.1 | ENSG00000184897.7 |
| ENSG00000307512.1 | ENSG00000242999.3 | ENSG00000231501.1 | ENSG00000125740.15 |
| ENSG00000131951.12 | ENSG00000211513.5 | ENSG00000309790.1 | ENSG00000226963.1 |
| ENSG00000295813.1 | ENSG00000272905.1 | ENSG00000304580.1 | ENSG00000262652.1 |
| ENSG00000233334.4 | ENSG00000214326.2 | ENSG00000235892.1 | ENSG00000219135.1 |
| ENSG00000267463.1 | ENSG00000180221.6 | ENSG00000244717.1 | ENSG00000127955.17 |

| | | | |
|---|---|---|---|
| ENSG00000244267.1 | ENSG00000242252.2 | ENSG00000214193.12 | ENSG00000290070.1 |
| ENSG00000233947.1 | ENSG00000250027.2 | ENSG00000213448.3 | ENSG00000235995.1 |
| ENSG00000169253.3 | ENSG00000228415.3 | ENSG00000115457.10 | ENSG00000226903.4 |
| ENSG00000293050.1 | ENSG00000219951.4 | ENSG00000176343.5 | ENSG00000253973.4 |
| ENSG00000251544.1 | ENSG00000164142.16 | ENSG00000234585.7 | ENSG00000259683.1 |
| ENSG00000285815.1 | ENSG00000249047.2 | ENSG00000260597.1 | ENSG00000144406.20 |
| ENSG00000272568.7 | ENSG00000215002.2 | ENSG00000310216.1 | ENSG00000216316.1 |
| ENSG00000258545.8 | ENSG00000283627.2 | ENSG00000241409.1 | ENSG00000306538.1 |
| ENSG00000299814.1 | ENSG00000223550.1 | ENSG00000242737.1 | ENSG00000302492.1 |
| ENSG00000216938.3 | ENSG00000257366.1 | ENSG00000280401.1 | ENSG00000293689.1 |
| ENSG00000232043.1 | ENSG00000261291.1 | ENSG00000309692.1 | ENSG00000188483.8 |
| ENSG00000309943.1 | ENSG00000243129.2 | ENSG00000259982.1 | ENSG00000140398.14 |
| ENSG00000217178.1 | ENSG00000274933.6 | ENSG00000250251.7 | ENSG00000279502.1 |
| ENSG00000241112.1 | ENSG00000225300.1 | ENSG00000189167.12 | ENSG00000064989.13 |
| ENSG00000250075.8 | ENSG00000259228.1 | ENSG00000259130.3 | ENSG00000168589.15 |
| ENSG00000228754.1 | ENSG00000227813.1 | ENSG00000256358.1 | ENSG00000273356.1 |
| ENSG00000224315.2 | ENSG00000224598.1 | ENSG00000106483.12 | ENSG00000223640.1 |
| ENSG00000161509.14 | ENSG00000202222.1 | ENSG00000276368.2 | ENSG00000231307.1 |
| ENSG00000224490.6 | ENSG00000259201.1 | ENSG00000236189.2 | ENSG00000297168.1 |
| ENSG00000259915.2 | ENSG00000154898.17 | ENSG00000230203.1 | ENSG00000162746.15 |
| ENSG00000258454.2 | ENSG00000174945.14 | ENSG00000244048.1 | ENSG00000228365.2 |
| ENSG00000286307.1 | ENSG00000291185.2 | ENSG00000240435.2 | ENSG00000276517.1 |
| ENSG00000252739.1 | ENSG00000277634.1 | ENSG00000213880.3 | ENSG00000069399.15 |
| ENSG00000223692.1 | ENSG00000296823.1 | ENSG00000254463.1 | ENSG00000101160.15 |
| ENSG00000206976.1 | ENSG00000218416.5 | ENSG00000240513.1 | ENSG00000240311.1 |
| ENSG00000307963.1 | ENSG00000304647.1 | ENSG00000291194.2 | ENSG00000196747.5 |
| ENSG00000220305.1 | ENSG00000247809.12 | ENSG00000288888.1 | ENSG00000310164.1 |
| ENSG00000246130.2 | ENSG00000144619.15 | ENSG00000165181.17 | ENSG00000225778.6 |
| ENSG00000303445.1 | ENSG00000124092.13 | ENSG00000303940.1 | ENSG00000204396.11 |
| ENSG00000214259.3 | ENSG00000224055.1 | ENSG00000241213.3 | ENSG00000215236.3 |
| ENSG00000252614.1 | ENSG00000119771.15 | ENSG00000290791.2 | ENSG00000267526.1 |
| ENSG00000307379.1 | ENSG00000230084.9 | ENSG00000182168.16 | ENSG00000231525.1 |
| ENSG00000202058.1 | ENSG00000242814.2 | ENSG00000223969.8 | ENSG00000217680.1 |
| ENSG00000139549.4 | ENSG00000257119.1 | ENSG00000229753.1 | ENSG00000276868.1 |
| ENSG00000288080.2 | ENSG00000223429.1 | ENSG00000196967.11 | ENSG00000248885.1 |
| ENSG00000201121.1 | ENSG00000260816.2 | ENSG00000169495.5 | ENSG00000310327.1 |

| | | | |
|---|---|---|---|
| ENSG00000279428.1 | ENSG00000220557.1 | ENSG00000210049.1 | ENSG00000223505.2 |
| ENSG00000286672.1 | ENSG00000272810.1 | ENSG00000229168.4 | ENSG00000224464.2 |
| ENSG00000176171.12 | ENSG00000284703.2 | ENSG00000230249.1 | ENSG00000242706.2 |
| ENSG00000214297.3 | ENSG00000294216.1 | ENSG00000283269.2 | ENSG00000232015.2 |
| ENSG00000154310.17 | ENSG00000255198.7 | ENSG00000233876.2 | ENSG00000215264.4 |
| ENSG00000262848.1 | ENSG00000242411.2 | ENSG00000265093.2 | ENSG00000227227.1 |
| ENSG00000277043.1 | ENSG00000229663.1 | ENSG00000256712.1 | ENSG00000307798.1 |
| ENSG00000235232.1 | ENSG00000156711.17 | ENSG00000207445.1 | ENSG00000230572.6 |
| ENSG00000230753.7 | ENSG00000234272.1 | ENSG00000143603.19 | ENSG00000220614.1 |
| ENSG00000232848.1 | ENSG00000297004.1 | ENSG00000229272.3 | ENSG00000280381.1 |
| ENSG00000114268.12 | ENSG00000279700.1 | ENSG00000246582.4 | ENSG00000229985.2 |
| ENSG00000124194.17 | ENSG00000302820.1 | ENSG00000288795.2 | ENSG00000271605.7 |
| ENSG00000236489.1 | ENSG00000238221.2 | ENSG00000224003.1 | ENSG00000168280.18 |
| ENSG00000290103.2 | ENSG00000294902.1 | ENSG00000249489.1 | ENSG00000254653.1 |
| ENSG00000288025.1 | ENSG00000231329.8 | ENSG00000255409.1 | ENSG00000121905.10 |
| ENSG00000225639.1 | ENSG00000226545.1 | ENSG00000287450.1 | ENSG00000266824.2 |
| ENSG00000234265.3 | ENSG00000183801.8 | ENSG00000253621.1 | ENSG00000259105.1 |
| ENSG00000236567.2 | ENSG00000297713.1 | ENSG00000243188.1 | ENSG00000215357.2 |
| ENSG00000246379.10 | ENSG00000175518.8 | ENSG00000158014.15 | ENSG00000258673.3 |
| ENSG00000203811.2 | ENSG00000214374.2 | ENSG00000171119.3 | ENSG00000300847.1 |
| ENSG00000300920.1 | ENSG00000256093.1 | ENSG00000180720.8 | ENSG00000271010.1 |
| ENSG00000251637.7 | ENSG00000215325.4 | ENSG00000301061.1 | ENSG00000231560.1 |
| ENSG00000268564.2 | ENSG00000264204.2 | ENSG00000259416.3 | ENSG00000234590.1 |
| ENSG00000258469.2 | ENSG00000278967.1 | ENSG00000143595.13 | ENSG00000295591.1 |
| ENSG00000287814.2 | ENSG00000242088.1 | ENSG00000239516.1 | ENSG00000259659.1 |
| ENSG00000253332.1 | ENSG00000112309.11 | ENSG00000255663.1 | ENSG00000289173.3 |
| ENSG00000288603.1 | ENSG00000287193.1 | ENSG00000277327.1 | ENSG00000278299.6 |
| ENSG00000286245.1 | ENSG00000215559.9 | ENSG00000246851.1 | ENSG00000232199.1 |
| ENSG00000116254.18 | ENSG00000286894.2 | ENSG00000233652.1 | ENSG00000297800.1 |
| ENSG00000158270.12 | ENSG00000176428.6 | ENSG00000201850.1 | ENSG00000238045.10 |
| ENSG00000223912.1 | ENSG00000232293.2 | ENSG00000131480.9 | ENSG00000207378.1 |
| ENSG00000118997.14 | ENSG00000289837.2 | ENSG00000197380.11 | ENSG00000207342.1 |
| ENSG00000300068.1 | ENSG00000250303.7 | ENSG00000213352.3 | ENSG00000289425.1 |
| ENSG00000294095.1 | ENSG00000213068.3 | ENSG00000243396.1 | ENSG00000306655.1 |
| ENSG00000295182.1 | ENSG00000221500.1 | ENSG00000214727.2 | ENSG00000213787.3 |
| ENSG00000290981.2 | ENSG00000237930.1 | ENSG00000298556.1 | ENSG00000236248.1 |

| | | | |
|---|---|---|---|
| ENSG00000288813.2 | ENSG00000101049.18 | ENSG00000264217.2 | ENSG00000274341.1 |
| ENSG00000225726.1 | ENSG00000275532.3 | ENSG00000214776.13 | ENSG00000240024.8 |
| ENSG00000231018.1 | ENSG00000213033.4 | ENSG00000288598.3 | ENSG00000306078.1 |
| ENSG00000302922.1 | ENSG00000260644.7 | ENSG00000172247.4 | ENSG00000304377.1 |
| ENSG00000255991.2 | ENSG00000161921.17 | ENSG00000223676.1 | ENSG00000287045.3 |
| ENSG00000229716.2 | ENSG00000213318.4 | ENSG00000114023.16 | ENSG00000253564.1 |
| ENSG00000285972.2 | ENSG00000293309.2 | ENSG00000302221.1 | ENSG00000230445.5 |
| ENSG00000250602.7 | ENSG00000170927.15 | ENSG00000203362.2 | ENSG00000252915.1 |
| ENSG00000225443.1 | ENSG00000249266.1 | ENSG00000270800.4 | ENSG00000234377.9 |
| ENSG00000297712.1 | ENSG00000219547.1 | ENSG00000269997.1 | ENSG00000279812.1 |
| ENSG00000244705.1 | ENSG00000237186.1 | ENSG00000303128.1 | ENSG00000296159.1 |
| ENSG00000299119.1 | ENSG00000208772.1 | ENSG00000276523.1 | ENSG00000059728.11 |
| ENSG00000236003.1 | ENSG00000264812.1 | ENSG00000244331.1 | ENSG00000224161.3 |
| ENSG00000258891.1 | ENSG00000266179.2 | ENSG00000308113.1 | ENSG00000278970.5 |
| ENSG00000206418.5 | ENSG00000239465.1 | ENSG00000232145.1 | ENSG00000246889.4 |
| ENSG00000240068.1 | ENSG00000151012.13 | ENSG00000232732.12 | ENSG00000286943.1 |
| ENSG00000253347.1 | ENSG00000206659.1 | ENSG00000301158.1 | ENSG00000308906.1 |
| ENSG00000269365.2 | ENSG00000167306.21 | ENSG00000240412.1 | ENSG00000302714.1 |
| ENSG00000286342.4 | ENSG00000285793.2 | ENSG00000253966.1 | ENSG00000130203.10 |
| ENSG00000237273.1 | ENSG00000266049.1 | ENSG00000166012.17 | ENSG00000295896.1 |
| ENSG00000255074.1 | ENSG00000267558.2 | ENSG00000248415.1 | ENSG00000220925.2 |
| ENSG00000232768.1 | ENSG00000142233.14 | ENSG00000285745.1 | ENSG00000294365.1 |
| ENSG00000309736.1 | ENSG00000125385.9 | ENSG00000289508.3 | ENSG00000230894.2 |
| ENSG00000182687.4 | ENSG00000243544.3 | ENSG00000265142.9 | ENSG00000258904.1 |
| ENSG00000244217.1 | ENSG00000306081.1 | ENSG00000251629.8 | ENSG00000254604.1 |
| ENSG00000227845.2 | ENSG00000240074.1 | ENSG00000275839.1 | ENSG00000230507.5 |
| ENSG00000158113.13 | ENSG00000235797.2 | ENSG00000224427.1 | ENSG00000307378.1 |
| ENSG00000279071.1 | ENSG00000279707.1 | ENSG00000123901.9 | ENSG00000293913.1 |
| ENSG00000300630.1 | ENSG00000170409.7 | ENSG00000274967.1 | ENSG00000228111.1 |
| ENSG00000169075.7 | ENSG00000231470.1 | ENSG00000309953.1 | ENSG00000225142.2 |
| ENSG00000244752.3 | ENSG00000310485.1 | ENSG00000230166.1 | ENSG00000294256.1 |
| ENSG00000233820.3 | ENSG00000021461.18 | ENSG00000289283.1 | ENSG00000267277.3 |
| ENSG00000289915.2 | ENSG00000274452.1 | ENSG00000277299.1 | ENSG00000232625.1 |
| ENSG00000249072.1 | ENSG00000180815.15 | ENSG00000239183.1 | ENSG00000105696.9 |
| ENSG00000215467.2 | ENSG00000146592.17 | ENSG00000207113.1 | ENSG00000225901.1 |
| ENSG00000274618.2 | ENSG00000215244.5 | ENSG00000252700.1 | ENSG00000233300.1 |

| | | | |
|---|---|---|---|
| ENSG00000212695.5 | ENSG00000271919.1 | ENSG00000299210.1 | ENSG00000145192.13 |
| ENSG00000210184.1 | ENSG00000111186.13 | ENSG00000307927.1 | ENSG00000267560.2 |
| ENSG00000240669.1 | ENSG00000237806.1 | ENSG00000237846.1 | ENSG00000304656.1 |
| ENSG00000291022.2 | ENSG00000303113.1 | ENSG00000240919.1 | ENSG00000310528.1 |
| ENSG00000252174.1 | ENSG00000272323.2 | ENSG00000267258.1 | ENSG00000301979.1 |
| ENSG00000273345.6 | ENSG00000267301.1 | ENSG00000164463.12 | ENSG00000201820.1 |
| ENSG00000169313.10 | ENSG00000145248.7 | ENSG00000299405.1 | ENSG00000280620.2 |
| ENSG00000241671.1 | ENSG00000306093.1 | ENSG00000225092.2 | ENSG00000232875.1 |
| ENSG00000297426.1 | ENSG00000295792.1 | ENSG00000228744.1 | ENSG00000269821.2 |
| ENSG00000230732.4 | ENSG00000309828.1 | ENSG00000240052.1 | ENSG00000224349.3 |
| ENSG00000299221.1 | ENSG00000161940.11 | ENSG00000226646.1 | ENSG00000300303.1 |
| ENSG00000300371.1 | ENSG00000310521.1 | ENSG00000248626.1 | ENSG00000273513.2 |
| ENSG00000299767.1 | ENSG00000280069.1 | ENSG00000260417.1 | ENSG00000250068.1 |
| ENSG00000206738.1 | ENSG00000226237.5 | ENSG00000124006.15 | ENSG00000290660.2 |
| ENSG00000213113.3 | ENSG00000139890.10 | ENSG00000156113.25 | ENSG00000254263.1 |
| ENSG00000204033.10 | ENSG00000298762.1 | ENSG00000130182.8 | ENSG00000285840.3 |
| ENSG00000297192.1 | ENSG00000261529.1 | ENSG00000184454.7 | ENSG00000235300.5 |
| ENSG00000143217.9 | ENSG00000296306.1 | ENSG00000225569.1 | ENSG00000282968.1 |
| ENSG00000207468.1 | ENSG00000305693.1 | ENSG00000122733.12 | ENSG00000215156.6 |
| ENSG00000171885.18 | ENSG00000258354.1 | ENSG00000198682.13 | ENSG00000240096.1 |
| ENSG00000228275.1 | ENSG00000228599.1 | ENSG00000105583.11 | ENSG00000232786.1 |
| ENSG00000187475.6 | ENSG00000242866.10 | ENSG00000124103.10 | ENSG00000213295.3 |
| ENSG00000267752.1 | ENSG00000300254.1 | ENSG00000264538.8 | ENSG00000242137.1 |
| ENSG00000237296.12 | ENSG00000232608.1 | ENSG00000225662.2 | ENSG00000310009.1 |
| ENSG00000144857.15 | ENSG00000291047.2 | ENSG00000309127.1 | ENSG00000249119.1 |
| ENSG00000259687.5 | ENSG00000271127.2 | ENSG00000308736.1 | ENSG00000155026.17 |
| ENSG00000134259.6 | ENSG00000280057.1 | ENSG00000228190.2 | ENSG00000169402.16 |
| ENSG00000253454.1 | ENSG00000226292.1 | ENSG00000205500.9 | ENSG00000254311.1 |
| ENSG00000130821.17 | ENSG00000291060.2 | ENSG00000261330.1 | ENSG00000198785.7 |
| ENSG00000235085.3 | ENSG00000293009.1 | ENSG00000267066.1 | ENSG00000166183.16 |
| ENSG00000300122.1 | ENSG00000259396.1 | ENSG00000269755.1 | ENSG00000298843.1 |
| ENSG00000290817.1 | ENSG00000290165.1 | ENSG00000295206.1 | ENSG00000231699.1 |
| ENSG00000215457.5 | ENSG00000304009.1 | ENSG00000213484.2 | ENSG00000236913.1 |
| ENSG00000147697.10 | ENSG00000235351.1 | ENSG00000301348.1 | ENSG00000230282.1 |
| ENSG00000215386.16 | ENSG00000241105.1 | ENSG00000271378.1 | ENSG00000243328.1 |
| ENSG00000228799.6 | ENSG00000237130.1 | ENSG00000309134.1 | ENSG00000244451.1 |

| | | | |
|---|---|---|---|
| ENSG00000215197.4 | ENSG00000251680.7 | ENSG00000232390.3 | ENSG00000249061.1 |
| ENSG00000225358.1 | ENSG00000212331.1 | ENSG00000168490.14 | ENSG00000251587.1 |
| ENSG00000280107.1 | ENSG00000305762.1 | ENSG00000243560.3 | ENSG00000244361.1 |
| ENSG00000287129.2 | ENSG00000263002.8 | ENSG00000296978.1 | ENSG00000121904.19 |
| ENSG00000087258.17 | ENSG00000269680.1 | ENSG00000271207.1 | ENSG00000279108.1 |
| ENSG00000243250.1 | ENSG00000231193.1 | ENSG00000248858.8 | ENSG00000288349.3 |
| ENSG00000225661.7 | ENSG00000285595.1 | ENSG00000235253.1 | ENSG00000279347.1 |
| ENSG00000241400.1 | ENSG00000230148.9 | ENSG00000133740.11 | ENSG00000253993.1 |
| ENSG00000310204.1 | ENSG00000239622.1 | ENSG00000271250.1 | ENSG00000270702.1 |
| ENSG00000133665.13 | ENSG00000258111.1 | ENSG00000170486.11 | ENSG00000103832.10 |
| ENSG00000263307.3 | ENSG00000244674.1 | ENSG00000231788.4 | ENSG00000270269.1 |
| ENSG00000232273.1 | ENSG00000259182.6 | ENSG00000059804.18 | ENSG00000235013.1 |
| ENSG00000088827.14 | ENSG00000294196.1 | ENSG00000230757.1 | ENSG00000011105.14 |
| ENSG00000229222.1 | ENSG00000294533.1 | ENSG00000230002.3 | ENSG00000261126.7 |
| ENSG00000175604.2 | ENSG00000223576.2 | ENSG00000223718.3 | ENSG00000099953.10 |
| ENSG00000269066.1 | ENSG00000201785.1 | ENSG00000299264.1 | ENSG00000140749.9 |
| ENSG00000259160.1 | ENSG00000232818.2 | ENSG00000272574.1 | ENSG00000283481.1 |
| ENSG00000295229.1 | ENSG00000243164.1 | ENSG00000302984.1 | ENSG00000296510.1 |
| ENSG00000285865.1 | ENSG00000232928.2 | ENSG00000240138.1 | ENSG00000263069.6 |
| ENSG00000226330.2 | ENSG00000235644.1 | ENSG00000240654.6 | ENSG00000225885.8 |
| ENSG00000249249.3 | ENSG00000293223.1 | ENSG00000289725.2 | ENSG00000207088.1 |
| ENSG00000258544.1 | ENSG00000102385.13 | ENSG00000273951.1 | ENSG00000241886.1 |
| ENSG00000274512.6 | ENSG00000310070.1 | ENSG00000200816.1 | ENSG00000303909.1 |
| ENSG00000236580.1 | ENSG00000242634.1 | ENSG00000164675.11 | ENSG00000297527.1 |
| ENSG00000286835.1 | ENSG00000244390.2 | ENSG00000170160.19 | ENSG00000234253.1 |
| ENSG00000291112.2 | ENSG00000214041.4 | ENSG00000236888.1 | ENSG00000232587.1 |
| ENSG00000243101.1 | ENSG00000236090.2 | ENSG00000213041.4 | ENSG00000294048.1 |
| ENSG00000293363.3 | ENSG00000285909.2 | ENSG00000213717.3 | ENSG00000228844.1 |
| ENSG00000236526.2 | ENSG00000291280.2 | ENSG00000305831.1 | ENSG00000272158.1 |
| ENSG00000302592.1 | ENSG00000238003.1 | ENSG00000230269.8 | ENSG00000262172.1 |
| ENSG00000163694.15 | ENSG00000154654.15 | ENSG00000225124.1 | ENSG00000155761.14 |
| ENSG00000286904.1 | ENSG00000240870.2 | ENSG00000264840.1 | ENSG00000302979.1 |
| ENSG00000216642.1 | ENSG00000255248.11 | ENSG00000214652.7 | ENSG00000289130.3 |
| ENSG00000254162.1 | ENSG00000303219.1 | ENSG00000249838.1 | ENSG00000232992.2 |
| ENSG00000244753.2 | ENSG00000261665.2 | ENSG00000306469.1 | ENSG00000254758.1 |
| ENSG00000258051.1 | ENSG00000179460.5 | ENSG00000230289.2 | ENSG00000227770.1 |

| | | | |
|---|---|---|---|
| ENSG00000227309.1 | ENSG00000293167.1 | ENSG00000225933.1 | ENSG00000226520.1 |
| ENSG00000286786.1 | ENSG00000303381.1 | ENSG00000308733.1 | ENSG00000226510.3 |
| ENSG00000237793.3 | ENSG00000214908.3 | ENSG00000130829.18 | ENSG00000187144.11 |
| ENSG00000278983.1 | ENSG00000226928.1 | ENSG00000240634.1 | ENSG00000233189.1 |
| ENSG00000149925.22 | ENSG00000182383.8 | ENSG00000256167.2 | ENSG00000304366.1 |
| ENSG00000213269.2 | ENSG00000227578.1 | ENSG00000265626.1 | ENSG00000223916.1 |
| ENSG00000294990.1 | ENSG00000231164.1 | ENSG00000237709.1 | ENSG00000270948.1 |
| ENSG00000219410.7 | ENSG00000244196.1 | ENSG00000184916.9 | ENSG00000148600.15 |
| ENSG00000280378.1 | ENSG00000234559.1 | ENSG00000144821.11 | ENSG00000280012.2 |
| ENSG00000238149.1 | ENSG00000235847.2 | ENSG00000299077.1 | ENSG00000281348.1 |
| ENSG00000207380.1 | ENSG00000238711.1 | ENSG00000243806.1 | ENSG00000268257.3 |
| ENSG00000241472.9 | ENSG00000176007.10 | ENSG00000270589.1 | ENSG00000201221.1 |
| ENSG00000292370.2 | ENSG00000257475.2 | ENSG00000297888.1 | ENSG00000225460.1 |
| ENSG00000301832.1 | ENSG00000231636.1 | ENSG00000308368.1 | ENSG00000257662.1 |
| ENSG00000255968.1 | ENSG00000217030.1 | ENSG00000227230.2 | ENSG00000296519.1 |
| ENSG00000274038.1 | ENSG00000267793.1 | ENSG00000126583.12 | ENSG00000235595.1 |
| ENSG00000229893.3 | ENSG00000159433.12 | ENSG00000219863.5 | ENSG00000213970.5 |
| ENSG00000281183.1 | ENSG00000262823.2 | ENSG00000165188.14 | ENSG00000260796.1 |
| ENSG00000266944.1 | ENSG00000204103.4 | ENSG00000305978.1 | ENSG00000257391.2 |
| ENSG00000261596.2 | ENSG00000227663.1 | ENSG00000230116.1 | ENSG00000267474.1 |
| ENSG00000101665.10 | ENSG00000246731.4 | ENSG00000197989.16 | ENSG00000262492.1 |
| ENSG00000232496.2 | ENSG00000232751.1 | ENSG00000170745.12 | ENSG00000233885.7 |
| ENSG00000296297.1 | ENSG00000226617.1 | ENSG00000279202.1 | ENSG00000300389.1 |
| ENSG00000200834.1 | ENSG00000291073.2 | ENSG00000235296.3 | ENSG00000306812.1 |
| ENSG00000178381.13 | ENSG00000303016.1 | ENSG00000267669.2 | ENSG00000290796.1 |
| ENSG00000257222.2 | ENSG00000277447.1 | ENSG00000228655.7 | ENSG00000279985.1 |
| ENSG00000229115.1 | ENSG00000201388.1 | ENSG00000236866.6 | ENSG00000236804.1 |
| ENSG00000241859.8 | ENSG00000297639.1 | ENSG00000251298.2 | ENSG00000259118.6 |
| ENSG00000224888.4 | ENSG00000244244.2 | ENSG00000260220.7 | ENSG00000287933.2 |
| ENSG00000228976.1 | ENSG00000220541.4 | ENSG00000198467.16 | ENSG00000229519.2 |
| ENSG00000223263.1 | ENSG00000181982.20 | ENSG00000188305.6 | ENSG00000267251.2 |
| ENSG00000259550.2 | ENSG00000185130.6 | ENSG00000287650.1 | ENSG00000253198.1 |
| ENSG00000244429.1 | ENSG00000308821.1 | ENSG00000249183.1 | ENSG00000256351.1 |
| ENSG00000231137.1 | ENSG00000290394.3 | ENSG00000270455.1 | ENSG00000254079.1 |
| ENSG00000155367.16 | ENSG00000243498.2 | ENSG00000116194.13 | ENSG00000225602.5 |
| ENSG00000257731.2 | ENSG00000305766.1 | ENSG00000177710.6 | ENSG00000244561.1 |

| | | | |
|---|---|---|---|
| ENSG00000260206.1 | ENSG00000261739.3 | ENSG00000237986.4 | ENSG00000255340.1 |
| ENSG00000161249.21 | ENSG00000122884.13 | ENSG00000255141.1 | ENSG00000243197.8 |
| ENSG00000279129.1 | ENSG00000293220.2 | ENSG00000173262.12 | ENSG00000112245.14 |
| ENSG00000153363.16 | ENSG00000280800.1 | ENSG00000226532.1 | ENSG00000234855.3 |
| ENSG00000211450.11 | ENSG00000227667.1 | ENSG00000300210.1 | ENSG00000267044.1 |
| ENSG00000215450.2 | ENSG00000248826.2 | ENSG00000279225.1 | ENSG00000234832.2 |
| ENSG00000233493.4 | ENSG00000308665.1 | ENSG00000244053.1 | ENSG00000266236.1 |
| ENSG00000262248.1 | ENSG00000205097.7 | ENSG00000215458.9 | ENSG00000261868.1 |
| ENSG00000207523.1 | ENSG00000306748.1 | ENSG00000289157.2 | ENSG00000201492.1 |
| ENSG00000241651.4 | ENSG00000159186.4 | ENSG00000100625.9 | ENSG00000197409.9 |
| ENSG00000253558.2 | ENSG00000279098.1 | ENSG00000214896.4 | ENSG00000303195.1 |
| ENSG00000287064.1 | ENSG00000307995.1 | ENSG00000227924.2 | ENSG00000258443.1 |
| ENSG00000303671.1 | ENSG00000274898.1 | ENSG00000278206.1 | ENSG00000221879.3 |
| ENSG00000294288.1 | ENSG00000255670.1 | ENSG00000281181.1 | ENSG00000122691.13 |
| ENSG00000273066.5 | ENSG00000243730.2 | ENSG00000241217.3 | ENSG00000309043.1 |
| ENSG00000300094.1 | ENSG00000223802.9 | ENSG00000246477.3 | ENSG00000221716.1 |
| ENSG00000309098.1 | ENSG00000179528.16 | ENSG00000215054.3 | ENSG00000237763.10 |
| ENSG00000241652.3 | ENSG00000212493.1 | ENSG00000242198.1 | ENSG00000310029.1 |
| ENSG00000308808.1 | ENSG00000280047.1 | ENSG00000279077.1 | ENSG00000161610.1 |
| ENSG00000166922.9 | ENSG00000254815.7 | ENSG00000272831.3 | ENSG00000227729.5 |
| ENSG00000201998.1 | ENSG00000244723.3 | ENSG00000250346.1 | ENSG00000279796.1 |
| ENSG00000308179.1 | ENSG00000233990.1 | ENSG00000237818.1 | ENSG00000287454.2 |
| ENSG00000244259.1 | ENSG00000240210.3 | ENSG00000308871.1 | ENSG00000236607.3 |
| ENSG00000253995.1 | ENSG00000265656.1 | ENSG00000273759.1 | ENSG00000198585.12 |
| ENSG00000281383.1 | ENSG00000237017.1 | ENSG00000239317.1 | ENSG00000264270.1 |
| ENSG00000253299.1 | ENSG00000290706.2 | ENSG00000231519.1 | ENSG00000244627.6 |
| ENSG00000289419.1 | ENSG00000254756.2 | ENSG00000235264.1 | ENSG00000309590.1 |
| ENSG00000285849.1 | ENSG00000290386.1 | ENSG00000214047.4 | ENSG00000234937.1 |
| ENSG00000309392.1 | ENSG00000227014.7 | ENSG00000263823.3 | ENSG00000200755.1 |
| ENSG00000249870.1 | ENSG00000231241.1 | ENSG00000184423.5 | ENSG00000309256.1 |
| ENSG00000269546.1 | ENSG00000166405.16 | ENSG00000168209.6 | ENSG00000303185.1 |
| ENSG00000215003.4 | ENSG00000228041.2 | ENSG00000272211.1 | ENSG00000251313.1 |
| ENSG00000254985.1 | ENSG00000255933.1 | ENSG00000295436.1 | ENSG00000241219.3 |
| ENSG00000231587.1 | ENSG00000293164.1 | ENSG00000272885.1 | ENSG00000107611.16 |
| ENSG00000280614.1 | ENSG00000230756.1 | ENSG00000253875.3 | ENSG00000182809.11 |
| ENSG00000228347.1 | ENSG00000243280.2 | ENSG00000282936.3 | ENSG00000234067.1 |

| | | | |
|---|---|---|---|
| ENSG00000293064.2 | ENSG00000278487.1 | ENSG00000205041.1 | ENSG00000236998.2 |
| ENSG00000232228.1 | ENSG00000243368.2 | ENSG00000274756.1 | ENSG00000271350.1 |
| ENSG00000116983.13 | ENSG00000230779.1 | ENSG00000272181.1 | ENSG00000289437.1 |
| ENSG00000188818.13 | ENSG00000258841.1 | ENSG00000266891.1 | ENSG00000266563.1 |
| ENSG00000236498.1 | ENSG00000296921.1 | ENSG00000199053.3 | ENSG00000109686.21 |
| ENSG00000223878.1 | ENSG00000308294.1 | ENSG00000258901.2 | ENSG00000245904.7 |
| ENSG00000272426.2 | ENSG00000230406.2 | ENSG00000307891.1 | ENSG00000289115.2 |
| ENSG00000267605.9 | ENSG00000249456.1 | ENSG00000298740.1 | ENSG00000243672.1 |
| ENSG00000180172.7 | ENSG00000240023.1 | ENSG00000213172.3 | ENSG00000276842.1 |
| ENSG00000291250.2 | ENSG00000260948.1 | ENSG00000241120.1 | ENSG00000106819.13 |
| ENSG00000204991.11 | ENSG00000310496.1 | ENSG00000201818.1 | ENSG00000218018.5 |
| ENSG00000290461.2 | ENSG00000267934.1 | ENSG00000240589.3 | ENSG00000224494.1 |
| ENSG00000231741.2 | ENSG00000286561.2 | ENSG00000165496.5 | ENSG00000251158.2 |
| ENSG00000235354.1 | ENSG00000226498.2 | ENSG00000244176.1 | ENSG00000308620.1 |
| ENSG00000260536.1 | ENSG00000235377.1 | ENSG00000258957.3 | ENSG00000305447.1 |
| ENSG00000302671.1 | ENSG00000310131.1 | ENSG00000301500.1 | ENSG00000274658.1 |
| ENSG00000213872.3 | ENSG00000250752.2 | ENSG00000244019.1 | ENSG00000245711.2 |
| ENSG00000198920.11 | ENSG00000154118.13 | ENSG00000240003.2 | ENSG00000228686.2 |
| ENSG00000228028.2 | ENSG00000251880.1 | ENSG00000235066.8 | ENSG00000236546.2 |
| ENSG00000277581.1 | ENSG00000229766.8 | ENSG00000228195.2 | ENSG00000303592.1 |
| ENSG00000102057.11 | ENSG00000201944.1 | ENSG00000236051.7 | ENSG00000278330.1 |
| ENSG00000248971.2 | ENSG00000242052.1 | ENSG00000067715.15 | ENSG00000254336.1 |
| ENSG00000233690.1 | ENSG00000269067.2 | ENSG00000293347.2 | ENSG00000188739.15 |
| ENSG00000223552.4 | ENSG00000182888.5 | ENSG00000261535.1 | ENSG00000243353.1 |
| ENSG00000308217.1 | ENSG00000307746.1 | ENSG00000293956.1 | ENSG00000206674.1 |
| ENSG00000307363.1 | ENSG00000252311.1 | ENSG00000179157.4 | ENSG00000288794.1 |
| ENSG00000240729.1 | ENSG00000297356.1 | ENSG00000290469.1 | ENSG00000237170.3 |
| ENSG00000225918.1 | ENSG00000221819.6 | ENSG00000289074.3 | ENSG00000238085.1 |
| ENSG00000228429.1 | ENSG00000299398.1 | ENSG00000198868.3 | ENSG00000303133.1 |
| ENSG00000262902.1 | ENSG00000143882.12 | ENSG00000204071.11 | ENSG00000280010.1 |
| ENSG00000240463.1 | ENSG00000232238.1 | ENSG00000273284.1 | ENSG00000300824.1 |
| ENSG00000230469.1 | ENSG00000226933.1 | ENSG00000145708.11 | ENSG00000306896.1 |
| ENSG00000199977.1 | ENSG00000186352.9 | ENSG00000299425.1 | ENSG00000227602.1 |
| ENSG00000213613.2 | ENSG00000165113.13 | ENSG00000305087.1 | ENSG00000182253.15 |
| ENSG00000296280.1 | ENSG00000304862.1 | ENSG00000235829.1 | ENSG00000274808.6 |
| ENSG00000137460.10 | ENSG00000240388.2 | ENSG00000232797.1 | ENSG00000235315.2 |

| | | | |
|---|---|---|---|
| ENSG00000184163.3 | ENSG00000278662.6 | ENSG00000185221.6 | ENSG00000287061.1 |
| ENSG00000197520.11 | ENSG00000232172.1 | ENSG00000291013.2 | ENSG00000242229.1 |
| ENSG00000281376.1 | ENSG00000243023.2 | ENSG00000223822.2 | ENSG00000129048.7 |
| ENSG00000286305.1 | ENSG00000227533.8 | ENSG00000174946.7 | ENSG00000127951.8 |
| ENSG00000258928.1 | ENSG00000261114.1 | ENSG00000235670.1 | ENSG00000255717.10 |
| ENSG00000295151.1 | ENSG00000270906.1 | ENSG00000236993.2 | ENSG00000270276.3 |
| ENSG00000153902.14 | ENSG00000230751.1 | ENSG00000229652.1 | ENSG00000145362.21 |
| ENSG00000293325.2 | ENSG00000243844.1 | ENSG00000248483.7 | ENSG00000270137.1 |
| ENSG00000287587.1 | ENSG00000105220.17 | ENSG00000105642.16 | ENSG00000295697.1 |
| ENSG00000278770.1 | ENSG00000274062.1 | ENSG00000272489.1 | ENSG00000230929.5 |
| ENSG00000279845.1 | ENSG00000302472.1 | ENSG00000229752.1 | ENSG00000223945.2 |
| ENSG00000204637.4 | ENSG00000101489.20 | ENSG00000288899.1 | ENSG00000244151.1 |
| ENSG00000216917.2 | ENSG00000154485.5 | ENSG00000135519.8 | ENSG00000249148.2 |
| ENSG00000164849.10 | ENSG00000218459.1 | ENSG00000241146.1 | ENSG00000226318.1 |
| ENSG00000242439.1 | ENSG00000232177.1 | ENSG00000201501.1 | ENSG00000225591.2 |
| ENSG00000306199.1 | ENSG00000293970.1 | ENSG00000227474.1 | ENSG00000256148.1 |
| ENSG00000244229.1 | ENSG00000286326.2 | ENSG00000267152.1 | ENSG00000305613.1 |
| ENSG00000287962.1 | ENSG00000237783.2 | ENSG00000179363.7 | ENSG00000184060.12 |
| ENSG00000239481.1 | ENSG00000260287.4 | ENSG00000270903.1 | ENSG00000275834.1 |
| ENSG00000228261.2 | ENSG00000251482.1 | ENSG00000300127.1 | ENSG00000254459.1 |
| ENSG00000259238.1 | ENSG00000243494.1 | ENSG00000120162.10 | ENSG00000262147.2 |
| ENSG00000257121.1 | ENSG00000239870.1 | ENSG00000253153.1 | ENSG00000288922.3 |
| ENSG00000285567.3 | ENSG00000203721.9 | ENSG00000256243.2 | ENSG00000183723.13 |
| ENSG00000259326.1 | ENSG00000305725.1 | ENSG00000308845.1 | ENSG00000242029.1 |
| ENSG00000213169.3 | ENSG00000186008.7 | ENSG00000213025.2 | ENSG00000294851.1 |
| ENSG00000242142.1 | ENSG00000290020.2 | ENSG00000299659.1 | ENSG00000290692.2 |
| ENSG00000230615.9 | ENSG00000236404.13 | ENSG00000242640.1 | ENSG00000295686.1 |
| ENSG00000267143.1 | ENSG00000231890.11 | ENSG00000274611.4 | ENSG00000241002.1 |
| ENSG00000300957.1 | ENSG00000135116.9 | ENSG00000298413.1 | ENSG00000269191.1 |
| ENSG00000223374.1 | ENSG00000106089.12 | ENSG00000214362.2 | ENSG00000239272.1 |
| ENSG00000130707.18 | ENSG00000273183.1 | ENSG00000269637.1 | ENSG00000295288.1 |
| ENSG00000282924.1 | ENSG00000303404.1 | ENSG00000218472.2 | ENSG00000233839.1 |
| ENSG00000288050.1 | ENSG00000143590.14 | ENSG00000225487.2 | ENSG00000181359.6 |
| ENSG00000289023.1 | ENSG00000254634.4 | ENSG00000212452.1 | ENSG00000303192.1 |
| ENSG00000214944.10 | ENSG00000222162.1 | ENSG00000271913.10 | ENSG00000231857.1 |
| ENSG00000279356.1 | ENSG00000234884.3 | ENSG00000271727.3 | ENSG00000278233.1 |

| | | | |
|---|---|---|---|
| ENSG00000232656.11 | ENSG00000124762.15 | ENSG00000250825.1 | ENSG00000267430.1 |
| ENSG00000226673.2 | ENSG00000266171.1 | ENSG00000075043.21 | ENSG00000280721.2 |
| ENSG00000301562.1 | ENSG00000254997.4 | ENSG00000113594.10 | ENSG00000243265.1 |
| ENSG00000088899.16 | ENSG00000240738.1 | ENSG00000279977.1 | ENSG00000233560.2 |
| ENSG00000243499.1 | ENSG00000251435.1 | ENSG00000307981.1 | ENSG00000234750.1 |
| ENSG00000270882.3 | ENSG00000236991.7 | ENSG00000220506.2 | ENSG00000144802.11 |
| ENSG00000243160.1 | ENSG00000228816.1 | ENSG00000249987.1 | ENSG00000279744.1 |
| ENSG00000211794.3 | ENSG00000250229.1 | ENSG00000280347.1 | ENSG00000214760.3 |
| ENSG00000242602.1 | ENSG00000127083.8 | ENSG00000299262.1 | ENSG00000274677.1 |
| ENSG00000258846.1 | ENSG00000259461.1 | ENSG00000265936.1 | ENSG00000217334.1 |
| ENSG00000224438.3 | ENSG00000119547.6 | ENSG00000177374.13 | ENSG00000262500.1 |
| ENSG00000113739.11 | ENSG00000287855.1 | ENSG00000166337.11 | ENSG00000107968.10 |
| ENSG00000258086.2 | ENSG00000164199.18 | ENSG00000255533.1 | ENSG00000251440.2 |
| ENSG00000276334.1 | ENSG00000249815.3 | ENSG00000284783.2 | ENSG00000163406.11 |
| ENSG00000207425.1 | ENSG00000302537.1 | ENSG00000243122.1 | ENSG00000287555.1 |
| ENSG00000055955.17 | ENSG00000107249.24 | ENSG00000291166.2 | ENSG00000033327.13 |
| ENSG00000219881.1 | ENSG00000213782.7 | ENSG00000294203.1 | ENSG00000225644.1 |
| ENSG00000259845.1 | ENSG00000247853.2 | ENSG00000286018.1 | ENSG00000156475.19 |
| ENSG00000280020.1 | ENSG00000227355.6 | ENSG00000249096.8 | ENSG00000267523.1 |
| ENSG00000280143.1 | ENSG00000261997.1 | ENSG00000260691.8 | ENSG00000253810.1 |
| ENSG00000283235.3 | ENSG00000308748.1 | ENSG00000261007.2 | ENSG00000290972.1 |
| ENSG00000307836.1 | ENSG00000270385.1 | ENSG00000218187.2 | ENSG00000273950.1 |
| ENSG00000259179.1 | ENSG00000261124.1 | ENSG00000289974.2 | ENSG00000064270.14 |
| ENSG00000300065.1 | ENSG00000291024.2 | ENSG00000233952.1 | ENSG00000279500.1 |
| ENSG00000272183.1 | ENSG00000299120.1 | ENSG00000227207.2 | ENSG00000278900.1 |
| ENSG00000288787.2 | ENSG00000151846.9 | ENSG00000279092.1 | ENSG00000255051.1 |
| ENSG00000259271.1 | ENSG00000224662.2 | ENSG00000286864.1 | ENSG00000233595.2 |
| ENSG00000300750.1 | ENSG00000125409.13 | ENSG00000244743.1 | ENSG00000230837.1 |
| ENSG00000258613.1 | ENSG00000167183.3 | ENSG00000270935.1 | ENSG00000233544.1 |
| ENSG00000293427.2 | ENSG00000225447.1 | ENSG00000213653.4 | ENSG00000226803.10 |
| ENSG00000214093.3 | ENSG00000178809.12 | ENSG00000254441.1 | ENSG00000273828.3 |
| ENSG00000007516.15 | ENSG00000251062.1 | ENSG00000248616.1 | ENSG00000271754.1 |
| ENSG00000240236.1 | ENSG00000234125.5 | ENSG00000288993.2 | ENSG00000305474.1 |
| ENSG00000119139.21 | ENSG00000301129.1 | ENSG00000255508.7 | ENSG00000206903.1 |
| ENSG00000288745.1 | ENSG00000230935.1 | ENSG00000051108.15 | ENSG00000239483.1 |
| ENSG00000244641.2 | ENSG00000246763.8 | ENSG00000230595.1 | ENSG00000241668.2 |

| | | | |
|---|---|---|---|
| ENSG00000197360.9 | ENSG00000244585.1 | ENSG00000278774.1 | ENSG00000225770.1 |
| ENSG00000226003.2 | ENSG00000300298.1 | ENSG00000276691.1 | ENSG00000182240.16 |
| ENSG00000268658.5 | ENSG00000231684.3 | ENSG00000173926.6 | ENSG00000181690.8 |
| ENSG00000298552.1 | ENSG00000228409.9 | ENSG00000141750.7 | ENSG0000256278.1 |
| ENSG00000296014.1 | ENSG00000213671.3 | ENSG00000234352.10 | ENSG00000290949.2 |
| ENSG00000164509.17 | ENSG00000213579.3 | ENSG00000267024.2 | ENSG00000240531.1 |
| ENSG00000256967.2 | ENSG00000141753.7 | ENSG00000248794.1 | ENSG00000260167.2 |
| ENSG00000182272.12 | ENSG00000251306.1 | ENSG00000177453.7 | ENSG00000197575.6 |
| ENSG00000257907.2 | ENSG00000174130.13 | ENSG00000057657.17 | ENSG00000293906.1 |
| ENSG00000308157.1 | ENSG00000181634.8 | ENSG00000201104.1 | ENSG00000293258.1 |
| ENSG00000185203.13 | ENSG00000243417.1 | ENSG00000047648.23 | ENSG00000237676.1 |
| ENSG00000233278.1 | ENSG00000240238.1 | ENSG00000285151.1 | ENSG00000271869.3 |
| ENSG00000137070.19 | ENSG00000173486.14 | ENSG00000234985.2 | ENSG00000055609.21 |
| ENSG00000243894.1 | ENSG00000263753.11 | ENSG00000237875.1 | ENSG00000241350.1 |
| ENSG00000198553.9 | ENSG00000242163.1 | ENSG00000235390.4 | ENSG00000276667.1 |
| ENSG00000255566.1 | ENSG00000303573.1 | ENSG00000286145.3 | ENSG00000188994.14 |
| ENSG00000251991.1 | ENSG00000294221.1 | ENSG00000262482.2 | ENSG00000223467.2 |
| ENSG00000213529.3 | ENSG00000224282.3 | ENSG00000241030.1 | ENSG00000175155.11 |
| ENSG00000239559.2 | ENSG00000260318.1 | ENSG00000286067.1 | ENSG00000274374.1 |
| ENSG00000198590.12 | ENSG00000277157.2 | ENSG00000296452.1 | ENSG00000268785.1 |
| ENSG00000298580.1 | ENSG00000270781.1 | ENSG00000239323.1 | ENSG00000182389.20 |
| ENSG00000133812.18 | ENSG00000307702.1 | ENSG00000177508.12 | ENSG00000243312.3 |
| ENSG00000236148.5 | ENSG00000270431.1 | ENSG00000261243.1 | ENSG00000132613.16 |
| ENSG00000250111.3 | ENSG00000237757.2 | ENSG00000249679.4 | ENSG00000213900.2 |
| ENSG00000300853.1 | ENSG00000309270.1 | ENSG00000230370.1 | |
| ENSG00000280407.3 | ENSG00000250098.2 | ENSG00000273302.1 | |
| ENSG00000164309.15 | ENSG00000269934.3 | ENSG00000275757.1 | |

Table S24. Jurkat Cells - DEG representative genes from enriched pathway and process for upregulated genes – *ZNF683* used as quality control indicator

| Gene Symbol | Function |
|---|---|
| *CD44* | Maintenance of memory T cells<br>https://doi.org/10.3389/fimmu.2012.00023 |
| *IL9R* | Correlates with lower expression of the exhaustion markers<br>https://doi.org/10.4049/jimmunol.2300094 |
| *JUNB* | Essential transcription factor for cytotoxic CD8 T cell responses<br>https://doi.org/10.1093/intimm/dxae063 |
| *JAK3* | Associated with maintaining memory T cell homeostasis<br>https://doi.org/10.4110/in.2023.23.e10 |
| *IL2RB* | Associated with T cells rapidly attaining memory T cell traits<br>https://doi.org/10.1126/sciimmunol.abl6322 |
| *CCR2* | Modulates memory T cell response<br>https://doi.org/10.1186/s12974-024-03309-y |
| *CCR5* | Associated with early memory T cell response<br>https://doi.org/10.1016/j.immuni.2008.05.011 |
| *IL36A* | Associated with efficient early cytokine and chemokine production<br>https://doi.org/10.1038/mi.2016.107 |
| *IL12RB2* | Associated with expansion of natural killer cells<br>https://doi.org/10.1084/jem.20111760 |
| *IRF6* | Increased in resident memory T cells<br>https://doi.org/10.1016/j.immuni.2023.05.003 |
| *IRF5* | Associated with delayed occurrence of terminal differentiation markers<br>https://doi.org/10.1038/s44318-025-00485-2 |
| *CD40LG* | Allows T cells to promote their own expansion<br>https://doi.org/10.3389/fimmu.2017.01484 |
| *IL16* | Associated with memory T cell function<br>https://doi.org/10.1172/jci.insight.153963 |
| *TNFRSF8* | Expressed in memory T cells<br>https://doi.org/10.1126/sciimmunol.adf8776 |
| *TNFRSF11A* | Expressed in memory T cells<br>https://doi.org/10.3389/fimmu.2022.806906 |

Table S25. Jurkat Cells - DEG representative genes from enriched pathway and process for downregulated genes – *ZNF683* used as quality control indicator

| Gene Symbol | Function |
|---|---|
| *PPP1R15A* | Associated with cellular stress<br>https://doi.org/10.1016/j.medj.2024.03.015 |
| *ADM* | Associated with suppressing CD8 T cell proliferation and cytotoxic function<br>https://doi.org/10.3389/fimmu.2025.1606125 |
| *HMOX1* | Hinders antitumor $CD8^+$ T-cell function in the tumor microenvironment<br>https://doi.org/10.1002/cam4.5787 |

Table S26. Jurkat Cells - Comparison of ZNF683+ (HOBIT_POS) and ZNF683– Cells (HOBIT_NEG) – Memory T Cell-Associated Gene Signature from GSEA-MSigDB Human Gene Set from the following resources:
GSE10239_MEMORY_VS_KLRG1HIGH_EFF_CD8_TCELL_UP,
GSE16522_MEMORY_VS_NAIVE_ANTI_CD3CD28_STIM_CD8_TCELL_UP,
GSE32423_MEMORY_VS_NAIVE_CD8_TCELL_UP,
GO:0043382 GOBP_POSITIVE_REGULATION_OF_MEMORY_T_CELL_DIFFERENTIATION

| SYMBOL | $Log_2FC$ | FDR | SRR25658160_ HOBIT_POS | SRR25658161_ HOBIT_POS | SRR25658158_ HOBIT_POS | SRR25658156_ HOBIT_NEG | SRR25658159_ HOBIT_NEG |
|---|---|---|---|---|---|---|---|
| *TACR1* | 2.4665 | 7.E-04 | 0.1212 | 1.5308 | 0.1151 | -1.0673 | -0.6998 |
| *SAMHD1* | 2.0597 | 7.E-04 | 0.4739 | 0.1114 | 1.3642 | -0.9461 | -1.0035 |
| *MST1R* | 2.0377 | 1.E-02 | 1.4193 | 0.0274 | 0.4004 | -1.1672 | -0.6799 |
| *UCP3* | 1.9728 | 3.E-08 | 0.4305 | 0.9781 | 0.6879 | -1.3543 | -0.7422 |
| *CLCF1* | 1.9041 | 1.E-05 | 1.5102 | 0.1605 | 0.1555 | -0.8806 | -0.9455 |
| *COL25A1* | 1.6695 | 2.E-03 | 0.9488 | 0.3185 | 0.6456 | -0.3433 | -1.5696 |
| *TIMP1* | 1.3817 | 7.E-12 | 1.3289 | 0.4873 | 0.1616 | -1.0751 | -0.9026 |
| *TGFB1I1* | 1.3373 | 4.E-02 | 1.1599 | 0.3592 | 0.5573 | -1.2099 | -0.8666 |
| *HR* | 1.1976 | 1.E-32 | 0.3357 | 0.4816 | 1.038 | -0.2731 | -1.5822 |
| *MAP7* | 1.1878 | 2.E-18 | 0.046 | 0.3994 | 1.3334 | -0.4005 | -1.3784 |
| *NT5E* | 1.1523 | 1.E-104 | 0.4874 | 0.5789 | 0.7296 | -0.0972 | -1.6987 |
| *SYT12* | 1.1487 | 2.E-04 | 1.0586 | 0.8292 | 0.0291 | -0.538 | -1.379 |
| *HGH1* | 1.0282 | 1.E-125 | 0.0671 | 1.0798 | 0.5892 | -0.1697 | -1.5664 |
| *IKBKE* | 0.9828 | 2.E-293 | 0.3893 | 0.7966 | 0.7205 | -0.2907 | -1.6156 |
| *SASH3* | 0.9674 | 0.E+00 | 0.468 | 0.6051 | 0.6992 | -0.063 | -1.7094 |
| *ADCY7* | 0.9642 | 4.E-198 | 0.2955 | 0.9439 | 0.7272 | -0.4547 | -1.512 |
| *AIMP2* | 0.9568 | 7.E-252 | 0.1723 | 1.1069 | 0.5078 | -0.2262 | -1.5608 |
| *CTSW* | 0.9062 | 2.E-105 | 0.3293 | 0.5994 | 1.1552 | -0.9265 | -1.1574 |
| *CCR7* | 0.8646 | 6.E-25 | 0.0029 | 0.7661 | 0.9851 | -0.2047 | -1.5494 |
| *CCDC71* | 0.8528 | 5.E-137 | 0.4928 | 0.7258 | 0.669 | -0.2362 | -1.6514 |
| *GRAP* | 0.8399 | 3.E-44 | 0.4819 | 0.3396 | 0.8947 | -0.0277 | -1.6885 |
| *APOBEC3C* | 0.8254 | 2.E-84 | 0.5419 | 0.5877 | 0.879 | -0.4698 | -1.5387 |
| *CRMP1* | 0.8173 | 2.E-04 | 0.7125 | 0.5691 | 0.5175 | -0.0988 | -1.7003 |
| *RIMBP2* | 0.7981 | 3.E-06 | 0.0988 | 1.4982 | 0.199 | -0.682 | -1.114 |

| | | | | | | | |
|---|---|---|---|---|---|---|---|
| *GMIP* | 0.79 | 8.E-227 | 0.4889 | 0.8206 | 0.7578 | -0.5998 | -1.4675 |
| *LYPLA2* | 0.7525 | 2.E-255 | 0.4907 | 1.0296 | 0.5248 | -0.6148 | -1.4303 |
| *CD84* | 0.7463 | 0.E+00 | 0.3809 | 0.5599 | 0.7833 | -0.0131 | -1.711 |
| *EVL* | 0.7356 | 0.E+00 | 1.0421 | 0.1304 | 0.7157 | -0.3954 | -1.4928 |
| *RAB37* | 0.7193 | 0.E+00 | 0.7734 | 0.1182 | 0.8134 | -0.0547 | -1.6503 |
| *IQCC* | 0.7057 | 5.E-69 | 0.3429 | 0.6647 | 0.9018 | -0.3204 | -1.589 |
| *NUBP1* | 0.705 | 1.E-53 | 0.4182 | 0.7084 | 0.722 | -0.1848 | -1.6637 |
| *DOK1* | 0.7028 | 7.E-116 | 0.4744 | 0.67 | 0.6172 | -0.0461 | -1.7155 |
| *SLC10A3* | 0.7 | 3.E-60 | 0.2683 | 1.0901 | 0.726 | -0.9006 | -1.1838 |
| *TMC6* | 0.6973 | 1.E-174 | 0.9441 | 0.382 | 0.7583 | -0.7142 | -1.3702 |
| *NEB* | 0.6847 | 2.E-04 | 0.6964 | 0.1494 | 1.0936 | -0.5136 | -1.4258 |
| *ATXN7L2* | 0.6608 | 3.E-60 | 0.0628 | 0.7074 | 1.095 | -0.4049 | -1.4603 |
| *ABCF2* | 0.657 | 8.E-144 | 0.0057 | 1.1733 | 0.5973 | -0.3012 | -1.4751 |
| *XPC* | 0.6534 | 4.E-228 | 0.3346 | 0.7679 | 0.7046 | -0.1391 | -1.6681 |
| *LHFPL2* | 0.6527 | 2.E-21 | 0.3709 | 0.5625 | 0.9862 | -0.3553 | -1.5643 |
| *MRPL20* | 0.6448 | 5.E-285 | 0.1596 | 1.1629 | 0.3504 | -0.0948 | -1.5781 |
| *DDR1* | 0.6361 | 3.E-47 | 0.7462 | 0.5036 | 0.5571 | -0.1138 | -1.693 |
| *SLC12A7* | 0.6318 | 1.E-280 | 0.4064 | 0.798 | 0.9022 | -0.7657 | -1.3409 |
| *LRRC61* | 0.6014 | 3.E-90 | 0.474 | 1.0569 | 0.4132 | -0.4254 | -1.5187 |
| *HOXB13* | 0.588 | 7.E-17 | 0.357 | 1.0677 | 0.2833 | -0.0814 | -1.6266 |

Table S27. Jurkat Cells - Comparison of ZNF683+ (HOBIT_POS) and ZNF683– Cells (HOBIT_NEG) – Natural Killer Cell-Associated Gene Signature from GSEA-MSigDB Human Gene Set from the following resources:

GSE130646 RUBENSTEIN_SKELETAL_MUSCLE_NK_CELLS

GSE130148 TRAVAGLINI_LUNG_NATURAL_KILLER_T_CELL

BIOCARTA_NKT_PATHWAY

KEGG_NATURAL_KILLER_CELL_MEDIATED_CYTOTOXICITY

| SYMBOL | $Log_2FC$ | FDR | SRR25658160 _HOBIT_POS | SRR25658161 _HOBIT_POS | SRR25658158 _HOBIT_POS | SRR25658156 _HOBIT_NEG | SRR25658159 _HOBIT_NEG |
|---|---|---|---|---|---|---|---|
| *ACTB* | 0.8048 | 0.E+00 | 0.2597823 | 0.9285271 | 0.6512703 | -0.2290761 | -1.6105036 |
| *ADGRE5* | 0.7148 | 9.E-95 | 0.8959518 | 0.364811 | 0.7336113 | -0.4789045 | -1.5154696 |
| *AKNA* | 0.9960 | 0.E+00 | 0.5274715 | 0.549576 | 0.7578256 | -0.1551831 | -1.67969 |
| *CARD9* | 4.1316 | 9.E-06 | 0.0175245 | 1.6195582 | 0.0134927 | -0.740605 | -0.9099705 |
| *CASP4* | 3.0307 | 6.E-08 | 0.459946 | 0.2793412 | 1.2674599 | -0.790716 | -1.2160312 |
| *CCR7* | 0.8646 | 6.E-25 | 0.0029106 | 0.7661466 | 0.9850896 | -0.2047291 | -1.5494178 |
| *CD244* | 1.1678 | 4.E-88 | 0.4346512 | 0.580234 | 0.7969358 | -0.1319749 | -1.6798461 |
| *CD52* | 0.688 | 1.E-114 | 0.1906618 | 0.770747 | 0.7874119 | -0.0932597 | -1.655561 |
| *CD6* | 0.5816 | 3.E-195 | 0.4759976 | 0.6727516 | 0.8800181 | -0.5201915 | -1.5085757 |
| *CTSW* | 0.9062 | 2.E-105 | 0.3293468 | 0.5993592 | 1.1551626 | -0.9265132 | -1.1573555 |
| *EVL* | 0.735 | 0.E+00 | 1.0420659 | 0.1304434 | 0.7157136 | -0.3954111 | -1.4928118 |
| *EZR* | 0.6256 | 0.E+00 | 0.3564504 | 0.6873543 | 0.6832557 | -0.0143306 | -1.7127298 |
| *FGR* | 0.8798 | 7.E-13 | 0.1227047 | 1.0154431 | 0.8989936 | -0.8306127 | -1.2065288 |
| *GZMK* | 0.5862 | 2.E-05 | 0.346439 | 1.3280861 | 0.1937725 | -0.5258895 | -1.3424082 |
| *IL2RB* | 1.9998 | 6.E-61 | 0.2201511 | 0.9110189 | 0.9632825 | -1.059454 | -1.0349985 |
| *KIR3DL2* | 1.6226 | 7.E-08 | 0.0017454 | 1.2473804 | 0.4195184 | -0.1725605 | -1.4960837 |
| *KLRD1* | 1.0377 | 5.E-07 | 0.0501116 | 0.2004467 | 1.3438055 | -1.4606948 | -0.1336691 |
| *MCM5* | 0.5970 | 0.E+00 | 0.3804711 | 0.9077453 | 0.6646184 | -0.3921482 | -1.5606865 |
| *NCR2* | 1.3145 | 3.E-06 | 0.0380628 | 0.5025034 | 1.0790183 | -0.01285 | -1.6067346 |
| *NCR3* | 1.690 | 2.E-06 | 0.9814054 | 0.4969295 | 0.6261827 | -0.7495194 | -1.3549983 |
| *OIP5* | 0.6333 | 4.E-27 | 0.4739563 | 0.9945707 | 0.5232967 | -0.4817745 | -1.5100492 |
| *PIK3CD* | 0.7022 | 2.E-224 | 0.4147659 | 0.7693707 | 0.7765906 | -0.384243 | -1.5764842 |
| *PRF1* | 1.7138 | 3.E-202 | 0.6363983 | 0.5829887 | 0.6885725 | -0.2608584 | -1.6471011 |
| *PTPN6* | 0.7944 | 0.E+00 | 0.7261509 | 0.5871606 | 0.610121 | -0.2886994 | -1.6347331 |
| *SASH3* | 0.9673 | 0.E+00 | 0.4680248 | 0.6050726 | 0.6992487 | -0.0629823 | -1.7093638 |

| | | | | | | | |
|---|---|---|---|---|---|---|---|
| *SH3BP2* | 0.6932 | 1.E-100 | 0.2968173 | 0.6556793 | 1.0311837 | -0.5168924 | -1.4667879 |
| *SHC1* | 0.5828 | 1.E-181 | 0.5526819 | 1.0105206 | 0.4853576 | -0.6122608 | -1.4362993 |
| *SUPT4H1* | 1.0287 | 3.E-29 | 0.5263283 | 0.4448697 | 0.9729713 | -1.5588125 | -0.3853567 |
| *SURF4* | 0.6730 | 0 | 0.3914905 | 0.7250839 | 0.6119415 | -0.0120161 | -1.7164998 |
| *TUBA1A* | 0.6250 | 0 | 0.6451509 | 0.6985892 | 0.4127655 | -0.0467716 | -1.709734 |
| *TUBA1B* | 0.930 | 0 | 0.7035007 | 0.7443983 | 0.4710385 | -0.2931654 | -1.6257721 |
| *UCP2* | 0.7787 | 0 | 0.4902072 | 0.7261265 | 0.5756436 | -0.0911656 | -1.7008117 |
| *UNC13D* | 0.894 | 0 | 0.5873367 | 0.6181103 | 0.6697749 | -0.2074918 | -1.6677302 |
| *WAS* | 0.6955 | 8.670e-321 | 0.4913505 | 0.7996123 | 0.6275715 | -0.2940748 | -1.6244596 |

Table S28. Jurkat Cells - Comparison of ZNF683+ (HOBIT_POS) and ZNF683– Cells (HOBIT_NEG) – Stress-Related Response Gene Signature from GSEA-MSigDB Human Gene Set

GO:0035966 GOBP_RESPONSE_TO_TOPOLOGICALLY_INCORRECT_PROTEIN

GO:0033554 GOBP_CELLULAR_RESPONSE_TO_STRESS

| SYMBOL | $Log_2FC$ | FDR | SRR25658160 _HOBIT _POS | SRR25658161 _HOBIT _POS | SRR25658158 _HOBIT _POS | SRR25658156 _HOBIT _NEG | SRR25658159 _HOBIT _NEG |
|---|---|---|---|---|---|---|---|
| *ADM* | -4.82.E+00 | 0.E+00 | -0.550205261 | -0.86109436 | -0.667061885 | 0.620238132 | 1.458123374 |
| *ADRB1* | -1.31.E+00 | 2.E-77 | -0.434551979 | -0.755746846 | -0.660996552 | 0.18759364 | 1.663701736 |
| *AMFR* | -8.97.E-01 | 0.E+00 | -0.20046879 | -1.16751441 | -0.659667424 | 0.783740838 | 1.243909786 |
| *AREG* | -2.98.E+00 | 7.E-05 | -0.981597195 | -0.656239984 | -0.514687425 | 1.186334546 | 0.966190058 |
| *BHLHE40* | -2.76.E+00 | 0.E+00 | -0.43193959 | -1.023363101 | -0.604467546 | 0.656088099 | 1.403682139 |
| *BNIP3* | -1.97.E+00 | 0.E+00 | -0.583346281 | -0.983198935 | -0.578852437 | 0.90448574 | 1.240911912 |
| *CAV1* | -3.13.E+01 | 5.E-04 | -0.58811489 | -0.58811489 | -0.58811489 | 1.72059543 | 0.04374924 |
| *CCND1* | -2.22.E+00 | 5.E-18 | -0.582155945 | -0.630188324 | -0.584392317 | 0.089569315 | 1.707167271 |
| *CDKN2D* | -6.34.E-01 | 1.E-231 | -0.344530623 | -0.861277304 | -0.582236766 | 0.118797059 | 1.669247634 |
| *CREB3L2* | -7.56.E-01 | 5.E-163 | -0.211772329 | -1.045645062 | -0.398031841 | 0.011301485 | 1.644147747 |
| *CREBBP* | -9.21.E-01 | 3.E-199 | -0.4291305 | -0.77591953 | -0.543594598 | 0.040829125 | 1.707815503 |
| *CXCL12* | -3.40.E+00 | 6.E-04 | -0.599421947 | -0.926956956 | -0.600882638 | 0.782771949 | 1.344489592 |
| *DDIT4* | -1.66.E+00 | 0.E+00 | -0.589596474 | -0.777599362 | -0.487920026 | 0.189681976 | 1.665433885 |
| *DSC2* | -3.40.E+00 | 2.E-02 | -0.263192717 | -0.920326964 | -0.920326964 | 1.160644989 | 0.943201655 |
| *ERN1* | -1.41.E+00 | 8.536e-316 | -0.309531702 | -0.760713349 | -0.673229648 | 0.049418934 | 1.694055765 |
| *FOXA3* | -1.09.E+00 | 1.E-10 | -0.337383428 | -0.600537134 | -0.826918815 | 0.081044672 | 1.683794705 |
| *FUT1* | -3.08.E+00 | 1.E-62 | -0.674349908 | -0.886368384 | -0.614075515 | 0.993454114 | 1.181339694 |
| *HK1* | -6.65.E-01 | 0.E+00 | -0.802393812 | -0.477027989 | -0.53518611 | 0.134069398 | 1.680538513 |
| *HK2* | -1.04.E+00 | 0.E+00 | -0.790754169 | -0.856251629 | -0.478462653 | 0.785419949 | 1.340048501 |
| *HRK* | -1.59.E+00 | 2.E-38 | -0.608478908 | -0.78093769 | -0.408767223 | 0.112940978 | 1.685242842 |
| *IGFBP2* | -2.00.E+00 | 0 | -0.564949256 | -0.562013455 | -0.611886763 | 0.009550022 | 1.729299453 |
| *IGFBP7* | -8.09.E-01 | 2.E-02 | -0.422509864 | -0.499686642 | -1.08615712 | 0.570979836 | 1.43737379 |
| *KCNJ11* | -9.96.E-01 | 9.E-21 | -0.042515681 | -0.725351871 | -1.057230471 | 0.325680451 | 1.499417572 |
| *KLF10* | -5.92.E-01 | 1.E-145 | -0.954355639 | -0.589334434 | -0.289635792 | 0.217570849 | 1.615755016 |
| *LDHA* | -1.46.E+00 | 0 | -0.622062875 | -0.568501407 | -0.663352914 | 0.174860746 | 1.67905645 |
| *LIPG* | -1.47.E+00 | 4.E-03 | -0.808464284 | -1.057041317 | -0.030796602 | 0.48440564 | 1.411896562 |
| *NUAK2* | -3.54.E+01 | 2.E-03 | -0.728454358 | -0.728454358 | -0.728454358 | 1.193072919 | 0.992290156 |

| | | | | | | | |
|---|---|---|---|---|---|---|---|
| *OTUD3* | -1.30.E+00 | 0 | -0.139230052 | -1.119385739 | -0.470403723 | 0.153328818 | 1.575690696 |
| *PAGE4* | -2.53.E+00 | 1.E-09 | -0.757225199 | -0.604356629 | -0.593074771 | 0.345155787 | 1.609500811 |
| *PFKFB3* | -6.63.E-01 | 1.E-277 | -0.96046853 | -0.780427614 | -0.404298781 | 1.027626322 | 1.117568602 |
| *PGK1* | -9.97.E-01 | 0 | -0.603417128 | -0.621751371 | -0.802920823 | 0.490566864 | 1.537522457 |
| *PPP1R15B* | -1.33.E+00 | 0 | -0.321286295 | -1.069612543 | -0.321954993 | 0.08762428 | 1.625229551 |
| *SESN3* | -1.09.E+00 | 8.E-92 | -0.10796941 | -1.233498361 | -0.309196109 | 0.11509376 | 1.53557012 |
| *STC1* | -3.22.E+00 | 3.E-19 | -0.131418552 | -0.812249143 | -0.812249143 | 0.129089322 | 1.626827517 |
| *STC2* | -1.57.E+00 | 0 | -0.173750203 | -1.068193786 | -0.793980655 | 1.268833846 | 0.767090798 |
| *UCP1* | -2.17.E+00 | 2.E-23 | -0.454197379 | -0.949394228 | -0.52658295 | 1.578389336 | 0.35178522 |
| *VEGFA* | -2.79.E+00 | 0 | -0.008383275 | -1.057073051 | -0.799743176 | 0.43087645 | 1.434323053 |
| *VGF* | -8.03.E-01 | 6.E-04 | -0.65234177 | -0.219489237 | -1.185189025 | 1.082989181 | 0.974030851 |

Table S29. APC - Single Nuclei RNA-seq Nuclei Distribution -

All Conditions – Raw Counts

| Raw Counts after Reclustering | CTRL _Adt | ASyS _Jo-1 | ASyS _OJ | ASyS _PL7 | DM _MDA5 | DM _Mi-2 | DM _NXP-2 | DM _TIF1-γ | DM _SAE | IBM | IMNM _HMGCR | IMNM _SRP |
|---|---|---|---|---|---|---|---|---|---|---|---|---|
| M1/2 | 2 | 800 | 1036 | 19 | 30 | 25 | 26 | 115 | 1 | 149 | 843 | 187 |
| M1 | 6 | 431 | 81 | 11 | 102 | 321 | 584 | 973 | 295 | 25 | 55 | 50 |
| M2 | 149 | 141 | 871 | 349 | 298 | 173 | 50 | 87 | 53 | 1234 | 477 | 1297 |
| Prolif_APC | 1 | 79 | 18 | 12 | 7 | 8 | 4 | 3 | 9 | 22 | 275 | 12 |
| cDC1 | 2 | 56 | 19 | 6 | 14 | 9 | 10 | 10 | 15 | 53 | 10 | 10 |
| CD2+DC | 0 | 12 | 20 | 4 | 1 | 3 | 11 | 5 | 2 | 70 | 21 | 21 |
| cDC2 | 17 | 11 | 21 | 2 | 9 | 2 | 2 | 1 | 10 | 45 | 6 | 13 |
| St_APC | 13 | 2 | 4 | 0 | 16 | 2 | 6 | 1 | 0 | 1 | 19 | 1 |
| Total APC | 190 | 1532 | 2070 | 403 | 477 | 543 | 693 | 1195 | 385 | 1599 | 1706 | 1591 |

Table S30. APC - Single Nuclei RNA-seq Nuclei Distribution - All Conditions – Proportion

| % after Reclustering | CTRL _Adt | ASyS _Jo-1 | ASyS _OJ | ASyS _PL7 | DM _MDA5 | DM _Mi-2 | DM _NXP-2 | DM _TIF1-γ | DM _SAE | IBM | IMNM _HMGCR | IMNM _SRP |
|---|---|---|---|---|---|---|---|---|---|---|---|---|
| M1/2 | 1.05 | 52.22 | 50.05 | 4.71 | 6.29 | 4.60 | 3.75 | 9.62 | 0.26 | 9.32 | 49.41 | 11.75 |
| M1 | 3.16 | 28.13 | 3.91 | 2.73 | 21.38 | 59.12 | 84.27 | 81.42 | 76.62 | 1.56 | 3.22 | 3.14 |
| M2 | 78.42 | 9.20 | 42.08 | 86.60 | 62.47 | 31.86 | 7.22 | 7.28 | 13.77 | 77.17 | 27.96 | 81.52 |
| Prolif_APC | 0.53 | 5.16 | 0.87 | 2.98 | 1.47 | 1.47 | 0.58 | 0.25 | 2.34 | 1.38 | 16.12 | 0.75 |
| cDC1 | 1.05 | 3.66 | 0.92 | 1.49 | 2.94 | 1.66 | 1.44 | 0.84 | 3.90 | 3.31 | 0.59 | 0.63 |
| CD2+DC | 0.00 | 0.78 | 0.97 | 0.99 | 0.21 | 0.55 | 1.59 | 0.42 | 0.52 | 4.38 | 1.23 | 1.32 |
| cDC2 | 8.95 | 0.72 | 1.01 | 0.50 | 1.89 | 0.37 | 0.29 | 0.08 | 2.60 | 2.81 | 0.35 | 0.82 |
| St_APC | 6.84 | 0.13 | 0.19 | 0.00 | 3.35 | 0.37 | 0.87 | 0.08 | 0.00 | 0.06 | 1.11 | 0.06 |
| Total APC | 100 | 100 | 100 | 100 | 100 | 100 | 100 | 100 | 100 | 100 | 100 | 100 |

Table S31. IBM vs. Controls only APC , Individual Sample Comparison – Raw Counts

| Nuclei Type | CTRL_1 | CTRL_2 | IBM_1 | IBM_2 | IBM_3 | IBM_4 |
|---|---|---|---|---|---|---|
| M1/2 | 1 | 1 | 14 | 19 | 70 | 46 |
| M1 | 2 | 4 | 5 | 5 | 13 | 2 |
| M2 | 115 | 34 | 328 | 434 | 190 | 282 |
| Prolif_APC | 1 | 0 | 5 | 12 | 3 | 2 |
| cDC1 | 1 | 1 | 17 | 21 | 8 | 7 |
| CD2+DC | 0 | 0 | 13 | 16 | 14 | 27 |
| cDC2 | 15 | 2 | 2 | 8 | 17 | 18 |
| St_APC | 13 | 0 | 0 | 1 | 0 | 0 |

Table S32. cDC1 and cDC2 versus TC – Topological Orientation Sample

| ID Number | Age Group | Sex | Duration | Biopsy Site |
|---|---|---|---|---|
| IBM_DC1 | 51-60 | F | >24 months | L VL |

L- left; VL – vastus lateralis

Table S33. Interactome Computation – ZNF683+IL7R+TRM to Targets

| source | target | ligand.complex | receptor.complex | aggregate_rank |
|---|---|---|---|---|
| ZNF683+IL7R+TRM | cDC1 | XCL1 | XCR1 | 5.19E-07 |
| ZNF683+IL7R+TRM | M1/2 | IFNG | IFNGR1_IFNGR2 | 0.009717502 |
| ZNF683+IL7R+TRM | cDC2 | IFNG | IFNGR1_IFNGR2 | 0.009717502 |
| ZNF683+IL7R+TRM | M1 | IFNG | IFNGR1_IFNGR2 | 0.009717502 |
| ZNF683+IL7R+TRM | CD2+DC | IFNG | IFNGR1_IFNGR2 | 0.018713815 |
| ZNF683+IL7R+TRM | cDC1 | IFNG | IFNGR1_IFNGR2 | 0.021427406 |
| ZNF683+IL7R+TRM | M2 | IFNG | IFNGR1_IFNGR2 | 0.03482732 |
| ZNF683+IL7R+TRM | CD2+DC | FASLG | TNFRSF1A | 0.130488769 |
| ZNF683+IL7R+TRM | M1 | FASLG | TNFRSF10B | 0.130488769 |
| ZNF683+IL7R+TRM | M2 | FASLG | TNFRSF1A | 0.130488769 |
| ZNF683+IL7R+TRM | M1/2 | FASLG | TNFRSF1A | 0.178806511 |
| ZNF683+IL7R+TRM | M2 | FASLG | TNFRSF10B | 0.242123994 |
| ZNF683+IL7R+TRM | cDC1 | FASLG | TNFRSF10B | 0.252732558 |
| ZNF683+IL7R+TRM | CD2+DC | FASLG | TNFRSF10B | 0.291411609 |
| ZNF683+IL7R+TRM | cDC2 | FASLG | TNFRSF10B | 0.309315573 |
| ZNF683+IL7R+TRM | M1 | FASLG | TNFRSF1A | 0.34248543 |
| ZNF683+IL7R+TRM | M1/2 | FASLG | TNFRSF10B | 0.393373803 |
| ZNF683+IL7R+TRM | cDC2 | FASLG | TNFRSF1A | 0.54936532 |
| ZNF683+IL7R+TRM | cDC1 | FASLG | TNFRSF1A | 1 |
| ZNF683+IL7R+TRM | M1 | TNFSF10 | TNFRSF10B | 0.261625304 |
| ZNF683+IL7R+TRM | cDC2 | TNFSF10 | TNFRSF10A | 0.439675946 |
| ZNF683+IL7R+TRM | CD2+DC | TNFSF10 | TNFRSF10B | 0.897741929 |
| ZNF683+IL7R+TRM | cDC1 | TNFSF10 | TNFRSF10B | 1 |
| ZNF683+IL7R+TRM | cDC2 | TNFSF10 | TNFRSF10B | 1 |
| ZNF683+IL7R+TRM | M2 | TNFSF10 | TNFRSF10B | 1 |
| ZNF683+IL7R+TRM | M1/2 | TNFSF10 | TNFRSF10B | 1 |
| ZNF683+IL7R+TRM | M1 | TNFSF10 | TNFRSF10A | 1 |
| ZNF683+IL7R+TRM | CD2+DC | TNFSF10 | TNFRSF10A | 1 |
| ZNF683+IL7R+TRM | M1/2 | TNFSF10 | TNFRSF10A | 1 |
| ZNF683+IL7R+TRM | MyoS | CCL5 | CCR5 | 0.000973528 |
| ZNF683+IL7R+TRM | MyoF | CCL5 | CCR5 | 0.086291215 |
| ZNF683+IL7R+TRM | MyoR | CCL5 | CCR5 | 0.130488769 |

| | | | | |
|---|---|---|---|---|
| ZNF683+IL7R+TRM | FAP | IFNG | IFNGR1_IFNGR2 | 0.130488769 |
| ZNF683+IL7R+TRM | EC | IFNG | IFNGR1_IFNGR2 | 0.130488769 |
| ZNF683+IL7R+TRM | EC | FASLG | TNFRSF10B | 0.13301279 |
| ZNF683+IL7R+TRM | MyoR | FASLG | TNFRSF10B | 0.209607129 |
| ZNF683+IL7R+TRM | PC | IFNG | IFNGR1_IFNGR2 | 0.249131878 |
| ZNF683+IL7R+TRM | MyoS | IFNG | IFNGR1_IFNGR2 | 0.350214123 |
| ZNF683+IL7R+TRM | MyoF | IFNG | IFNGR1_IFNGR2 | 0.369168512 |
| ZNF683+IL7R+TRM | MuSC | FASLG | TNFRSF10B | 0.413003344 |
| ZNF683+IL7R+TRM | MyoR | IFNG | IFNGR1_IFNGR2 | 0.511023144 |
| ZNF683+IL7R+TRM | FAP | FASLG | TNFRSF10B | 0.629195118 |
| ZNF683+IL7R+TRM | MyoF | FASLG | TNFRSF10B | 1 |
| ZNF683+IL7R+TRM | MyoS | FASLG | TNFRSF10B | 1 |

Table S34. Interactome Computation – Other Nuclei Types to ZNF683+IL7R+TRM

| source | target | ligand.complex | receptor.complex | aggregate_rank |
|---|---|---|---|---|
| M1 | ZNF683+IL7R+TRM | IL15 | IL15RA_IL2RB_IL2RG | 0.009717502 |
| cDC2 | ZNF683+IL7R+TRM | IL15 | IL15RA_IL2RB_IL2RG | 0.009717502 |
| M1 | ZNF683+IL7R+TRM | TNFSF10 | TNFRSF10A | 0.010357903 |
| M1/2 | ZNF683+IL7R+TRM | IL15 | IL15RA_IL2RB_IL2RG | 0.028789729 |
| M1 | ZNF683+IL7R+TRM | CXCL11 | CXCR3 | 0.028830292 |
| M1 | ZNF683+IL7R+TRM | IL7 | IL2RG_IL7R | 0.03023518 |
| CD2+DC | ZNF683+IL7R+TRM | IL15 | IL15RA_IL2RB_IL2RG | 0.045137054 |
| M2 | ZNF683+IL7R+TRM | IL15 | IL15RA_IL2RB_IL2RG | 0.051102973 |
| cDC1 | ZNF683+IL7R+TRM | IL15 | IL15RA_IL2RB_IL2RG | 0.058905576 |
| M2 | ZNF683+IL7R+TRM | CD59 | CD2 | 0.062008858 |
| cDC2 | ZNF683+IL7R+TRM | IL7 | IL2RG_IL7R | 0.065895131 |
| M1/2 | ZNF683+IL7R+TRM | IL7 | IL2RG_IL7R | 0.075173697 |
| CD2+DC | ZNF683+IL7R+TRM | IL7 | IL2RG_IL7R | 0.091357068 |
| M2 | ZNF683+IL7R+TRM | IL7 | IL2RG_IL7R | 0.109534077 |
| cDC1 | ZNF683+IL7R+TRM | IL7 | IL2RG_IL7R | 0.120896427 |
| CD2+DC | ZNF683+IL7R+TRM | CD59 | CD2 | 0.122068064 |
| M2 | ZNF683+IL7R+TRM | CCL13 | CCR5 | 0.130488769 |
| M2 | ZNF683+IL7R+TRM | CCL2 | CCR5 | 0.130488769 |
| M1/2 | ZNF683+IL7R+TRM | TNFSF10 | TNFRSF10A | 0.130488769 |
| cDC2 | ZNF683+IL7R+TRM | TNFSF10 | TNFRSF10A | 0.130488769 |
| M1 | ZNF683+IL7R+TRM | CCL2 | CCR5 | 0.151129274 |
| CD2+DC | ZNF683+IL7R+TRM | CCL13 | CCR5 | 0.185983084 |
| M1/2 | ZNF683+IL7R+TRM | CCL2 | CCR5 | 0.193657999 |
| CD2+DC | ZNF683+IL7R+TRM | TNFSF10 | TNFRSF10A | 0.213745021 |
| CD2+DC | ZNF683+IL7R+TRM | CCL2 | CCR5 | 0.344750929 |
| cDC1 | ZNF683+IL7R+TRM | TNFSF10 | TNFRSF10A | 1 |
| M2 | ZNF683+IL7R+TRM | TNFSF10 | TNFRSF10A | 1 |
| FAP | ZNF683+IL7R+TRM | C3 | CD46 | 0.009717502 |
| EC | ZNF683+IL7R+TRM | CD58 | CD2 | 0.067438074 |
| MuSC | ZNF683+IL7R+TRM | CD58 | CD2 | 0.130488769 |
| FAP | ZNF683+IL7R+TRM | CD58 | CD2 | 0.130488769 |
| PC | ZNF683+IL7R+TRM | CD58 | CD2 | 0.130488769 |

| | | | | |
|---|---|---|---|---|
| MyoR | ZNF683+IL7R+TRM | CD58 | CD2 | 0.130488769 |
| MyoS | ZNF683+IL7R+TRM | CD58 | CD2 | 0.212778986 |
| MyoF | ZNF683+IL7R+TRM | CD58 | CD2 | 0.38178019 |
| EC | ZNF683+IL7R+TRM | CD59 | CD2 | 0.000186355 |
| MyoS | ZNF683+IL7R+TRM | CD59 | CD2 | 0.009717502 |
| PC | ZNF683+IL7R+TRM | CD59 | CD2 | 0.009717502 |
| MyoF | ZNF683+IL7R+TRM | CD59 | CD2 | 0.009717502 |
| FAP | ZNF683+IL7R+TRM | CD59 | CD2 | 0.020062971 |
| MyoR | ZNF683+IL7R+TRM | CD59 | CD2 | 0.036134929 |
| EC | ZNF683+IL7R+TRM | CXCL12 | CXCR4 | 0.130488769 |
| FAP | ZNF683+IL7R+TRM | CXCL12 | CXCR4 | 0.137252455 |
| PC | ZNF683+IL7R+TRM | CXCL12 | CXCR4 | 0.235137609 |
| EC | ZNF683+IL7R+TRM | HLA-E | KLRK1 | 0.000661254 |
| FAP | ZNF683+IL7R+TRM | HLA-E | KLRK1 | 0.009717502 |
| MuSC | ZNF683+IL7R+TRM | HLA-E | KLRK1 | 0.01304345 |
| PC | ZNF683+IL7R+TRM | HLA-E | KLRK1 | 0.024644219 |
| MyoF | ZNF683+IL7R+TRM | HLA-E | KLRK1 | 0.121575164 |
| MyoR | ZNF683+IL7R+TRM | HLA-E | KLRK1 | 0.130488769 |
| MyoS | ZNF683+IL7R+TRM | HLA-E | KLRK1 | 0.2207294 |
| MyoS | ZNF683+IL7R+TRM | HMGB1 | CXCR4 | 0.119705211 |
| MyoF | ZNF683+IL7R+TRM | HMGB1 | CXCR4 | 0.212896713 |
| MyoR | ZNF683+IL7R+TRM | HMGB1 | CXCR4 | 0.278251841 |
| FAP | ZNF683+IL7R+TRM | HMGB1 | CXCR4 | 0.287725777 |
| MuSC | ZNF683+IL7R+TRM | HMGB1 | CXCR4 | 0.335508575 |
| PC | ZNF683+IL7R+TRM | HMGB1 | CXCR4 | 1 |
| EC | ZNF683+IL7R+TRM | HMGB1 | CXCR4 | 1 |
| EC | ZNF683+IL7R+TRM | IL15 | IL15RA_IL2RB_IL2RG | 0.124778107 |
| MyoF | ZNF683+IL7R+TRM | IL15 | IL15RA_IL2RB_IL2RG | 0.309826529 |
| MyoS | ZNF683+IL7R+TRM | IL15 | IL15RA_IL2RB_IL2RG | 0.467468965 |
| MuSC | ZNF683+IL7R+TRM | IL15 | IL15RA_IL2RB_IL2RG | 0.483673254 |
| FAP | ZNF683+IL7R+TRM | IL15 | IL15RA_IL2RB_IL2RG | 0.585895288 |
| MyoR | ZNF683+IL7R+TRM | IL15 | IL15RA_IL2RB_IL2RG | 0.585895288 |
| PC | ZNF683+IL7R+TRM | IL15 | IL15RA_IL2RB_IL2RG | 1 |
| FAP | ZNF683+IL7R+TRM | IL7 | IL2RG_IL7R | 0.017892911 |
| MuSC | ZNF683+IL7R+TRM | IL7 | IL2RG_IL7R | 0.048933995 |

| | | | | |
|---|---|---|---|---|
| PC | ZNF683+IL7R+TRM | IL7 | IL2RG_IL7R | 0.425529169 |
| EC | ZNF683+IL7R+TRM | IL7 | IL2RG_IL7R | 0.549310963 |
| MyoR | ZNF683+IL7R+TRM | IL7 | IL2RG_IL7R | 0.862483828 |
| MyoF | ZNF683+IL7R+TRM | IL7 | IL2RG_IL7R | 1 |
| EC | ZNF683+IL7R+TRM | TNFSF10 | TNFRSF10A | 0.075935419 |
| FAP | ZNF683+IL7R+TRM | TNFSF10 | TNFRSF10A | 0.5133941 |

The HLA-E–KLRK1 edge is best viewed as an inferred network-level interaction reflecting modulation of ZNF683$^{+}$ IL7R$^{+}$ TRM within the IBM microenvironment. Together with MICA/KLRK1 signaling and IFN-γ-driven HLA-E expression, it supports a model in which resident T cells are functionally remodeled toward a persistent cytokine/chemokine-producing surveillance state while retaining KLRK1 expression, rather than undergoing classical exhaustion or complete loss of cytotoxic capability.

Table S35. CXCL9-CXCR3 Custom Interaction Computation

(Non-Native to R package interaction dataset calibrated using native interaction dataset IFNG and FASLG interactions)

| source | target | ligand.complex | receptor.complex | weight_expression (cell-type specificity) | weight_cells (tissue impact) |
|---|---|---|---|---|---|
| M2 | ZNF683+IL7R+TRM | CXCL9 | CXCR3 | 0.016201079 | 13454.70486 |
| M1 | ZNF683+IL7R+TRM | CXCL9 | CXCR3 | 0.556079605 | 9356.03935 |
| M1/2 | ZNF683+IL7R+TRM | CXCL9 | CXCR3 | 0.054351513 | 5450.206693 |
| cDC1 | ZNF683+IL7R+TRM | CXCL9 | CXCR3 | 0.112415303 | 4009.741447 |
| CD2+DC | ZNF683+IL7R+TRM | CXCL9 | CXCR3 | 0.044926451 | 2116.485084 |

CXCL9–CXCR3 is a canonical, experimentally established interaction present in curated consensus resources but retained here through custom computation because its transcripts fell below LIANA's expression and rank-aggregation thresholds in sparse single-nucleus data rather than through biological absence. CXCL9 is an interferon-γ-inducible ligand of CXCR3, a receptor rapidly induced on activated T cells and preferentially retained on effector $CD8^{+}$ T cells, type-1 helper T cells, and innate-type lymphocytes including natural killer and NKT cells. The axis mediates recruitment of these populations into inflamed tissue. The interaction weight was computed from the product of normalised ligand and receptor expression and scaled to the range of inferred LIANA weights using IFNG–IFNGR1/2 and FASLG–TNFRSF10A/B as reference interactions, and is flagged as supplemented throughout filtering and visualisation. Reference: Groom JR, Luster AD. CXCR3 ligands: redundant, collaborative and antagonistic functions. *Immunol Cell Biol.* 2011;89(2):207–215.

Table S36. CXCL10-CXCR3 Custom Interaction Computation

(Non-Native to R package interaction dataset calibrated using native interaction dataset IFNG and FASLG interactions)

| source | target | ligand.complex | receptor.complex | weight_expression (cell-type specificity) | weight_cells (tissue impact) |
|---|---|---|---|---|---|
| M2 | ZNF683+IL7R+TRM | CXCL10 | CXCR3 | 0.082509823 | 68522.9228 |
| M1 | ZNF683+IL7R+TRM | CXCL10 | CXCR3 | 3.582561404 | 60276.59563 |
| M1/2 | ZNF683+IL7R+TRM | CXCL10 | CXCR3 | 0.147862054 | 14827.16315 |
| cDC1 | ZNF683+IL7R+TRM | CXCL10 | CXCR3 | 0.283665972 | 13363.50394 |
| CD2+DC | ZNF683+IL7R+TRM | CXCL10 | CXCR3 | 0.086619018 | 3089.613766 |

CXCL10–CXCR3 is a canonical, experimentally established interaction present in curated consensus resources but retained here through custom computation because its transcripts fell below LIANA's expression and rank-aggregation thresholds in sparse single-nucleus data rather than through biological absence. CXCL10 (IP-10) is an interferon-γ-inducible CXCR3 ligand. In immune-mediated tissue destruction, infiltrating $CD8^+$ cytotoxic T cells express CXCR3 and store CXCL10 within their cytolytic granules, supporting a self-recruiting effector-cell mechanism conceptually parallel to the retention niche described in this study. The interaction weight was computed from the product of normalised ligand and receptor expression and scaled to the range of inferred LIANA weights using IFNG–IFNGR1/2 and FASLG–TNFRSF10A/B as reference interactions, and is flagged as supplemented throughout filtering and visualisation. Reference: Iijima W, Ohtani H, Nakayama T, et al. Infiltrating $CD8^+$ T cells in oral lichen planus predominantly express CCR5 and CXCR3 and carry respective chemokine ligands RANTES/CCL5 and IP-10/CXCL10 in their cytolytic granules: a potential self-recruiting mechanism. *Am J Pathol.* 2003;163(1):261–268.

Table S37. CCL18-CCR8 Custom Interaction Computation

(Non-Native to R package interaction dataset calibrated using native interaction dataset IFNG and FASLG interactions)

| source | target | ligand.complex | receptor.complex | weight_expression (cell-type specificity) | weight_cells (tissue impact) |
|---|---|---|---|---|---|
| M2 | ZNF683+IL7R+TRM | CCL18 | CCR8 | 0.005 | 4164.857 |
| CD2+DC | ZNF683+IL7R+TRM | CCL18 | CCR8 | 0.003 | 125.556 |
| M1/2 | ZNF683+IL7R+TRM | CCL18 | CCR8 | 0.001 | 106.253 |
| M1 | ZNF683+IL7R+TRM | CCL18 | CCR8 | 0.001 | 12.193 |
| cDC1 | ZNF683+IL7R+TRM | CCL18 | CCR8 | 0.0002 | 7.330 |
| cDC2 | ZNF683+IL7R+TRM | CCL18 | CCR8 | 0 | 0 |

CCL18–CCR8 is an experimentally established interaction present in curated consensus resources but retained here through custom computation because its transcripts fell below LIANA's expression and rank-aggregation thresholds in sparse single-nucleus data rather than through biological absence. CCL18 is a chemokine produced predominantly by alternatively activated (M2) macrophages and is among the most highly expressed chemokines in human chronic inflammatory disease. Human CCR8 has been identified as a functional CCL18 receptor mediating chemotaxis and calcium flux, consistent with the M2-dominant antigen-presenting-cell compartment observed in IBM. The receptor assignment for CCL18 has been debated in the literature, and CCR8 represents the most established G-protein-coupled receptor candidate. The interaction weight was computed from the product of normalised ligand and receptor expression and scaled to the range of inferred LIANA weights using IFNG–IFNGR1/2 and FASLG–TNFRSF10A/B as reference interactions, and is flagged as supplemented throughout filtering and visualisation. Reference: Islam SA, Ling MF, Leung J, Shreffler WG, Luster AD. Identification of human CCR8 as a CCL18 receptor. *J Exp Med.* 2013;210(10):1889–1898.

Table S38. CCL5-CCR5 Custom Interaction Computation
(Non-Native to R package interaction dataset calibrated using native interaction dataset IFNG and FASLG interactions)

| source | target | ligand.complex | receptor.complex | weight_expression (cell-type specificity) | weight_cells (tissue impact) |
|---|---|---|---|---|---|
| FAP | ZNF683+IL7R+TRM | CCL5 | CCR5 | 0.058972 | 62865.54 |
| MyoR | ZNF683+IL7R+TRM | CCL5 | CCR5 | 0.044209 | 61499.02 |
| MyoF | ZNF683+IL7R+TRM | CCL5 | CCR5 | 0.013326 | 33065.48 |
| EC | ZNF683+IL7R+TRM | CCL5 | CCR5 | 0.036805 | 29277.79 |
| MyoS | ZNF683+IL7R+TRM | CCL5 | CCR5 | 0.008762 | 18067.28 |
| PC | ZNF683+IL7R+TRM | CCL5 | CCR5 | 0.022156 | 11675.5 |
| MuSC | ZNF683+IL7R+TRM | CCL5 | CCR5 | 0.017104 | 10348.13 |

CCL5–CCR5 is a canonical, experimentally established interaction present in curated consensus resources but retained here through custom computation because its transcripts fell below LIANA's expression and rank-aggregation thresholds in sparse single-nucleus data rather than through biological absence. CCL5 (RANTES) signals through CCR5 to recruit cytotoxic $CD8^+$ T cells to sites of inflammation. Tissue-infiltrating $CD8^+$ T cells express CCR5 and store CCL5 within their cytolytic granules, establishing a self-recruiting circuit for effector cytotoxic T cells in chronic immune-mediated tissue injury. The interaction weight was computed from the product of normalised ligand and receptor expression and scaled to the range of inferred LIANA weights using IFNG–IFNGR1/2 and FASLG–TNFRSF10A/B as reference interactions, and is flagged as supplemented throughout filtering and visualisation. Reference: Iijima W, Ohtani H, Nakayama T, et al. Infiltrating $CD8^+$ T cells in oral lichen planus predominantly express CCR5 and CXCR3 and carry respective chemokine ligands RANTES/CCL5 and IP-10/CXCL10 in their cytolytic granules: a potential self-recruiting mechanism. *Am J Pathol.* 2003;163(1):261–268.

Table S39. Upregulated genes in snRNAseq and bulk RNAseq associated with CD1d-ligand loading for iNKT/NKT TCR activation and non-classical MHC-I signaling.

| gene | log2FC | padj | category | lipid_or_process | dataset_label |
|---|---|---|---|---|---|
| *ANXA1* | 1.618961373 | 2.48E-156 | Phagocytic uptake signaling | Annexin-mediated efferocytosis | snRNAseq MyoF |
| *ANXA1* | 1.20016995 | 5E-324 | Phagocytic uptake signaling | Annexin-mediated efferocytosis | snRNAseq MyoR |
| *ANXA1* | 2.056501044 | 5.25E-14 | Phagocytic uptake signaling | Annexin-mediated efferocytosis | bulk RNAseq |
| *GM2A* | 1.337744692 | 7.18E-90 | Lysosomal CD1 lipid-loading support | GM2 activator protein / glycolipid transfer | snRNAseq MyoF |
| *GM2A* | 1.281551555 | 7.67E-07 | Lysosomal CD1 lipid-loading support | GM2 activator protein / glycolipid transfer | bulk RNAseq |
| *PLA2G4A* | 1.517196885 | 5.34E-14 | Lysophospholipid pool generation and lipid oxidation substrate release | LPC / arachidonic-acid / oxidizable phospholipid pool | snRNAseq MyoF |
| *PLA2G4A* | 2.368967539 | 9.37E-10 | Lysophospholipid pool generation and lipid oxidation substrate release | LPC / arachidonic-acid / oxidizable phospholipid pool | bulk RNAseq |
| *HLA-E* | 1.541322880 | 6.35E-24 | Modulator of cytopathic activity of T cells in expressing tissues | Expression increased downstream of IFN- γ signaling | snRNAseq MyoF |
| *HLA-E* | 2.624202802 | 9.45E-12 | Modulator of cytopathic activity of T cells in expressing tissues | Expression increased downstream of IFN- γ signaling | bulk RNAseq |

Table S40. Public Domain Data Information with Spatial Transcriptomics

h5ad data from Single-nucleus RNA sequencing and spatial transcriptomics decode cell type-specific drivers underlying IBM pathogenesis

https://explore.data.humancellatlas.org/projects/d5c91e92-2e7f-473d-8cf3-ab03bbae21c2

Clinical Information from: Cell type mapping of inflammatory muscle diseases highlights selective myofiber vulnerability in inclusion body myositis

https://www.nature.com/articles/s43587-024-00645-9#Sec23

https://static-content.springer.com/esm/art%3A10.1038%2Fs43587-024-00645-9/MediaObjects/43587_2024_645_MOESM2_ESM.xlsx

| Patient Number | Condition | Age Group | Sex | Biopsy Site |
|---|---|---|---|---|
| CTRL 47 | Non-inflammatory control | 51-60 | M | Quadriceps |
| CTRL 50 | Non-inflammatory control | 61-70 | M | Quadriceps |
| IMNM 1 | Immune mediated necrotizing myopathy | 41-50 | M | Quadriceps |
| IMNM 4 | Immune mediated necrotizing myopathy | 61-70 | F | Quadriceps |
| IBM 29 | Inclusion body myositis | 51-60 | M | Quadriceps |
| IBM 35 | Inclusion body myositis | 71-80 | F | Quadriceps |

Table S41. Distribution of Nuclei from Spatial Transcriptomics Data

| Condition | SuperGroup | CellCategory | Gene Signature Criteria and expression level | n |
|---|---|---|---|---|
| IBM | Myonuclei / MuSC | MuSC | *PAX7*>0, *NCAM1*>0 | 72 |
| IBM | Myonuclei / MuSC | MyoS+MyoR | *MYH7*>1, *MYH3*>1, *MYH8*>1 | 335 |
| IBM | Myonuclei / MuSC | MyoS | *MYH7*>1 | 167 |
| IBM | Myonuclei / MuSC | MyoF+MyoR | *MYH1*>1 or *MYH2*>1, *MYH3*>1, *MYH8*>1 | 664 |
| IBM | Myonuclei / MuSC | MyoF_2A | *MYH2*>1 | 151 |
| IBM | Myonuclei / MuSC | MyoF_2X | *MYH1*>1 | 93 |
| IBM | Immune | ZNF683+TRM | *ZNF683*>1, *CD247*>1, *ZBTB16*<0.5 | 266 |
| IBM | Immune | iNKT-like_TC | *ZBTB16*>1, *CD247*>1, *ZNF683* < 0.5 | 28 |
| IBM | Immune | cDC1 | *XCR1*>1 | 109 |
| IBM | Immune | cDC2 | *FLT3*>0, *CD1C*>1 | 9 |
| IBM | Immune | M2 | *MRC1*>1, *MSR1*>1 | 290 |
| IBM | Immune | M1 | *CD80*>0, *CD86*>0 | 1 |
| IBM | Immune | CD2+DC | *CD2*>1, *CD14*>1, *MRC1*>1 | 361 |
| IBM | Interstitial | FAP/MSC | *PDGFRA*>1 | 527 |
| IBM | Interstitial | EC | *VWF*>1, *PECAM1*>1 | 224 |
| IBM | Interstitial | PC | *PDGFRB*>1 | 161 |
| IMNM | Myonuclei / MuSC | MuSC | *PAX7*>0, *NCAM1*>0 | 9 |
| IMNM | Myonuclei / MuSC | MyoS+MyoR | *MYH7*>1, *MYH3*>1, *MYH8*>1 | 45 |
| IMNM | Myonuclei / MuSC | MyoS | *MYH7*>1 | 69 |
| IMNM | Myonuclei / MuSC | MyoF+MyoR | *MYH1*>1 or *MYH2*>1, *MYH3*>1, *MYH8*>1 | 455 |
| IMNM | Myonuclei / MuSC | MyoF_2A | *MYH2*>1 | 304 |
| IMNM | Myonuclei / MuSC | MyoF_2X | *MYH1*>1 | 184 |
| IMNM | Immune | M2 | *MRC1*>1, *MSR1*>1 | 12 |
| IMNM | Interstitial | FAP/MSC | *PDGFRA*>1 | 287 |
| IMNM | Interstitial | EC | *VWF*>1, *PECAM1*>1 | 92 |
| IMNM | Interstitial | PC | *PDGFRB*>1 | 41 |
| CTRL | Myonuclei / MuSC | MuSC | *PAX7*>0, *NCAM1*>0 | 3 |
| CTRL | Myonuclei / MuSC | MyoS+MyoR | *MYH7*>1, *MYH3*>1, *MYH8*>1 | 36 |

| | | | | |
|---|---|---|---|---|
| CTRL | Myonuclei / MuSC | MyoS | *MYH7*>1 | 932 |
| CTRL | Myonuclei / MuSC | MyoF+MyoR | *MYH1*>1 or *MYH2*>1, *MYH3*>1, *MYH8*>1 | 34 |
| CTRL | Myonuclei / MuSC | MyoF_2A | *MYH2*>1 | 460 |
| CTRL | Myonuclei / MuSC | MyoF_2X | *MYH1*>1 | 250 |
| CTRL | Immune | M2 | *MRC1*>1, *MSR1*>1 | 1 |
| CTRL | Interstitial | FAP/MSC | *PDGFRA*>1 | 100 |
| CTRL | Interstitial | EC | *VWF*>1, *PECAM1*>1 | 8 |
| CTRL | Interstitial | PC | *PDGFRB*>1 | 18 |

Note: The ZNF683+TRM population in this table corresponds to the ZNF683+IL7R+TRM population described in the main text and in Table S6–9 and Table S33–34. *IL7R* was not used as a classification criterion for spatial transcriptomics analysis because transcript detection in spatial data reflects both nuclear and cytoplasmic RNA compartments, and classification was based on *ZNF683* and *CD247* co-expression with *ZBTB16* exclusion to maintain consistency with the snRNA-seq-derived gene signature criteria (see also Table S42 legend).

Table S42. Proportion of Myonuclei in IBM expressing *HMGB1* and *G3BP1* log-normalized expression value > 0

| Nuclei Type | Count_all nuclei in Subtype | Count_target nuclei with gene only (HMGB1 and G3BP1 only) | Proportion vs. all myonuclei (Prop_all) | Proportion of nuclei with target gene only (Prop_target) | Enrichment (Prop_target / Prop_all) | PosRate (Count_target / Count_all) |
|---|---|---|---|---|---|---|
| MyoS+MyoR | 1118 | 348 | 0.294987 | 0.376623 | 1.28 | 0.311 |
| MyoS | 260 | 28 | 0.068602 | 0.030303 | 0.44 | 0.108 |
| MyoF+MyoR | 2050 | 525 | 0.540897 | 0.568182 | 1.05 | 0.256 |
| MyoF_2A | 236 | 17 | 0.062269 | 0.018398 | 0.3 | 0.072 |
| MyoF_2X | 126 | 6 | 0.033245 | 0.006494 | 0.2 | 0.048 |

Interpretation of Enrichment if ≥ 1 – enriched , if < 1 – depleted

Interpretation of PosRate – higher score indicates more prone to stress/regeneration program

Note: Count_all values in this table reflect nuclei pooled across all IBM spatial transcriptomics samples per condition. Values in this table will therefore exceed per-sample counts reported in Table S41.

Table S43. Proportion of FAP/MSC in IBM (n=527) expressing *HMGB1*, *CXCL12*, and *G3BP1* log-normalized expression value > 0

| Nuclei Type | Count_all nuclei in Subtype | Count_target nuclei with combination defined per row | Proportion vs. all myonuclei (Prop_all) | Proportion of nuclei with target gene only (Prop_target) | Enrichment (Prop_target / Prop_all) | PosRate (Count_target / Count_all) |
|---|---|---|---|---|---|---|
| FAP_HC_G (HMGB1+ CXCL12+ G3BP1+) | 310 | 310 | 0.588235 | 1 | 1.7 | 1 |
| FAP_H_G (HMGB1+ G3BP1+) | 7 | 0 | 0.013283 | 0 | 0 | 0 |
| FAP_CX_G (CXCL12+ G3BP1+) | 53 | 0 | 0.100569 | 0 | 0 | 0 |

Interpretation of Enrichment if ≥ 1 – enriched , if < 1 – depleted

Interpretation of PosRate – higher score indicates more prone to stress/regeneration program

Note: The total FAP/MSC count stated in the table title (n = 527) reflects all *PDGFRA* > 1 nuclei identified in the IBM spatial transcriptomics dataset. The three rows shown represent mutually exclusive marker co-expression subgroups (FAP_HC_G: HMGB1+CXCL12+G3BP1+, n = 310; FAP_H_G: HMGB1+G3BP1+ only, n = 7; FAP_CX_G: CXCL12+G3BP1+ only, n = 53; sum = 370). The remaining 157 nuclei (527 − 370) are FAP/MSC nuclei expressing none of the listed marker combinations (HMGB1−/CXCL12−/G3BP1−) and are excluded from this table as they do not contribute to the inflammatory stromal signal described in the main text.

### Table S44. Quantification of Spatial TRM-Associated Stress Microdomains in IBM Muscle

| hex_id | n_cells | n_TRM | frac_stress | frac_FAP | niche_TRM_ x_stress | niche_TRM_ x_stress_x_FAP | hex_area_um2 | spatial_1 | spatial_2 | in_TRM_ KDE_hotspot | in_TRM_ KDE_core | TRM_hotspot_ proximity |
|---|---|---|---|---|---|---|---|---|---|---|---|---|
| 1527 | 2 | 1 | 1 | 1 | 1 | 1 | 5607.6 | 4501 | 3201 | 0 | 0 | 0.498 |
| 1950 | 2 | 1 | 1 | 1 | 1 | 1 | 5607.6 | 2184 | 3731 | 0 | 0 | 0.292 |
| 115 | 1 | 1 | 1 | 1 | 1 | 1 | 5607.6 | 3723 | 1754.5 | 0 | 0 | 0.11 |
| 525 | 1 | 1 | 1 | 1 | 1 | 1 | 5607.6 | 3492 | 2278 | 0 | 0 | 0.184 |
| 735 | 1 | 1 | 1 | 1 | 1 | 1 | 5607.6 | 3551.3333 | 2472 | 0 | 0 | 0.192 |
| 945 | 1 | 1 | 1 | 1 | 1 | 1 | 5607.6 | 2630.5 | 2682 | 0 | 0 | 0.34 |
| 1559 | 1 | 1 | 1 | 1 | 1 | 1 | 5607.6 | 3309.5 | 3239 | 1 | 0 | 0.654 |
| 1690 | 1 | 1 | 1 | 1 | 1 | 1 | 5607.6 | 1585 | 3420 | 1 | 1 | 0.684 |
| 1757 | 1 | 1 | 1 | 1 | 1 | 1 | 5607.6 | 4125 | 3441 | 1 | 1 | 0.74 |
| 1864 | 1 | 1 | 1 | 1 | 1 | 1 | 5607.6 | 2714 | 3619.5 | 0 | 0 | 0.194 |
| 2301 | 1 | 1 | 1 | 1 | 1 | 1 | 5607.6 | 1554 | 4206 | 0 | 0 | 0.457 |
| 2591 | 1 | 1 | 1 | 1 | 1 | 1 | 5607.6 | 2225 | 4700 | 0 | 0 | 0.154 |
| 2747 | 1 | 1 | 1 | 1 | 1 | 1 | 5607.6 | 2758 | 4977 | 0 | 0 | 0.225 |
| 1596 | 3 | 1 | 1 | 0.6667 | 1 | 0.6667 | 5607.6 | 2035 | 3288 | 1 | 1 | 0.852 |
| 1808 | 3 | 2 | 1 | 0.3333 | 2 | 0.6667 | 5607.6 | 4772 | 3500 | 1 | 0 | 0.641 |
| 1017 | 2 | 1 | 1 | 0.5 | 1 | 0.5 | 5607.6 | 3708 | 2745 | 0 | 0 | 0.306 |
| 1120 | 2 | 1 | 1 | 0.5 | 1 | 0.5 | 5607.6 | 3097.6667 | 2838.66667 | 0 | 0 | 0.268 |
| 1384 | 2 | 1 | 1 | 0.5 | 1 | 0.5 | 5607.6 | 1376 | 3093 | 0 | 0 | 0.415 |
| 1878 | 2 | 1 | 1 | 0.5 | 1 | 0.5 | 5607.6 | 3816 | 3586 | 1 | 1 | 0.737 |
| 2088 | 2 | 1 | 1 | 0.5 | 1 | 0.5 | 5607.6 | 3765 | 3846 | 1 | 1 | 0.849 |
| 2231 | 2 | 1 | 1 | 0.5 | 1 | 0.5 | 5607.6 | 4343 | 4049 | 1 | 0 | 0.52 |
| 2319 | 2 | 1 | 1 | 0.5 | 1 | 0.5 | 5607.6 | 4090 | 4209 | 0 | 0 | 0.497 |
| 2395 | 2 | 1 | 1 | 0.5 | 1 | 0.5 | 5607.6 | 2213 | 4402 | 0 | 0 | 0.153 |
| 1803 | 2 | 1 | 0.5 | 1 | 0.5 | 0.5 | 5607.6 | 4388 | 3520 | 1 | 1 | 0.722 |
| 812 | 3 | 1 | 0.6667 | 0.6667 | 0.6667 | 0.4444 | 5607.6 | 5106 | 2526 | 0 | 0 | 0.311 |
| 1173 | 3 | 1 | 0.6667 | 0.6667 | 0.6667 | 0.4444 | 5607.6 | 2790 | 2884.5 | 0 | 0 | 0.263 |
| 1387 | 3 | 2 | 0.6667 | 0.3333 | 1.3333 | 0.4444 | 5607.6 | 1766 | 3076 | 1 | 1 | 0.768 |
| 1880 | 3 | 2 | 0.6667 | 0.3333 | 1.3333 | 0.4444 | 5607.6 | 3943 | 3579 | 1 | 1 | 0.773 |
| 1665 | 3 | 1 | 1 | 0.3333 | 1 | 0.3333 | 5607.6 | 3927 | 3339.5 | 1 | 1 | 0.686 |
| 1319 | 2 | 1 | 0.5 | 0.5 | 0.5 | 0.25 | 5607.6 | 5274 | 3000 | 0 | 0 | 0.334 |

| | | | | | | | | | | | | |
|---|---|---|---|---|---|---|---|---|---|---|---|---|
| 1553 | 2 | 1 | 0.5 | 0.5 | 0.5 | 0.25 | 5607.6 | 2837 | 3231 | 0 | 0 | 0.264 |
| 1618 | 2 | 1 | 0.5 | 0.5 | 0.5 | 0.25 | 5607.6 | 3855 | 3285 | 1 | 0 | 0.641 |
| 2017 | 2 | 1 | 0.5 | 0.5 | 0.5 | 0.25 | 5607.6 | 4269.5 | 3768.5 | 1 | 1 | 0.835 |
| 2021 | 2 | 1 | 0.5 | 0.5 | 0.5 | 0.25 | 5607.6 | 4527 | 3779 | 1 | 1 | 0.877 |
| 2086 | 2 | 1 | 0.5 | 0.5 | 0.5 | 0.25 | 5607.6 | 3637 | 3853 | 1 | 1 | 0.765 |
| 2100 | 2 | 1 | 0.5 | 0.5 | 0.5 | 0.25 | 5607.6 | 826 | 3901 | 0 | 0 | 0.181 |
| 2108 | 2 | 1 | 0.5 | 0.5 | 0.5 | 0.25 | 5607.6 | 1671 | 3902 | 0 | 0 | 0.359 |
| 2160 | 2 | 1 | 0.5 | 0.5 | 0.5 | 0.25 | 5607.6 | 3763.5 | 3977.5 | 1 | 1 | 0.894 |
| 2322 | 2 | 1 | 0.5 | 0.5 | 0.5 | 0.25 | 5607.6 | 4416 | 4233 | 0 | 0 | 0.396 |
| 2324 | 2 | 1 | 0.5 | 0.5 | 0.5 | 0.25 | 5607.6 | 4676 | 4221 | 0 | 0 | 0.23 |
| 2402 | 2 | 1 | 0.5 | 0.5 | 0.5 | 0.25 | 5607.6 | 3188 | 4397 | 1 | 0 | 0.591 |
| 2609 | 2 | 1 | 0.5 | 0.5 | 0.5 | 0.25 | 5607.6 | 3786 | 4708 | 1 | 1 | 0.796 |
| 891 | 3 | 1 | 0.6667 | 0.3333 | 0.6667 | 0.2222 | 5607.6 | 2820 | 2635.5 | 0 | 0 | 0.292 |
| 1032 | 3 | 1 | 0.3333 | 0.6667 | 0.3333 | 0.2222 | 5607.6 | 4742 | 2724 | 0 | 0 | 0.333 |
| 1244 | 3 | 2 | 0.3333 | 0.3333 | 0.6667 | 0.2222 | 5607.6 | 3779 | 2938.5 | 1 | 0 | 0.575 |
| 1320 | 3 | 1 | 0.3333 | 0.3333 | 0.3333 | 0.1111 | 5607.6 | 5337 | 2960 | 0 | 0 | 0.312 |
| 1453 | 3 | 1 | 0.3333 | 0.3333 | 0.3333 | 0.1111 | 5607.6 | 2961 | 3151 | 0 | 0 | 0.363 |
| 1663 | 3 | 1 | 0.3333 | 0.3333 | 0.3333 | 0.1111 | 5607.6 | 3798 | 3345.5 | 1 | 0 | 0.652 |
| 2022 | 3 | 1 | 0.3333 | 0.3333 | 0.3333 | 0.1111 | 5607.6 | 4591.5 | 3753 | 1 | 1 | 0.821 |
| 2161 | 3 | 1 | 0.3333 | 0.3333 | 0.3333 | 0.1111 | 5607.6 | 3859 | 3954.5 | 1 | 1 | 0.858 |
| 1522 | 3 | 1 | 1 | 0 | 1 | 0 | 5607.6 | 4180 | 3199 | 0 | 0 | 0.46 |
| 1332 | 2 | 1 | 1 | 0 | 1 | 0 | 5607.6 | 1959 | 3030 | 1 | 0 | 0.641 |
| 1458 | 2 | 1 | 1 | 0 | 1 | 0 | 5607.6 | 3344 | 3132 | 1 | 1 | 0.702 |
| 1875 | 2 | 1 | 1 | 0 | 1 | 0 | 5607.6 | 3617.5 | 3613.5 | 0 | 0 | 0.505 |
| 1946 | 2 | 1 | 1 | 0 | 1 | 0 | 5607.6 | 1598 | 3718 | 1 | 0 | 0.53 |
| 2259 | 2 | 1 | 1 | 0 | 1 | 0 | 5607.6 | 4087 | 4135 | 1 | 0 | 0.589 |
| 469 | 1 | 1 | 1 | 0 | 1 | 0 | 5607.6 | 3152.3333 | 2195.33333 | 0 | 0 | 0.148 |
| 811 | 1 | 1 | 1 | 0 | 1 | 0 | 5607.6 | 5049 | 2508 | 0 | 0 | 0.321 |
| 879 | 1 | 1 | 1 | 0 | 1 | 0 | 5607.6 | 1876 | 2623 | 0 | 0 | 0.254 |
| 943 | 1 | 1 | 1 | 0 | 1 | 0 | 5607.6 | 2516 | 2680 | 0 | 0 | 0.315 |
| 954 | 1 | 1 | 1 | 0 | 1 | 0 | 5607.6 | 3250 | 2667 | 0 | 0 | 0.215 |
| 1245 | 1 | 1 | 1 | 0 | 1 | 0 | 5607.6 | 3843 | 2935.5 | 1 | 0 | 0.565 |
| 1385 | 1 | 1 | 1 | 0 | 1 | 0 | 5607.6 | 1506 | 3087 | 1 | 0 | 0.531 |
| 1390 | 1 | 1 | 1 | 0 | 1 | 0 | 5607.6 | 1962 | 3105 | 1 | 1 | 0.767 |
| 1457 | 1 | 1 | 1 | 0 | 1 | 0 | 5607.6 | 3330 | 3158 | 1 | 1 | 0.706 |

| | | | | | | | | | | | | |
|---|---|---|---|---|---|---|---|---|---|---|---|---|
| 1471 | 1 | 1 | 1 | 0 | 1 | 0 | 5607.6 | 4242 | 3159 | 0 | 0 | 0.42 |
| 1692 | 1 | 1 | 1 | 0 | 1 | 0 | 5607.6 | 1715 | 3414 | 1 | 1 | 0.868 |
| 1694 | 1 | 1 | 1 | 0 | 1 | 0 | 5607.6 | 1975 | 3403 | 1 | 1 | 0.919 |
| 1738 | 1 | 1 | 1 | 0 | 1 | 0 | 5607.6 | 2041 | 3438 | 1 | 1 | 0.767 |
| 1759 | 1 | 1 | 1 | 0 | 1 | 0 | 5607.6 | 4257 | 3453 | 1 | 1 | 0.695 |
| 1804 | 1 | 1 | 1 | 0 | 1 | 0 | 5607.6 | 4450 | 3480 | 1 | 1 | 0.688 |
| 1829 | 1 | 1 | 1 | 0 | 1 | 0 | 5607.6 | 3239 | 3579 | 0 | 0 | 0.211 |
| 1835 | 1 | 1 | 1 | 0 | 1 | 0 | 5607.6 | 3776 | 3550.5 | 1 | 1 | 0.699 |
| 1947 | 1 | 1 | 1 | 0 | 1 | 0 | 5607.6 | 1728 | 3713 | 1 | 0 | 0.591 |
| 2037 | 1 | 1 | 1 | 0 | 1 | 0 | 5607.6 | 2867 | 3819 | 0 | 0 | 0.413 |
| 2049 | 1 | 1 | 1 | 0 | 1 | 0 | 5607.6 | 3820.5 | 3826.5 | 1 | 1 | 0.798 |
| 2087 | 1 | 1 | 1 | 0 | 1 | 0 | 5607.6 | 3694 | 3869.5 | 1 | 1 | 0.847 |
| 2115 | 1 | 1 | 1 | 0 | 1 | 0 | 5607.6 | 2972 | 3921 | 0 | 0 | 0.451 |
| 2178 | 1 | 1 | 1 | 0 | 1 | 0 | 5607.6 | 2878 | 4039 | 0 | 0 | 0.358 |
| 2192 | 1 | 1 | 1 | 0 | 1 | 0 | 5607.6 | 3829.5 | 4011.5 | 1 | 1 | 0.838 |
| 2249 | 1 | 1 | 1 | 0 | 1 | 0 | 5607.6 | 3331 | 4126 | 0 | 0 | 0.435 |
| 2258 | 1 | 1 | 1 | 0 | 1 | 0 | 5607.6 | 4028 | 4113.5 | 1 | 0 | 0.605 |
| 2302 | 1 | 1 | 1 | 0 | 1 | 0 | 5607.6 | 1621 | 4241 | 0 | 0 | 0.438 |
| 2372 | 1 | 1 | 1 | 0 | 1 | 0 | 5607.6 | 3328 | 4331.5 | 1 | 0 | 0.516 |
| 2401 | 1 | 1 | 1 | 0 | 1 | 0 | 5607.6 | 3087 | 4359 | 1 | 0 | 0.538 |
| 2473 | 1 | 1 | 1 | 0 | 1 | 0 | 5607.6 | 3258 | 4507 | 0 | 0 | 0.512 |
| 2477 | 1 | 1 | 1 | 0 | 1 | 0 | 5607.6 | 3517 | 4495 | 0 | 0 | 0.47 |
| 2612 | 1 | 1 | 1 | 0 | 1 | 0 | 5607.6 | 4046 | 4697 | 1 | 1 | 0.669 |
| 2717 | 1 | 1 | 1 | 0 | 1 | 0 | 5607.6 | 2495 | 4913 | 0 | 0 | 0.194 |
| 2732 | 1 | 1 | 1 | 0 | 1 | 0 | 5607.6 | 3989 | 4886 | 1 | 0 | 0.516 |
| 2018 | 3 | 2 | 0.6667 | 0 | 1.3333 | 0 | 5607.6 | 4338 | 3780 | 1 | 1 | 0.892 |
| 1389 | 3 | 1 | 0.6667 | 0 | 0.6667 | 0 | 5607.6 | 1895 | 3070 | 1 | 1 | 0.757 |
| 1737 | 3 | 1 | 0.6667 | 0 | 0.6667 | 0 | 5607.6 | 1911 | 3443 | 1 | 1 | 0.926 |
| 2020 | 3 | 1 | 0.6667 | 0 | 0.6667 | 0 | 5607.6 | 4463 | 3759 | 1 | 1 | 0.949 |
| 1531 | 2 | 2 | 0.5 | 0 | 1 | 0 | 5607.6 | 4759 | 3188.5 | 0 | 0 | 0.486 |
| 1733 | 2 | 2 | 0.5 | 0 | 1 | 0 | 5607.6 | 1522 | 3460 | 1 | 0 | 0.614 |
| 395 | 2 | 1 | 0.5 | 0 | 0.5 | 0 | 5607.6 | 4546 | 2105 | 0 | 0 | 0.112 |
| 596 | 2 | 1 | 0.5 | 0 | 0.5 | 0 | 5607.6 | 5178 | 2294 | 0 | 0 | 0.169 |
| 965 | 2 | 1 | 0.5 | 0 | 0.5 | 0 | 5607.6 | 4088.5 | 2664.5 | 0 | 0 | 0.205 |
| 1164 | 2 | 1 | 0.5 | 0 | 0.5 | 0 | 5607.6 | 2082 | 2875 | 0 | 0 | 0.326 |

| | | | | | | | | | | | | |
|---|---|---|---|---|---|---|---|---|---|---|---|---|
| 1322 | 2 | 1 | 0.5 | 0 | 0.5 | 0 | 5607.6 | 983 | 3035 | 0 | 0 | 0.223 |
| 1381 | 2 | 1 | 0.5 | 0 | 0.5 | 0 | 5607.6 | 1116 | 3104 | 0 | 0 | 0.286 |
| 1386 | 2 | 1 | 0.5 | 0 | 0.5 | 0 | 5607.6 | 1636 | 3081 | 1 | 1 | 0.662 |
| 1448 | 2 | 1 | 0.5 | 0 | 0.5 | 0 | 5607.6 | 2418 | 3123 | 0 | 0 | 0.218 |
| 1460 | 2 | 1 | 0.5 | 0 | 0.5 | 0 | 5607.6 | 3538 | 3159 | 1 | 1 | 0.73 |
| 1693 | 2 | 1 | 0.5 | 0 | 0.5 | 0 | 5607.6 | 1845 | 3409 | 1 | 1 | 0.986 |
| 1766 | 2 | 1 | 0.5 | 0 | 0.5 | 0 | 5607.6 | 4834 | 3460 | 1 | 0 | 0.577 |
| 1945 | 2 | 1 | 0.5 | 0 | 0.5 | 0 | 5607.6 | 1532 | 3684 | 0 | 0 | 0.49 |
| 1949 | 2 | 1 | 0.5 | 0 | 0.5 | 0 | 5607.6 | 1988 | 3702 | 0 | 0 | 0.499 |
| 1976 | 2 | 1 | 0.5 | 0 | 0.5 | 0 | 5607.6 | 4462 | 3700 | 1 | 1 | 0.94 |
| 2019 | 2 | 1 | 0.5 | 0 | 0.5 | 0 | 5607.6 | 4398.5 | 3762.5 | 1 | 1 | 0.942 |
| 2116 | 2 | 1 | 0.5 | 0 | 0.5 | 0 | 5607.6 | 3064 | 3919 | 0 | 0 | 0.426 |
| 2187 | 2 | 1 | 0.5 | 0 | 0.5 | 0 | 5607.6 | 3455 | 4046 | 1 | 0 | 0.583 |
| 2188 | 2 | 1 | 0.5 | 0 | 0.5 | 0 | 5607.6 | 3539 | 4007 | 1 | 1 | 0.71 |
| 2265 | 2 | 1 | 0.5 | 0 | 0.5 | 0 | 5607.6 | 902 | 4159 | 0 | 0 | 0.175 |
| 2329 | 2 | 1 | 0.5 | 0 | 0.5 | 0 | 5607.6 | 1231 | 4257 | 0 | 0 | 0.336 |
| 2332 | 2 | 1 | 0.5 | 0 | 0.5 | 0 | 5607.6 | 1557 | 4281 | 0 | 0 | 0.443 |
| 2333 | 2 | 1 | 0.5 | 0 | 0.5 | 0 | 5607.6 | 2828 | 4299 | 0 | 0 | 0.324 |
| 2399 | 2 | 1 | 0.5 | 0 | 0.5 | 0 | 5607.6 | 2959 | 4366 | 0 | 0 | 0.449 |
| 2474 | 2 | 1 | 0.5 | 0 | 0.5 | 0 | 5607.6 | 3321 | 4466 | 1 | 0 | 0.558 |
| 2540 | 2 | 1 | 0.5 | 0 | 0.5 | 0 | 5607.6 | 3847 | 4593 | 1 | 1 | 0.683 |
| 2541 | 2 | 1 | 0.5 | 0 | 0.5 | 0 | 5607.6 | 3976 | 4588 | 1 | 0 | 0.651 |
| 2543 | 2 | 1 | 0.5 | 0 | 0.5 | 0 | 5607.6 | 4236 | 4577 | 0 | 0 | 0.325 |
| 2610 | 2 | 1 | 0.5 | 0 | 0.5 | 0 | 5607.6 | 3850 | 4668 | 1 | 1 | 0.804 |
| 2817 | 2 | 1 | 0.5 | 0 | 0.5 | 0 | 5607.6 | 3413 | 5098 | 0 | 0 | 0.18 |
| 2819 | 2 | 1 | 0.5 | 0 | 0.5 | 0 | 5607.6 | 3739 | 5121 | 0 | 0 | 0.374 |
| 2820 | 2 | 1 | 0.5 | 0 | 0.5 | 0 | 5607.6 | 3869 | 5116 | 0 | 0 | 0.454 |
| 2821 | 2 | 1 | 0.5 | 0 | 0.5 | 0 | 5607.6 | 3999 | 5110 | 0 | 0 | 0.467 |
| 882 | 3 | 1 | 0.3333 | 0 | 0.3333 | 0 | 5607.6 | 2133 | 2618 | 0 | 0 | 0.328 |
| 1027 | 3 | 1 | 0.3333 | 0 | 0.3333 | 0 | 5607.6 | 4414 | 2723 | 0 | 0 | 0.237 |
| 1246 | 3 | 1 | 0.3333 | 0 | 0.3333 | 0 | 5607.6 | 3907.5 | 2933 | 1 | 0 | 0.521 |
| 1318 | 3 | 1 | 0.3333 | 0 | 0.3333 | 0 | 5607.6 | 5207 | 2965 | 0 | 0 | 0.334 |
| 2158 | 3 | 1 | 0.3333 | 0 | 0.3333 | 0 | 5607.6 | 3635 | 3983.5 | 1 | 1 | 0.84 |
| 881 | 1 | 1 | 0 | 1 | 0 | 0 | 5607.6 | 2064 | 2592 | 0 | 0 | 0.317 |
| 1312 | 1 | 1 | 0 | 1 | 0 | 0 | 5607.6 | 4716.5 | 2987 | 0 | 0 | 0.278 |

| | | | | | | | | | | | | |
|---|---|---|---|---|---|---|---|---|---|---|---|---|
| 1948 | 1 | 1 | 0 | 1 | 0 | 0 | 5607.6 | 1858 | 3707 | 1 | 0 | 0.57 |
| 2810 | 1 | 1 | 0 | 1 | 0 | 0 | 5607.6 | 2764 | 5126 | 0 | 0 | 0.197 |
| 2025 | 3 | 1 | 0 | 0.6667 | 0 | 0 | 5607.6 | 1082 | 3815 | 0 | 0 | 0.201 |
| 615 | 2 | 1 | 0 | 0.5 | 0 | 0 | 5607.6 | 2691 | 2340 | 0 | 0 | 0.138 |
| 1515 | 2 | 1 | 0 | 0.5 | 0 | 0 | 5607.6 | 3656 | 3181 | 1 | 1 | 0.668 |
| 1765 | 2 | 1 | 0 | 0.5 | 0 | 0 | 5607.6 | 4768 | 3427 | 1 | 0 | 0.646 |
| 2468 | 2 | 1 | 0 | 0.5 | 0 | 0 | 5607.6 | 2542 | 4500 | 0 | 0 | 0.196 |
| 975 | 2 | 1 | 0 | 0 | 0 | 0 | 5607.6 | 4858 | 2686 | 0 | 0 | 0.348 |
| 1405 | 2 | 1 | 0 | 0 | 0 | 0 | 5607.6 | 3278 | 3098 | 1 | 0 | 0.603 |
| 1409 | 2 | 1 | 0 | 0 | 0 | 0 | 5607.6 | 3528.5 | 3098.5 | 1 | 1 | 0.691 |
| 1546 | 2 | 1 | 0 | 0 | 0 | 0 | 5607.6 | 1968 | 3254 | 1 | 1 | 0.897 |
| 1595 | 2 | 1 | 0 | 0 | 0 | 0 | 5607.6 | 1905 | 3294 | 1 | 1 | 0.964 |
| 1658 | 2 | 1 | 0 | 0 | 0 | 0 | 5607.6 | 3290 | 3319 | 0 | 0 | 0.494 |
| 1755 | 2 | 1 | 0 | 0 | 0 | 0 | 5607.6 | 3997 | 3447.5 | 1 | 1 | 0.786 |
| 1811 | 2 | 1 | 0 | 0 | 0 | 0 | 5607.6 | 876 | 3562 | 0 | 0 | 0.133 |
| 2176 | 2 | 1 | 0 | 0 | 0 | 0 | 5607.6 | 1807 | 4046 | 0 | 0 | 0.282 |
| 2261 | 2 | 1 | 0 | 0 | 0 | 0 | 5607.6 | 4347 | 4123 | 1 | 0 | 0.516 |
| 2330 | 2 | 1 | 0 | 0 | 0 | 0 | 5607.6 | 1361 | 4252 | 0 | 0 | 0.425 |
| 2611 | 2 | 1 | 0 | 0 | 0 | 0 | 5607.6 | 3916 | 4703 | 1 | 1 | 0.807 |
| 2674 | 2 | 1 | 0 | 0 | 0 | 0 | 5607.6 | 3856 | 4817 | 1 | 1 | 0.687 |
| 2823 | 2 | 1 | 0 | 0 | 0 | 0 | 5607.6 | 4195 | 5139 | 0 | 0 | 0.342 |
| 806 | 1 | 1 | 0 | 0 | 0 | 0 | 5607.6 | 4663 | 2526 | 0 | 0 | 0.243 |
| 1043 | 1 | 1 | 0 | 0 | 0 | 0 | 5607.6 | 1297 | 2760 | 0 | 0 | 0.137 |
| 1629 | 1 | 1 | 0 | 0 | 0 | 0 | 5607.6 | 4633 | 3286 | 1 | 0 | 0.593 |
| 1645 | 1 | 1 | 0 | 0 | 0 | 0 | 5607.6 | 1908 | 3369 | 1 | 1 | 0.997 |
| 1980 | 1 | 1 | 0 | 0 | 0 | 0 | 5607.6 | 4720 | 3734 | 1 | 0 | 0.63 |
| 2035 | 1 | 1 | 0 | 0 | 0 | 0 | 5607.6 | 2739 | 3825 | 0 | 0 | 0.354 |
| 2606 | 1 | 1 | 0 | 0 | 0 | 0 | 5607.6 | 3590 | 4679 | 1 | 0 | 0.568 |
| 2671 | 1 | 1 | 0 | 0 | 0 | 0 | 5607.6 | 3660 | 4788 | 1 | 0 | 0.577 |
| 2854 | 1 | 1 | 0 | 0 | 0 | 0 | 5607.6 | 4068 | 5219 | 0 | 0 | 0.368 |

**Definition of labels:**

**Spatial microdomain (hexbin)**: A spatial microdomain represents a discrete tissue neighborhood defined by aggregating spatially adjacent nuclei using hexagonal binning of spatial transcriptomics coordinates. Each microdomain summarizes the local cellular composition and molecular features within that region. Hexagonal binning was applied to treat each spatial unit uniformly regardless of local nuclear density, consistent with an analytical framework focused on tissue niche co-occurrence rather than cellular clustering. All microdomains share a uniform area of 5,607.6 $\mu m^2$ per hexagonal unit, derived from a bin size of 30 applied across a coordinate span of 4,831 pixels, converted using the Visium platform scale factor of 0.4997 µm per pixel.

**Table Ssort order**: Rows are ordered by decreasing niche completeness. Microdomains with maximal stress signal and FAP/MSC co-occurrence appear first (frac_stress = 1 and frac_FAP > 0, n = 24), followed by microdomains with maximal stress signal only (frac_stress = 1 and frac_FAP = 0, n = 40), microdomains with partial stress signal and FAP/MSC co-occurrence (n = 26), microdomains with partial stress signal only (n = 43), and microdomains containing TRM without detected stress or FAP/MSC signal (frac_stress = 0, n = 32). Within each group, rows are further ordered by decreasing niche_TRM_x_stress_x_FAP, frac_stress, frac_FAP, and n_TRM.

**n_cells**: Total number of nuclei captured within a given spatial microdomain.

**n_TRM**: Number of ZNF683+ TRM nuclei present within a given spatial microdomain.

**TRM (ZNF683+TC): Nuclei classified as tissue-resident memory T cells based on co-expression of *ZNF683* > 1 and *CD247* > 1, with exclusion of nuclei expressing *ZBTB16* ≥ 0.5 to avoid overlap with the iNKT-like_TC population. Classification criteria for all spatially mapped populations are given in Table S41. *IL7R*, while expressed in this population in the snRNA-seq dataset, was not used as a classification criterion for spatial transcriptomics analysis.**

**frac_stress**: Fraction of nuclei within a spatial microdomain classified as stress-associated, defined by co-expression of *HMGB1* > 0 and *G3BP1* > 0, consistent with activation of cellular stress and stress granule-associated pathways. Unlike single-nucleus RNA sequencing, spatial transcriptomics captures polyadenylated RNA from both nuclear and cytoplasmic compartments, enabling detection of cytoplasmic stress granule-associated transcripts including *G3BP1* that may be underrepresented in nuclear preparations. A value of 1 indicates that all nuclei within the microdomain expressed both stress markers.

**frac_FAP**: Fraction of nuclei within a spatial microdomain classified as FAP/MSC, defined by expression of *PDGFRA* > 1, consistent with the gene signature criteria applied in the snRNA-seq subclustering analysis (Table S41). For spatial microdomains, FAP/MSC nuclei with additional inflammatory co-expression of *HMGB1* > 0 and/or *CXCL12* > 1 are represented in Fig. 6E to reflect the stromal signaling environments implicated in immune retention. In this table, frac_FAP reflects the broader *PDGFRA* > 1 classification only.

**niche_TRM_x_stress**: Derived metric calculated as the product of n_TRM and frac_stress, reflecting the degree to which TRM abundance coincides with stress-associated cellular environments within a spatial microdomain. Higher values indicate microdomains in which more TRM are present alongside a greater fraction of stress-associated nuclei.

**niche_TRM_x_stress_x_FAP**: Derived metric calculated as the product of n_TRM, frac_stress, and frac_FAP, reflecting the degree to which TRM abundance coincides with stress-associated nuclei and FAP/MSC stromal support within the same spatial microdomain. This metric identifies microdomains in which all three components of the inflammatory–myofiber–interstitial axis are simultaneously present. A value of 0 indicates absence of at least one component. The maximum observed value reflects the microdomain with the greatest convergence of all three signals.

**hex_area_um2**: Area of each hexagonal spatial microdomain expressed in square micrometres ($\mu m^2$). All microdomains share a uniform area of 5,607.6 $\mu m^2$ by construction of the hexagonal binning grid.

**spatial_1, spatial_2**: Pixel coordinates corresponding to the geometric center of each spatial microdomain in the full-resolution tissue image, provided for reference and traceability to the original spatial transcriptomics map (Supplementary Fig. 6).

**in_TRM_KDE_hotspot**: Binary indicator (1 = yes, 0 = no) denoting whether a microdomain centroid falls within the TRM kernel density estimation (KDE) enrichment zone. KDE was computed over microdomain centroid coordinates weighted by n_TRM using a Gaussian kernel with Silverman bandwidth, evaluated on a 300 × 300 grid spanning the full tissue coordinate range. The hotspot threshold was defined as the 90th percentile of positive TRM KDE density values, identifying the top 10% of the tissue area by TRM signal concentration. This threshold was applied uniformly across all niche types to enable direct spatial comparison. Of 165 TRM-containing microdomains, 81 (49.1%) fell within the TRM KDE hotspot zone. Two complementary area measures are reported throughout: a count-based microdomain area, calculated as the number of qualifying microdomains multiplied by the uniform hexagonal unit area (5,607.6 µm²), and a KDE zone area, calculated as the spatial extent of the density-thresholded enrichment polygon. These two measures are not directly comparable, as the former sums discrete hexagonal units while the latter measures a continuous density-thresholded polygon. Count-based areas are used for all microdomain area values reported in the main text, including the total TRM-occupied area (165 microdomains, 925,254 µm², 0.93 mm²) and the stress-containing subset (133 microdomains with frac_stress > 0, 745,811 µm², 80.6% of TRM-containing microdomains). The 81 hotspot microdomains correspond to a count-based area of 454,216 µm² (0.45 mm²) and a KDE zone area of 505,681 µm² (0.51 mm²). The former is the value reported in the main text. Zone-to-zone overlap comparisons below are computed on KDE zone areas. The stress myonuclear KDE zone (550,350 µm²) overlapped the TRM hotspot at a Jaccard index of 0.640, with 74.9% of the stress KDE zone area falling within the TRM hotspot zone. The FAP/MSC inflammatory KDE zone (447,135 µm²) showed a Jaccard index of 0.415 against the TRM hotspot, with 62.5% of the FAP/MSC KDE zone area falling within the TRM hotspot zone. Microdomains in which all three niche components were simultaneously present (frac_stress > 0 and frac_FAP > 0, equivalently niche_TRM_x_stress_x_FAP > 0, n = 50, 30.3%) defined the triple-niche convergence zone, corresponding to a count-based area of 280,380 µm² (0.28 mm²), the value reported in the main text. For reference, the density-thresholded triple-niche KDE zone spanned 544,001 µm² (hotspot) and 211,543 µm² (core) (Jaccard vs TRM hotspot = 0.383).

**in_TRM_KDE_core**: Binary indicator (1 = yes, 0 = no) denoting whether a microdomain centroid falls within the TRM KDE core zone, defined as the top 5% of positive TRM KDE density values. Of 165 TRM-containing microdomains, 50 (30.3%) fell within the core zone, corresponding to the region of highest TRM signal concentration within IBM tissue: a count-based area of 280,380 µm² (0.28 mm²) and a KDE zone area of 252,841 µm² (0.25 mm²). This count is equal to, but defined independently of, the 50 microdomains constituting the triple-niche convergence zone: the core zone is defined by TRM density, whereas the triple-niche zone is defined by marker composition. The two sets share 16 microdomains (Jaccard index = 0.190), indicating that the equal counts do not reflect a common set of tissue regions.

Membership under each criterion is given in the in_TRM_KDE_core and niche_TRM_x_stress_x_FAP columns.

**TRM_hotspot_proximity**: Continuous score ranging from 0 to 1, representing the normalised TRM KDE density at each microdomain centroid relative to the maximum observed TRM KDE density across the tissue. A score of 1.0 indicates the microdomain of peak TRM signal concentration. Scores approaching 0 indicate microdomains at the periphery of or outside the TRM enrichment zone. This score provides a spatially continuous measure of each microdomain's position within the TRM retention landscape, complementing the binary hotspot classification.

Table S45. List of antibodies used for identifying proteins of interest (primary antibody presented in order of appearance in the article)

| Antibody | Isotype | Clone | Assay | Dilution | Source |
|---|---|---|---|---|---|
| Anti-PLZF | Mouse $IgG_{1, \kappa}$ | D-9 | IF | 1:100 | Santa Cruz Biotechnology |
| CD8A | Rat $IgG_{2, \beta}$ | YTC182.20 | IF | 1:100 | Abcam |
| KLRK1 | Mouse $IgG_1$ | 1D11 | IF | 1:100 | Novus Biologicals |
| ZNF683 | Rabbit IgG | Custom sequence position 505-524* | IF | 1:100 | Cosmo Bio Co |
| FLT3 | Rabbit IgG | EP1140Y | IF | 1:100 | Abcam |
| CD1C | Goat polyclonal IgG | - | IF | 1:100 | R&D Systems |
| CAV3 | Goat polyclonal IgG | - | IF | 1:100 | GeneTex |
| CD8 | Mouse $IgG_1$ | DK25 | IF/IHC | 1:100 | Dako/Agilent |
| CD68 | Mouse $IgG_1$ | KP1 | IF/IHC | 1:100 | Dako/Agilent |
| p62 | Rabbit polyclonal IgG | - | IF | 1:100 | Enzo Life Sciences |
| Alexa Fluor™ 488, 568, and 674 | Goat polyclonal IgG | - | IF | 1:1000 | Molecular Probes |

IF – immunofluorescence

* – “[H]CRAVSLSSAGTPLVMGQDQNN[OH]

Table S46. List of primers used for identifying genes of interest

| Target Gene | Primer Direction | Sequence (5'-3') |
|---|---|---|
| *ZNF683* | Forward | CATATGTGGCAAGAGCTTTGG |
| *ZNF683* | Reverse | GGCAAGTTGAGTGAAGCTCT |
| *POLR2A* | Forward | ATCTCTCCTGCCATGACACC |
| *POLR2A* | Reverse | AGACCAGGCAGGGGAGTAAC |
